# Risk of Severe Outcomes From COVID-19 in Immunocompromised People During the Omicron Era: A Systematic Review and Meta-Analysis

**DOI:** 10.1101/2024.11.25.24317895

**Authors:** Akvile Chapman, Francis Berenbaum, Giuseppe Curigliano, Triantafyllos Pliakas, Aziz Sheikh, Sultan Abduljawad

**Author notes:** **Correspondence to:** Sultan Abduljawad, BioNTech UK Ltd., London, UK.

## Abstract

**Key Points:** **Question:** What are the risks of severe outcomes from COVID-19 in people with immunocompromising/immunosuppressive (IC/IS) conditions in the Omicron era?

**Findings:** This systematic review and meta-analysis found increased risk of severe outcomes for people with IC/IS conditions (e.g., autoimmunity, cancer, liver disease, renal disease, transplant) compared with people without the respective conditions.

Of all meta-analyzed conditions, transplant recipients had the highest risk of severe COVID-19 outcomes, compared with non-transplant recipients or the general population.

**Meaning:** People with IC/IS conditions remain at increased risk of severe outcomes from COVID-19 during the Omicron era; continued preventative measures and personalized care are crucial.

**Importance:** This is the first meta-analysis to investigate the risk of severe outcomes for individuals with immunocompromising/immunosuppressive (IC/IS) conditions specifically in the Omicron era.

**Objective:** To assess the risk of mortality and hospitalization from COVID-19 in people with IC/IS conditions compared with people without IC/IS conditions during the Omicron era.

**Data Sources:** A systematic search of Embase, MEDLINE, PubMed, Europe PMC, Latin American and Caribbean Health Sciences Literature, Cochrane COVID-19 Study Register, and WHO COVID-19 Database was performed to identify studies published between 1 January 2022 and 13 March 2024.

**Study Selection:** Inclusion criteria were observational studies that included people (all ages) with at least 1 of the following conditions: IC/IS unspecified groups, transplant (solid organ, stem cells, or bone marrow), any malignancy, autoimmune diseases, any liver diseases, chronic or end-stage kidney disease, and advanced/untreated HIV. In total, 72 studies were included in the review, of which 66 were included in the meta-analysis.

**Data Extraction and Synthesis:** Data were extracted by one reviewer and verified by a second. Studies were synthesized quantitively (meta-analysis) using random-effect models. PRISMA guidelines were followed.

**Main Outcomes and Measures:** Evaluated outcomes were risks of death, hospitalization, intensive care unit (ICU) admission, and any combination of these outcomes. Odds ratios, hazard ratios, and rate ratios were extracted; pooled relative risk (RR) and 95% confidence intervals (CI) were calculated.

**Results:** Minimum numbers of participants per IC/IS condition ranged from 12 634 to 3 287 816. Risks of all outcomes were increased in people with all meta-analyzed IC/IS conditions compared with people without the respective conditions. Of all meta-analyzed IC/IS conditions, transplant recipients had the highest risk of death (RR, 6.78; 95% CI, 4.41-10.43; *P*<.001), hospitalization (RR, 6.75; 95% CI, 3.41-13.37; *P*<.001), and combined outcomes (RR, 8.65; 95% CI, 4.01-18.65; *P*<.001), while participants in the unspecified IC/IS group had the highest risk of ICU admission (RR, 3.38; 95% CI, 2.37-4.83; *P*<.001) compared with participants without the respective IC/IS conditions or general population.

**Conclusions:** In the Omicron era, people with IC/IS conditions have a substantially higher risk of death and hospitalization from COVID-19 than people without these conditions.

## Introduction

SARS-CoV-2, the virus causing COVID-19, emerged in 2019 and was declared a pandemic in March 2020 by the World Health Organization (WHO).^1,2^ In November 2021, the Omicron variant was identified and designated a variant of concern,^3^ displaying higher transmissibility but fewer severe outcomes than Delta.^4–8^ To date, there are over 7 million deaths worldwide,^9^ and although COVID-19 is no longer designated a global health emergency, it remains a threat due to evolving Omicron subvariants causing spikes in infections and fatalities.^10,11^

Despite effective preventative measures, COVID-19 still imposes a high burden on immunocompromised people. Immunocompromising or immunosuppressive (IC/IS) conditions vary in type and severity (i.e., moderate to severe), and negatively impact the ability of the immune system to combat pathogens, such as SARS-CoV-2.^12–14^ IC/IS conditions may be genetically acquired, caused directly by a disease (HIV/AIDS), or result from immunosuppressive therapies (e.g., medications for transplant recipients or autoimmune diseases).^15^ Though often used interchangeably in the literature, the term “immunocompromised” is used here to describe people with an impaired immune system due to a health condition, while “immunosuppression” is considered a result of treatment or medication.

People with IC/IS conditions may experience persistent SARS-CoV-2 infection, which can drive the evolution of new variants.^16,17^ Additionally, previous studies show that people with IC/IS conditions tend to have a higher risk of severe COVID-19 and death than people without IC/IS conditions.^14,15,18^ The ability to clear the virus and the level of risk for severe outcomes can vary widely depending on the IC/IS etiology and severity.^16^

The continuous evolution and global circulation of Omicron subvariants remains a significant threat to the IC/IS population.^14,19^ Thus, developing a comprehensive understanding of the burden of Omicron subvariants on people with IC/IS conditions is crucial for improving prevention, treatment methods, and public health policy. This systematic literature review (SLR) and meta-analysis aimed to assess the risk of hospitalization and mortality from COVID-19 in people with IC/IS conditions compared with people without IC/IS conditions in the Omicron era.

## Methods

The SLR protocol is registered with PROSPERO (CRD42024501163). This SLR and meta-analysis adheres to the Preferred Reporting Items for Systematic Reviews and Meta-Analysis guidelines (PRISMA).

### Search Strategy

The following databases were searched: Embase, MEDLINE, PubMed, Europe PMC (including medRix and bioRxiv preprints), Latin American and Caribbean Health Sciences Literature, the Cochrane COVID-19 Study Register, and the WHO COVID-19 Database. Search strategies were structured using terms related to COVID-19 infection, risk, and burden of illness (**eMethods**).

### Eligibility Criteria

Inclusion criteria for studies are detailed in **eTable 1**. Participants (all ages) were included in the review if they were (i) defined as ‘IC/IS’ by the study authors (for clarity, referred as “IC/IS unspecified” thereafter), (ii) taking immunosuppressive drugs, (iii) receiving radiotherapy treatment, or (iv) had multiple immunocompromising conditions, including transplant (solid organ, stem cell, or bone marrow), any malignancy, liver disease, kidney disease (chronic or end-stage), or advanced/untreated HIV.

Individuals without the respective IC/IS condition or the general population (as defined by study authors) were used as a comparator group. Evaluated outcomes were the risk of hospitalization (for any reason), intensive care unit (ICU) admission (for any reason, with or without ventilatory support), or death. Additionally, a combined outcome was evaluated, which was defined as a combination of any of the above outcomes. COVID-19 outcomes were determined by including studies where either all patients had COVID-19 at the start of the study or all deaths and hospitalizations were related to COVID-19 (defined by the studies).

Included studies were observational (cohort, case-control, cross-sectional), published between 1 January 2022 and 13 March 2024, and had full texts published in English.

### Data Synthesis and Analysis

#### Qualitative Data Synthesis

All studies included in the review were assessed qualitatively to identify which studies could be combined in a meta-analysis.

#### Statistical Analysis

For the primary analysis, pairwise meta-analyses were performed for the risk of death, hospitalization, ICU admission, and the combined outcome for each IC/IS condition, using the most adjusted reported outcome estimates. The robustness of the results was assessed using ‘Leave-1-out’,^20^ ‘Least adjusted’, ‘Only adjusted’, and ‘Excluding studies for population overlap’ sensitivity analyses. Further details are available in the **eMethods**.

Subgroup analyses were conducted for the ‘Hospitalized’ or ‘General’ populations, which included only people who were or were not already hospitalized when they started the study, respectively. Additional subgroup analyses are described in **Section 1.5.2** of the **eMethods.**

All statistical analyses were performed in R version 4.1.1 (R Foundation for Statistical Computing) using the meta package. A statistically significant (*P* < 0.05) result is referred to as significant thereafter.

## Results

In total, 21 937 records were identified through searches and 1 study was identified via reference checking. Following elimination of duplicates, 11 593 remaining studies underwent title and abstract screening, of which, 3 123 studies were assessed in full text screening. A total of 72 studies were selected for inclusion in the review, 66 of which were included in the meta-analyses (**Figure 1**).

**Figure 1.**
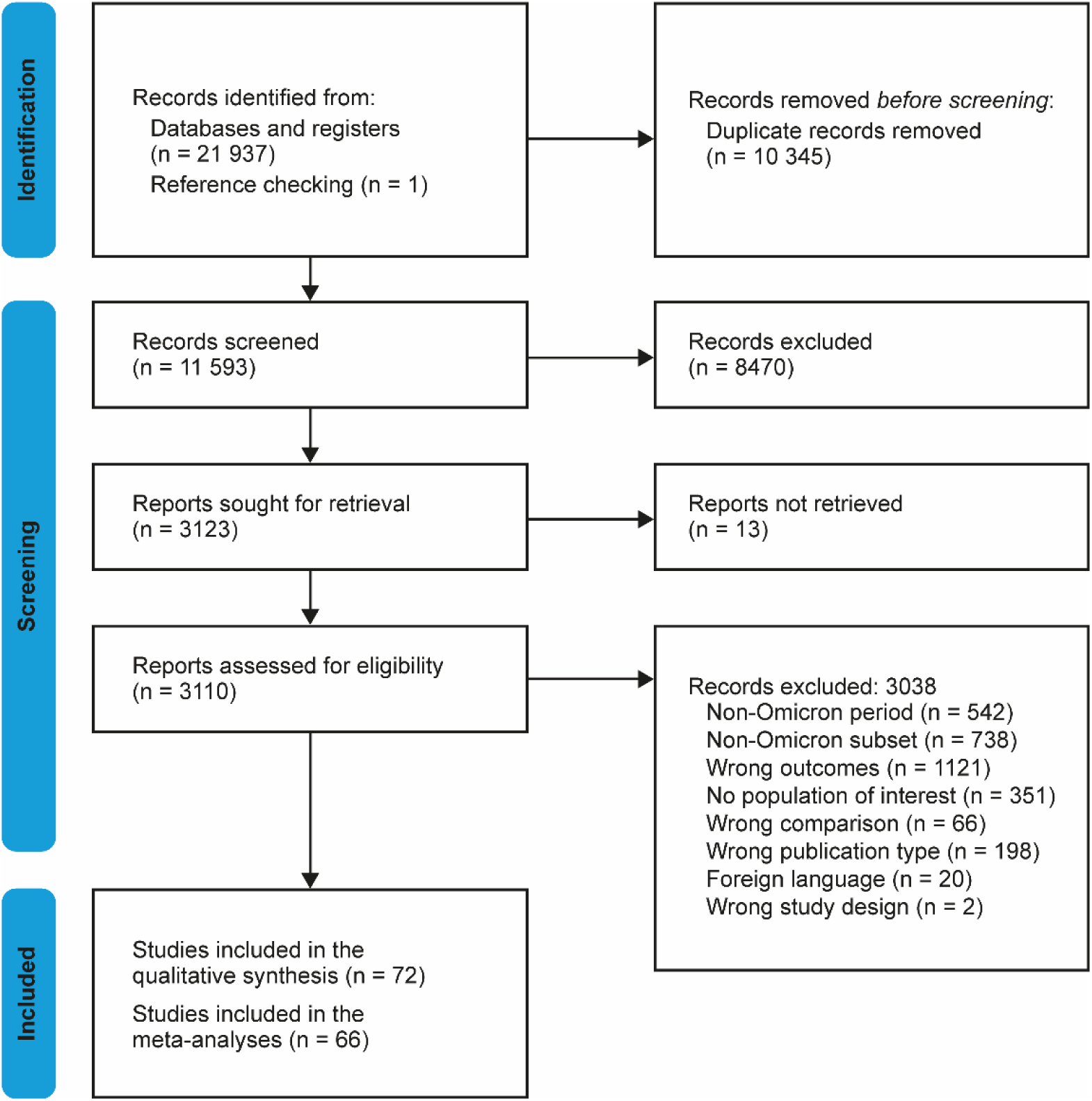
PRISMA Flow Diagram.

### Study Characteristics

Studies were performed in 25 different countries, primarily in European countries (n = 20), China (n = 13), and the USA (n = 11). Most were retrospective cohort studies (n = 55), followed by prospective cohort (n = 8), cross-sectional (n = 6), and case-control studies (n = 3) (**Table 1**). The ‘Death’, ‘Hospitalization’, ‘ICU’, and ‘Combined’ outcomes were reported in 43, 22, 16, and 19 studies, respectively. Most studies did not report Omicron subvariants, but for those that did (n = 20), BA.1 was the most common (**Table 1**). The Omicron period for each study is shown in **eTable 2**.

**Table 1.**
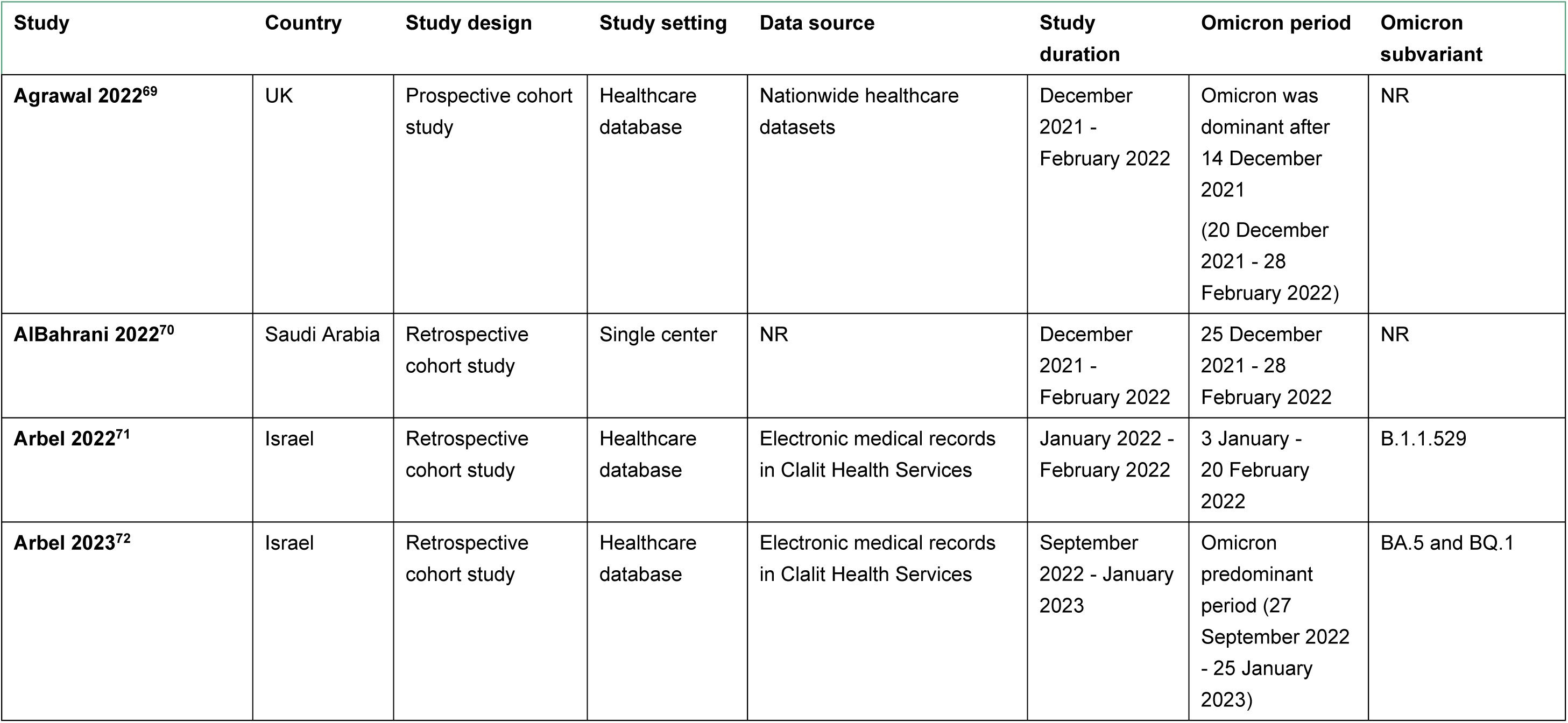
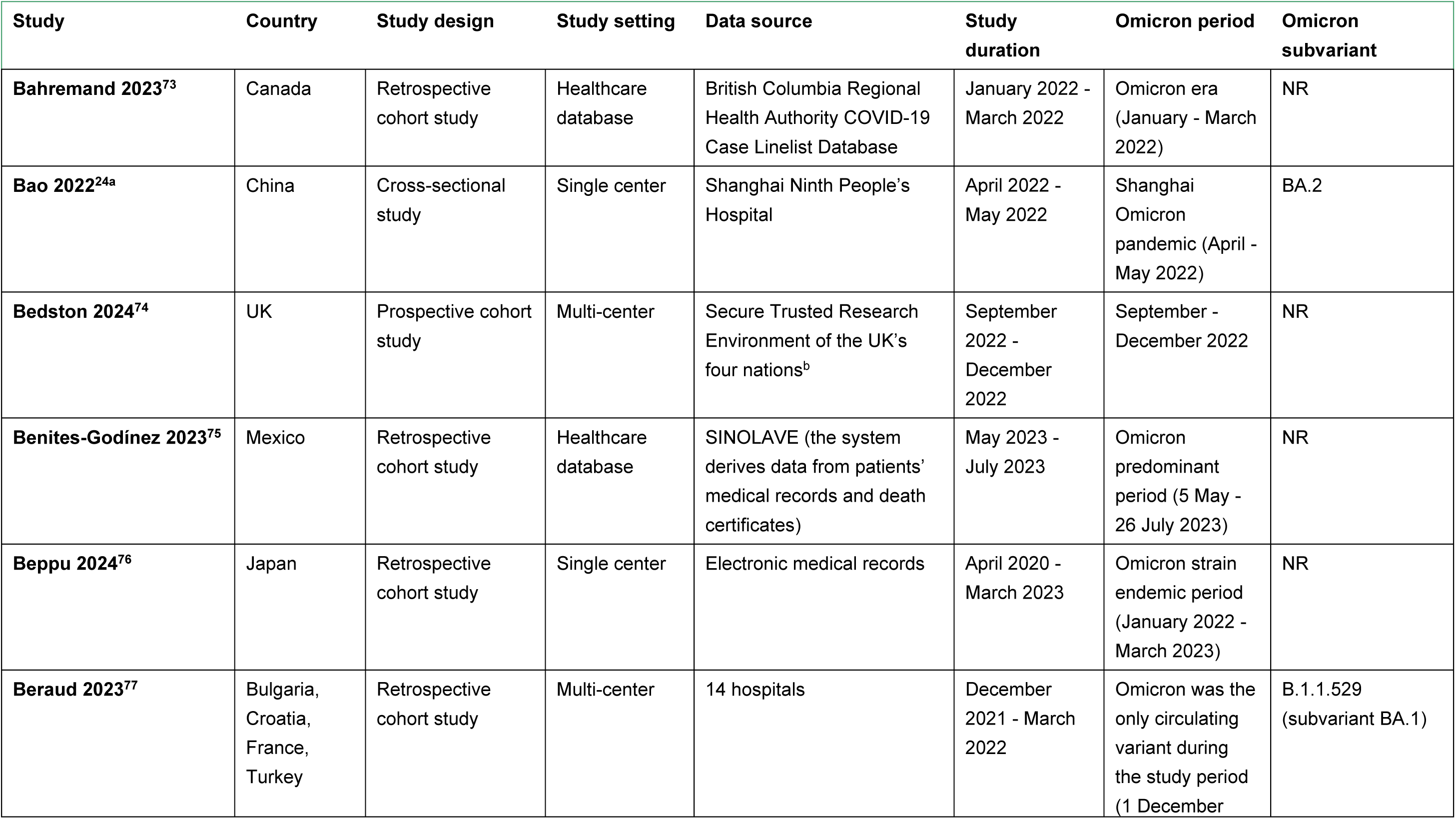
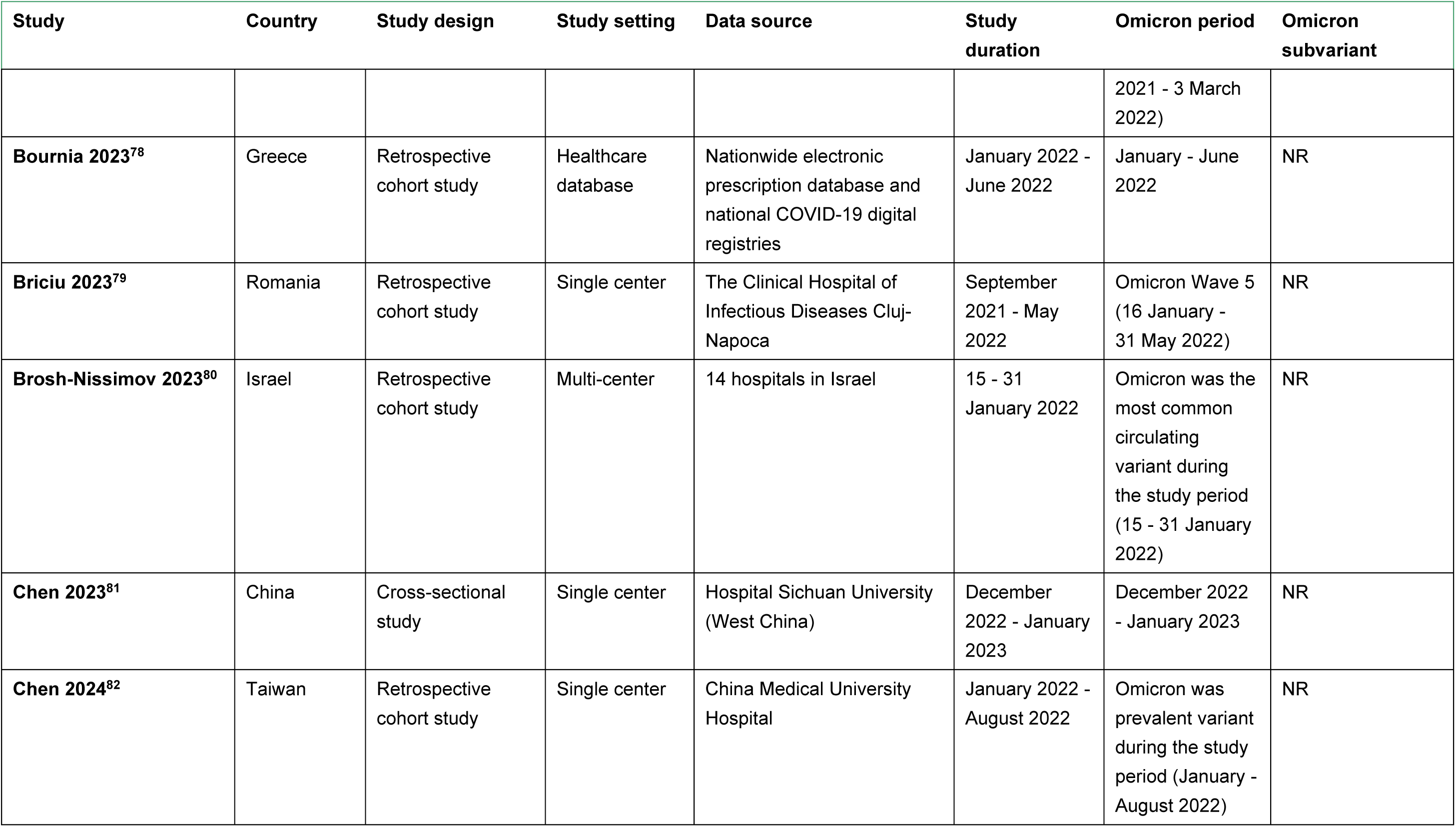
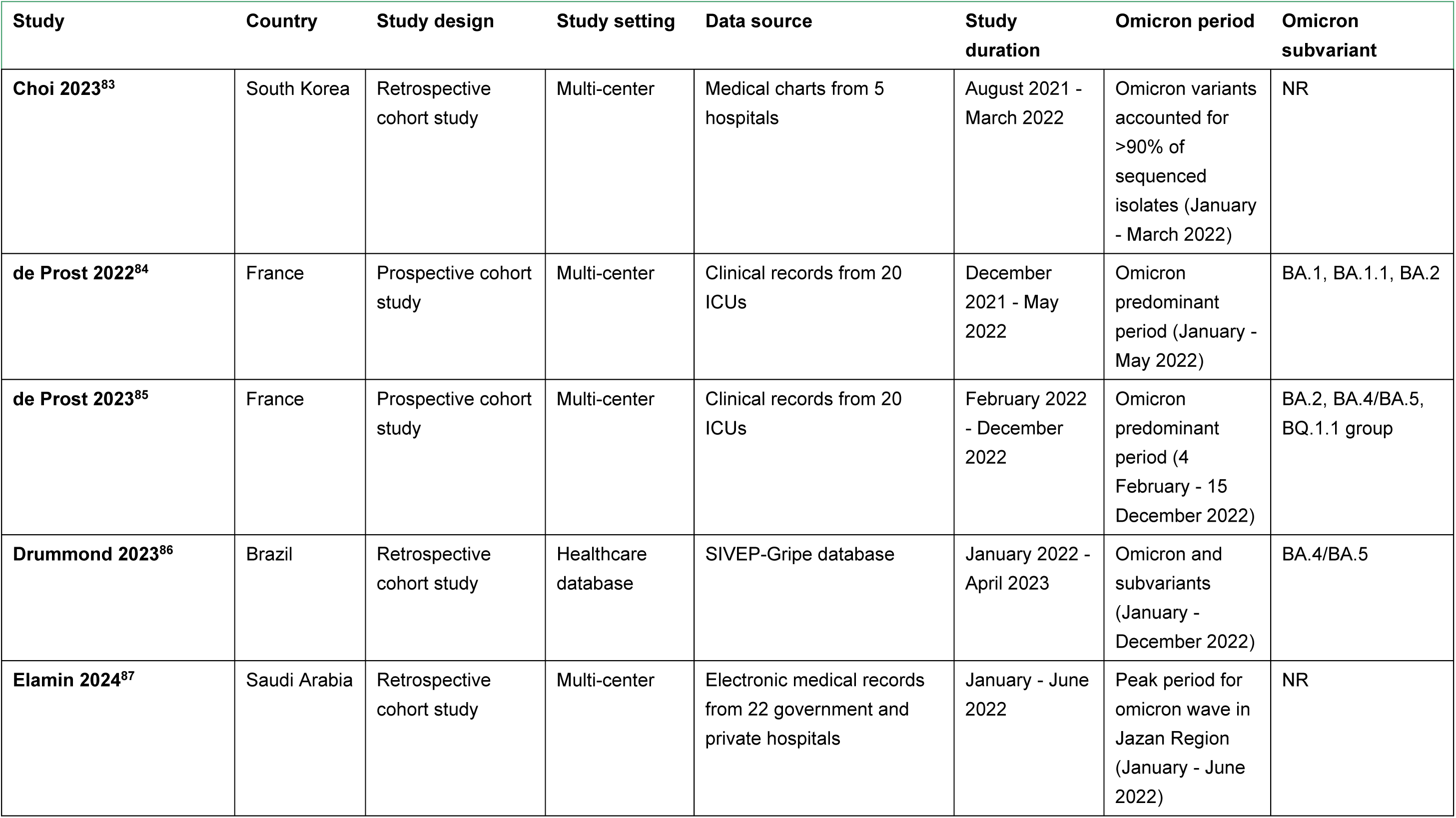
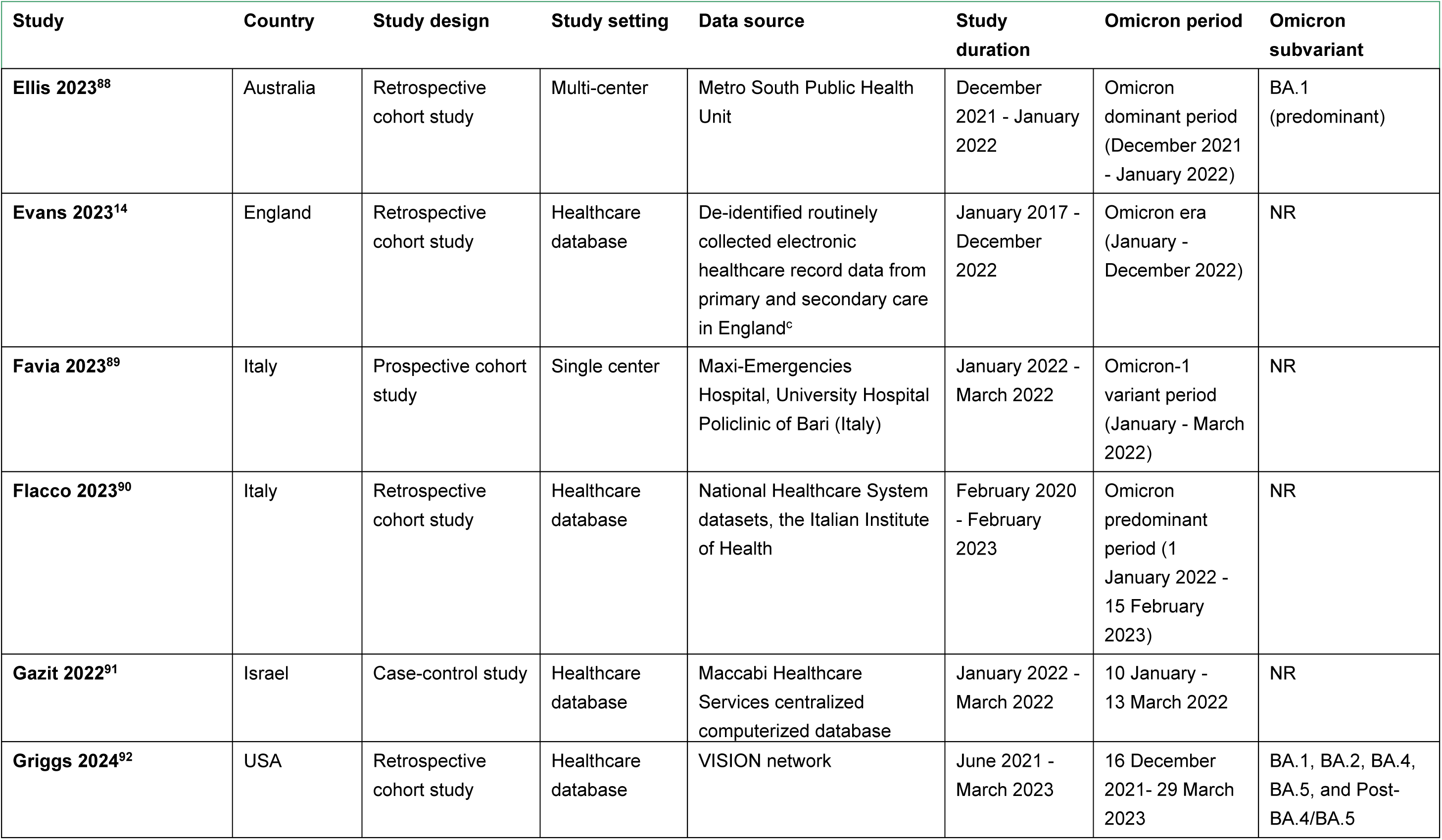
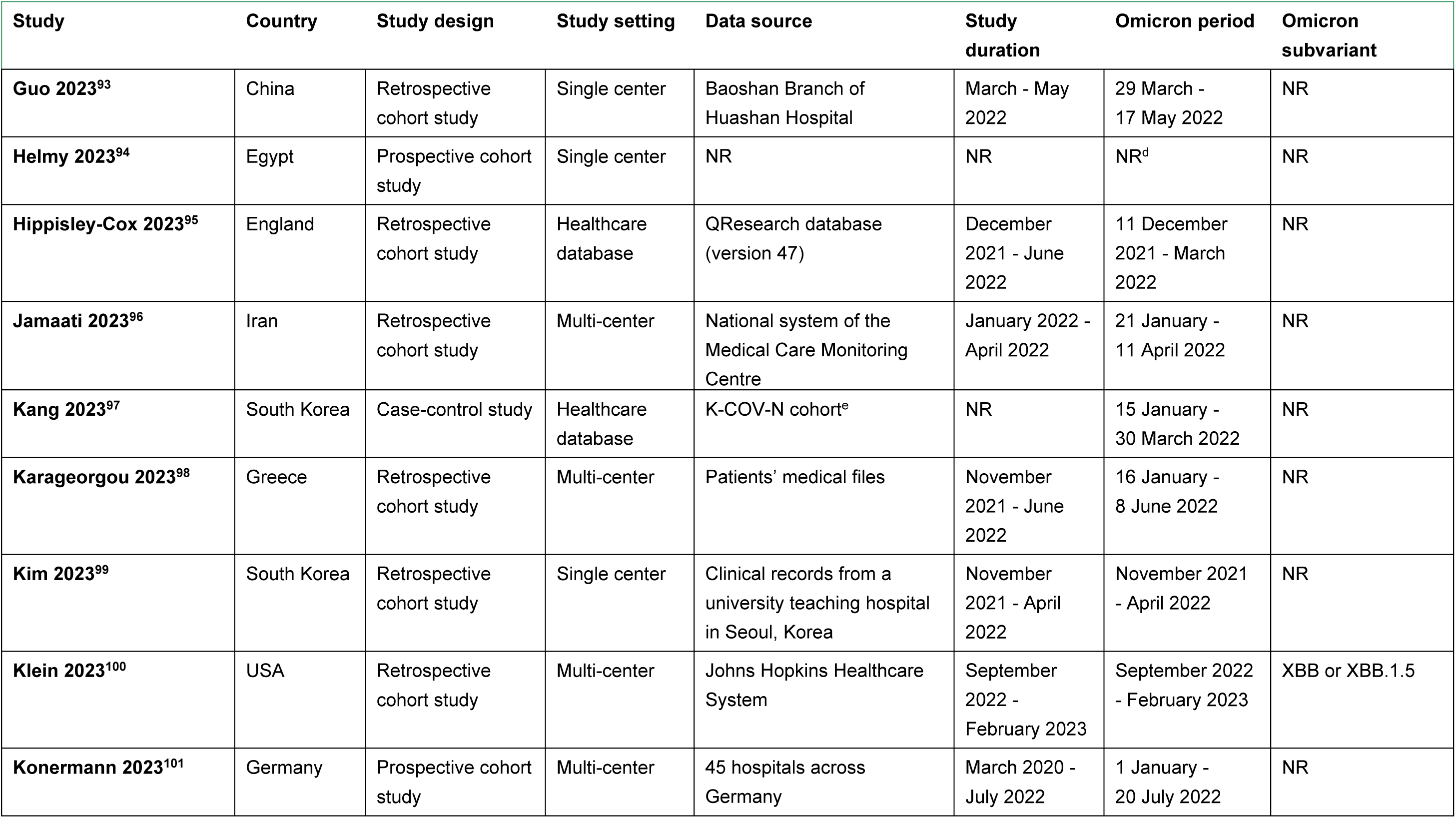
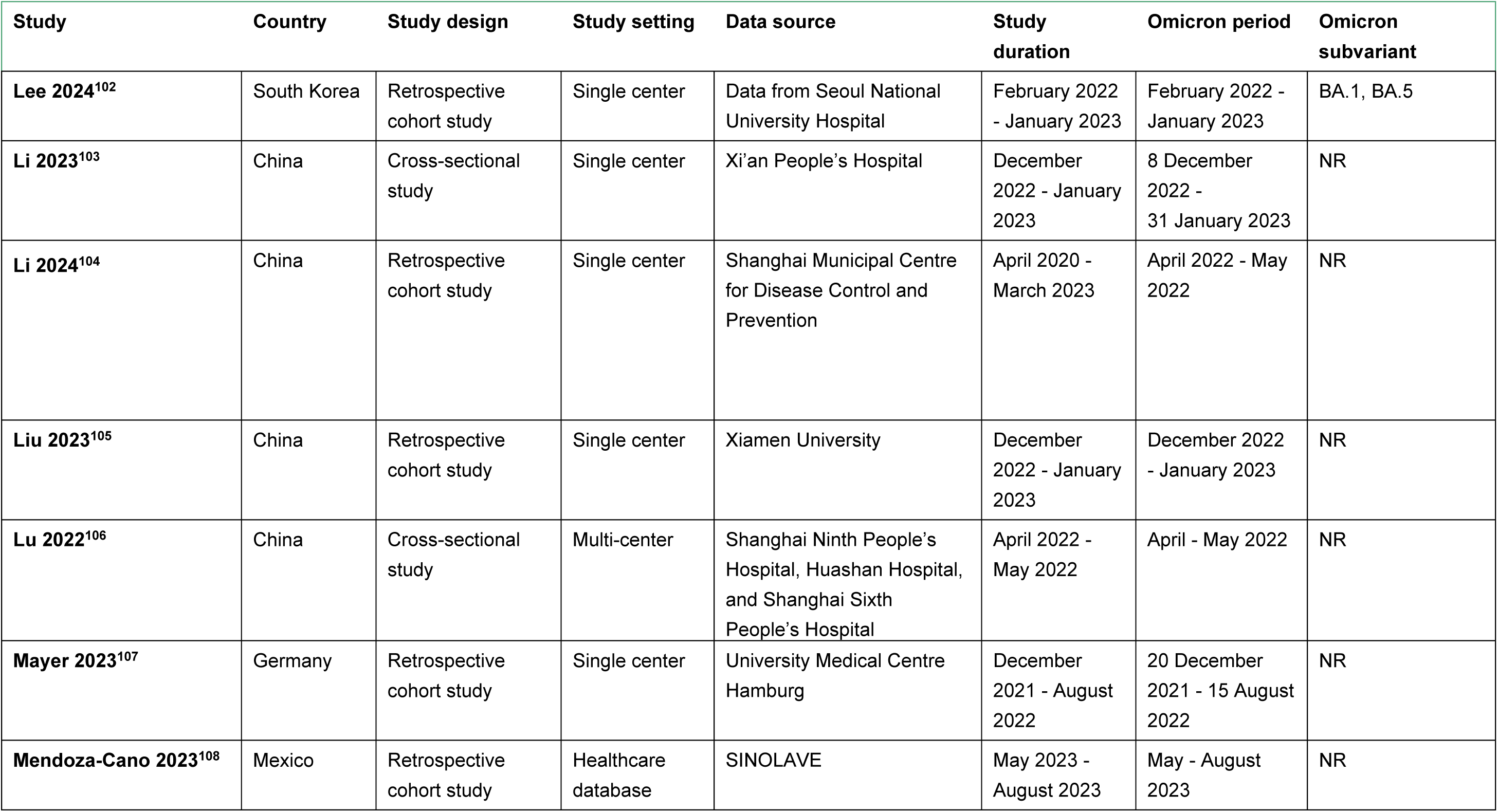
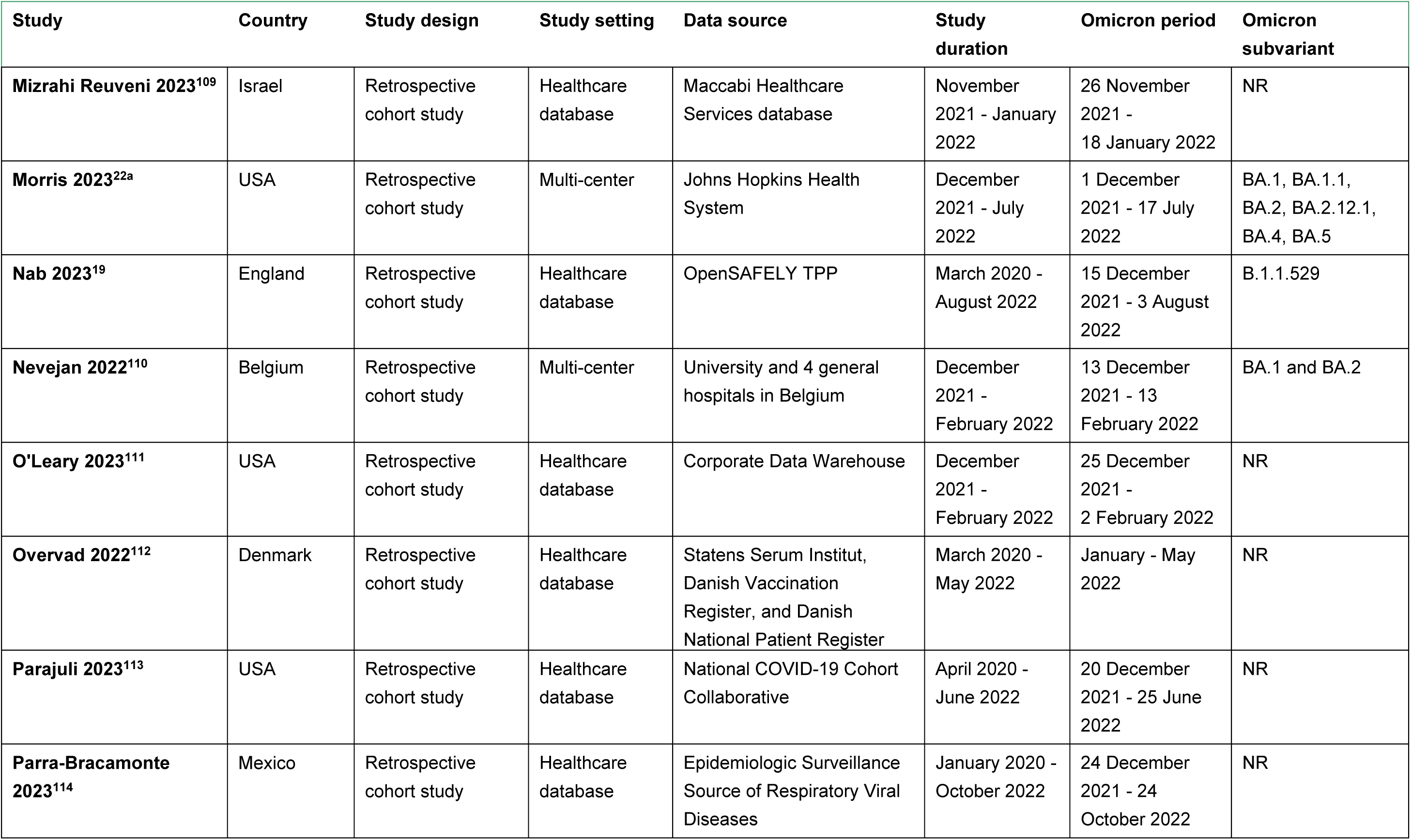
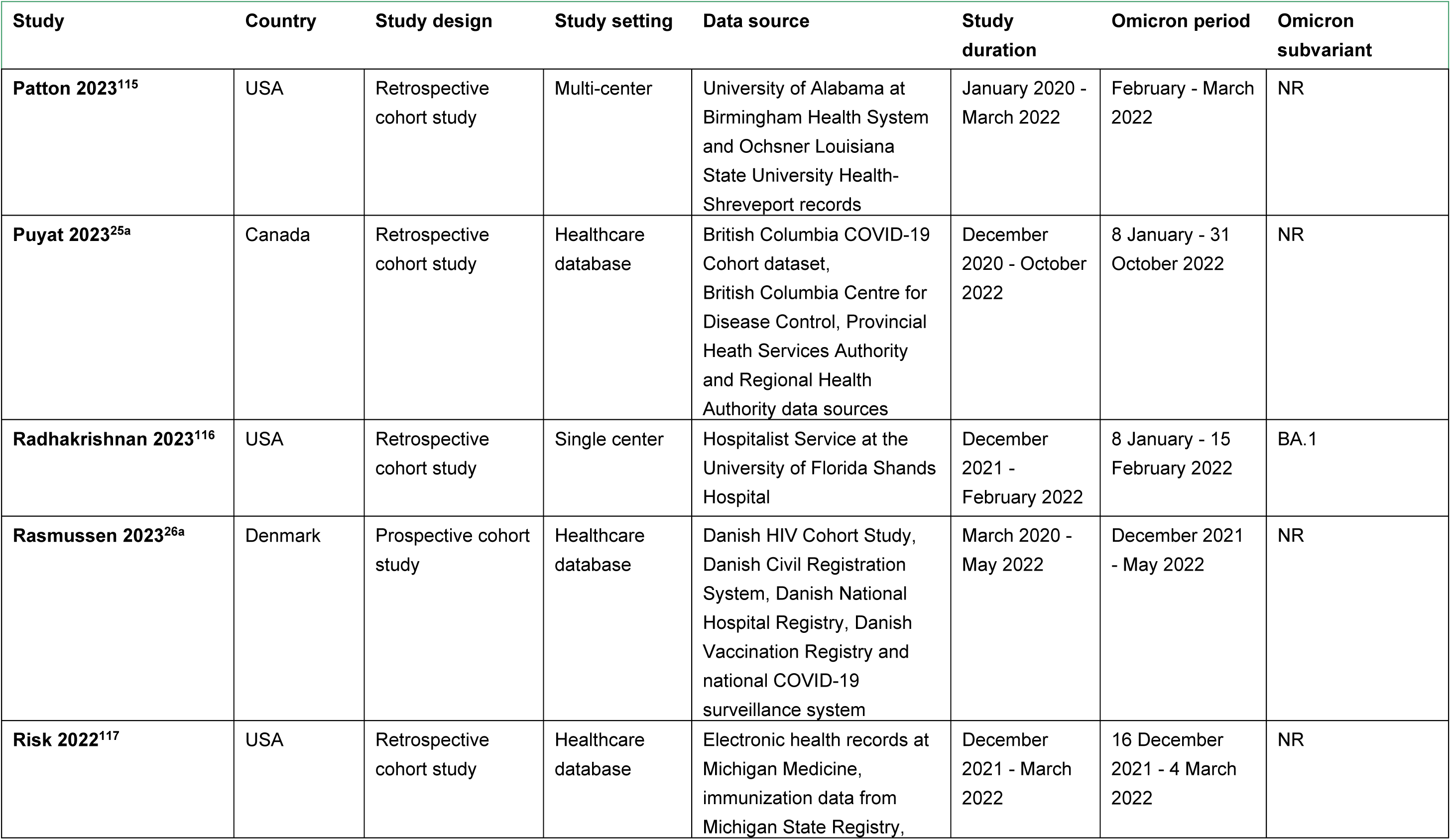
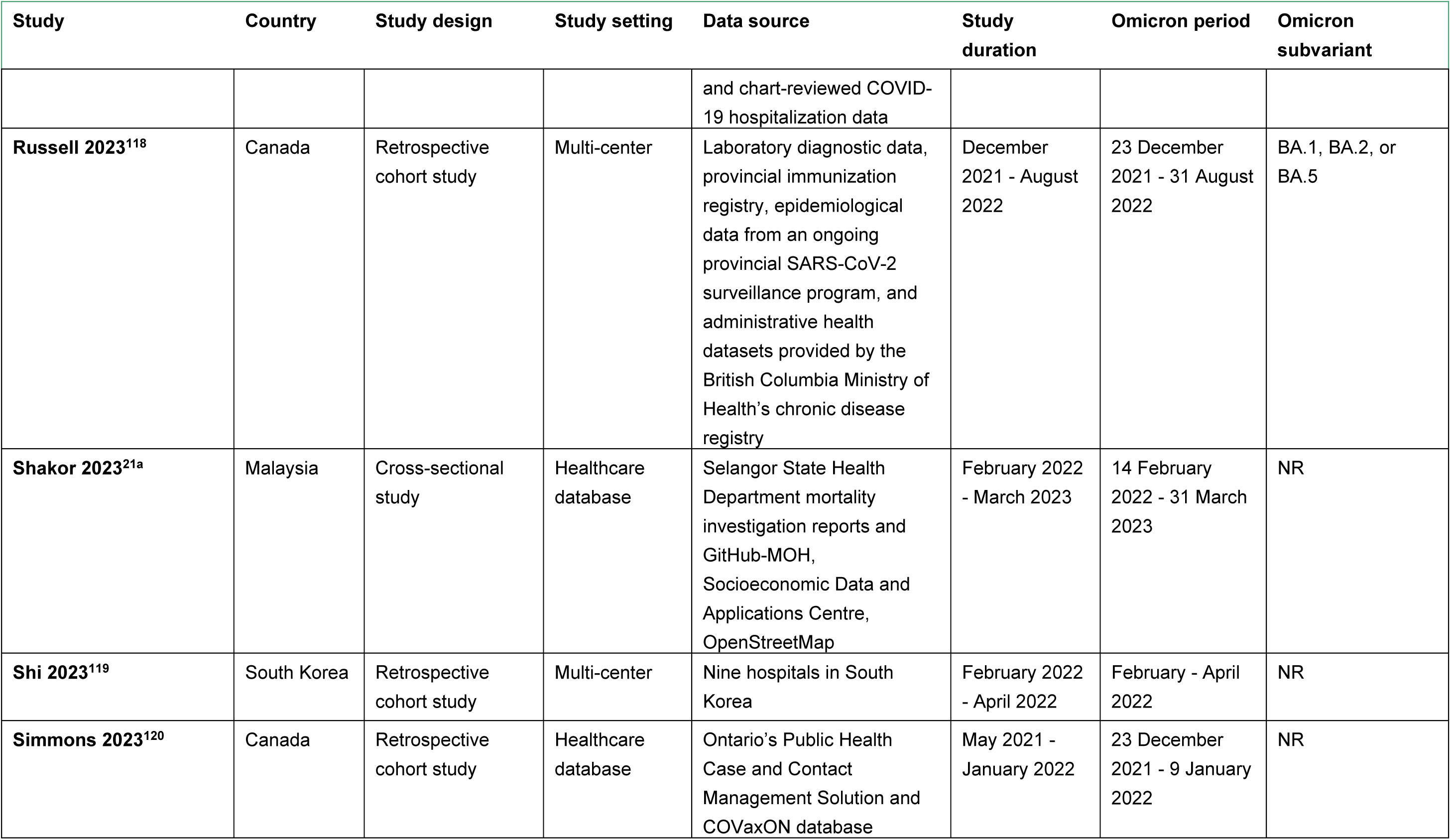
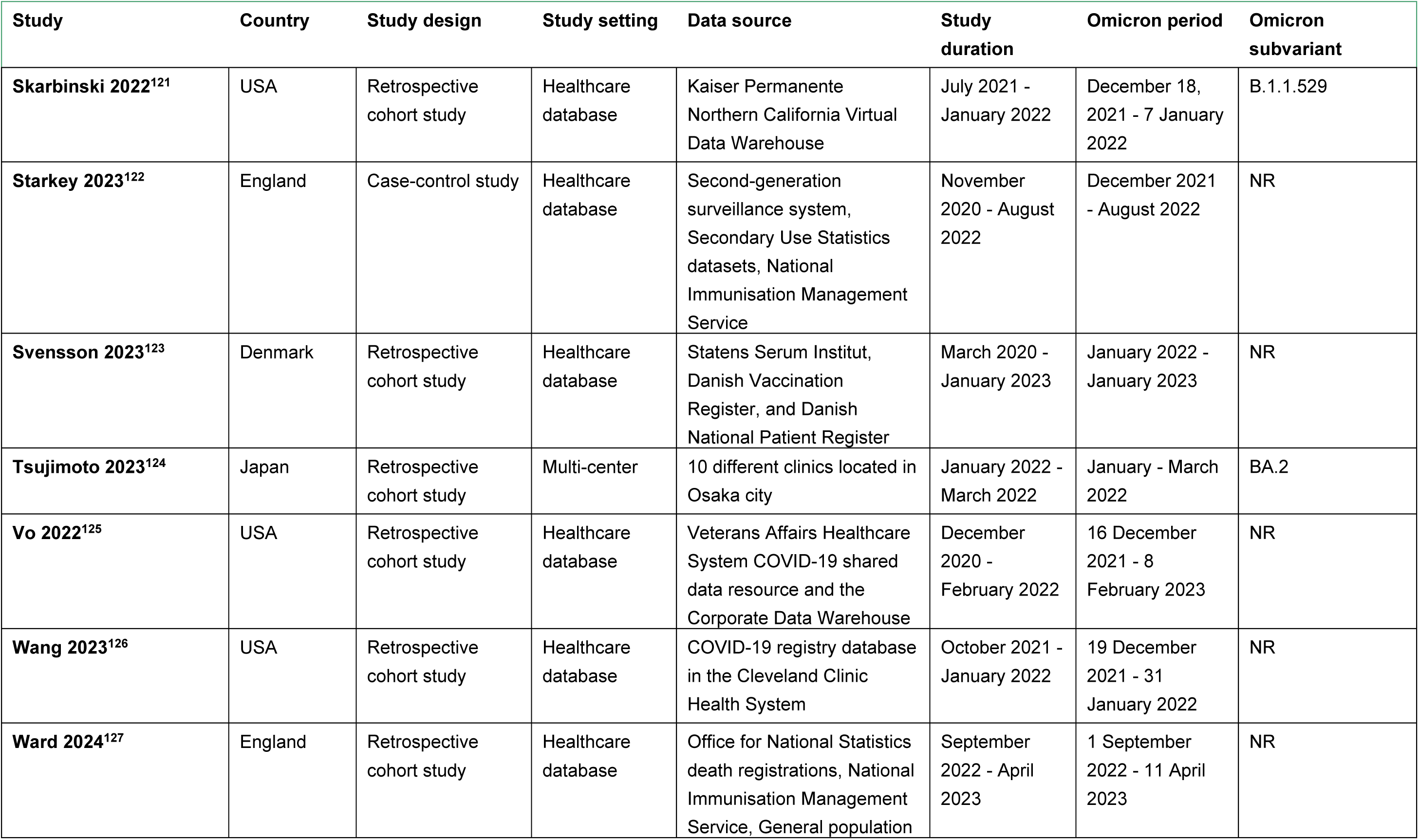
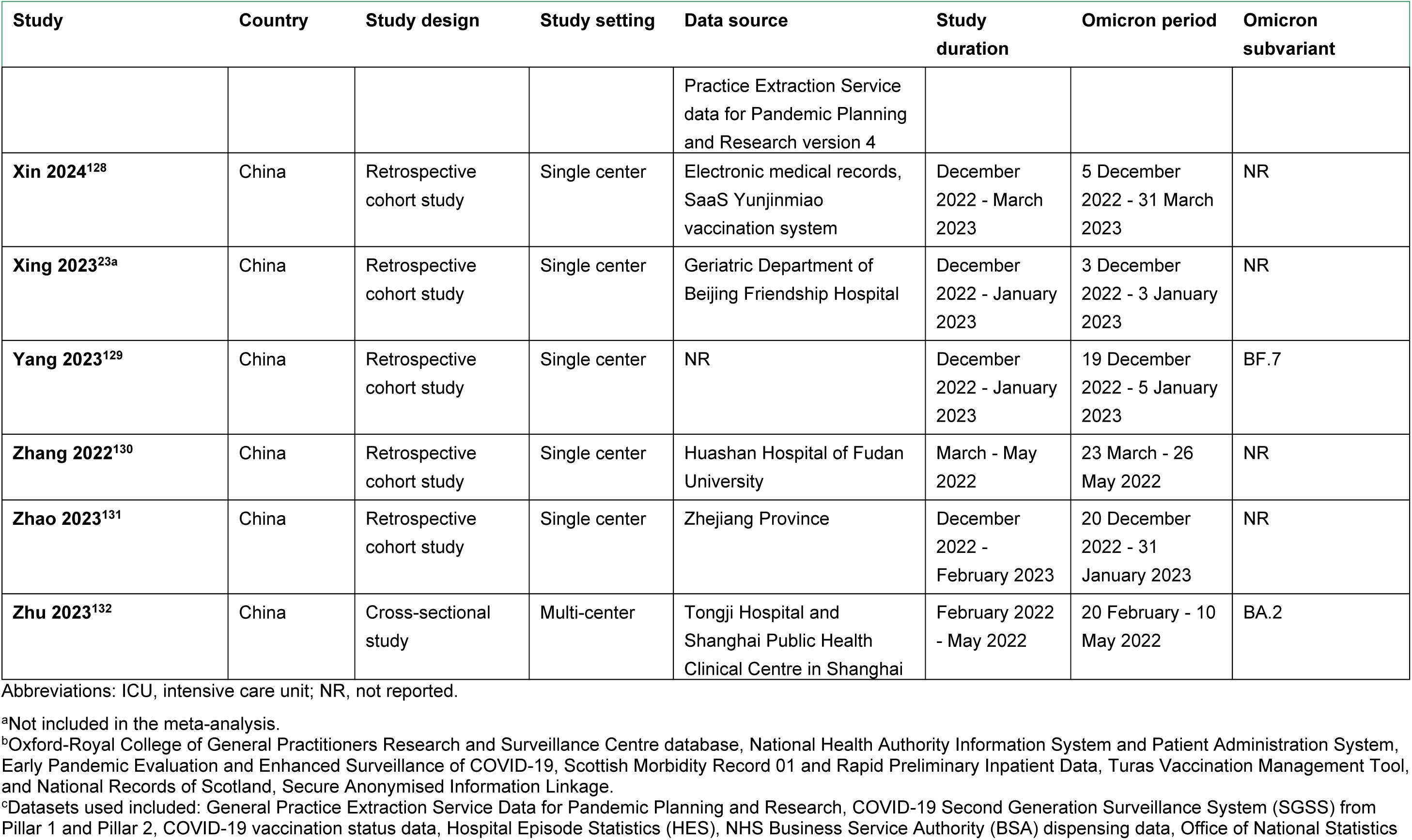
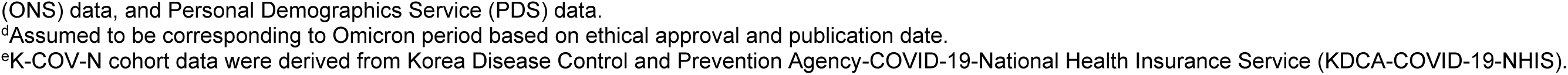
Characteristics of the Included Studies.

### Participant Characteristics

Most studies (n = 37) included a ‘Hospitalized Population’, which refers to people who were already hospitalized at the start of the study. The rest of the studies (n = 35) included a ‘General Population’ (not hospitalized at the start of the study) (**Table 2**). Minimum numbers of participants included in the analyses for each IC/IS condition are reported in **Table 3**. People with renal disease were the most commonly reported group (n = 48 studies), and people with HIV were the least commonly reported (n = 5 studies) (**Table 3**). Though 16 studies did not report vaccination status, the majority of studies (>50%) that reported vaccination status included fully vaccinated people (defined as those who received the initial vaccination).

**Table 2.**
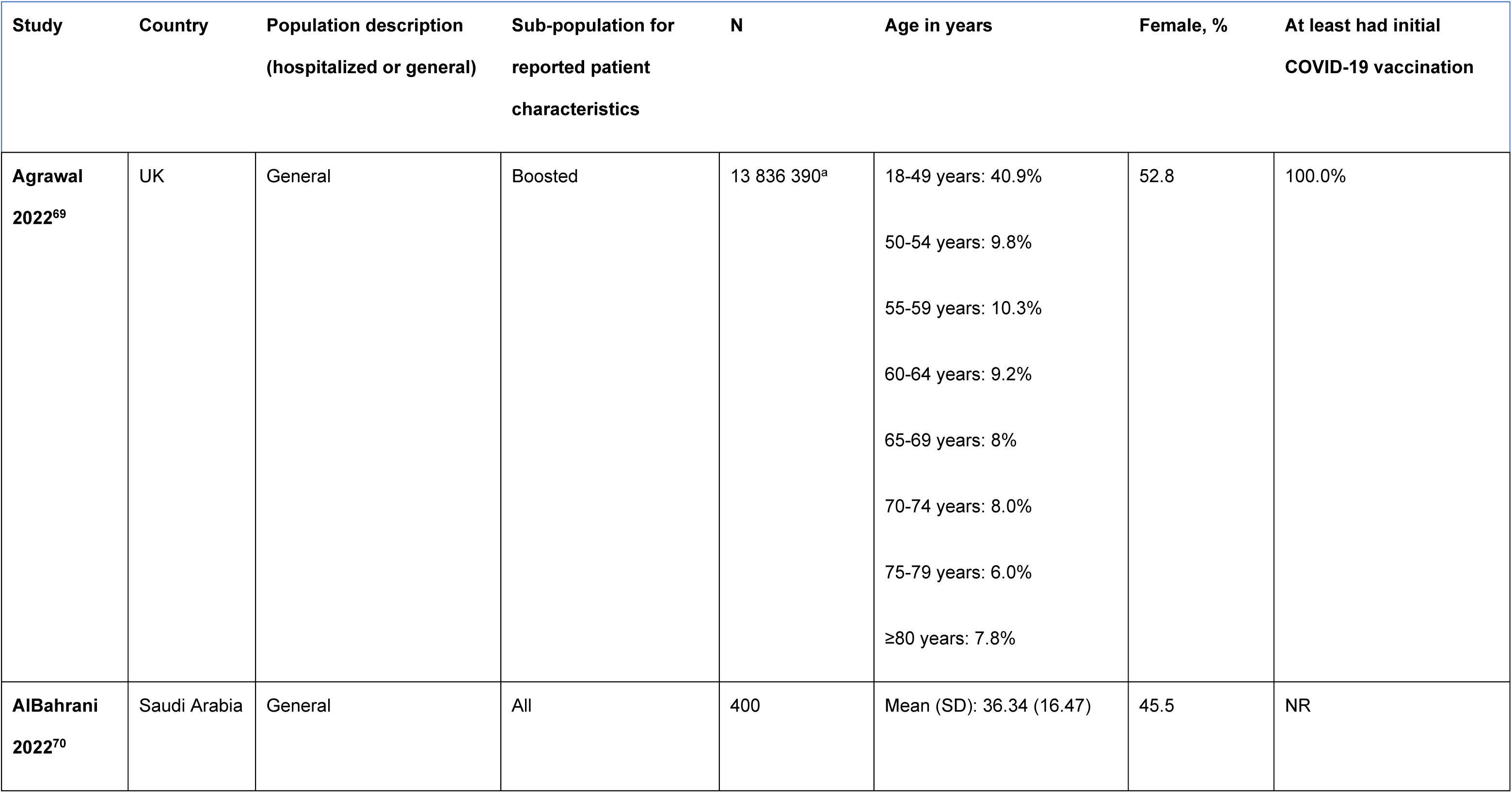
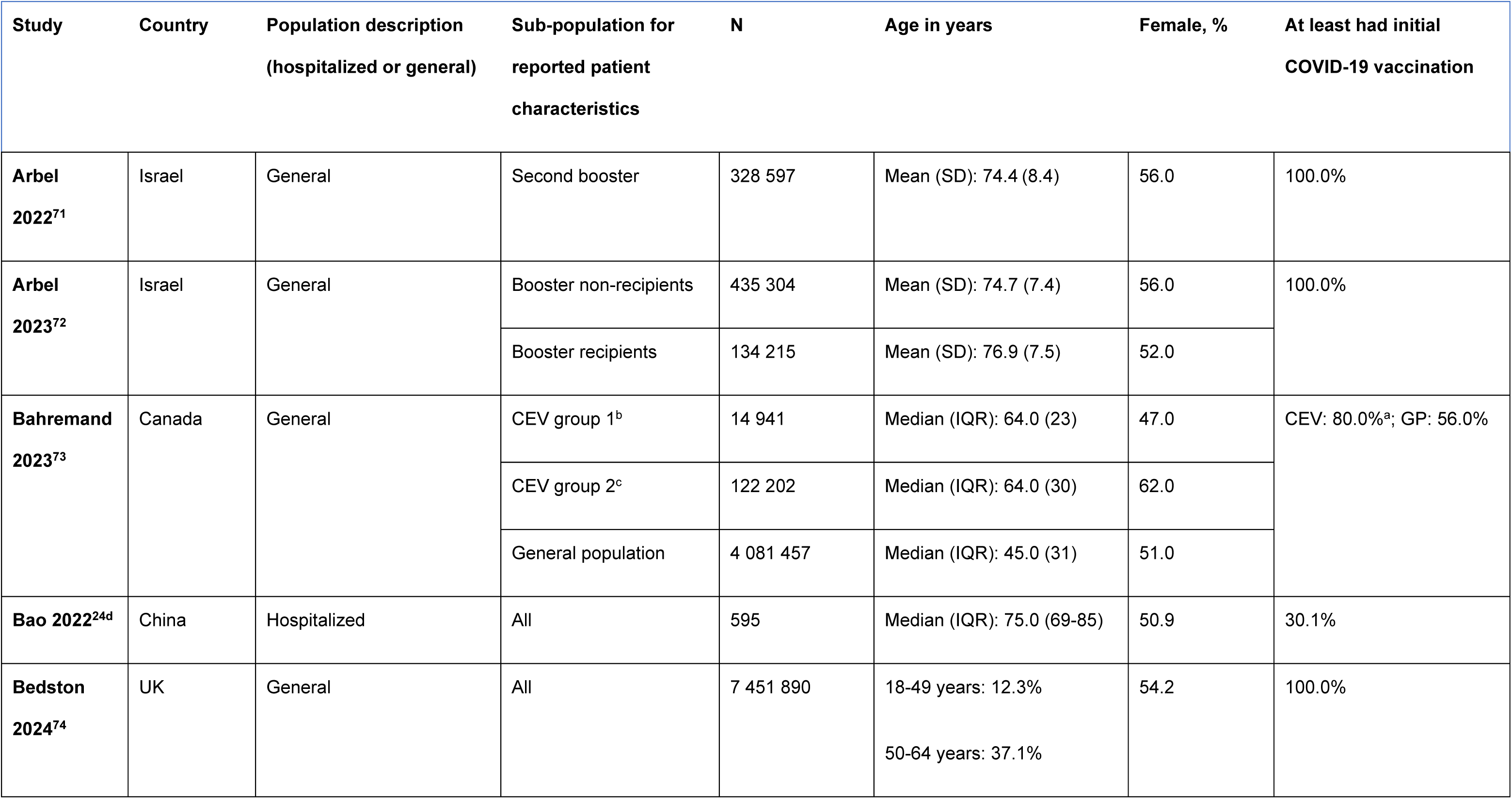
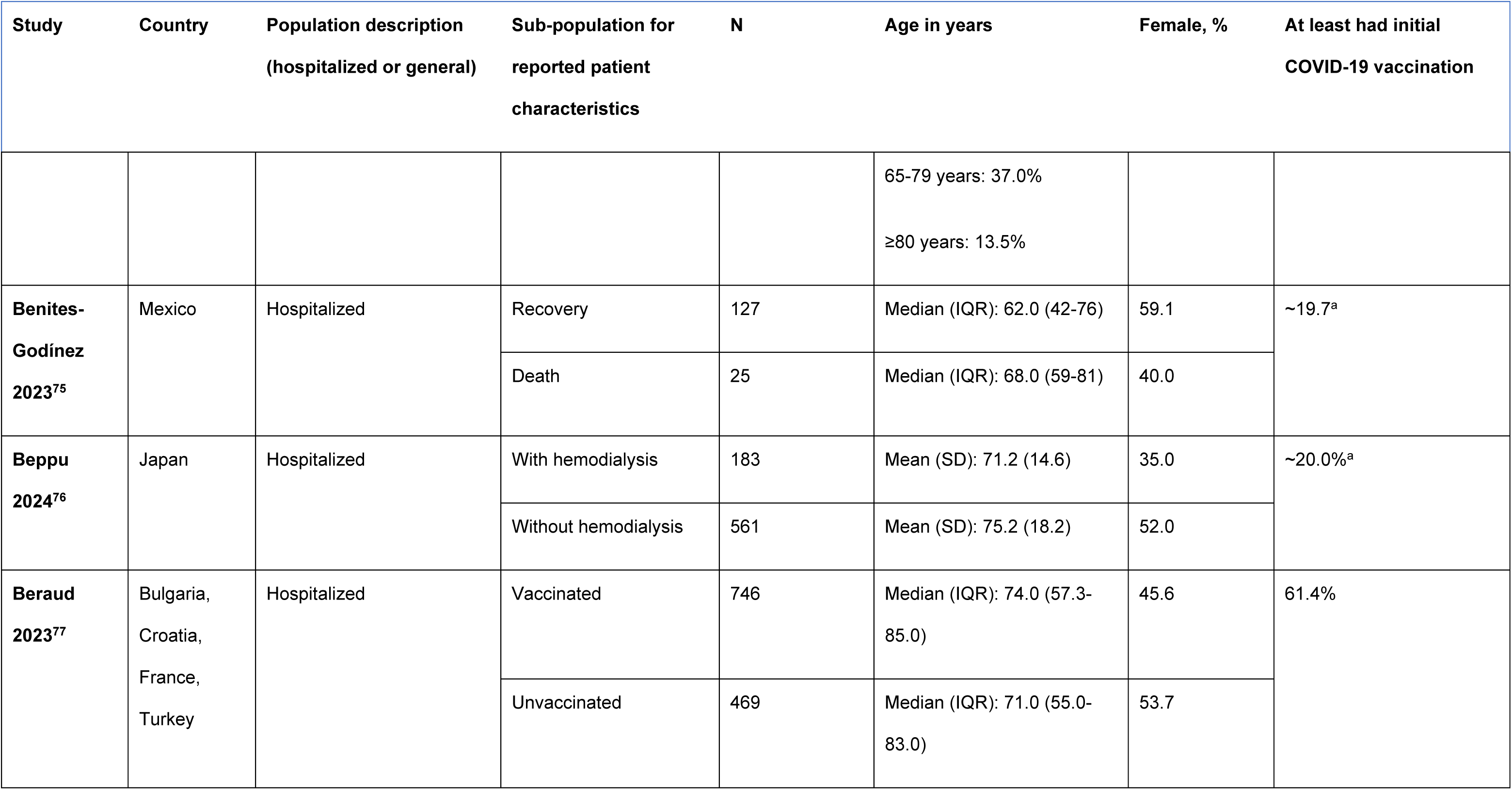
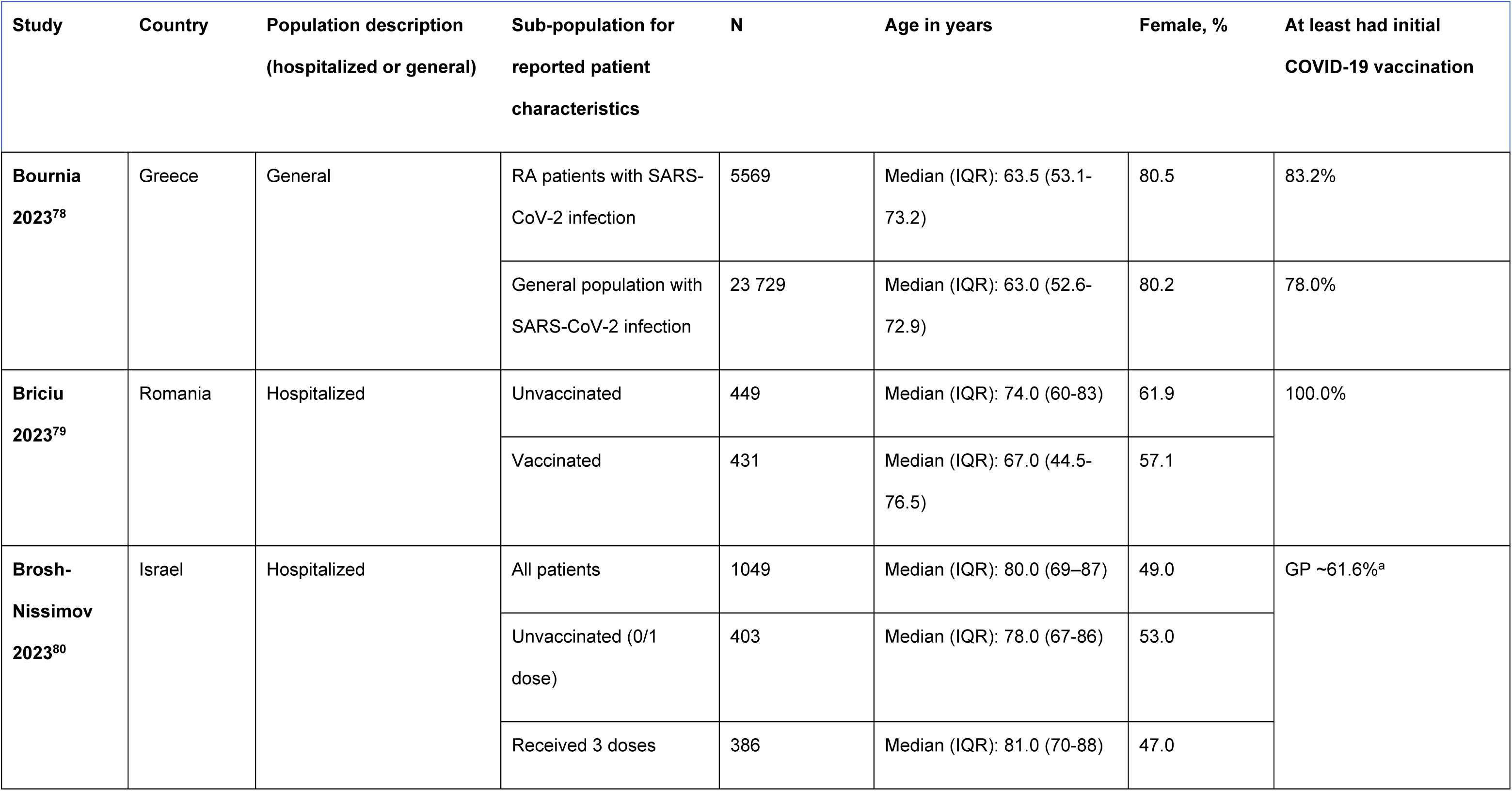
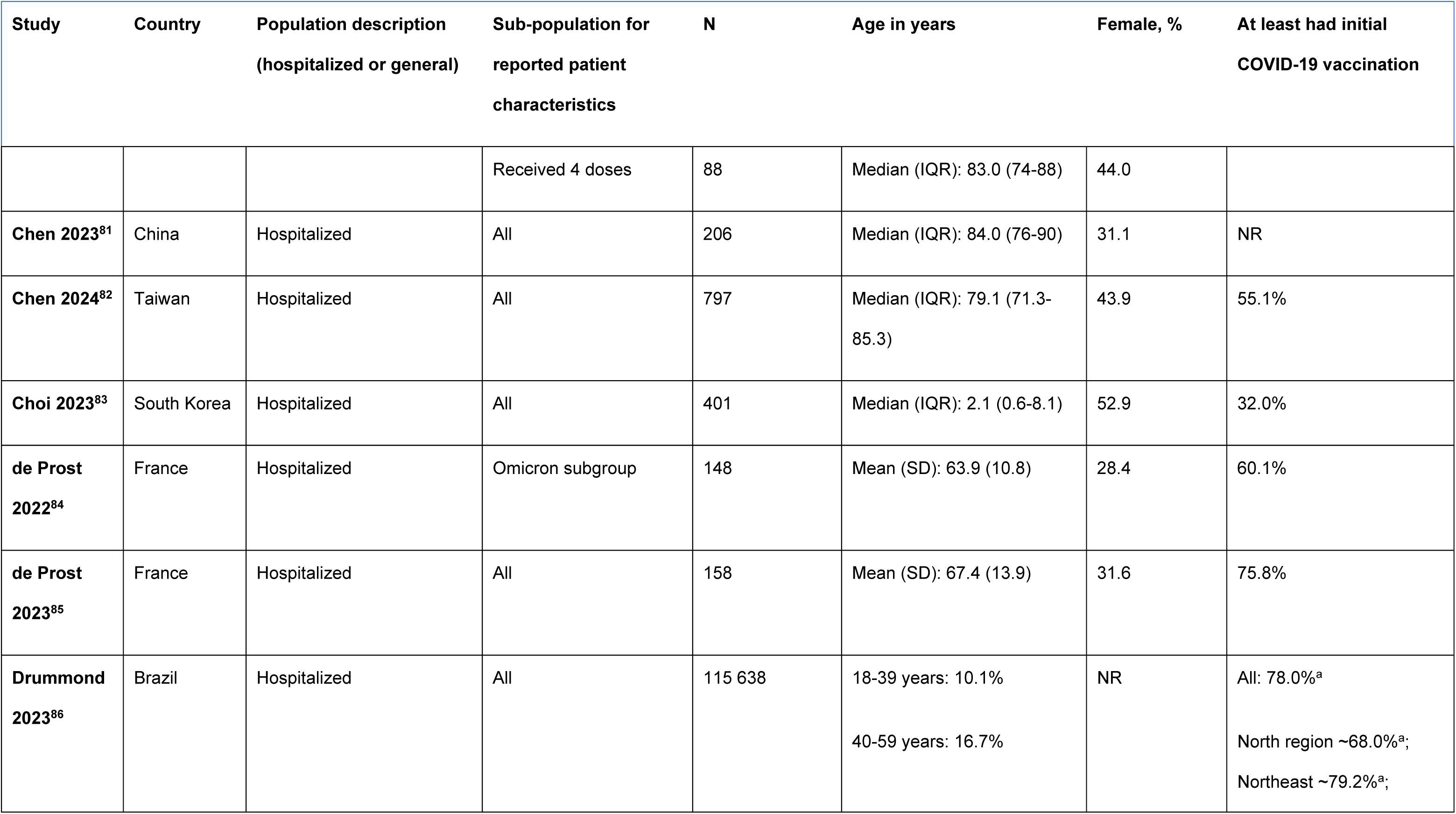
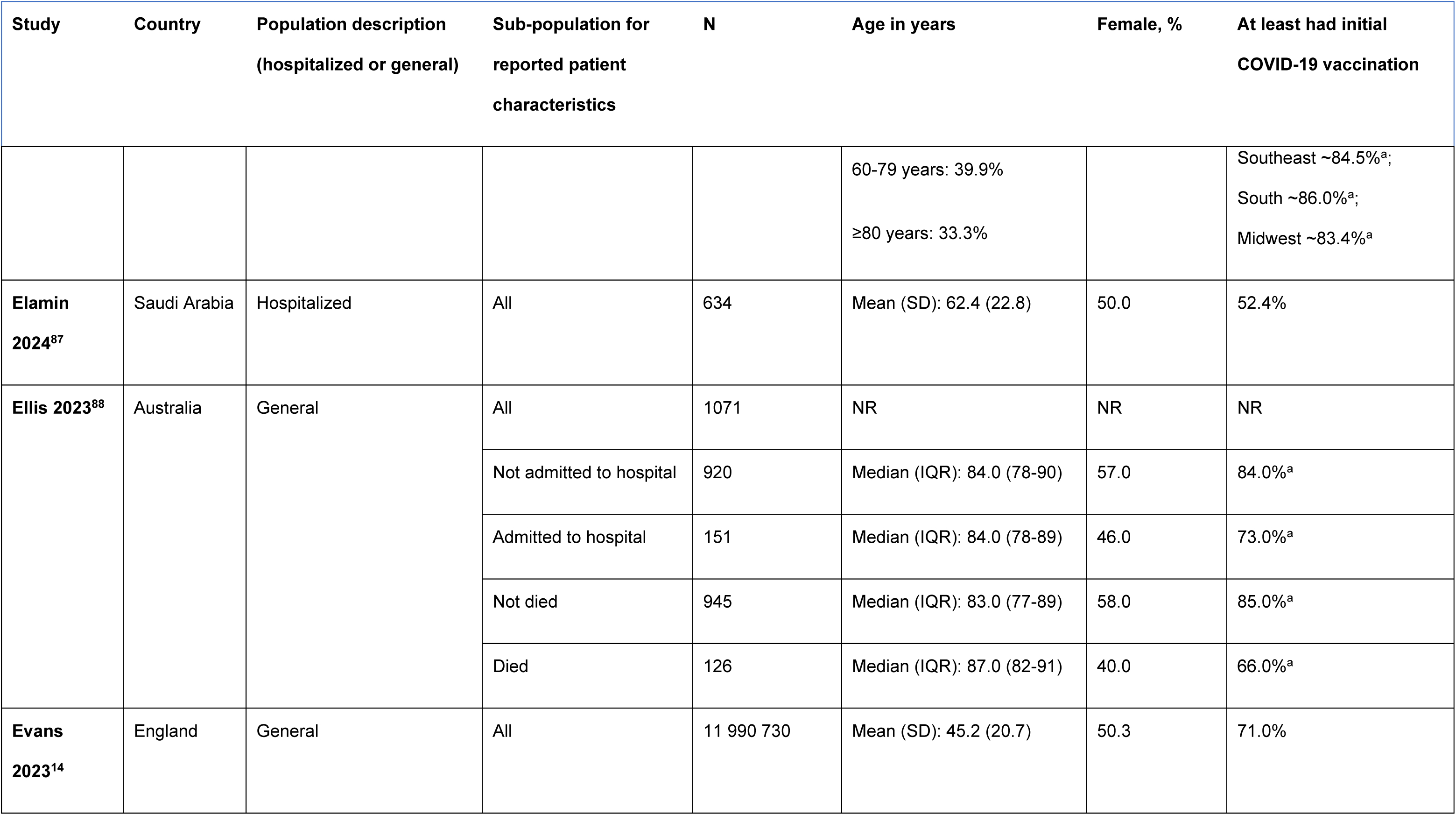
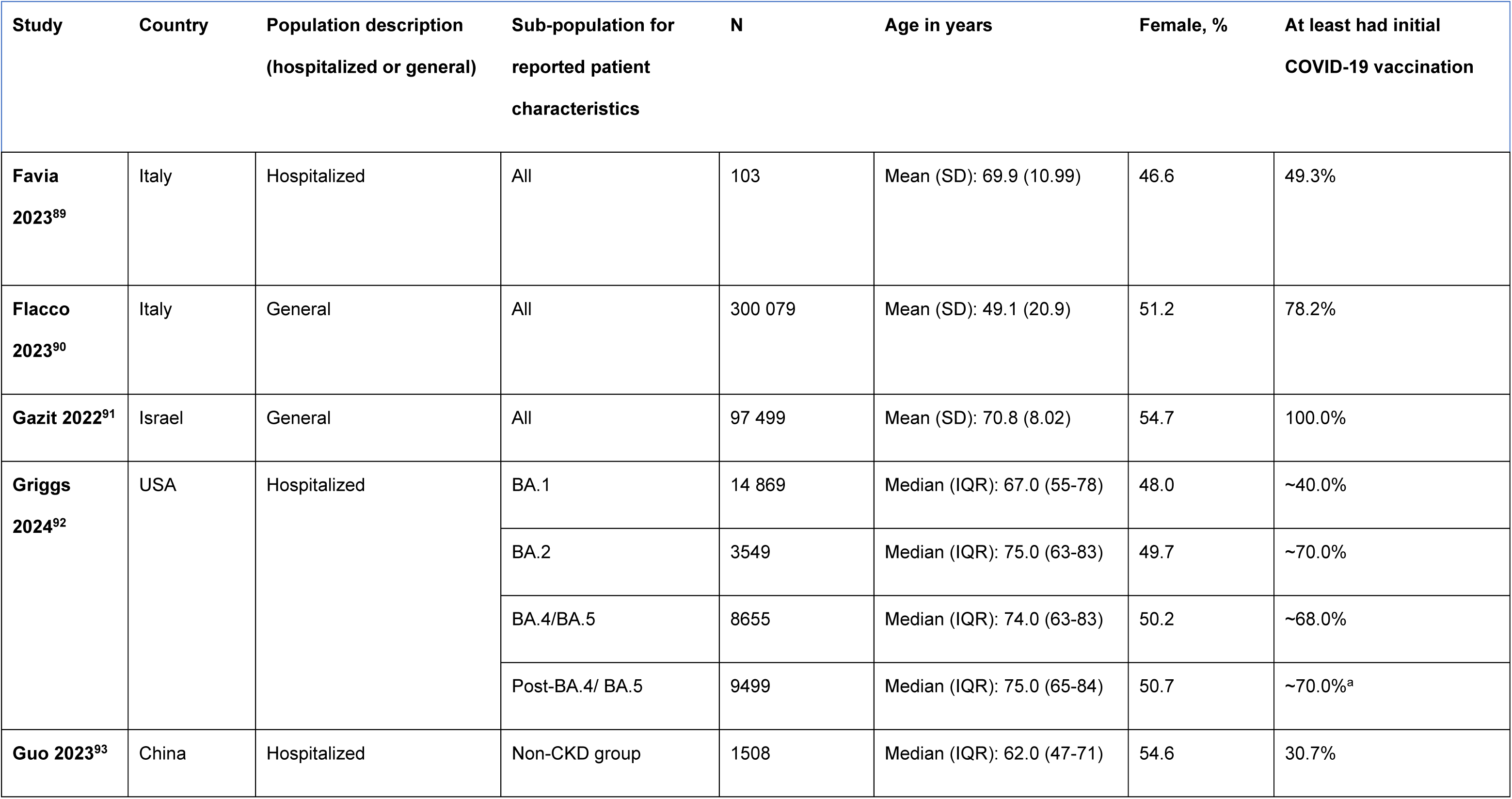
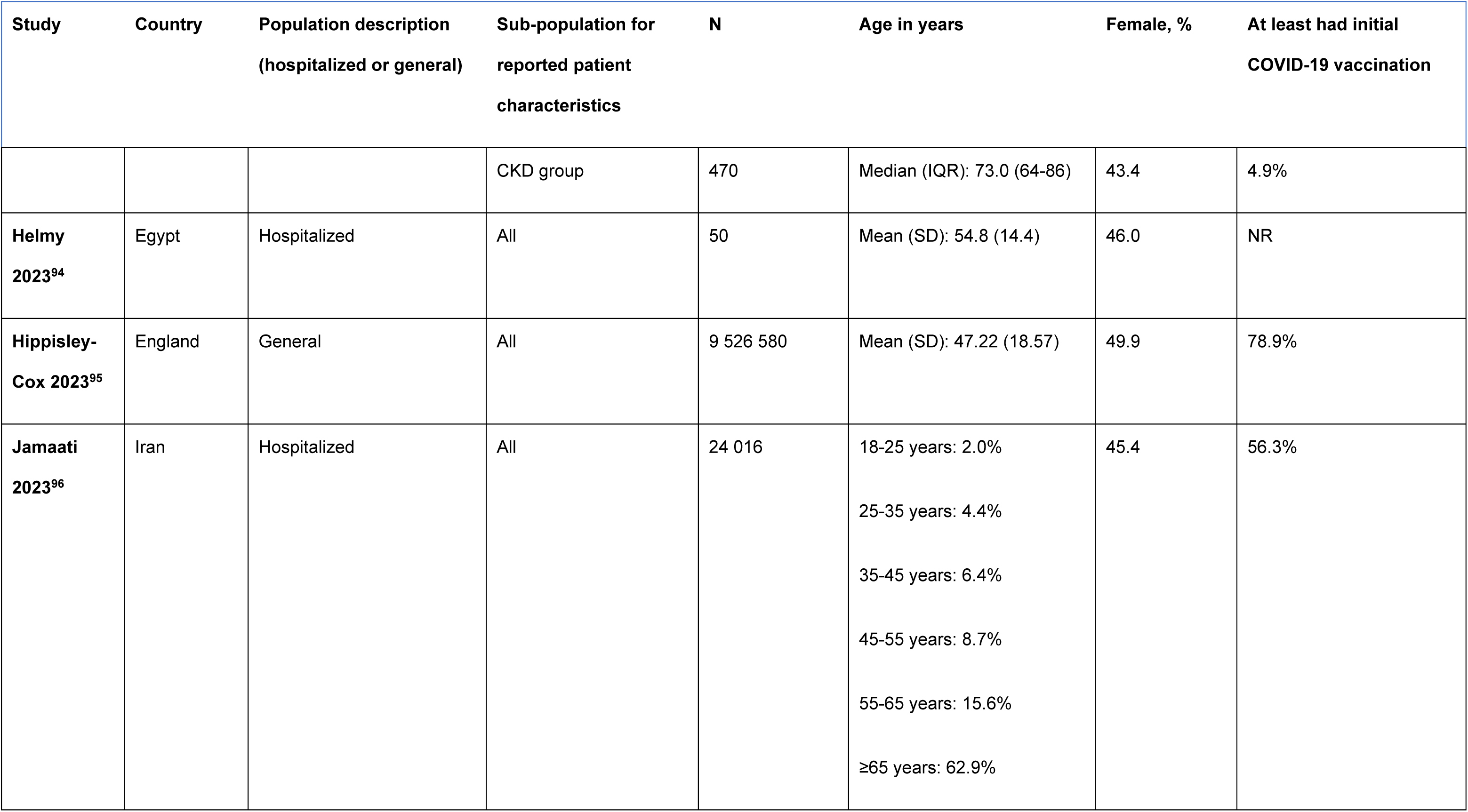
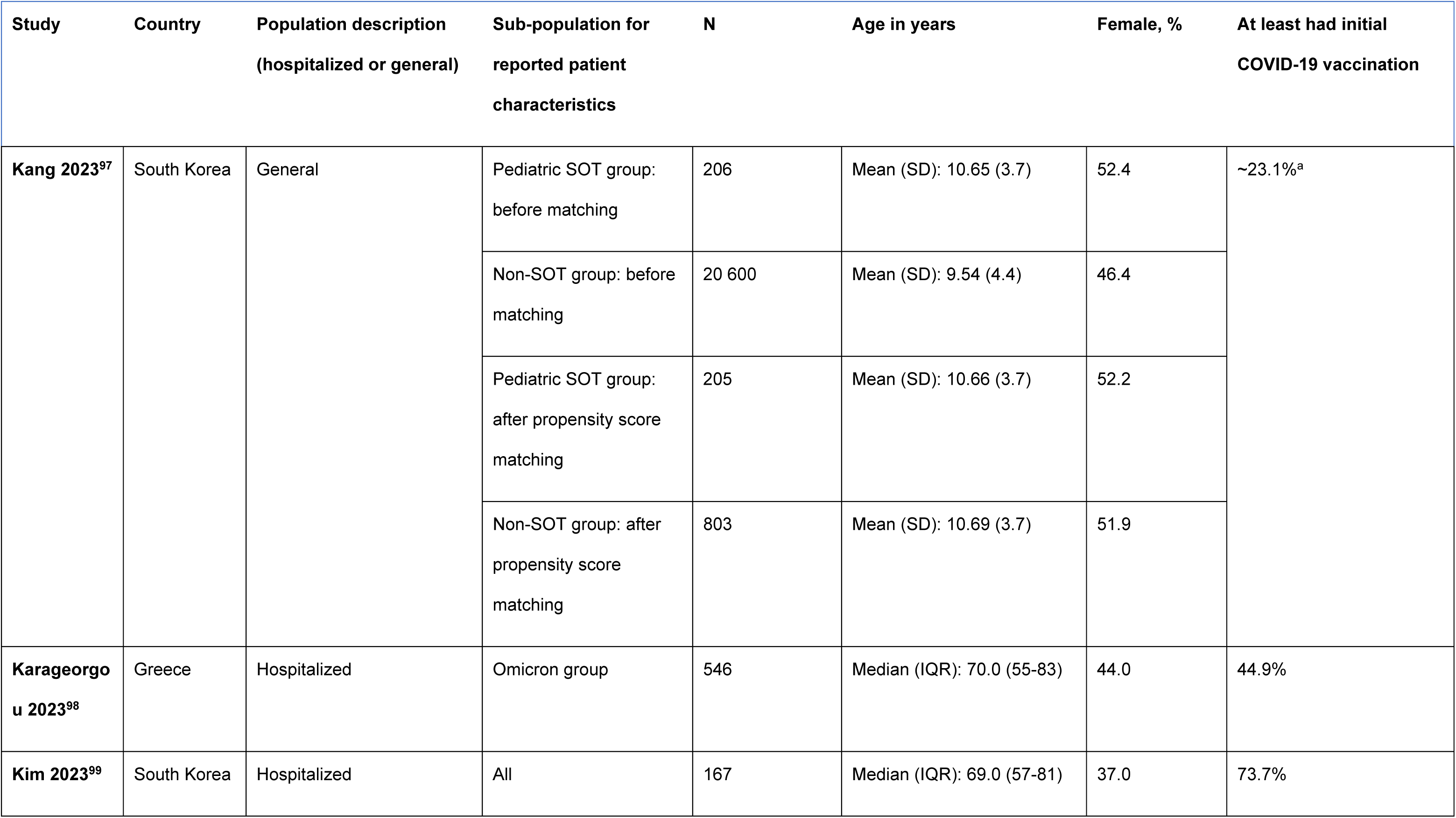
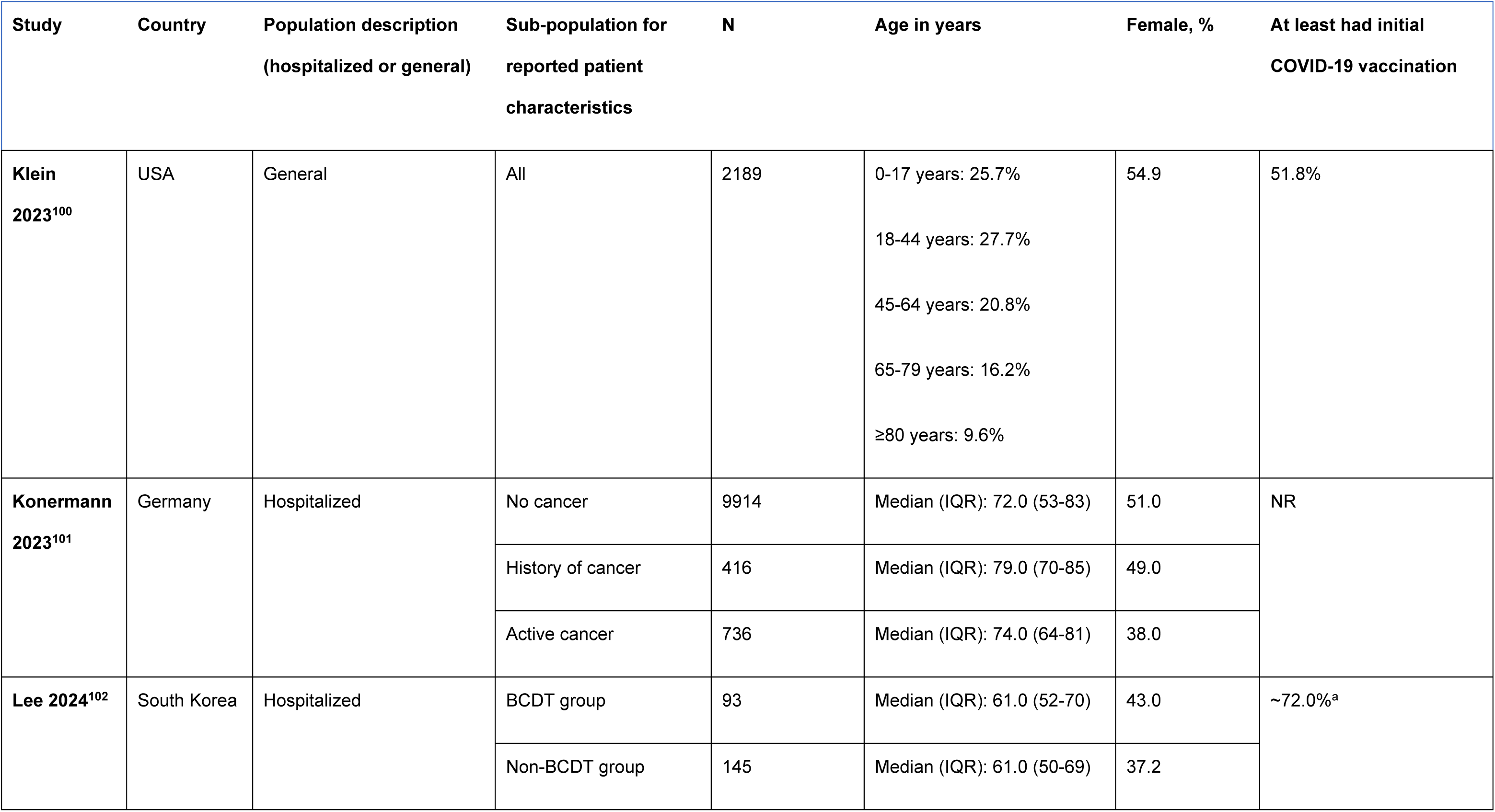
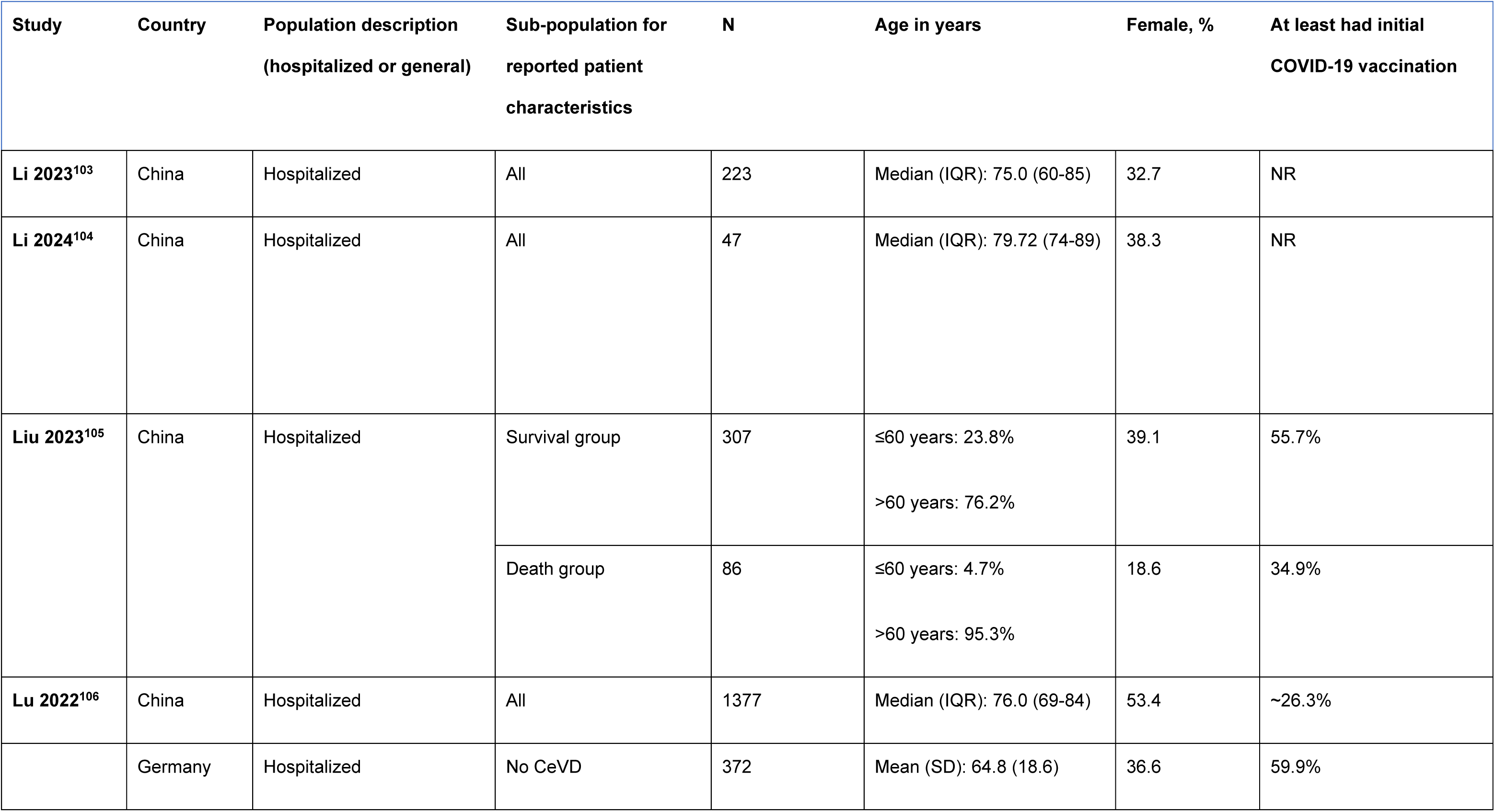
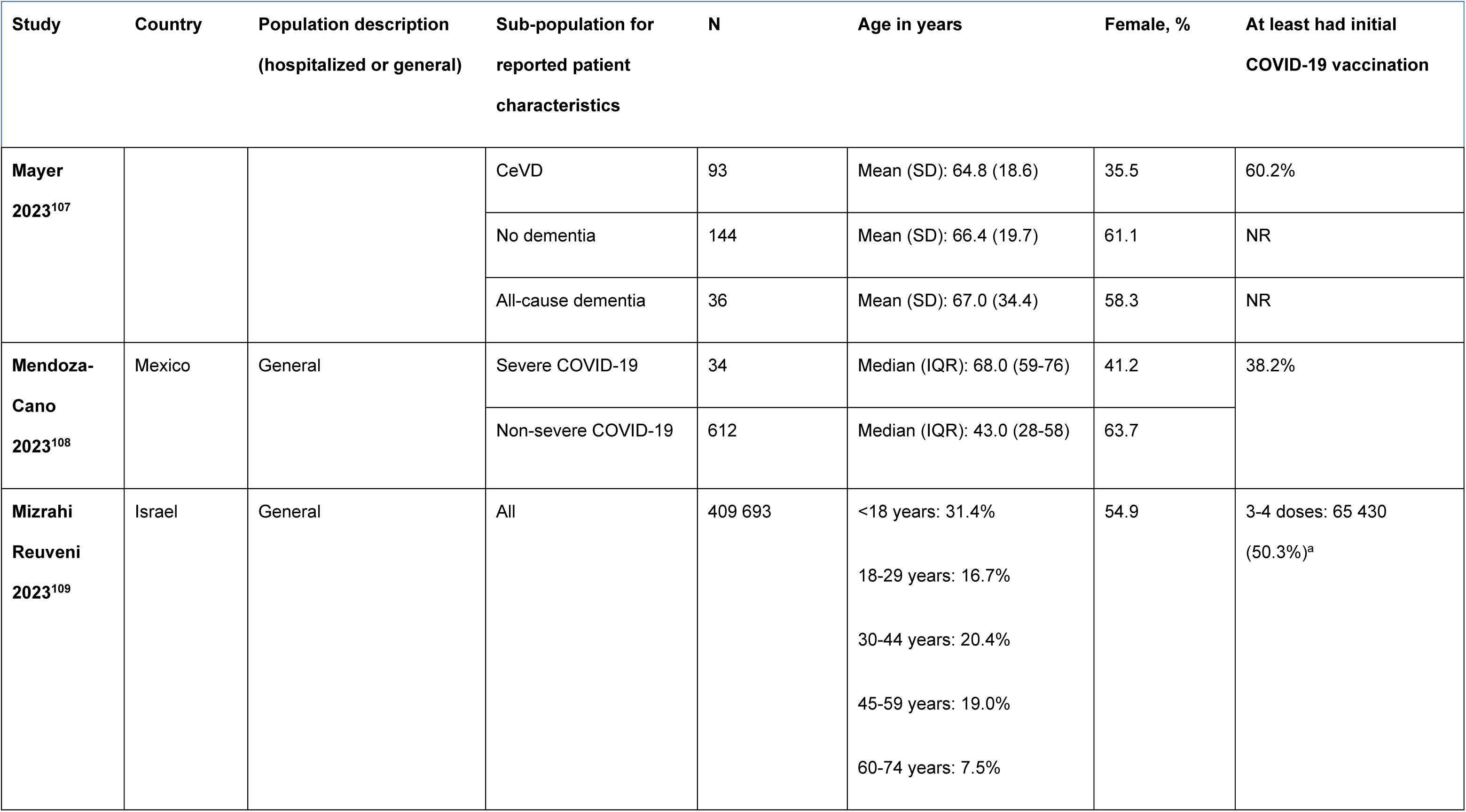
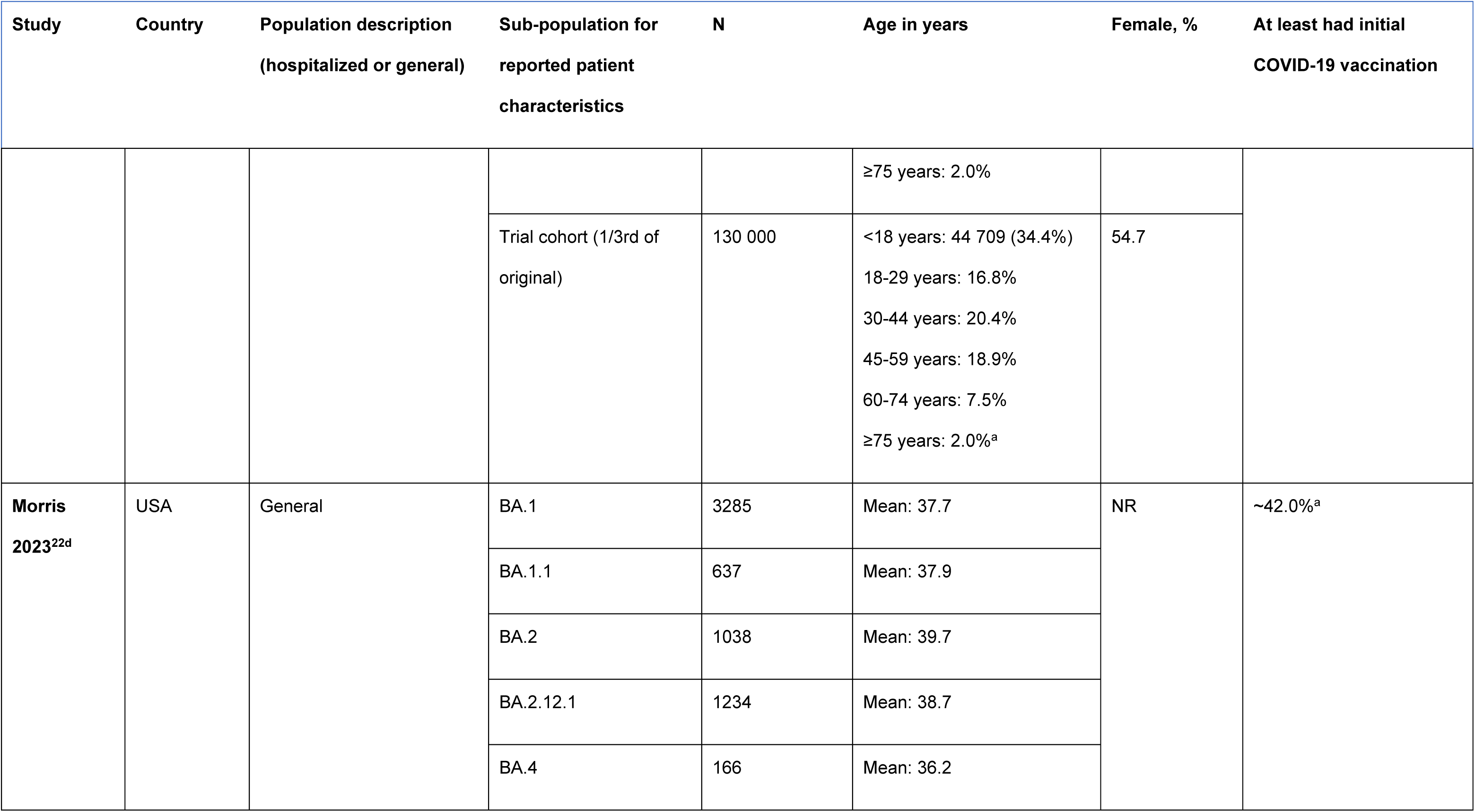
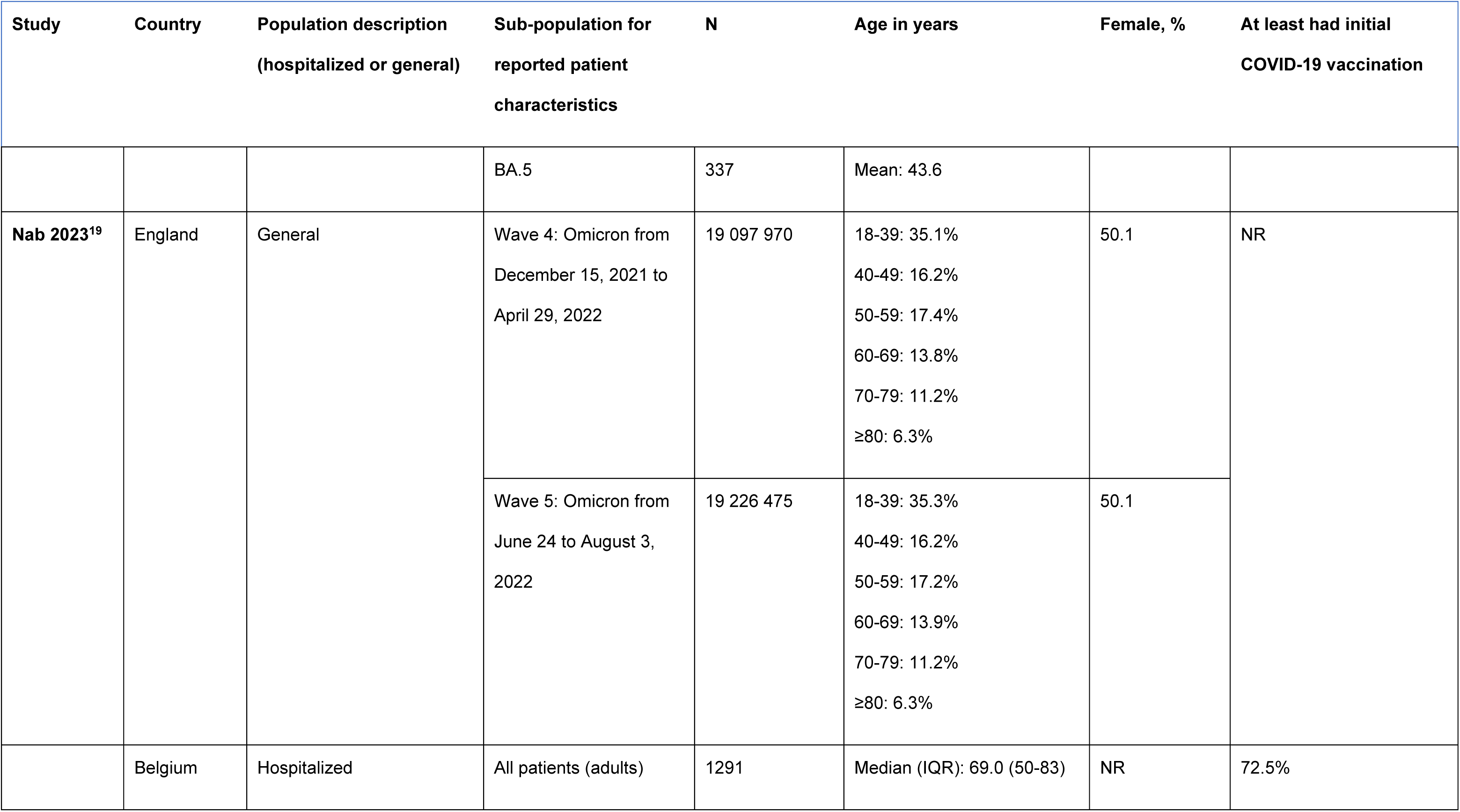
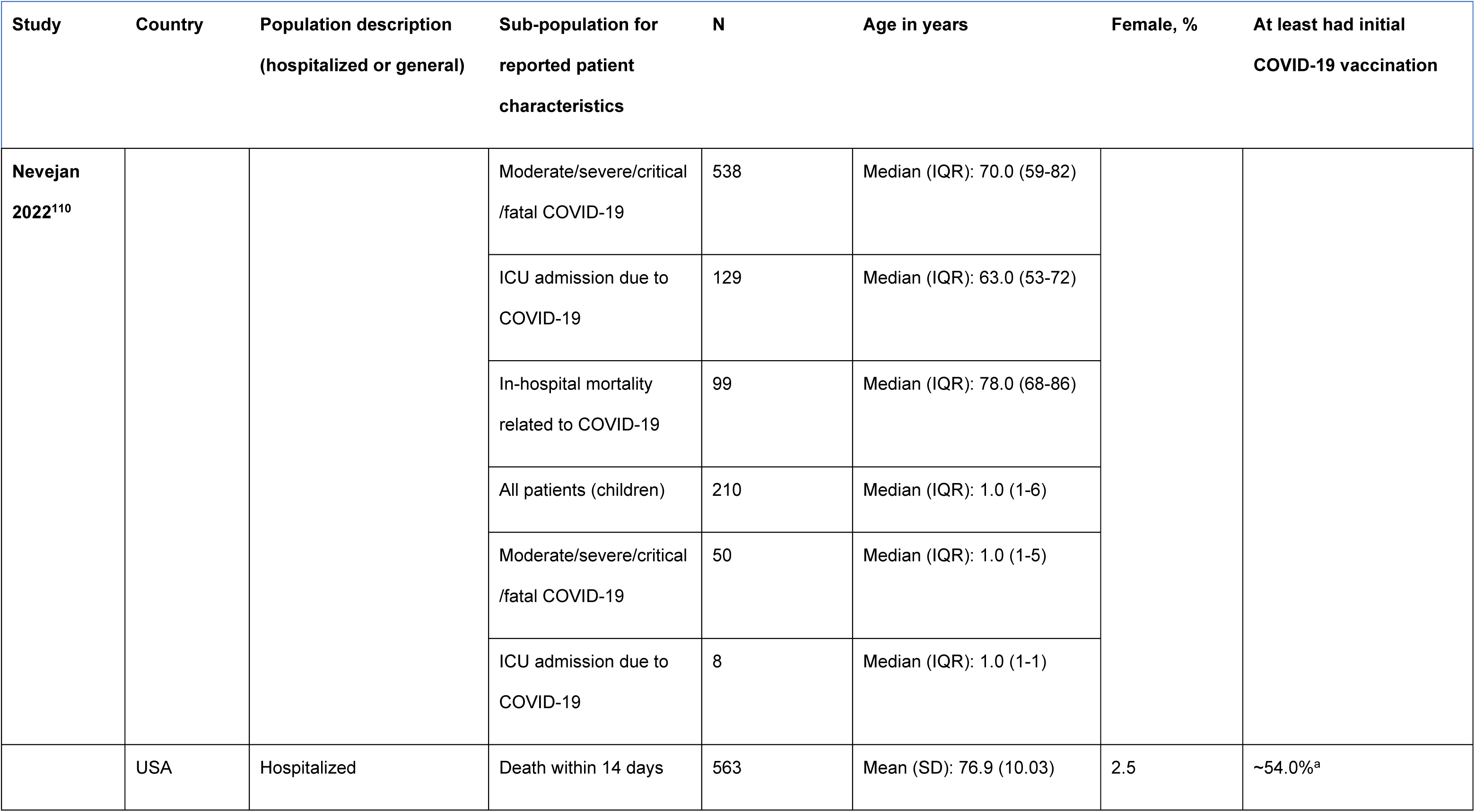
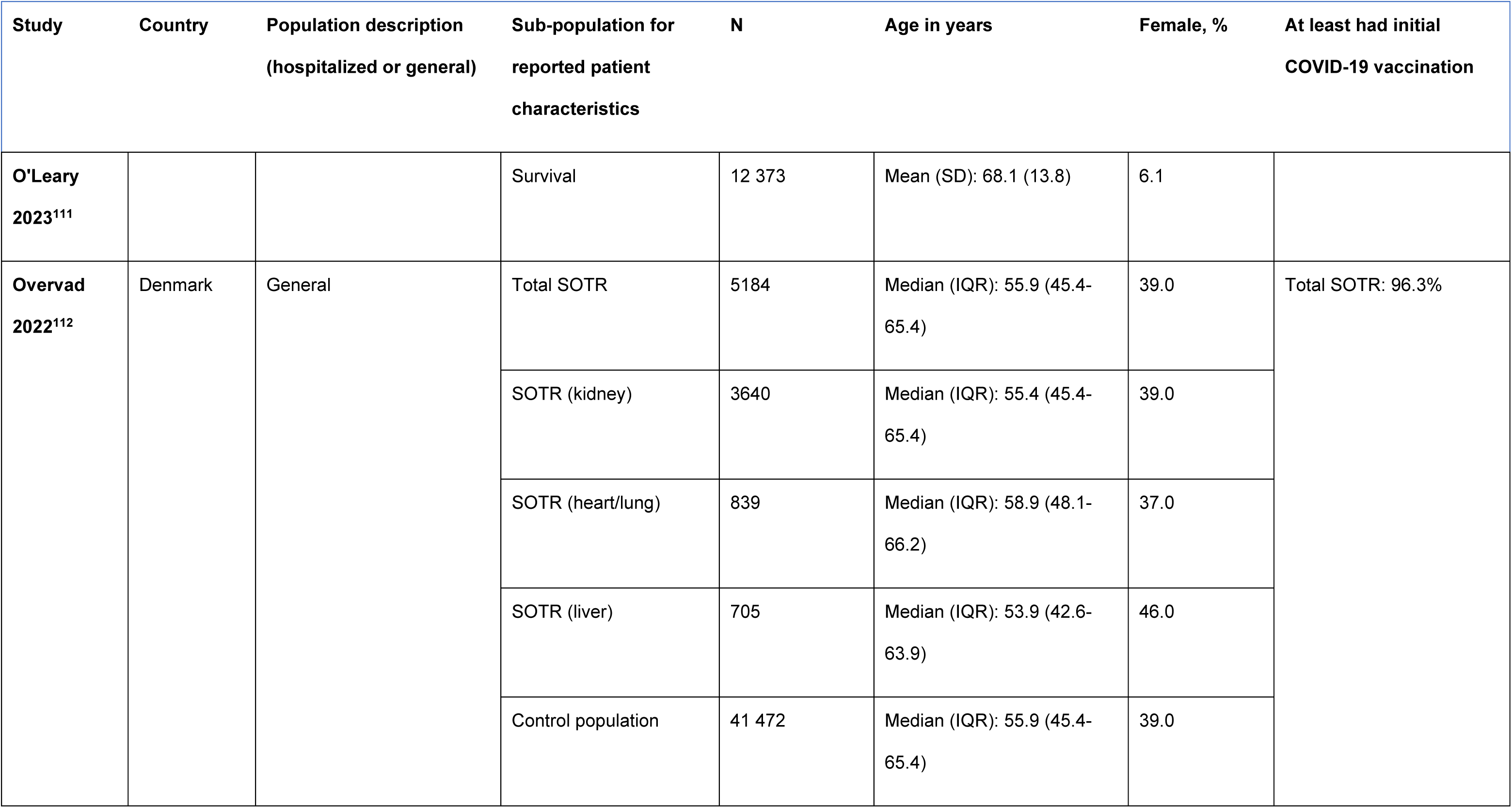
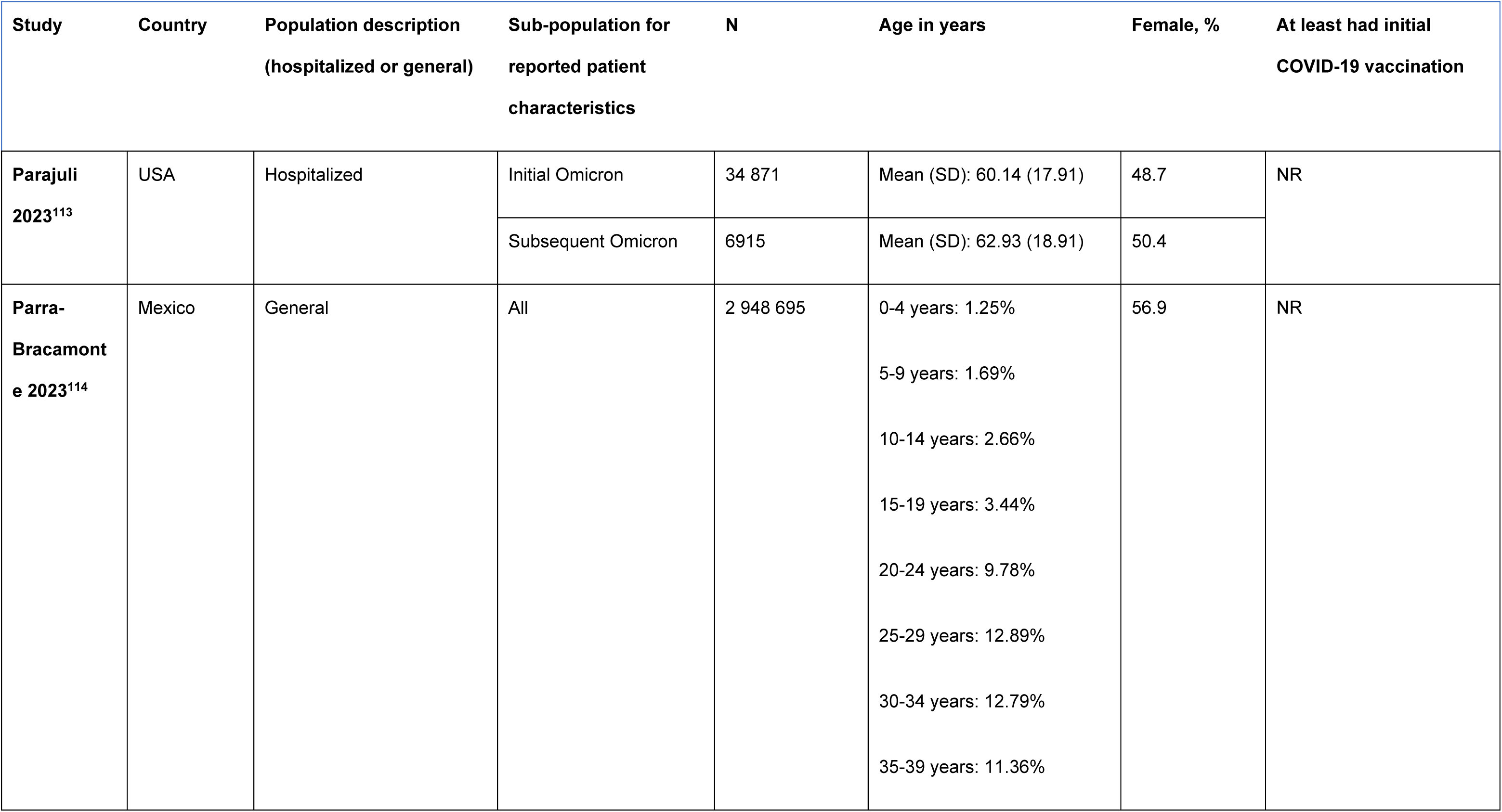
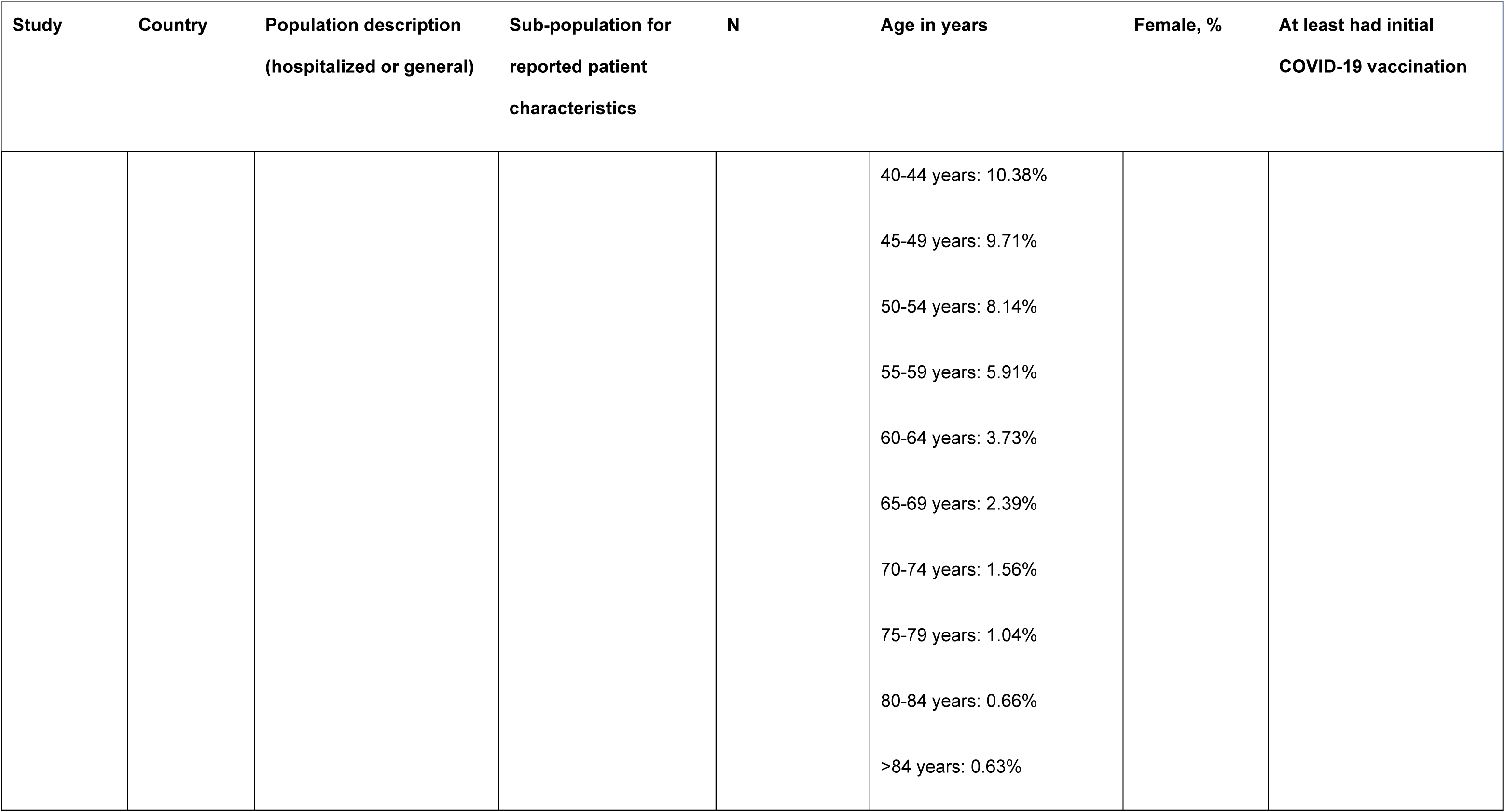
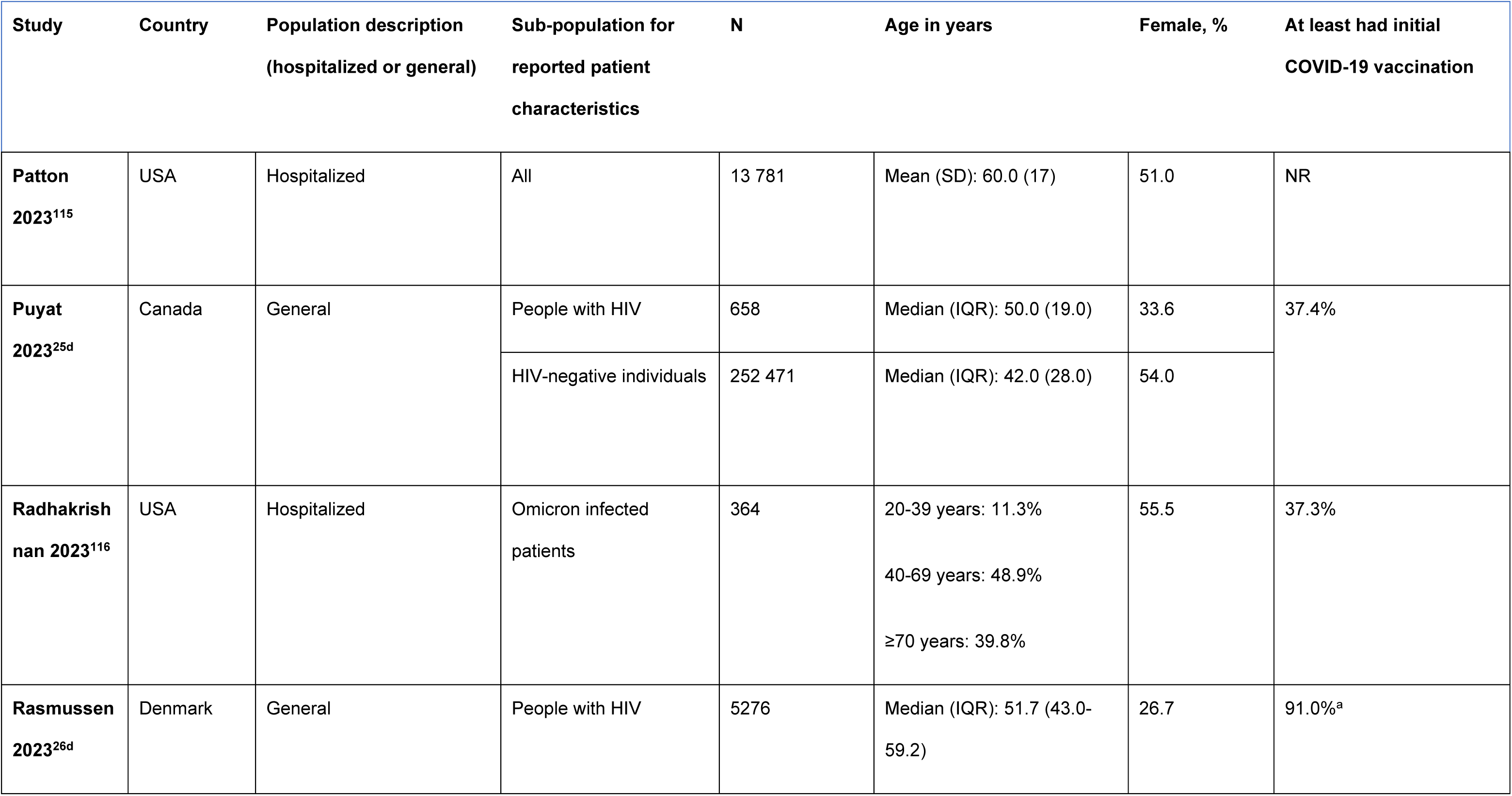
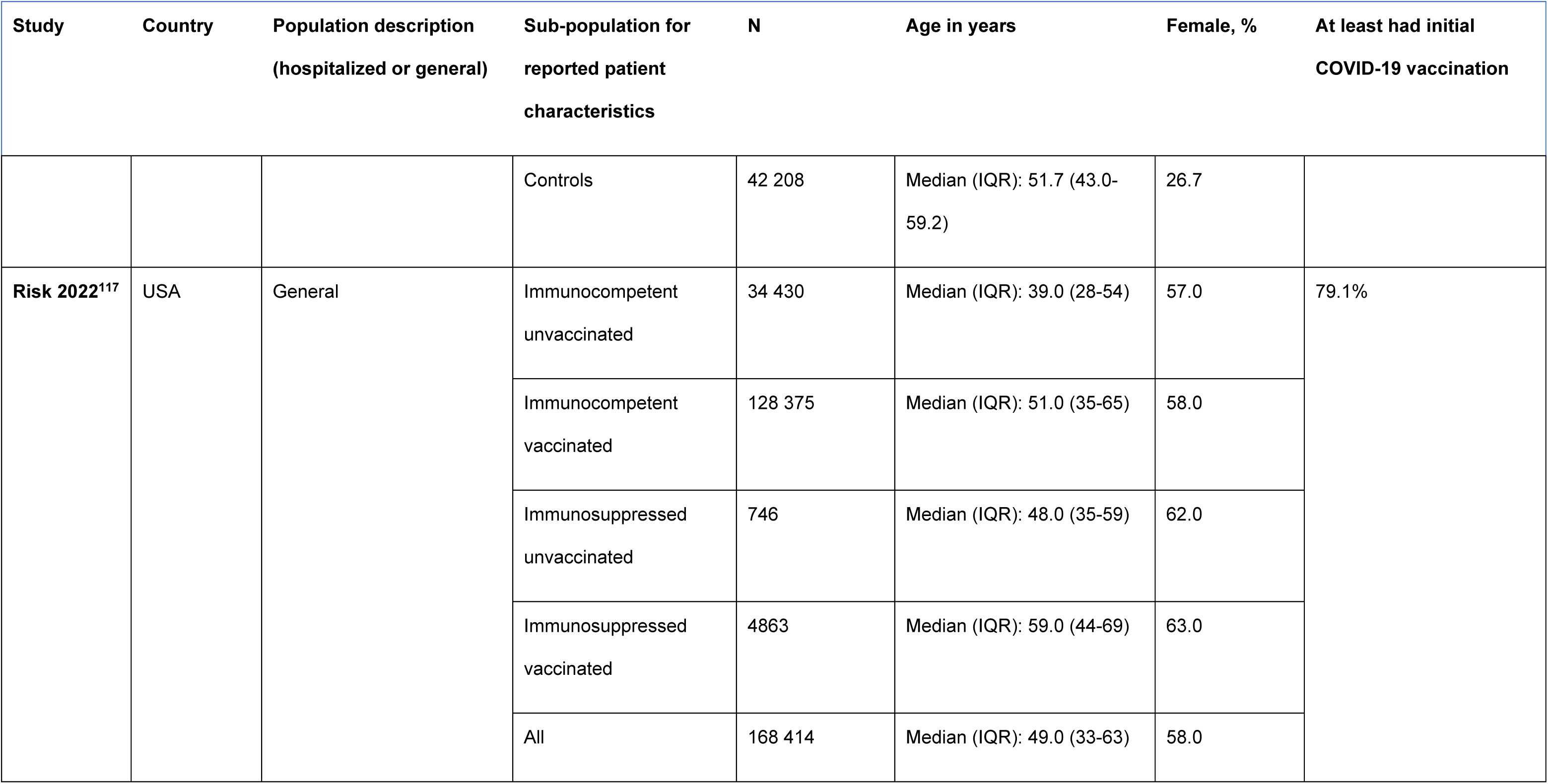
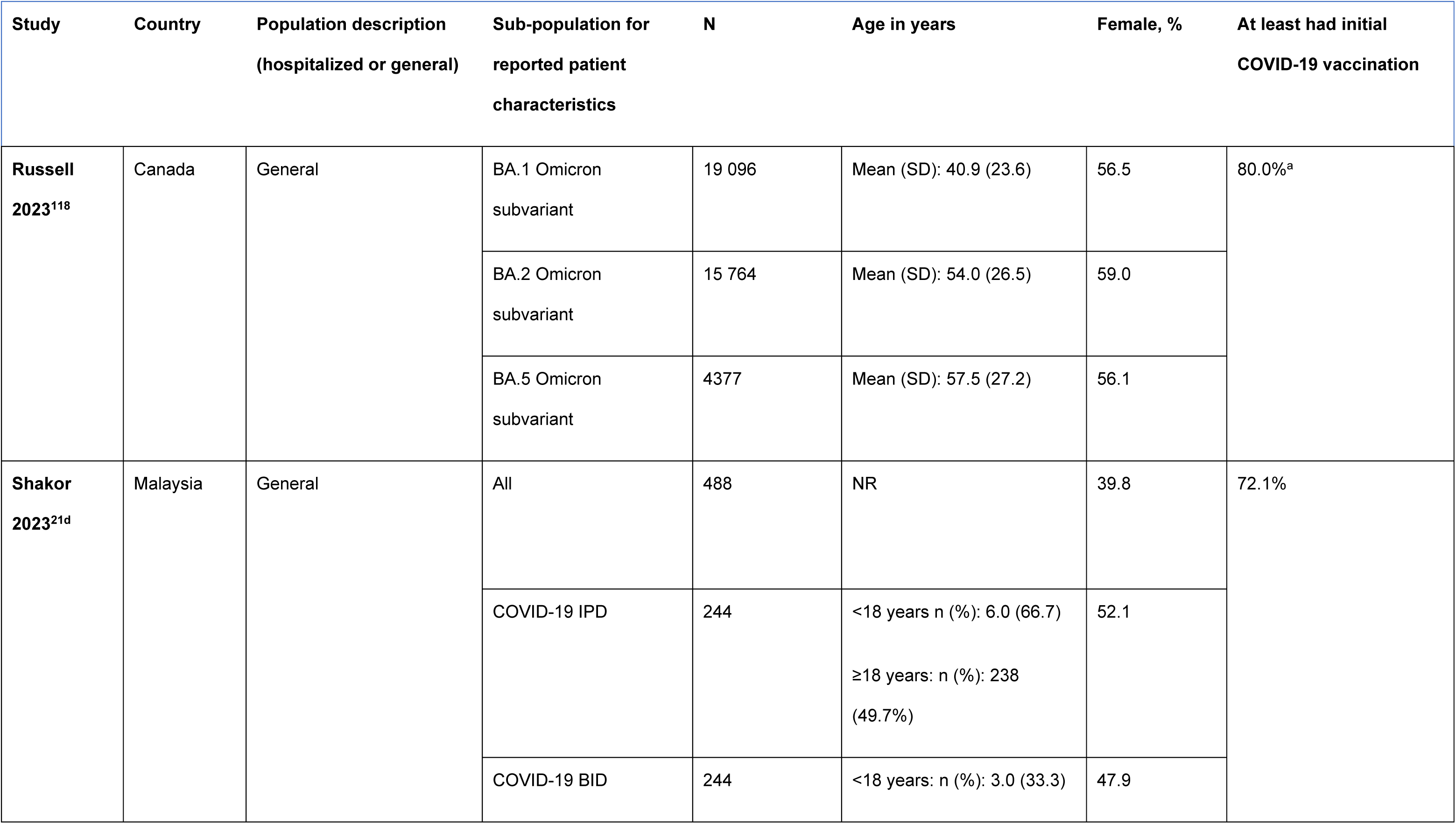
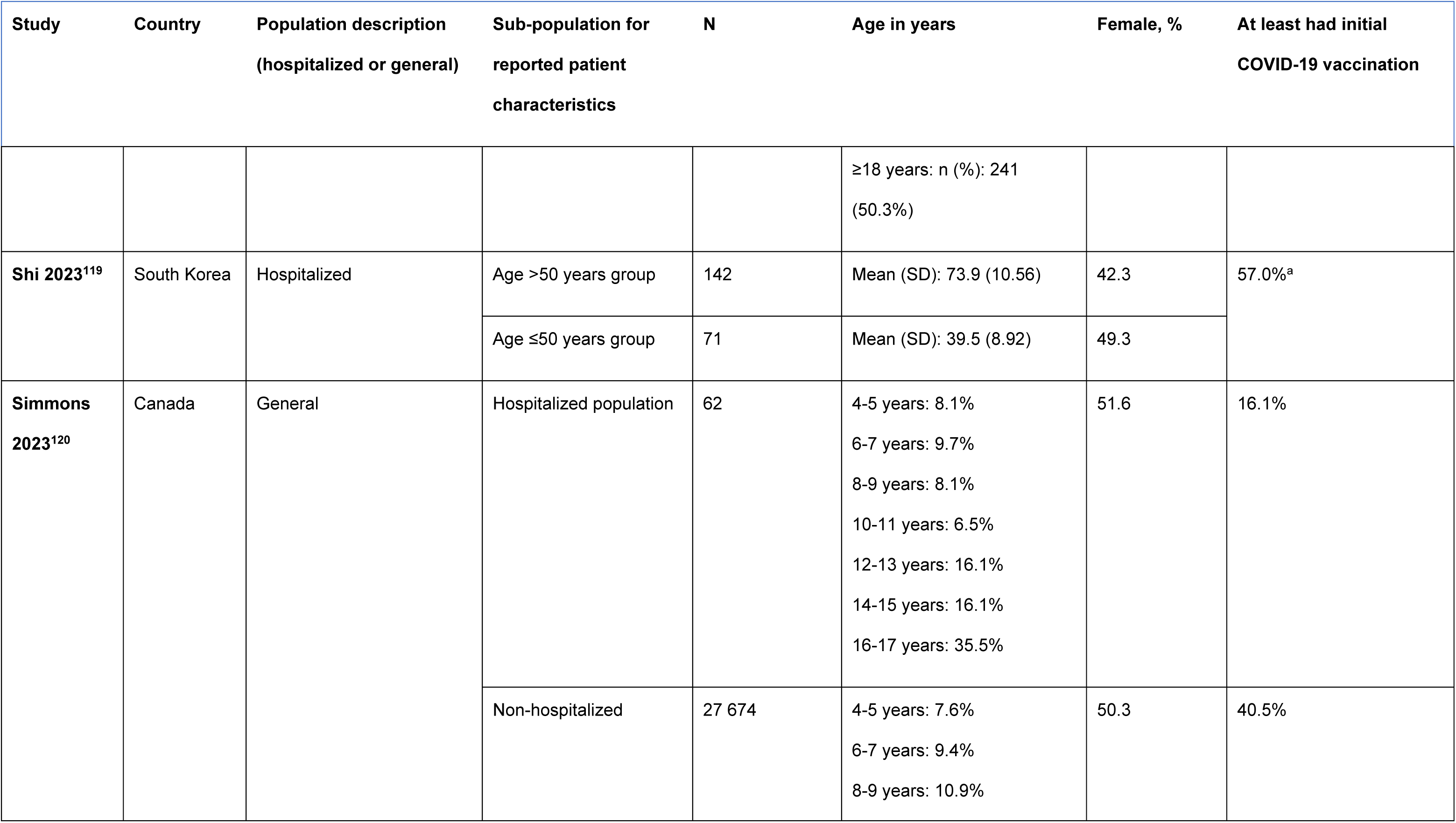
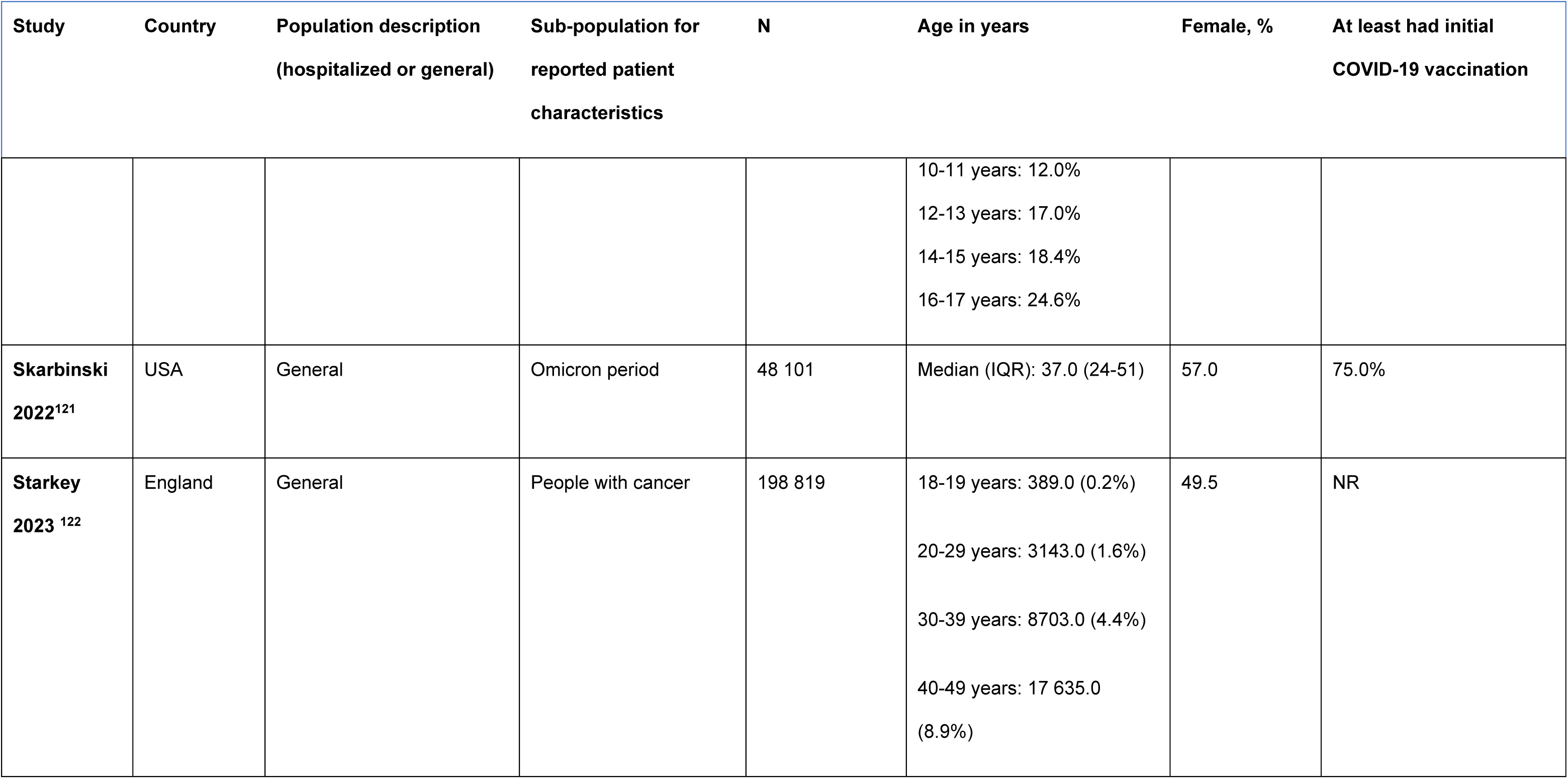
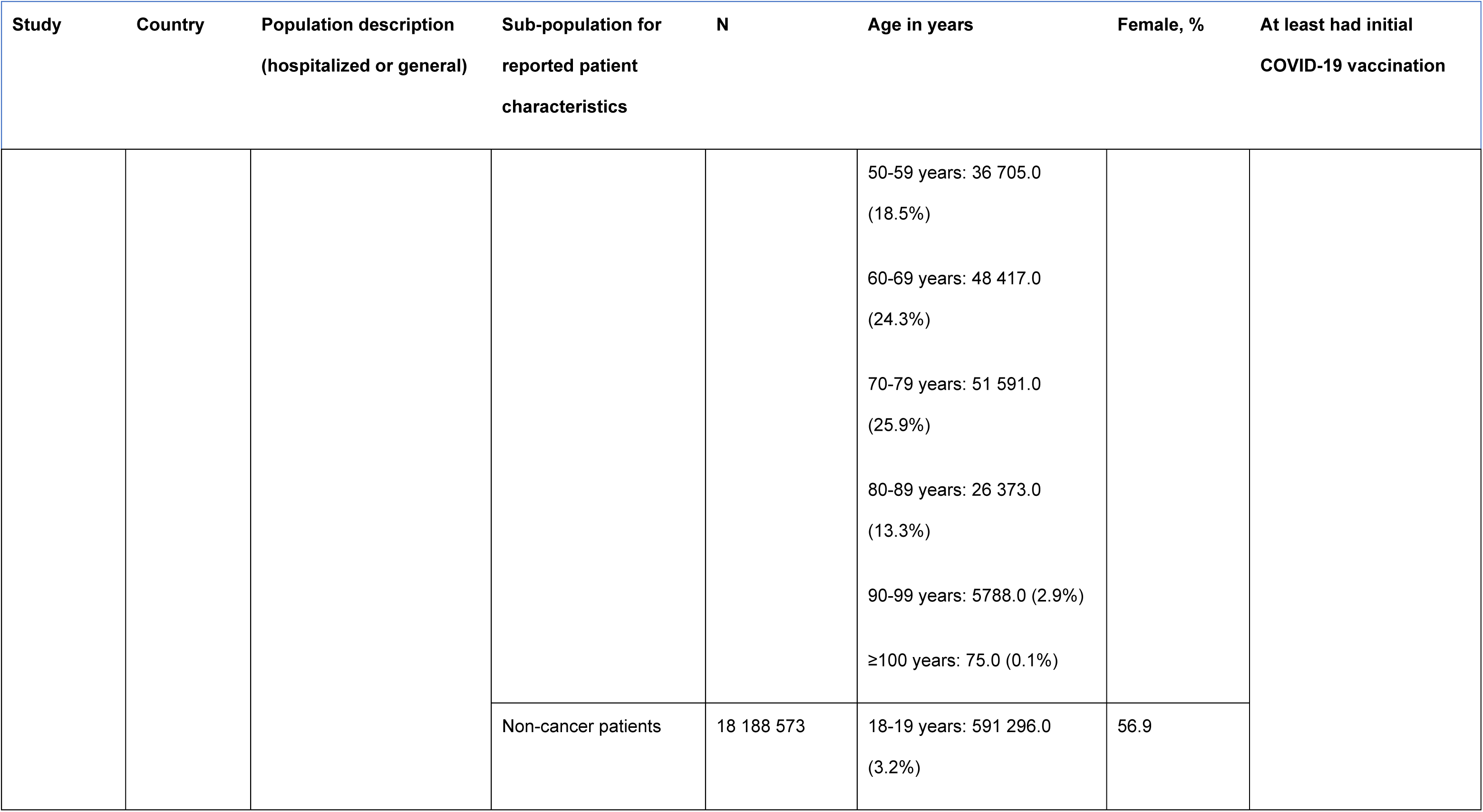
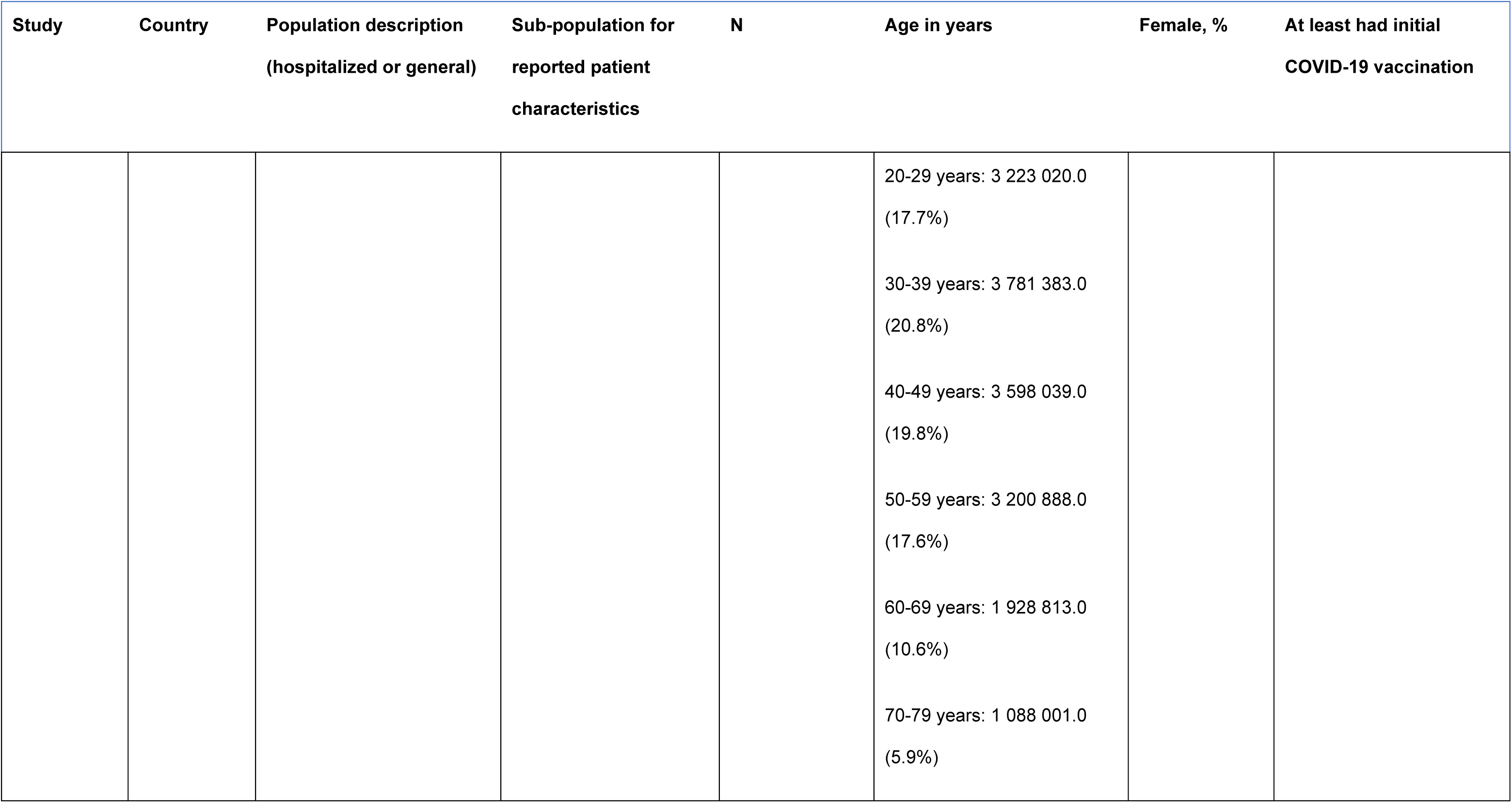
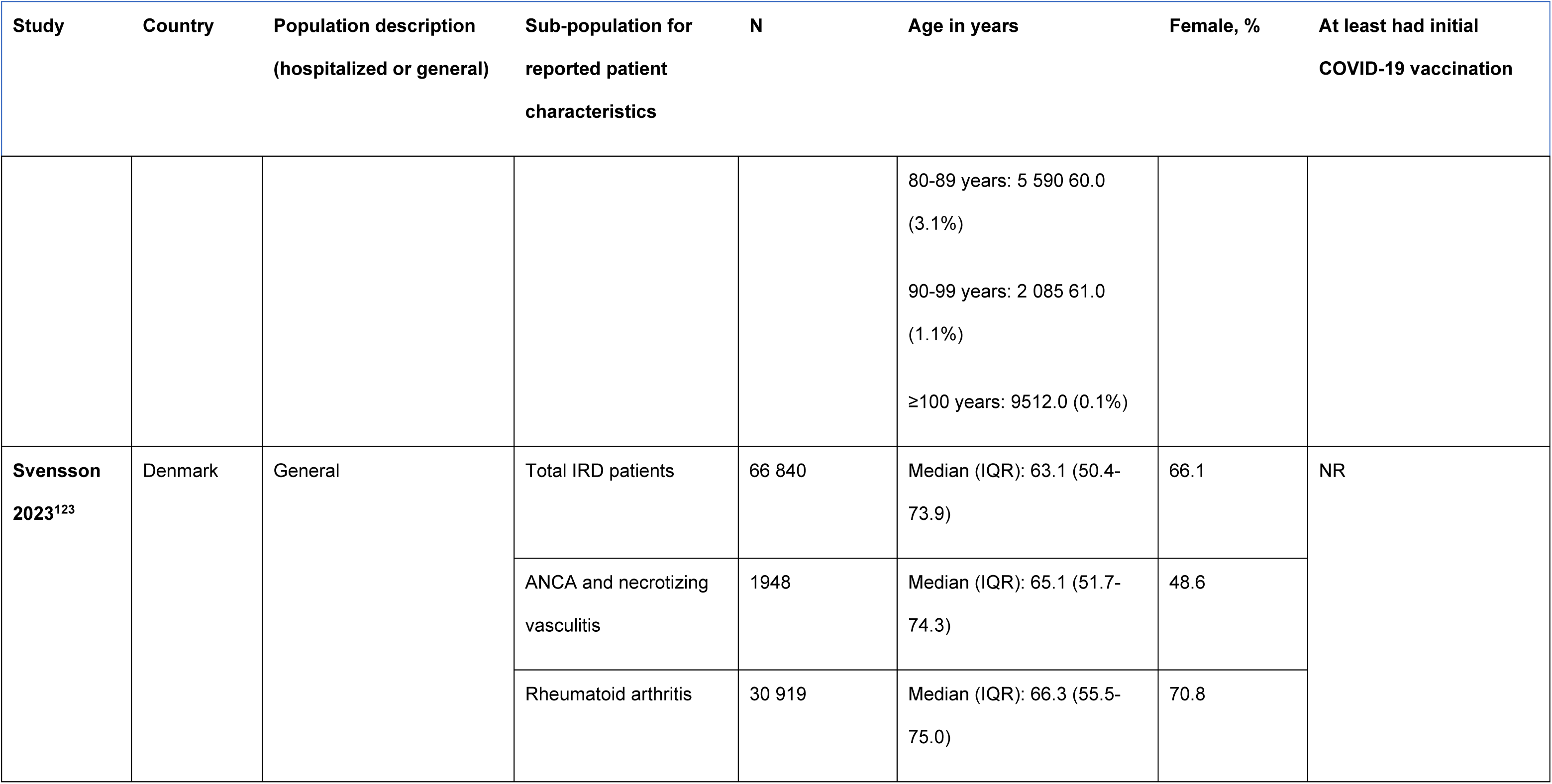
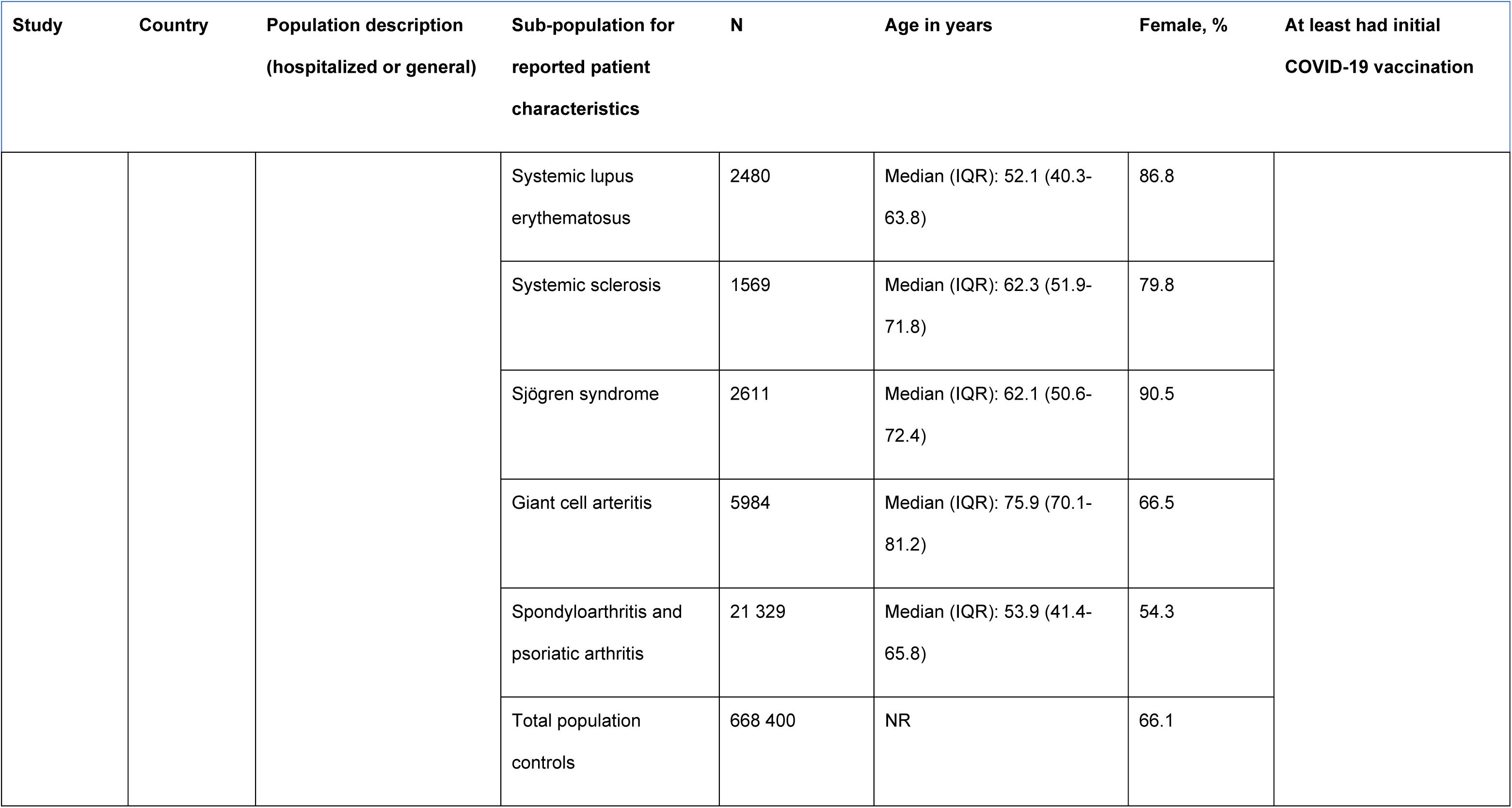
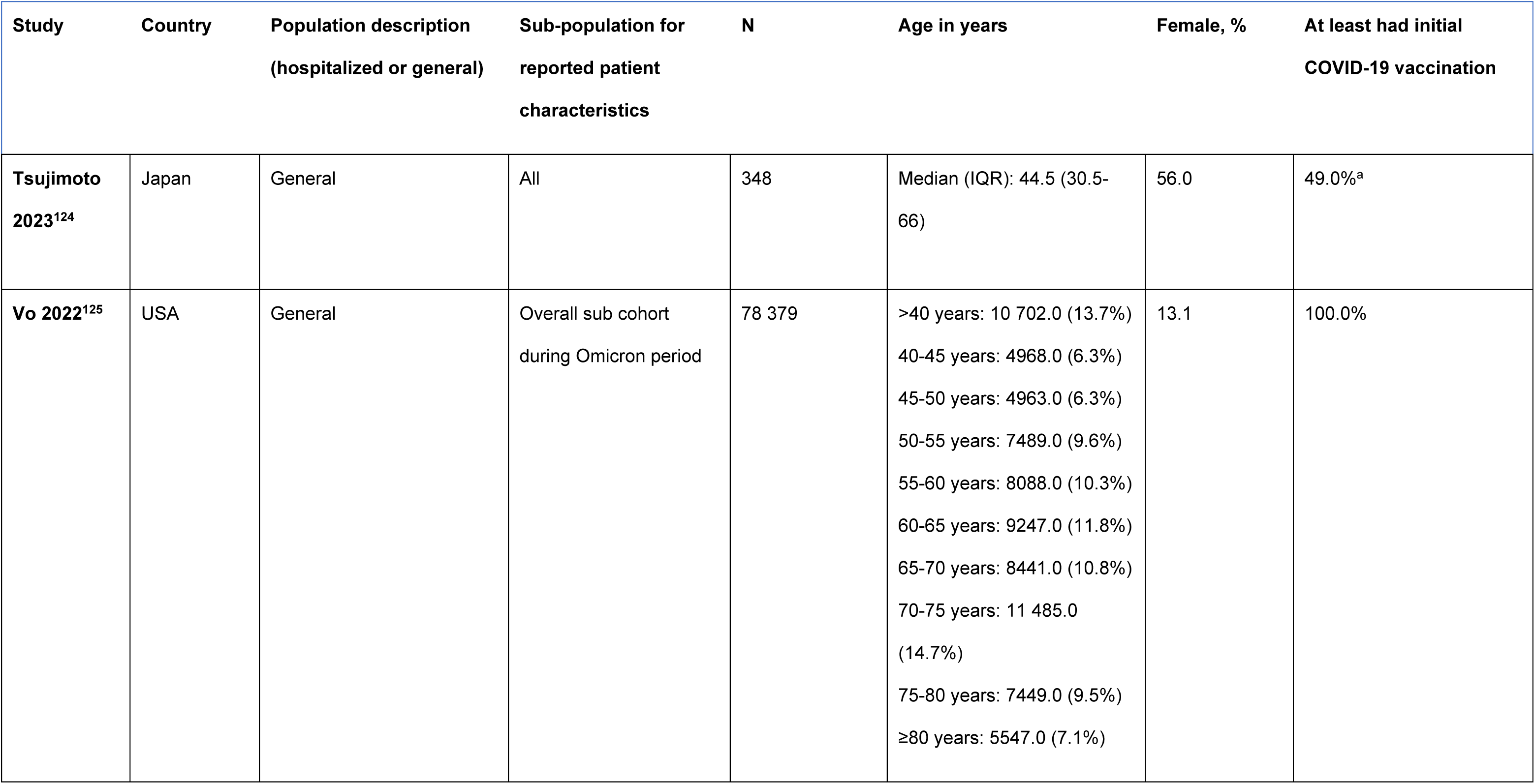
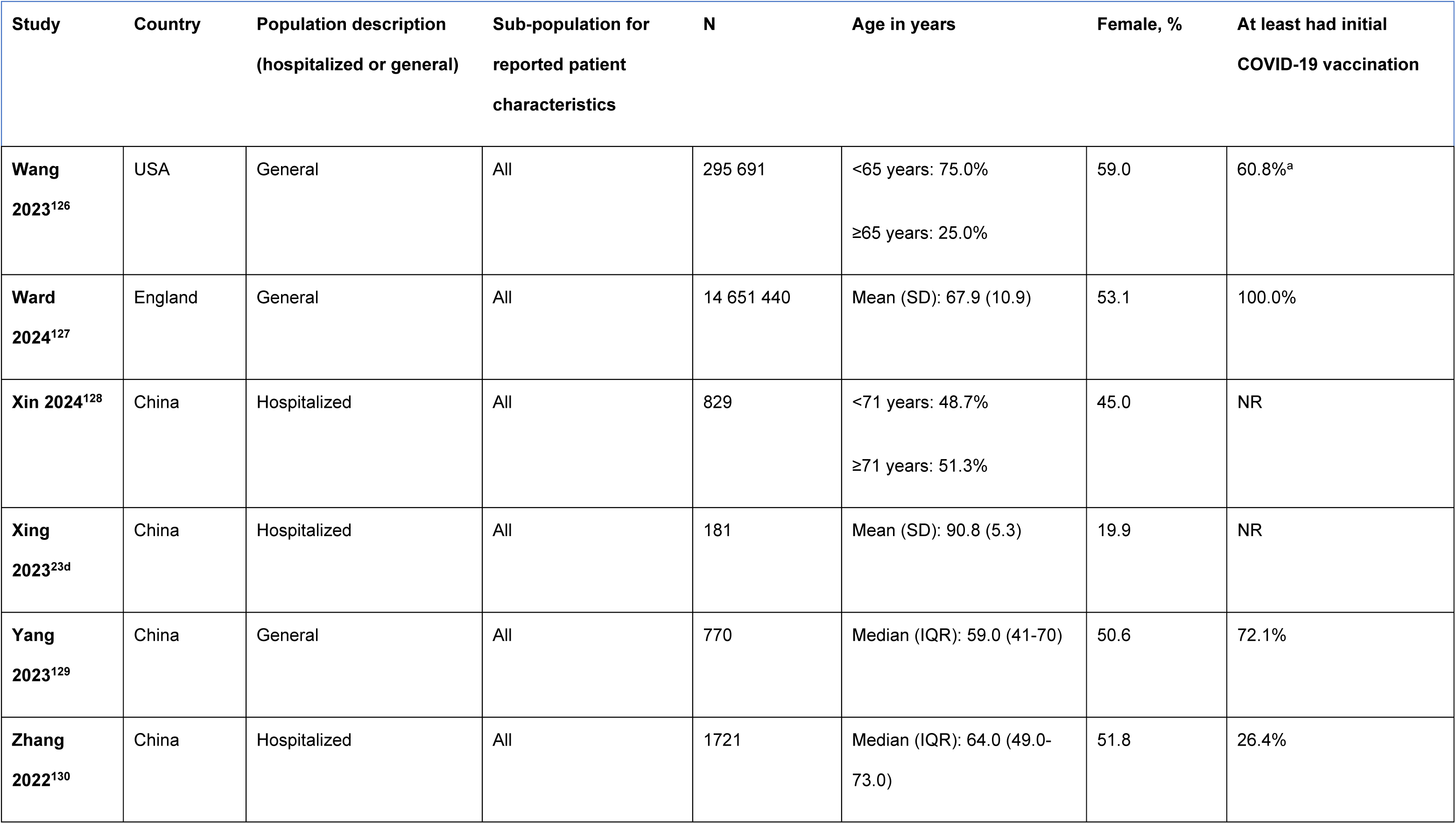
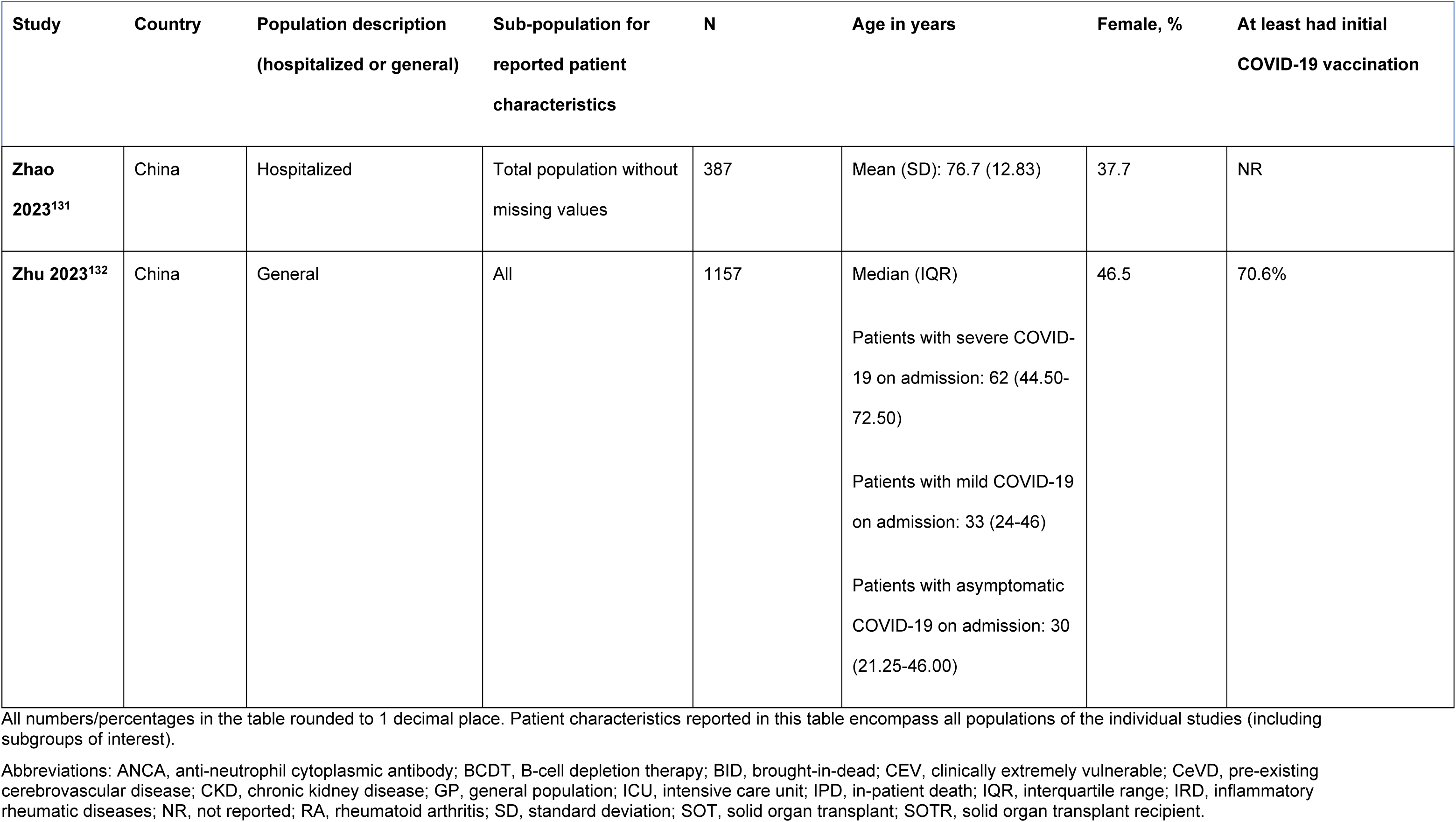
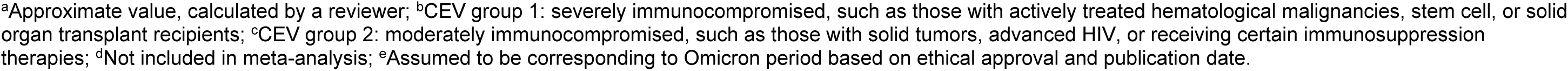
Patient Characteristics of the Included Studies.

**Table 3.** Number of Studies, Subgroups, and Minimum Number of Participants Included in Each IC/IS Condition Analysis.

| IC/IS condition | Number of studies reporting IC/IS condition | Number of studies included in the 'Main' meta-analysis | Number of subgroups<sup>a</sup> included in each 'Main' IC/IS condition analysis by outcome |  |  |  | Minimum number of participants<sup>b</sup> |
| --- | --- | --- | --- | --- | --- | --- | --- |
|  |  |  | Death | Hospitalization | ICU | Combined |  |
| **Autoimmune** | 13 | 13 | 11 | 8 | 2<sup>d</sup> | 5 | 2 557 105 |
| **Cancer** | 35 | 32 | 39 | 18 | 9 | 18 | 3 189 465 |
| **HIV** | 5 | 0<sup>c</sup> | 1<sup>d</sup> | 3<sup>d</sup> | 1<sup>d</sup> | 2<sup>d</sup> | 12 634 |
| **IC/IS unspecified** | 33 | 30 | 24 | 16 | 5 | 17 | 1 681 408 |
| **Liver disease** | 19 | 19 | 14 | 2<sup>d</sup> | 5 | 11 | 372 221 |
| **Renal disease** | 48 | 45 | 44 | 17 | 10 | 21 | 3 287 816 |
| **Transplant** | 10 | 9 | 13 | 8 | 2<sup>d</sup> | 5 | 82 955 |

Abbreviations: ICU, intensive care unit; IC/IS, immunocompromised/ immunosuppressed.
<sup>a</sup>Subgroups distinguish between the IC/IS conditions of interest and may contain additional information that differentiates the condition from another 1 included in the same meta-analysis by the same study (e.g. 'men who take immunosuppressants' and 'women who take immunosuppressants'). Some studies reported multiple subgroups that are included in the same meta-analysis; for clarity, they are referred to in this report as 'subgroups'.
<sup>b</sup>Breakdown of the number of participants by outcome not provided as most studies included more than 1 outcome of interest.
<sup>c</sup>Studies with HIV were narratively described but were not included in the main analysis because there were too few studies.
<sup>d</sup>Meta-analyses were not conducted for analyses with fewer than 5 subgroups.
Darker green indicates greater number of studies, subgroups, or participants.

### Risk of Bias

Risk of bias (**eTable 3**) was assessed in all included studies (n = 72) and was low or medium in 59 and 12 studies, respectively. One study was assessed as high risk of bias and excluded from the analyses.^21^ Three additional studies were excluded from the analyses due to not reporting confidence intervals ([CI] n = 1),^22^ a likely mistake in the publication (n = 1),^23^ and conflicting interpretation of the results (n = 1).^24^ Two studies were only summarized qualitatively as they only reported on IC/IS conditions that had insufficient data to conduct meta-analyses.^25,26^

### Meta-Analyses

A total of 66 studies were included in the meta-analyses. Individuals with the following IC/IS conditions were included in the analyses: autoimmune diseases, cancer, HIV, IC/IS unspecified, liver disease, renal disease, and transplant recipients. Conditions that could not be meta-analyzed are discussed in **Section 2.1** of the **eResults**.

### Risk of Death

For all assessed IC/IS conditions (autoimmune diseases, cancer, liver disease, renal diseases, IC/IS unspecified or transplant), people with the condition had a significantly increased risk of death (**Figure 2A**) in comparison with people without the condition. Statistical heterogeneity ranged from considerable to substantial for all IC/IS conditions (**Figure 2A**). Publication bias, as evidenced by the statistical results, was present in the renal disease and transplant groups (**eTable 4**).

**Figure 2.**
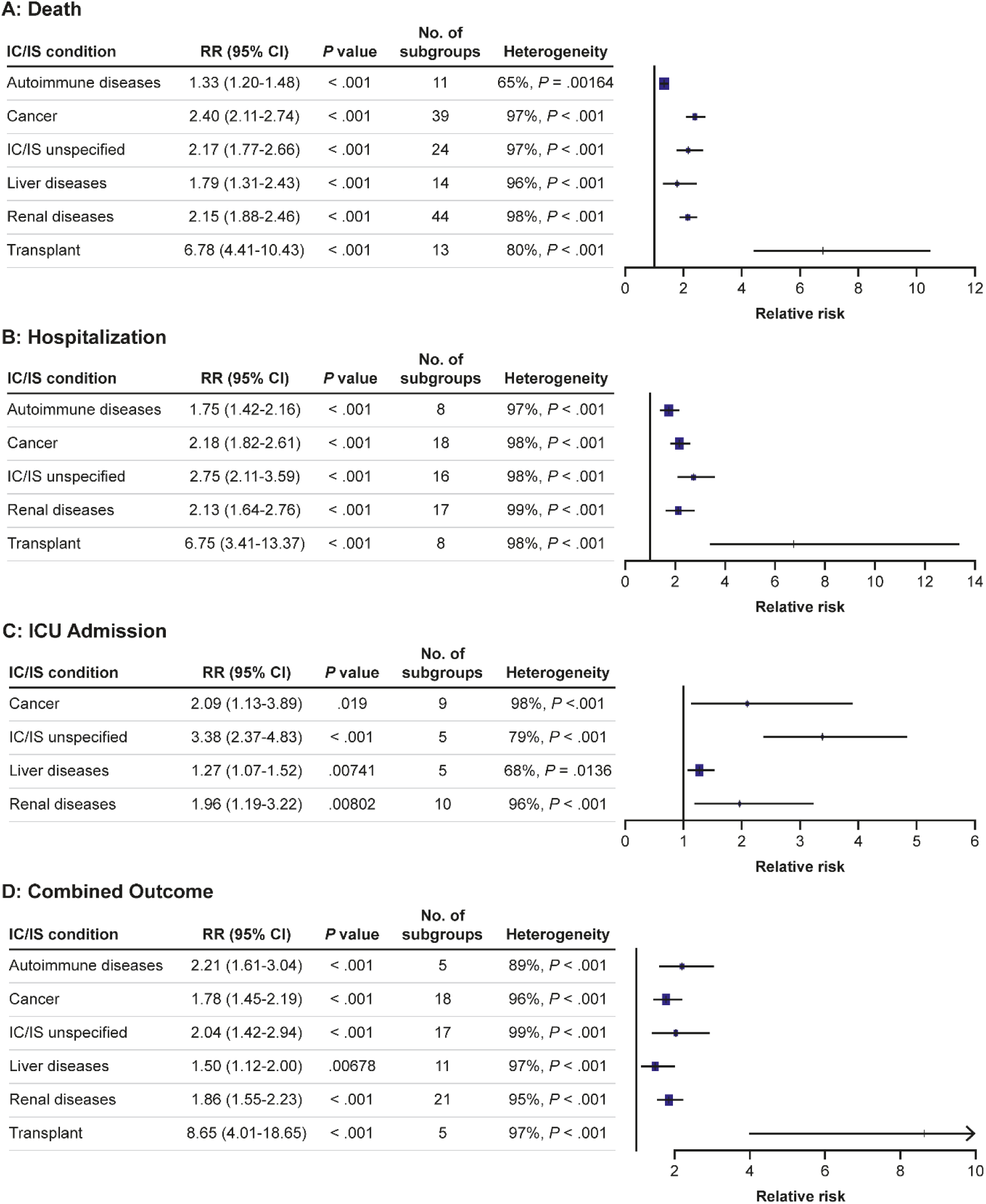
**Co**mbined Forest Plots of the A) Death, B) Hospitalization, C) ICU Admission, and D) Combined Outcomes by IC/IS Condition. CI, confidence interval; IC/IS, immunocompromised/immunosuppressed; ICU, intensive care unit; RR, risk ratio.

All sensitivity analyses results had the same degree of significance and direction of effect as the main analysis for the risk of death (**eTable 4**), indicating the robustness of the main analysis results. In subgroup analyses (**eResults, Section 2.2**), people with advanced renal disease (i.e., chronic kidney disease [CKD] stage 5, renal failure, end-stage kidney disease, and renal replacement therapy subgroups) had a much higher risk of death (RR, 3.57; 95% CI, 2.06-6.19) than people with ‘CKD stage 3’ (RR, 1.38; 95% CI, 1.17-1.64) (**eTable 5**).

### Risk of Hospitalization

For all assessed IC/IS conditions (autoimmune diseases, cancer, renal diseases, IC/IS unspecified and transplant), people with the condition had a significantly increased risk of hospitalization (**Figure 2B**). Statistical heterogeneity was considerable (>90%) across all IC/IS conditions (**Figure 2B**). Publication bias was not detected in any of the groups, but could not be assessed in studies of people with autoimmune diseases or transplant recipients due to a low number of studies (**eTable 6**).

All sensitivity analyses results had the same degree of significance as the main analysis for the risk of hospitalization (**eTable 6**), indicating the robustness of the main analysis results. All subgroup analyses were consistent with the main analysis (**eTable 7**).

### Risk of ICU Admission

For all assessed IC/IS conditions (cancer, liver diseases, renal diseases or IC/IS unspecified), people with the condition had a significantly increased risk of ICU admission (**Figure 2C**). Statistical heterogeneity ranged from substantial to considerable across the IC/IS conditions (**Figure 2C**). Publication bias was not detected for studies of people with renal disease but could not be assessed for other conditions due to a low number of studies (**eTable 8**).

Nearly all sensitivity analyses resulted in the same degree of significance as the main analysis (**eTable 8**). Subgroup analyses are discussed in the **eResults** (**Section 2.3** and **eTable 9**).

### Risk of Any Combination of Outcomes (Death, Hospitalization, or ICU Admission)

For all assessed IC/IS conditions (autoimmune diseases, cancer, liver disease, renal diseases, IC/IS unspecified or transplant), people with the condition had a significantly increased risk of the ‘Combined’ outcome (**Figure 2D**). All sensitivity analyses resulted in the same significance and direction of effect as the main analyses, indicating the robustness of the results (**eTable 10**). All subgroup analyses resulted in the same direction of effect as the main analyses; those that resulted in the loss of significance are described in the **eResults** (**eTable 11**).

## Discussion

This SLR and meta-analysis found that people with IC/IS conditions had a significantly higher risk of severe outcomes from COVID-19 during the Omicron era compared with people without the respective conditions. Individuals with cancer, IC/IS unspecified, and renal disease were at least twice as likely to die, become hospitalized, or be admitted to the ICU. Individuals who had received transplants (including solid organ, stem cells, or bone marrow) were at least six times more likely to die (pooled RR, 6.78; 95% CI, 4.41-10.43), become hospitalized (pooled RR, 6.75; 95% CI, 3.41-13.37), or experience a combined outcome (pooled RR, 8.65; 95% CI, 4.01-18.65) during the Omicron era. People included in the IC/IS unspecified group had the highest risk of ICU admission (pooled RR, 3.38; 95% CI, 2.37-4.83]).

Generally, people with IC/IS conditions had a lower risk of any outcome if they were included in the ‘Hospitalized’ subgroup, compared with the risk observed in the main analysis. People with autoimmune or liver diseases had no significant risk of ‘Death’ or ‘Combined’ outcomes. Additionally, people with cancer or renal diseases had no significant risk of ICU admission. This may be due to the lower number of studies included in these sub-analyses. Alternatively, these people may have been protected from more severe COVID-19 outcomes due to already receiving hospital care and treatments.

Several pre-Omicron analyses have shown an increased risk of severe outcomes in people with autoimmune conditions,^27^ cancer,^28,29^ general IC/IS conditions,^30^ renal disease,^31^ and transplant recipients.^32^ Immunosuppressive therapies for such IC/IS conditions downregulate the immune response, increasing the risk of infection and severe outcomes without further preventative measures.^33–36^ Specific mechanisms for individual IC/IS conditions are discussed below.

### Autoimmune Diseases

Studies showed an increased risk of developing an autoimmune disease, such as inflammatory arthritis/rheumatoid arthritis (RA), after COVID-19.^37,38^ This indicates a potential positive feedback loop between autoimmunity and COVID-19. Additionally, people with autoimmune diseases may have a higher risk of severe COVID-19 due to comorbidities, use of immunosuppressive medications, or cytokine storm/hyperinflammation.^39,40^ Comorbidities may independently contribute to severe outcomes, as one study found that people with autoimmune diseases did not have significantly higher risks when adjusting for smoking and comorbidities.^39^ The balance of disease-specific therapies and level of inflammation can also lead to divergent outcomes. For example, rituximab use in people with RA was associated with an increased risk of COVID-19-related hospitalization, ICU admission, and invasive ventilation.^41^ Alternatively, anti-tumor necrosis factor and anti-interleukin (IL)-6 have been tested as medications to prevent severe outcomes from COVID-19.^42^ The observed increased risk of mortality in autoimmune diseases may relate to cytokine storm, enhanced by the disease or its treatment. However, some therapies, such as the RA drug baricitinib,^43^ could potentially offer protection, though this has not yet been proven.

### Cancer

One study suggested that the risk of COVID-19-related death for people with cancer is lower in the Omicron era than in the prior waves, though they are still at higher risk of severe outcomes, particularly if unvaccinated.^44^ Early pandemic reports indicated that people with cancer may be at higher risk of severe COVID-19-associated outcomes due to nosocomial exposure^45^ and reduced access to treatment and follow-ups.^29^ Additional risk factors include upregulation of proteins facilitating viral infection (TMPRSS2 in prostate cancer), immunosuppressive effects of the tumor or therapies, or cytokine storms exacerbated by cancer therapies and SARS-CoV-2 infection.^46^ SARS-CoV-2 can also cause indirect damage to organs by exacerbating cancer-associated, hypoxia-mediated systemic inflammation injury via upregulated IL-6.^47^ Notably, people with hematological malignancies have a significantly higher risk of COVID-19-related mortality compared with people with solid tumors, possibly due to a weaker immune system or an increased risk of thrombosis.^48^

### Immunocompromised/Immunosuppressed

A pre-Omicron SLR and meta-analysis indicated that immunosuppression and immunodeficiency were associated with an increased risk of severe COVID-19; however, unlike the analysis described here, the results were not significant.^30^ Omicron-era studies found that individuals with IC/IS had 4.3 to 23 times greater risk of hospitalization upon first COVID-19 diagnosis and a significantly increased risk of in-hospital mortality compared with people without IC/IS conditions.^13,49^

### Renal Diseases

People with renal diseases may experience an increased risk of severe COVID-19 outcomes due to immune system dysfunction, chronic systemic inflammation associated with kidney impairment, and CKD-associated comorbidities (e.g., cardiovascular disease, anemia, vitamin D deficiency, etc.).^50,51^ Additionally, SARS-CoV-2 can cause acute kidney injury through direct infection of tissues in the kidneys and by inducing chronic inflammation,^51^ which could further exacerbate existing kidney issues. Worsening CKD stage and comorbidities are independent risk factors for COVID-19-associated hospitalization and death in people with renal disease.^52,53^

### Transplant

Transplant recipients are at higher risk of severe COVID-19 outcomes due to immunosuppressive drugs downregulating innate and adaptive immunity,^54,55^ comorbidities, and suboptimal organ function.^56^ Moreover, solid-organ transplant recipients have lower response rates to SARS-CoV-2 vaccines^35^ highlighting the need for enhanced protective measures including additional doses.

### HIV

Systematic reviews before the Omicron era reported a higher risk of mortality^57^ and hospitalization^58^ from COVID-19 for people with HIV compared with people without HIV. However, some studies suggested no difference in COVID-19 mortality rates for people with well-controlled HIV compared with HIV-negative individuals. Among people with HIV, those with low CD4+ T cell counts or uncontrolled viral loads are more susceptible to severe COVID-19 outcomes.^57,59^ Understanding the risk of severe COVID-19 outcomes for people with HIV is complicated by variations in disease control, immunosuppression, and antiretroviral use.^57,58^ The lack of high-quality prospective studies on COVID-19 outcomes for people with HIV stratified by antiretroviral use and disease severity, presents an unmet need for this population.

### Quality of Evidence

#### Strengths

To our knowledge, this is the first comprehensive SLR and meta-analysis to assess severe outcomes from COVID-19 in people with IC/IS conditions during the Omicron period. Most of the sensitivity analyses showed no change in significance from the main analyses, indicating robust pooled estimates. Additionally, most of the results included in the meta-analyses were adjusted for age and comorbidities, increasing confidence in the findings.

#### Limitations

Several limitations were identified in this analysis. High clinical and statistical heterogeneity existed between studies due to variations in outcome definitions and adjustments (e.g. vaccination rates and comorbidities). Heterogeneity in vaccination status and confounding adjustments prevented subgroup analyses by vaccination status. The studies mainly covered earlier Omicron variants; however, it was not possible to stratify by subvariants. Additionally, many studies on disease flares following COVID-19 or vaccination were excluded as they did not directly address the research question.

### Implications for Practice and Future Research

This analysis demonstrates that the COVID-19 burden remains high in populations with IC/IS conditions, necessitating continuous adaptation of public health strategies to protect these individuals as SARS-CoV-2 evolves. Vaccination remains the most effective defense against severe outcomes from COVID-19, especially for people with IC/IS conditions, whereby a 3-dose primary series (as opposed to 2 doses) and additional booster doses are recommended.^60,61^ Carefully timing vaccine doses or temporarily adjusting immunosuppressive therapies post-vaccination may offer benefits.^62,63^ However, this should be tailored to each patient’s specific disease status and therapy regimen, balancing the benefits against potential risks.^62–64^ Given that people with IC/IS conditions often have multiple comorbidities and differing treatments, a personalized and multi-disciplinary approach to disease management and COVID-19 prevention is advantageous.^65^

This study identified a dearth of high-quality prospective research on severe outcomes from COVID-19 in people living with HIV, indicating a need for additional studies in this population. Additionally, given there are disparities in COVID-19 testing, vaccination, and outpatient therapeutics access based on race and ethnicity;^66–68^ further research into how these disparities impact people with IC/IS conditions is warranted. Future research should also investigate the impact of age, Omicron subvariant, vaccination status, and geographical region on people with IC/IS conditions.

## Conclusions

This SLR and meta-analysis demonstrated that people with IC/IS conditions are at an increased risk of severe outcomes from COVID-19 during the Omicron era. Of the meta-analyzed IC/IS conditions, transplant recipients were at the highest risk for hospitalization, death, and combined outcomes. Our study highlights the need for continued enhanced preventative measures for IC/IS populations, and a personalized multi-disciplinary approach to care.

## Data Availability

The datasets generated during and/or analyzed during the current study are available from the corresponding author on reasonable request.

## Acknowledgements

The authors would like to thank the individuals, their families, and all investigators involved in this study.

Guidance through the review process as well as contributions to systematic review processes, such as screening, risk of bias assessment, and data extraction, were provided by Nick Pooley, PhD (Maverex Ltd), Masoumeh Kisomi, PhD (Maverex Ltd), and Megha Garg, MD (Maverex Ltd). Statistical support, including the design and running of the meta-analyses, was provided by Medha Shrivastava, MSc (Maverex Ltd). Kate Misso, MSc/MCLIP (Maverex Ltd), designed and performed the electronic searches in this systematic review.

Medical writing support, including assisting authors with development of the outline and initial draft, incorporation of comments, figure preparation, referencing, and data checking was provided by Ashley Knox, PhD, and editorial support, including formatting, proofreading, and submission was provided by Michelle Seddon, Dip Psychol, all of Paragon (a division of Prime, Knutsford, UK). The study was supported by BioNTech SE, Mainz, Germany, according to Good Publication Practice guidelines (<u>Link</u>). The sponsor was involved in the study design, analysis and interpretation of data in the manuscript as well as data checking of information provided in the manuscript. However, ultimate responsibility for opinions, conclusions, and data interpretation lies with the authors.

## Conflicts of Interest Disclosures

AC is an employee of Maverex Ltd., who received consulting fees from BioNTech SE.

FB has no conflicts to disclose.

GC has been an advisory board member for Roche, Novartis, Lilly, Pfizer, Astra Zeneca, Daichii Sankyo, Ellipsis, Veracyte, Exact Science, Celcuity, Merck, BMS, Gilead, Sanofi, and Menarini.

TP receives consulting fees from BioNTech SE, GlaxoSmithKline, UNAIDS, and USAID.

SA is an employee of BioNTech SE.

## Funding

This study was funded by BioNTech SE, Mainz, Germany.

## Author Contributions

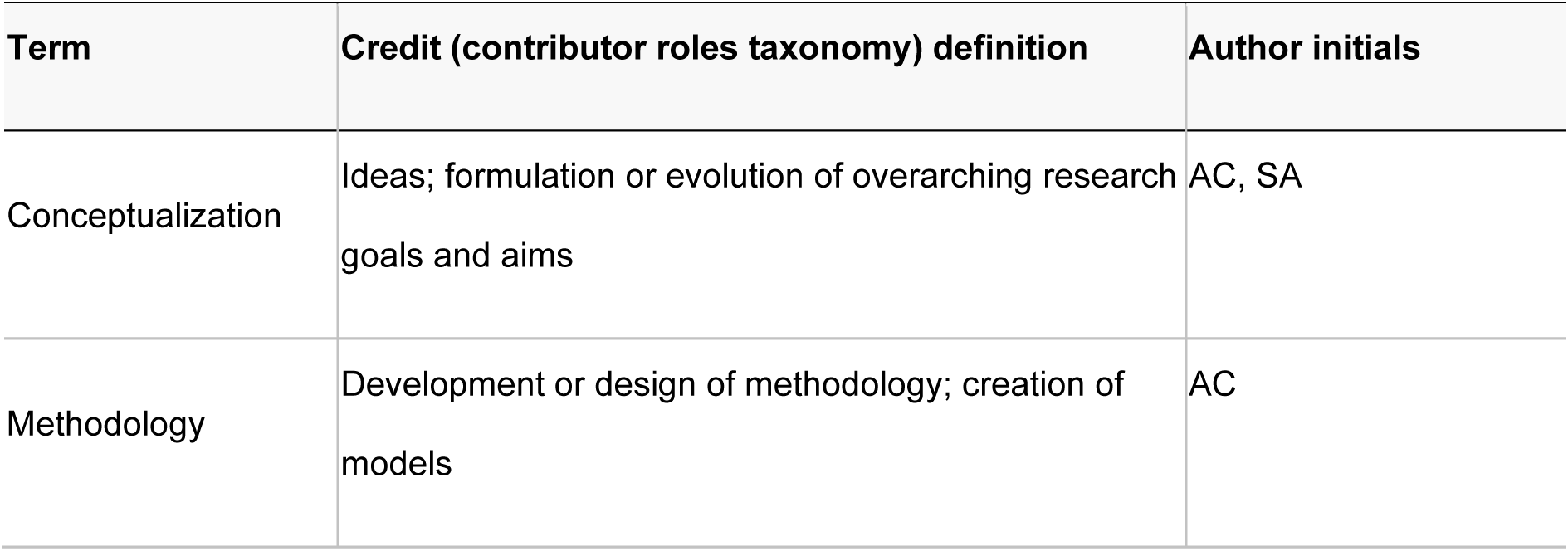
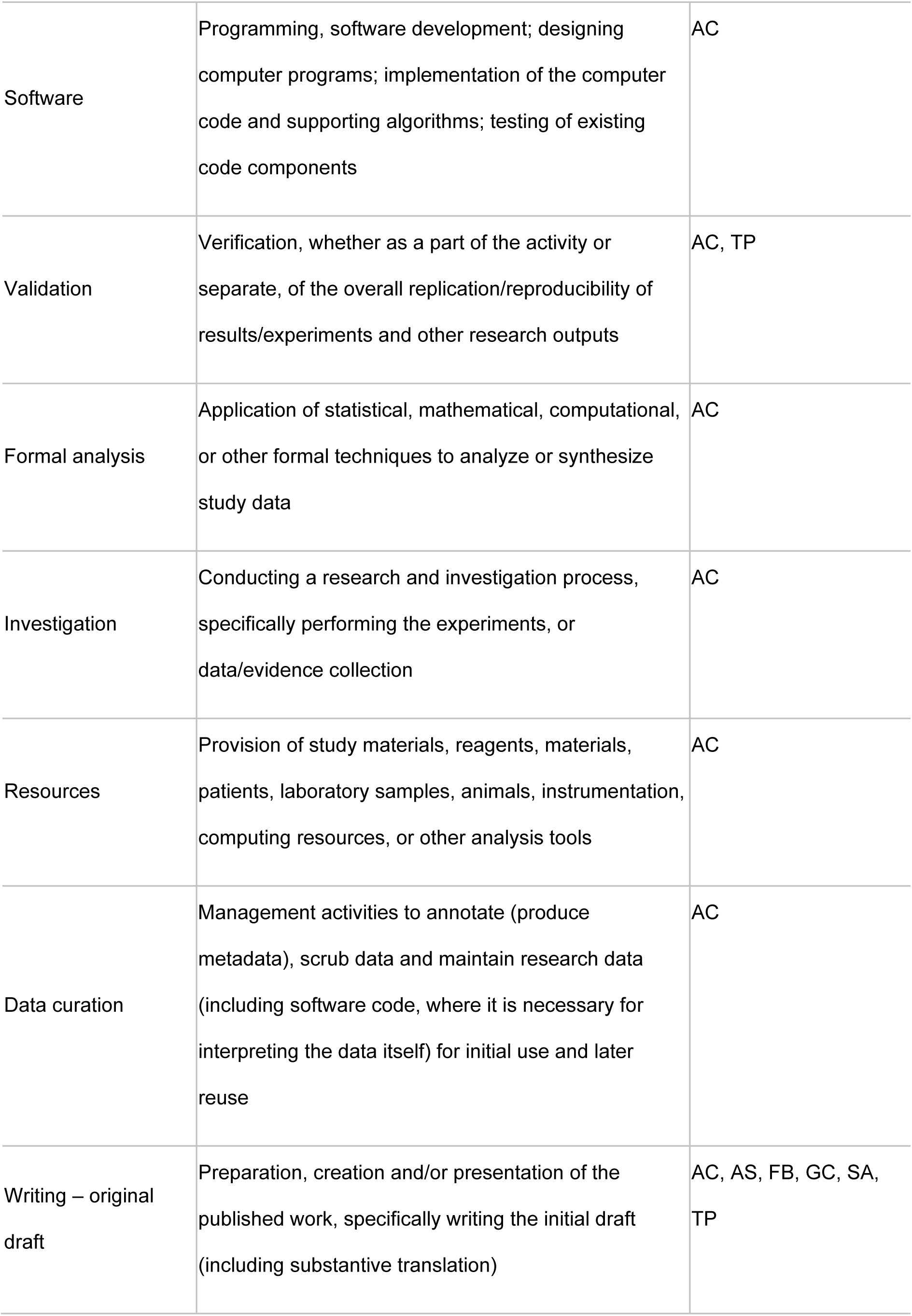
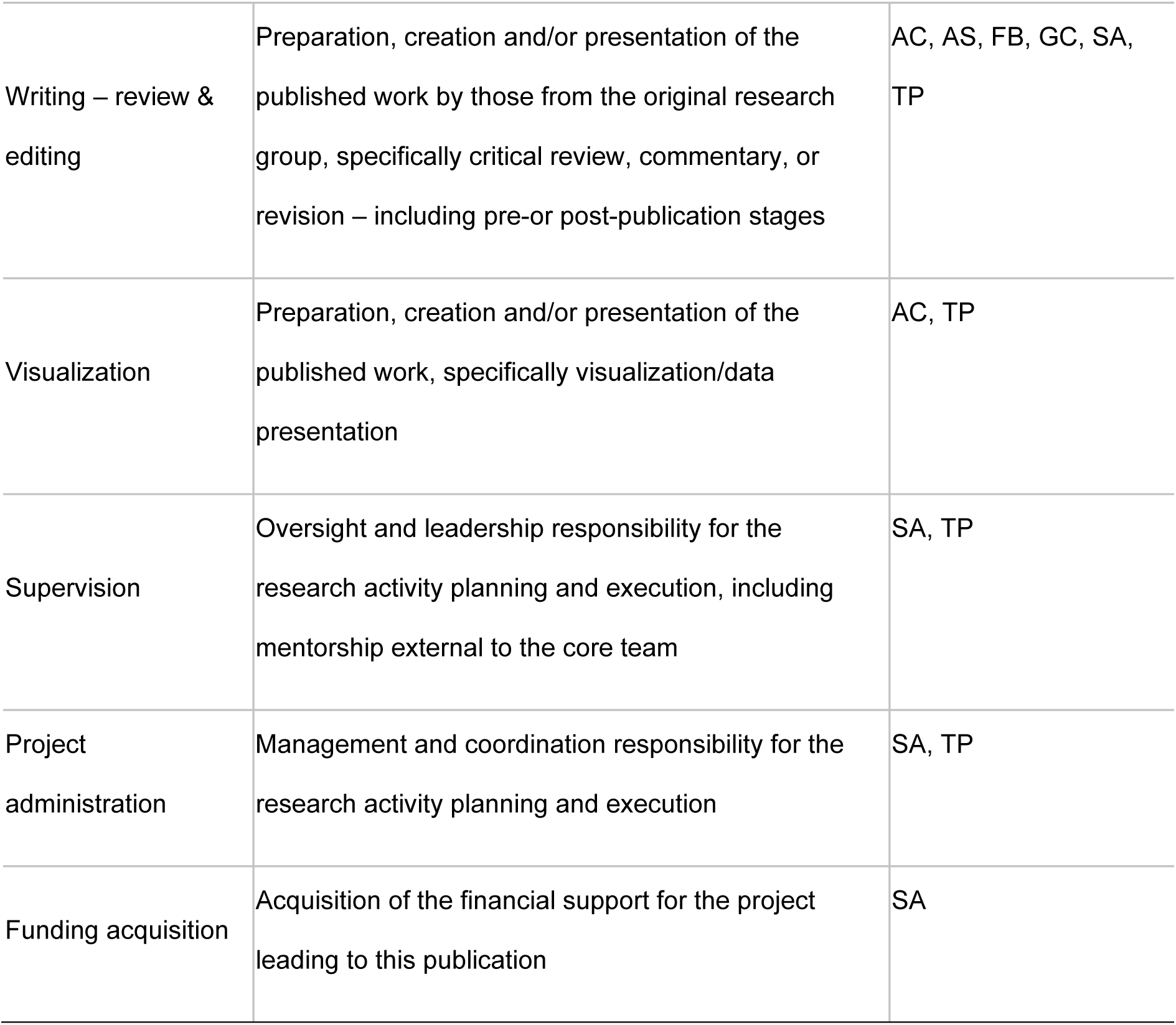

## Notes

### Author Declarations

All data obtained from published references' and then copy in the list of references in List of References Included in Systematic Review.

## References

1. World Health Organization. WHO Director-General’s statement on IHR Emergency Committee on Novel Coronavirus (2019-nCoV). Updated January 30, 2020.https://www.who.int/director-general/speeches/detail/who-director-general-s-statement-on-ihr-emergency-committee-on-novel-coronavirus-(2019-ncov)). Accessed July 3, 2024.

2. World Health Organization. Coronavirus disease 2019 (COVID-19) Situation Report – 51. Updated March 11, 2020. https://www.who.int/docs/default-source/coronaviruse/situation-reports/20200311-sitrep-51-covid-19.pdf?sfvrsn=1ba62e57_10. Accessed July 3, 2024.

3. World Health Organization. Classification of Omicron (B.1.1.529): SARS-CoV-2 variant of concern. Updated November 26, 2021.https://www.who.int/news/item/26-11-2021-classification-of-omicron-(b.1.1.529)-sars-cov-2-variant-of-concern. Accessed July 3, 2024.

4. Petrone D, Mateo-Urdiales A, Sacco C, et al. Reduction of the risk of severe COVID-19 due to Omicron compared to Delta variant in Italy (November 2021 - February 2022). Int J Infect Dis. 2023;129:135–141.

5. Bager P, Wohlfahrt J, Bhatt S, et al. Risk of hospitalisation associated with infection with SARS-CoV-2 omicron variant versus delta variant in Denmark: an observational cohort study. Lancet Infect Dis. 2022;22(7):967–976.

6. Nyberg T, Ferguson NM, Nash SG, et al. Comparative analysis of the risks of hospitalisation and death associated with SARS-CoV-2 omicron (B.1.1.529) and delta (B.1.617.2) variants in England: a cohort study. Lancet. 2022;399(10332):1303-1312.

7. Veneti L, Boas H, Brathen Kristoffersen A, et al. Reduced risk of hospitalisation among reported COVID-19 cases infected with the SARS-CoV-2 Omicron BA.1 variant compared with the Delta variant, Norway, December 2021 to January 2022. Euro Surveill. 2022;27(4):pii=2200077.

8. Sheikh A, Kerr S, Woolhouse M, et al. Severity of omicron variant of concern and effectiveness of vaccine boosters against symptomatic disease in Scotland (EAVE II): a national cohort study with nested test-negative design. Lancet Infect Dis. 2022:959–966.

9. World Health Organization. WHO COVID-19 Dashboard. https://data.who.int/dashboards/covid19/. Accessed August 16, 2024.

10. World Health Organization. WHO Director-General’s opening remarks at the media briefing – 5 May 2023. https://www.who.int/news-room/speeches/item/who-director-general-s-opening-remarks-at-the-media-briefing5-may-2023.

11. Centers for Disease Control and Prevention. COVID-19 can surge throughout the year. Updated July 3, 2024.https://www.cdc.gov/ncird/whats-new/covid-19-can-surge-throughout-the-year.html. Accessed August 16, 2024.

12. Belsky JA, Tullius BP, Lamb MG, et al. COVID-19 in immunocompromised patients: a systematic review of cancer, hematopoietic cell and solid organ transplant patients. J Infect. 2021;82(3):329–338.

13. Turtle L, Thorpe M, Drake TM, et al. Outcome of COVID-19 in hospitalised immunocompromised patients: an analysis of the WHO ISARIC CCP-UK prospective cohort study. PLoS Med. 2023;20(1):e1004086.

14. Evans RA, Dube S, Lu Y, et al. Impact of COVID-19 on immunocompromised populations during the Omicron era: insights from the observational population-based INFORM study. Lancet Reg Health Eur. 2023;35:100747.

15. Antinori A, Bausch-Jurken M. The burden of COVID-19 in the immunocompromised patient: implications for vaccination and needs for the future. J Infect Dis. 2023;228(Suppl 1):S4–S12.

16. Li Y, Choudhary MC, Regan J, et al. SARS-CoV-2 viral clearance and evolution varies by type and severity of immunodeficiency. Sci Transl Med. 2024;16(731):eadk1599.

17. Hogan JI, Duerr R, Dimartino D, et al. Remdesivir resistance in transplant recipients with persistent Coronavirus Disease 2019. Clin Infect Dis. 2023;76(2):342–345.

18. Fung M, Babik JM. COVID-19 in immunocompromised hosts: what we know so far. Clin Infect Dis. 2021;72(2):340–350.

19. Nab L, Parker EPK, Andrews CD, et al. Changes in COVID-19-related mortality across key demographic and clinical subgroups in England from 2020 to 2022: a retrospective cohort study using the OpenSAFELY platform. Lancet Public Health. 2023;8(5):e364–e377.

20. Viechtbauer W. Conducting meta-analyses in R with the metafor package. J Stat Softw. 2010;36(3):1–48.

21. Shakor ASaA, Samsudin EZ, Chen XW, Ghazali MH. Factors associated with COVID-19 brought-in deaths: a data-linkage comparative cross-sectional study. J Infect Public Health. 2023;16(12):2068–2078.

22. Morris CP, Eldesouki RE, Sachithanandham J, et al. Omicron subvariants: clinical, laboratory, and cell culture characterization. Clin Infect Dis. 2023;76(7):1276–1284.

23. Xing Y, Sun Y, Tang M, et al. Variables associated with 30-day mortality in very elderly COVID-19 patients. Clin Interv Aging. 2023;18:1155–1162.

24. Bao S, Lu G, Kang Y, et al. A diagnostic model for serious COVID-19 infection among older adults in Shanghai during the Omicron wave. Front Med (Lausanne). 2022;9:1018516.

25. Puyat JH, Fowokan A, Wilton J, et al. Risk of COVID-19 hospitalization in people living with HIV and HIV-negative individuals and the role of COVID-19 vaccination: a retrospective cohort study. Int J Infect Dis. 2023;135:49–56.

26. Rasmussen LD, Cowan S, Gerstoft J, et al. Outcomes following severe acute respiratory syndrome coronavirus 2 infection among individuals with and without HIV in Denmark. AIDS. 2023;37(2):311–321.

27. Yang H, Xu J, Shi L, Duan G, Wang Y. Correspondence on ‘Prevalence and clinical outcomes of COVID-19 in patients with autoimmune diseases: a systematic review and meta-analysis’. Ann Rheum Dis. 2023;82(4):e79.

28. Han S, Zhuang Q, Chiang J, et al. Impact of cancer diagnoses on the outcomes of patients with COVID-19: a systematic review and meta-analysis. BMJ Open. 2022;12(2):e044661.

29. Di Felice G, Visci G, Teglia F, Angelini M, Boffetta P. Effect of cancer on outcome of COVID-19 patients: a systematic review and meta-analysis of studies of unvaccinated patients. Elife. 2022;11:e74634.

30. Gao Y, Chen Y, Liu M, Shi S, Tian J. Impacts of immunosuppression and immunodeficiency on COVID-19: a systematic review and meta-analysis. J Infect. 2020;81(2):e93–e95.

31. Lin YC, Lai TS, Lin SL, et al. Outcomes of coronavirus 2019 infection in patients with chronic kidney disease: a systematic review and meta-analysis. Ther Adv Chronic Dis. 2021;12:2040622321998860.

32. Ao G, Wang Y, Qi X, et al. The association between severe or death COVID-19 and solid organ transplantation: a systematic review and meta-analysis. Transplant Rev (Orlando). 2021;35(3):100628.

33. Liao SY, Gerber AN, Zelarney P, Make B, Wechsler ME. Impaired SARS-CoV-2 mRNA vaccine antibody response in chronic medical conditions: a real-world analysis. Chest. 2022;161(6):1490–1493.

34. Liu H, Aviszus K, Zelarney P, et al. Vaccine-elicited B-and T-cell immunity to SARS-CoV-2 is impaired in chronic lung disease patients. ERJ Open Res. 2023;9(5):00400–02023.

35. Galmiche S, Luong Nguyen LB, Tartour E, et al. Immunological and clinical efficacy of COVID-19 vaccines in immunocompromised populations: a systematic review. Clin Microbiol Infect. 2022;28(2):163–177.

36. Prendecki M, Clarke C, Edwards H, et al. Humoral and T-cell responses to SARS-CoV-2 vaccination in patients receiving immunosuppression. Ann Rheum Dis. 2021;80(10):1322–1329.

37. Lim SH, Ju HJ, Han JH, et al. Autoimmune and autoinflammatory connective tissue disorders following COVID-19. JAMA Netw Open. 2023;6(10):e2336120.

38. Marin JS, Mazenett-Granados EA, Salazar-Uribe JC, et al. Increased incidence of rheumatoid arthritis after COVID-19. Autoimmun Rev. 2023;22(10):103409.

39. Faye AS, Lee KE, Laszkowska M, et al. Risk of adverse outcomes in hospitalized patients with autoimmune disease and COVID-19: a matched cohort study from New York City. J Rheumatol. 2021;48(3):454–462.

40. Mehta P, McAuley DF, Brown M, et al. COVID-19: consider cytokine storm syndromes and immunosuppression. Lancet. 2020;395(10229):1033–1034.

41. Singh N, Madhira V, Hu C, et al. Rituximab is associated with worse COVID-19 outcomes in patients with rheumatoid arthritis: a retrospective, nationally sampled cohort study from the U.S. National COVID Cohort Collaborative (N3C). Semin Arthritis Rheum. 2023;58:152149.

42. Knight JS, Caricchio R, Casanova JL, et al. The intersection of COVID-19 and autoimmunity. J Clin Invest. 2021;131(24):e154886.

43. Sun J, Wang S, Ma X, et al. Efficacy and safety of baricitinib for the treatment of hospitalized adults with COVID-19: a systematic review and meta-analysis. Eur J Med Res. 2023;28(1):536.

44. Pinato DJ, Aguilar-Company J, Ferrante D, et al. Outcomes of the SARS-CoV-2 omicron (B.1.1.529) variant outbreak among vaccinated and unvaccinated patients with cancer in Europe: results from the retrospective, multicentre, OnCovid registry study. Lancet Oncol. 2022;23(7):865-875.

45. Yu J, Ouyang W, Chua MLK, Xie C. SARS-CoV-2 transmission in patients with cancer at a tertiary care hospital in Wuhan, China. JAMA Oncol. 2020;6(7):1108–1110.

46. Sinha S, Kundu CN. Cancer and COVID-19: why are cancer patients more susceptible to COVID-19? Med Oncol. 2021;38(9):101.

47. Leyfman Y, Emmanuel N, Menon GP, et al. Cancer and COVID-19: unravelling the immunological interplay with a review of promising therapies against severe SARS-CoV-2 for cancer patients. J Hematol Oncol. 2023;16(1):39.

48. Hardy N, Vegivinti CTR, Mehta M, et al. Mortality of COVID-19 in patients with hematological malignancies versus solid tumors: a systematic literature review and meta-analysis. Clin Exp Med. 2023;23(6):1945–1959.

49. Ketkar A, Willey V, Glasser L, et al. Assessing the burden and cost of COVID-19 across variants in commercially insured immunocompromised populations in the United States: updated results and trends from the ongoing EPOCH-US study. Adv Ther. 2024;41(3):1075–1102.

50. D’Marco L, Puchades MJ, Romero-Parra M, et al. Coronavirus disease 2019 in chronic kidney disease. Clin Kidney J. 2020;13(3):297-306.

51. Rai V. COVID-19 and kidney: the importance of follow-up and long-term screening. Life (Basel). 2023;13(11):2137.

52. Artborg A, Caldinelli A, Wijkstrom J, et al. Risk factors for COVID-19 hospitalization and mortality in patients with chronic kidney disease: a nationwide cohort study. Clin Kidney J. 2024;17(1):sfad283.

53. Gur E, Levy D, Topaz G, et al. Disease severity and renal outcomes of patients with chronic kidney disease infected with COVID-19. Clin Exp Nephrol. 2022;26(5):445–452.

54. Rinaldi M, Bartoletti M, Bussini L, et al. COVID-19 in solid organ transplant recipients: no difference in survival compared to general population. Transpl Infect Dis. 2021;23(1):e13421.

55. Caillard S, Chavarot N, Francois H, et al. Is COVID-19 infection more severe in kidney transplant recipients? Am J Transplant. 2021;21(3):1295–1303.

56. Coll E, Fernandez-Ruiz M, Sanchez-Alvarez JE, et al. COVID-19 in transplant recipients: the Spanish experience. Am J Transplant. 2021;21(5):1825–1837.

57. Ssentongo P, Heilbrunn ES, Ssentongo AE, et al. Epidemiology and outcomes of COVID-19 in HIV-infected individuals: a systematic review and meta-analysis. Sci Rep. 2021;11(1):6283.

58. Danwang C, Noubiap JJ, Robert A, Yombi JC. Outcomes of patients with HIV and COVID-19 co-infection: a systematic review and meta-analysis. AIDS Res Ther. 2022;19(1):3.

59. Boffito M, Waters L. More evidence for worse COVID-19 outcomes in people with HIV. Lancet HIV. 2021;8(11):e661–e662.

60. Centers for Disease Control and Prevention. Vaccines for Moderately to Severely Immunocompromised People. Updated August 30, 2024.https://www.cdc.gov/covid/vaccines/immunocompromised-people.html. Accessed 1 October 2024.

61. Paranilam J, Arcioni F, Franco A, et al. Delphi panel consensus statement generation: COVID-19 vaccination recommendations for immunocompromised populations in the European Union. Infect Dis Ther. 2024;13(11):2227–2253.

62. Serra Lopez-Matencio JM, Vicente-Rabaneda EF, Alanon E, et al. COVID-19 vaccination and immunosuppressive therapy in immune-mediated inflammatory diseases. Vaccines (Basel). 2023;11(12):1813.

63. Curtis JR, Johnson SR, Anthony DD, et al. American College of Rheumatology guidance for COVID-19 vaccination in patients with rheumatic and musculoskeletal diseases: Version 5. Arthritis Rheumatol. 2023;75(1):E1–E16.

64. Curigliano G, Banerjee S, Cervantes A, et al. Managing cancer patients during the COVID-19 pandemic: an ESMO multidisciplinary expert consensus. Ann Oncol. 2020;31(10):1320–1335.

65. Russell CD, Lone NI, Baillie JK. Comorbidities, multimorbidity and COVID-19. Nat Med. 2023;29(2):334–343.

66. Rader B, Gertz A, Iuliano AD, et al. Use of at-home COVID-19 tests - United States, August 23, 2021-March 12, 2022. MMWR Morb Mortal Wkly Rep. 2022;71(13):489-494.

67. Pingali C, Meghani M, Razzaghi H, et al. COVID-19 vaccination coverage among insured persons aged >/=16 years, by race/ethnicity and other selected characteristics - eight integrated health care organizations, United States, December 14, 2020-May 15, 2021. MMWR Morb Mortal Wkly Rep. 2021;70(28):985-990.

68. Wiltz JL, Feehan AK, Molinari NM, et al. Racial and ethnic disparities in receipt of medications for treatment of COVID-19 - United States, March 2020-August 2021. MMWR Morb Mortal Wkly Rep. 2022;71(3):96-102.

69. Agrawal U, Bedston S, McCowan C, et al. Severe COVID-19 outcomes after full vaccination of primary schedule and initial boosters: pooled analysis of national prospective cohort studies of 30 million individuals in England, Northern Ireland, Scotland, and Wales. Lancet. 2022;400(10360):1305–1320.

70. AlBahrani S, AlBarrak A, Al-Musawi T, et al. COVID-19 vaccine had a significant positive impact on patients with SARS-COV-2 during the third (Omicron) wave in Saudi Arabia. J Infect Public Health. 2022;15(11):1169–1174.

71. Arbel R, Sergienko R, Friger M, et al. Effectiveness of a second BNT162b2 booster vaccine against hospitalization and death from COVID-19 in adults aged over 60 years. Nat Med. 2022;28(7):1486–1490.

72. Arbel R, Peretz A, Sergienko R, et al. Effectiveness of a bivalent mRNA vaccine booster dose to prevent severe COVID-19 outcomes: a retrospective cohort study. Lancet Infect Dis. 2023;23(8):914–921.

73. Bahremand T, Yao JA, Mill C, et al. COVID-19 hospitalisations in immunocompromised individuals in the Omicron era: a population-based observational study using surveillance data in British Columbia, Canada. Lancet Reg Health Am. 2023;20:100461.

74. Bedston S, Almaghrabi F, Patterson L, et al. Risk of severe COVID-19 outcomes after autumn 2022 COVID-19 booster vaccinations: a pooled analysis of national prospective cohort studies involving 7.4 million adults in England, Northern Ireland, Scotland and Wales. Lancet Reg Health Eur. 2024;37:100816.

75. Benites-Godínez V, Mendoza-Cano O, Trujillo X, et al. Survival analysis and contributing factors among PCR-confirmed adult inpatients during the endemic phase of COVID-19. Diseases. 2023;11(3):119.

76. Beppu H, Fukuda T, Otsubo N, et al. Comparative outcomes of hemodialysis patients facing pre-Omicron and Omicron COVID-19 epidemics. Ther Apher Dial. 2024;28(1):51–60.

77. Beraud G, Bouetard L, Civljak R, et al. Impact of vaccination on the presence and severity of symptoms in hospitalized patients with an infection of the Omicron variant (B.1.1.529) of the SARS-CoV-2 (subvariant BA.1). Clin Microbiol Infect. 2023;29(5):642-650.

78. Bournia V-K, Fragoulis GE, Mitrou P, et al. Outcomes of COVID-19 Omicron variant in patients with rheumatoid arthritis: a nationwide Greek cohort study. Rheumatology. 2023;63(4):1130–1138.

79. Briciu V, Topan A, Calin M, et al. Comparison of COVID-19 severity in vaccinated and unvaccinated patients during the Delta and Omicron wave of the pandemic in a Romanian tertiary infectious diseases hospital. Healthcare. 2023;11(3):373.

80. Brosh-Nissimov T, Hussein K, Wiener-Well Y, et al. Hospitalized patients with severe coronavirus disease 2019 during the Omicron wave in Israel: benefits of a fourth vaccine dose. Clin Infect Dis. 2022;76(3):e234–e239.

81. Chen Z, Tian F, Zeng Y. Polypharmacy, potentially inappropriate medications, and drug-drug interactions in older COVID-19 inpatients. BMC Geriatrics. 2023;23(1):774.

82. Chen CL, Teng CK, Chen WC, et al. Clinical characteristics and treatment outcomes among the hospitalized elderly patients with COVID-19 during the late pandemic phase in central Taiwan. J Microbiol Immunol Infect. 2024;57(2):257–268.

83. Choi S-H, Choi JH, Lee JK, et al. Clinical characteristics and outcomes of children with SARS-CoV-2 infection during the Delta and Omicron variant-dominant periods in Korea. J Korean Med Sci. 2023;38(9):e65.

84. de Prost N, Audureau E, Heming N, et al. Clinical phenotypes and outcomes associated with SARS-CoV-2 variant Omicron in critically ill French patients with COVID-19. Nat Commun. 2022;13(1):6025.

85. de Prost N, Audureau E, Préau S, et al. Clinical phenotypes and outcomes associated with SARS-CoV-2 Omicron variants BA.2, BA.5 and BQ.1.1 in critically ill patients with COVID-19: a prospective, multicenter cohort study. Intensive Care Med Exp. 2023;11(1):48.

86. Drummond PD, de Salles DB, de Souza NSH, et al. Profile and outcomes of hospitalized COVID-19 patients during the prevalence of the Omicron variant according to the Brazilian regions: a retrospective cohort study from 2022. Vaccines. 2023;11(10):1568.

87. Elamin MY, Maslamani YA, Alsheikh FA, et al. Impact of vaccination on morbidity and mortality in adults hospitalized with COVID-19 during the omicron wave in the Jazan Region, Saudi Arabia. Saudi Med J. 2024;45(2):179–187.

88. Ellis RJ, Moffatt CR, Aaron LT, et al. Factors associated with hospitalisations and deaths of residential aged care residents with COVID-19 during the Omicron (BA.1) wave in Queensland. Med J Aust. 2023;218(4):174-179.

89. Favia G, Barile G, Tempesta A, et al. Relationship between oral lesions and severe SARS-CoV-2 infection in intensive care unit patients. Oral Dis. 2024;30(3):1296–1303.

90. Flacco ME, Acuti Martellucci C, Soldato G, et al. Predictors of SARS-CoV-2 infection and severe and lethal COVID-19 after three years of follow-up: a population-wide study. Viruses. 2023;15(9):1794.

91. Gazit S, Saciuk Y, Perez G, et al. Short term, relative effectiveness of four doses versus three doses of BNT162b2 vaccine in people aged 60 years and older in Israel: retrospective, test negative, case-control study. BMJ. 2022;377:e071113.

92. Griggs EP, Mitchell PK, Lazariu V, et al. Clinical epidemiology and risk factors for critical outcomes among vaccinated and unvaccinated adults hospitalized with COVID-19-VISION Network, 10 states, June 2021-March 2023. Clin Infect Dis. 2024;78(2):338-348.

93. Guo Y, Guo Y, Ying H, et al. In-hospital adverse outcomes and risk factors among chronic kidney disease patients infected with the omicron variant of SARS-CoV-2: a single-center retrospective study. BMC Infect Dis. 2023;23(1):698.

94. Helmy MA, Milad LM, Hasanin AM, et al. Parasternal intercostal thickening at hospital admission: a promising indicator for mechanical ventilation risk in subjects with severe COVID-19. J Clin Monit Comput. 2023;37(5):1287–1293.

95. Hippisley-Cox J, Khunti K, Sheikh A, Nguyen-Van-Tam JS, Coupland CAC. Risk prediction of covid-19 related death or hospital admission in adults testing positive for SARS-CoV-2 infection during the omicron wave in England (QCOVID4): cohort study. BMJ. 2023;381:e072976.

96. Jamaati H, Karimi S, Ghorbani F, et al. Effectiveness of different vaccine platforms in reducing mortality and length of ICU stay in severe and critical cases of COVID-19 in the Omicron variant era: a national cohort study in Iran. J Med Virol. 2023;95(3):e28607.

97. Kang J-M, Kang M, Kim Y-E, et al. Severe coronavirus disease 2019 in pediatric solid organ transplant recipients: big data convergence study in Korea (K-COV-N cohort). Int J Infect Dis. 2023;134:220–227.

98. Karageorgou V, Papaioannou AI, Kallieri M, et al. Patients hospitalized for COVID-19 in the periods of Delta and Omicron variant dominance in Greece: determinants of severity and mortality. J Clin Med. 2023;12(18):5904.

99. Kim SH, Kim T, Choi H, Shin TR, Sim YS. Clinical outcome and prognosis of a nosocomial outbreak of COVID-19. J Clin Med. 2023;12(6):2279.

100. Klein EY, Fall A, Norton JM, et al. Severity outcomes associated with SARS-CoV-2 XBB variants, an observational analysis. J Clin Virol. 2023;165:105500.

101. Konermann FM, Gessler N, Wohlmuth P, et al. High in-hospital mortality in SARS-CoV-2-infected patients with active cancer disease during Omicron phase of the pandemic: insights from the CORONA Germany study. Oncol Res Treat. 2023;46(5):201–210.

102. Lee CM, Kim M, Park SW, et al. Clinical outcomes and immunological features of COVID-19 patients receiving B-cell depletion therapy during the Omicron era. Infect Dis (Lond). 2024;56(2):116–127.

103. Li H, Jia X, Wang Y, et al. Differences in the severity and mortality risk factors for patients hospitalized for COVID-19 pneumonia between the early wave and the very late stage of the pandemic. Front Med (Lausanne). 2023;10:1238713.

104. Li D-J, Zhou C-C, Huang F, Shen F-M, Li Y-C. Clinical features of omicron SARS-CoV-2 variants infection associated with co-infection and ICU-acquired infection in ICU patients. Front Public Health. 2024;11:1320340.

105. Liu Y, Qi Z, Bai M, et al. Combination of chest computed tomography value and clinical laboratory data for the prognostic risk evaluation of patients with COVID-19. Int J Gen Med. 2023;16:3829–3842.

106. Lu G, Zhang Y, Zhang H, et al. Geriatric risk and protective factors for serious COVID-19 outcomes among older adults in Shanghai Omicron wave. Emerg Microbes Infect. 2022;11(1):2045–2054.

107. Mayer C, Woo MS, Brehm TT, et al. History of cerebrovascular disease but not dementia increases the risk for secondary vascular events during SARS-CoV-2 infection with presumed Omicron variant: a retrospective observational study. Eur J Neurol. 2023;30(8):2297–2304.

108. Mendoza-Cano O, Trujillo X, Ríos-Silva M, et al. Association between vaccination status for COVID-19 and the risk of severe symptoms during the endemic phase of the disease. Vaccines. 2023;11(10):1512.

109. Mizrahi Reuveni M, Kertes J, Shapiro Ben David S, et al. Risk stratification model for severe COVID-19 disease: a retrospective cohort study. Biomedicines. 2023;11(3):767.

110. Nevejan L, Ombelet S, Laenen L, et al. Severity of COVID-19 among hospitalized patients: Omicron remains a severe threat for immunocompromised hosts. Viruses. 2022;14(12):2736.

111. O’Leary AL, Wattengel BA, Carter MT, Drye AF, Mergenhagen KA. Risk factors associated with mortality in hospitalized patients with laboratory confirmed SARS-CoV-2 infection during the period of omicron (B.1.1.529) variant predominance. Am J Infect Control. 2023;51(6):603-606.

112. Overvad M, Koch A, Jespersen B, et al. Outcomes following SARS-CoV-2 infection in individuals with and without solid organ transplantation-A Danish nationwide cohort study. Am J Transplant. 2022;22(11):2627–2636.

113. Parajuli P, Sabo R, Alsaadawi R, et al. Fibrosis-4 (FIB-4) index as a predictor for mechanical ventilation and 30-day mortality across COVID-19 variants. J Clin Transl Sci. 2023;7(1):e213.

114. Parra-Bracamonte GM, Lopez-Villalobos N, Velazquez MA, et al. Comparative analysis of risk factors for COVID-19 mortality before, during and after the vaccination programme in Mexico. Public Health. 2023;215:94–99.

115. Patton MJ, Orihuela CJ, Harrod KS, et al. COVID-19 bacteremic co-infection is a major risk factor for mortality, ICU admission, and mechanical ventilation. Crit Care. 2023;27(1):34.

116. Radhakrishnan N, Liu M, Idowu B, et al. Comparison of the clinical characteristics of SARS-CoV-2 Delta (B.1.617.2) and Omicron (B.1.1.529) infected patients from a single hospitalist service. BMC Infect Dis. 2023;23(1):747.

117. Risk M, Hayek SS, Schiopu E, et al. COVID-19 vaccine effectiveness against omicron (B.1.1.529) variant infection and hospitalisation in patients taking immunosuppressive medications: a retrospective cohort study. Lancet Rheumatol. 2022;4(11):e775-e784.

118. Russell SL, Klaver BRA, Harrigan SP, et al. Clinical severity of Omicron subvariants BA.1, BA.2, and BA.5 in a population-based cohort study in British Columbia, Canada. J Med Virol. 2023;95(1):e28423.

119. Shi HJ, Yang J, Eom JS, et al. Clinical characteristics and risk factors for mortality in critical COVID-19 patients aged 50 years or younger during Omicron wave in Korea: comparison with patients older than 50 years of age. J Korean Med Sci. 2023;38(28):e217.

120. Simmons AE, Amoako A, Grima AA, et al. Vaccine effectiveness against hospitalization among adolescent and pediatric SARS-CoV-2 cases between May 2021 and January 2022 in Ontario, Canada: a retrospective cohort study. PLoS One. 2023;18(3):e0283715.

121. Skarbinski J, Wood MS, Chervo TC, et al. Risk of severe clinical outcomes among persons with SARS-CoV-2 infection with differing levels of vaccination during widespread Omicron (B.1.1.529) and Delta (B.1.617.2) variant circulation in Northern California: a retrospective cohort study. Lancet Reg Health Am. 2022;12:100297.

122. Starkey T, Ionescu MC, Tilby M, et al. A population-scale temporal case– control evaluation of COVID-19 disease phenotype and related outcome rates in patients with cancer in England (UKCCP). Sci Rep. 2023;13(1):11327.

123. Svensson ALL, Emborg H-D, Bartels LE, et al. Outcomes following SARS-CoV-2 infection in individuals with and without inflammatory rheumatic diseases: a Danish nationwide cohort study. Ann Rheum Dis. 2023;82(10):1359–1367.

124. Tsujimoto Y, Kobayashi M, Oku T, et al. Outcomes in novel hospital-at-home model for patients with COVID-19: a multicentre retrospective cohort study. Fam Pract. 2023;40(5-6):662–670.

125. Vo AD, La J, Wu JT, et al. Factors associated with severe COVID-19 among vaccinated adults treated in US veterans affairs hospitals. JAMA Netw Open. 2022;5(10):e2240037.

126. Wang X, Zein J, Ji X, Lin DY. Impact of vaccination, prior infection, and therapy on Omicron infection and mortality. J Infect Dis. 2023;227(8):970–976.

127. Ward IL, Robertson C, Agrawal U, et al. Risk of COVID-19 death in adults who received booster COVID-19 vaccinations in England. Nat Commun. 2024;15(1):398.

128. Xin S, Chen W, Yu Q, Gao L, Lu G. Effect of the number of coronavirus disease 2019 (COVID-19) vaccination shots on the occurrence of pneumonia, severe pneumonia, and death in SARS-CoV-2-infected patients. Front Public Health. 2024;11:1330106.

129. Yang H, Wang Z, Zhang Y, et al. Clinical characteristics and factors for serious outcomes among outpatients infected with the Omicron subvariant BF.7. J Med Virol. 2023;95(8):e28977.

130. Zhang Y, Han J, Sun F, et al. A practical scoring model to predict the occurrence of critical illness in hospitalized patients with SARS-CoV-2 omicron infection. Front Microbiol. 2022;13:1031231.

131. Zhao Q, Zheng B, Han B, et al. Is azvudine comparable to nirmatrelvir-ritonavir in real-world efficacy and safety for hospitalized patients with COVID-19? a retrospective cohort study. Infect Dis Ther. 2023;12(8):2087–2102.

132. Zhu Z, Cai J, Tang Q, et al. Circulating eosinophils associated with responsiveness to COVID-19 vaccine and the disease severity in patients with SARS-CoV-2 omicron variant infection. BMC Pulm Med. 2023;23(1):177.

