## Supplemental Materials for "Risk of Severe Outcomes From COVID-19 in Immunocompromised People During the Omicron Era: A Systematic Review and Meta-Analysis"

Contents

1. Supplemental Methods ... 3
1.1.Search Strategy ... 3
1.2. Study Selection ... 21
1.3. Data Extraction ... 22
1.4. Quality Assessment ... 22
1.5. Data Analysis ... 22
1.5.1 Statistical Analysis ... 22
1.5.2 Additional Subgroup Analysis ... 22
2. Supplemental eResults ... 23
2.1 Studies With Insufficient Numbers for Meta-Analysis ... 23
2.2 Risk of Death – Other Subgroup Analyses ... 23
2.3 Risk of ICU – Sensitivity and Subgroup Analyses ... 23
2.4 Risk of Any Combination of Severity Outcomes (Death, Hospitalization or ICU Admission) Among Patients With IC/IS Conditions Compared With Patients Without IC/IS Conditions ... 23
2.5. Supplemental eTables ... 24
2.5.1. Supplemental eTable 1. Inclusion and Exclusion Criteria for Studies Included in This Analysis. ... 24
2.5.2 Supplemental eTable 2. Omicron Period in the Included Studies ... 26
2.5.3. Supplemental eTable 3. Risk of Bias of the Included Studies Using NOS and JBI Tools ... 30
2.5.4. Supplemental eTable 4. Main and Sensitivity Analyses Results of the 'Death' Outcome ... 33
2.5.5. Supplemental eTable 5. Main and Population Subgroup Analyses Results of the 'Death' Outcome for All Population Subgroups of Interest ... 34
2.5.6. Supplemental eTable 6. Main and Sensitivity Analyses Results of the 'Hospitalization' Outcome ... 35
2.5.7. Supplemental eTable 7. Main and Population Subgroup Analyses Results of the 'Hospitalization' Outcome for All Subgroups of Interest ... 36
2.5.8. Supplemental eTable 8. Main and Sensitivity Analyses Results of the 'ICU Admission' Outcome ... 37
2.5.9. Supplemental eTable 9. Main and Population Subgroup Analyses Results of the 'ICU' Outcome for All Population Subgroups of Interest ... 38
2.5.10. Supplemental eTable 10. Main and Sensitivity Analyses Results of the 'Combined' Outcome ... 39
2.5.11. Supplemental eTable 11. Main and Population Subgroup Analyses Results of the 'Combined' Outcome for All Population Subgroups of Interest ... 40
2.6. Meta-analyses for Each IC/IS Condition: Forest Plots of Pooled RR for Each IC/IS Condition and Outcome ... 41
2.6.1. Autoimmune Disease ... 41
2.6.2. Cancer ... 45
2.6.3. HIV ... 49

### 1. Supplemental eMethods

#### 1.1. Search Strategy

The following databases were used for searches: Embase, MEDLINE, PubMed, Europe PMC (including medRix and bioRxiv preprints), Latin American and Caribbean Health Sciences Literature (LILACS), Cochrane COVID-19 Study Register, and the World Health Organization (WHO) COVID-19 Database. Search strategies were structured using search terms related to COVID-19 infection, risk, and burden of illness, in combination with appropriate study design filters, as detailed below. Rolling searches were performed each month for studies available between 1 January 2022 and 13 March 2024, though the WHO COVID-19 database was no longer available before the end date (ceased June 2023). Hand searching was also performed, in which bibliographies of identified studies were checked, and for particularly relevant studies, citation tracking (Google Scholar) was performed.

Searching was carried out using 2 separate search approaches:
Search 1: Burden/risk of COVID-19 infection;
Search 2: Protective effect of vaccination on burden/risk of COVID-19 infection.

The following resources were searched for this systematic review:

| Database | Host | Date range | Date searched | S1: Burden results | S2: Vaccine results |
|---|---|---|---|---|---|
| **January 2024 searches** | | | | | |
| Embase | Ovid | 2022-2023/12/29 | 2.1.24 | 4172 | 1628 |
| Medline ALL | Ovid | 2022-2024/02/02 | 3.1.24 | 3385 | 825 |
| PubMed | NLM | 2022-2024/01/03 | 3.1.24 | 1047 | 281 |
| LILACS | www | 2022-2024/01/04 | 4.1.24 | 38 | 3 |
| Cochrane COVID Register | www | 2022-2024/01/04 | 4.1.24 | 3814 | 1792 |
| Europe PMC, includes bioRxiv & medRxiv | www | 2022-2024/01/04 | 4.1.24 | 10 | 10 |
| WHO COVID-19 database | www | 2022-2024/01/03 | 3.1.24 | 2268 | 477 |
| **January 2024 subtotal** | | | | **14734** | **5016** |
| **February 2024 update searches** | | | | | |
| Embase | Ovid | 2022-2024/02/01 | 1.2.24 | 460 | 110 |
| Medline ALL | Ovid | 2022-2024/02/01 | 1.2.24 | 363 | 51 |
| PubMed | NLM | 2022-2024/02/01 | 1.2.24 | 92 | 20 |
| LILACS | www | 2022-2024/02/01 | 2.2.24 | 3 | 0 |
| Cochrane COVID Register | www | 2022-2024/02/01 | 2.2.24 | 0 | 1 |

| | | | | | |
|---|---|---|---|---|---|
| Europe PMC, includes bioRxiv & medRxiv | www | 2022-2024/02/01 | 1.2.24 | 0 | 1 |
| WHO COVID-19 database* | www | Not updated | - | - | - |
| **February 2024 subtotal** | | | | **918** | **183** |
| **March 2024 update searches** | | | | | |
| Embase | Ovid | 2022-2024/03/01 | 1.3.24 | 404 | 86 |
| 68 | Ovid | 2022-2024/03/01 | 1.3.24 | 386 | 63 |
| PubMed | NLM | 2022-2024/03/01 | 12.3.24 | 68 | 17 |
| LILACS | www | 2022-2024/03/01 | 13.3.24 | 0 | 0 |
| Cochrane COVID Register | www | 2022-2024/03/01 | 13.3.24 | 59 | 0 |
| Europe PMC, includes bioRxiv & medRxiv | www | 2022-2024/03/01 | 1.3.24 | 3 | 0 |
| WHO COVID-19 database* | www | Not updated | - | - | - |
| **March 2024 update subtotal** | | | | **920** | **166** |
| **Sub total retrieved** | | | | **16572** | **5365** |
| **Sub total screened** | | | | **9425** | **2167** |
| **Sub total duplicates** | | | | **7147** | **3198** |
| **Combined total retrieved** | | | | **21937** | |
| **Combined total screened** | | | | **11592** | |
| **Combined total duplicates** | | | | **10345** | |

***** WHO COVID-19 database ceased on June 2023, therefore update searches were not necessary.

**Search 1: Burden/risk of COVID-19 infection search strategies**

**Embase (Ovid): 2022-2023/12/29**
**Searched 2.1.24**

1 exp coronavirus disease 2019/ 374674
2 sars-related coronavirus/ or exp Severe acute respiratory syndrome coronavirus 2/ 106902
3 (coronavirinae/ or betacoronavirus/ or coronavirus infection/) and (epidemic/ or pandemic/) 10615
4 (Coronavirus$ or "covid 19" or 2019-ncov).ti,ab,kw,kf,ot. 430070
5 (2019-ncov or 2019ncov or corona-virus$ or cov19 or cov-19 or 19nCoV or COVID19 or COVID2019 or "COVID-19 2019").ti,ab,kw,kf,ot. 15833
6 (ncov$ or "sars cov$" or sarscov$ or "sars coronavirus$" or coronovirus$ or corono$ virus$ or "19-nCoV$" or 19nCoV$).ti,ab,kw,kf,ot. 160670
7 (SARS2$ or "SARS-2$" or SARScoronavirus$ or SARS-coronavirus$ or SARScoronovirus$ or SARS-coronovirus$).ti,ab,kw,kf,ot. 2922
8 ("HCoV-19$" or HCoV19$ or "HCoV-2019$" or HCoV2019$).ti,ab,kw,kf,ot. 70
9 ("2019 novel$" or Ncov$).ti,ab,kw,kf,ot. 5843
10 ("Severe Acute Respiratory Syndrome Coronavirus 2" or "Severe Acute Respiratory Syndrome Corona Virus 2").ti,ab,kw,kf,ot. 36797
11 or/1-10 496535
12 ("2022" or "2023" or "2024").ti,ab,ot. 235450
13 11 and 12 26927
14 (strain or strains or variant$ or mutation$).ti,ab,ot,kf,kw. 2383716
15 11 and 14 36992
16 exp "SARS-CoV-2 (lineage B.1.1)"/ 8191
17 exp "SARS-CoV-2 Omicron"/ 7250
18 (omikron or Omicron or "B.1.1.529" or "B11529" or xbb$).af. 13566
19 or/16-18 14551
20 or/13,15,19 63058
21 Clinical study/ 165138
22 Case control study/ 211296
23 Family study/ 25764
24 Longitudinal study/ 203445
25 Retrospective study/ 1544157
26 Prospective study/ 899644
27 Randomized controlled trials/ 267337
28 26 not 27 888668
29 Cohort analysis/ 1096585
30 (Cohort adj (study or studies)).mp. 493516
31 (Case control adj (study or studies)).tw. 171865
32 (follow up adj (study or studies)).tw. 74874
33 (observational adj (study or studies)).tw. 264634
34 (epidemiologic$ adj (study or studies)).tw. 123898
35 (cross sectional adj (study or studies)).tw. 356386
36 or/21-25,28-35 4072059
37 animal/ or animal experiment/ 4720382
38 (rat or rats or mouse or mice or murine or rodent or rodents or hamster or hamsters or pig or pigs or porcine or rabbit or rabbits or animal or animals or dogs or dog or cats or cow or bovine or sheep or ovine or monkey or monkeys).ti,ab,ot,hw. 7724982
39 or/37-38 7724982
40 human experiment/ or exp humans/ 25973725
41 39 not (39 and 40) 5781960
42 36 not 41 4009600
43 limit 42 to yr="2022 -Current" 754329
44 43 not (letter or editorial or conference or "conference abstract" or "conference paper" or "conference review").pt. 555478
45 mortality/ or death/ or "cause of death"/ or survival rate/ 1478645

46 (mortalit$ or death$ or fatal$ or survival).ti,ab,ot,kf,kw. 4124730
47 exp intensive care/ or exp intensive care unit/ or high dependency unit/ 1066416
48 (close attention unit$ or intensive care or respiratory care unit or respiratory care units or special care unit or special care units or high dependency unit or high dependency units).ti,ab,ot,kf,kw. 298895
49 (ICU or ICUs or CCU or CCus or GICU or GICUs or HDU or HDUs or ITU or ITUs).ti,ab,ot,kf,kw. 187026
50 (Hospitaliz$ or Hospitalis$ or (hospital adj2 Admission$) or Critical Care).ti,ab,ot,kf,kw. 716741
51 hospitalization/ 546684
52 ("length of stay" or "duration of stay" or "extended stay" or "prolonged stay" or "hospital stay").ti,ab,ot,hw. 412206
53 ("in-patient stay" or "inpatient stay" or "in-patient stays" or "inpatient stays").ti,ab,ot,hw. 7835
54 or/45-53 5826795
55 exp risk/ or exp risk factor/ or exp risk assessment/ 3170934
56 hazard ratio/ or odds ratio/ 99395
57 mortality risk/ 46876
58 life table/ 5253
59 (risk$ or danger$ or association$ or peril or jeopard$ or threat$ or chance or chances or hazard$ or gamble$ or probabilit$ or "at stake" or endanger$ or associat$ or likelihood$ or possibilit$ or correlation$ or odds).ti,ab,ot,kf,kw. 12507127
60 (morbidit$ or comorbid$ or co-morbid$).ti,ab,ot,kf,kw. 1138678
61 (incidence or prevalence or predict$ or prognosis).ti,ab,ot,kf,kw. 5507851
62 (HR or RR or aiRR or aOR or "adjusted OR").ti,ab,ot,kf,kw. 753007
63 or/55-62 15711755
**64 20 and 54 and 63 and 44 4172**

***The Embase strategy was updated on 1.2.24 (460 records) and 1.3.24 (404 records).***

*COVID facet based on terms from:*

World Health Organization (26 May 2021) WHO COVID-19 Database Search Strategy. Systematic search of the COVID-19 literature performed Monday through Friday for the WHO Database. Search strategy as of 26 May 2021. Searches performed by Tomas Allen, Kavita Kothari, and Martha Knuth. Available from: https://www.who.int/docs/default-source/coronaviruse/who-covid-19-database/who-covid-19_sources_searchstrategy_20210526.pdf?sfvrsn=65209cc2_5

Canadian Agency for Drugs and Technologies in Health (2.9.21) CADTH COVID-19 Search Strings: COVID-19 — EMBASE (Internet). Available from: https://covid.cadth.ca/literature-searching-tools/cadth-covid-19-search-strings/

NICE (18 December 2020) [accessed 17.8.21] COVID-19 rapid guideline: managing the long-term effects of COVID-19 [NG188]. Search history record [PDF]. NICE: London. Available from: https://www.nice.org.uk/guidance/ng188/evidence/search-strategies-pdf-8957634445

*Observational study design filter adapted from:*

Scottish Intercollegiate Guidelines Network (SIGN). Search filters: observational studies. Embase. Edinburgh: SIGN, Last Modified 24/04/17 Available from: https://www.sign.ac.uk/what-we-do/methodology/search-filters

Observational Studies - Embase. In: CADTH Search Filters Database. Ottawa: CADTH; 2023 [accessed 7.11.23]: https://searchfilters.cadth.ca/link/37

**Medline ALL (Ovid): 2022-2024/01/02**
**Searched 3.1.24**

1 exp COVID-19/ 250794
2 exp Severe acute respiratory syndrome-related coronavirus/ 166686
3 coronaviridae/ or exp coronavirus/ 178833
4 Betacoronavirus/ 33277

5 Coronavirus Infections/ 46094
6 or/3-5 185838
7 epidemics/ or pandemics/ 139078
8 Disease Outbreaks/ 92374
9 or/7-8 226884
10 6 and 9 83151
11 (Coronavirus$ or "covid 19" or 2019-ncov).ti,ab,kw,kf,ot. 385791
12 (2019-ncov or 2019ncov or corona-virus$ or cov19 or cov-19 or 19nCoV or COVID19 or COVID2019 or "COVID-19 2019").ti,ab,kw,kf,ot. 10202
13 (ncov$ or "sars cov$" or sarscov$ or "sars coronavirus$" or coronovirus$ or corono$ virus$ or "19-nCoV$" or 19nCoV$).ti,ab,kw,kf,ot. 140222
14 (SARS2$ or "SARS-2$" or SARScoronavirus$ or SARS-coronavirus$ or SARScoronovirus$ or SARS-coronovirus$).ti,ab,kw,kf,ot. 2635
15 ("HCoV-19$" or HCoV19$ or "HCoV-2019$" or HCoV2019$).ti,ab,kw,kf,ot. 66
16 ("2019 novel$" or Ncov$).ti,ab,kw,kf,ot. 5236
17 ("Severe Acute Respiratory Syndrome Coronavirus 2" or "Severe Acute Respiratory Syndrome Corona Virus 2").ti,ab,kw,kf,ot. 36907
18 or/1-2,10-17 416995
19 ("2022" or "2023" or "2024").ti,ab,ot. 153808
20 18 and 19 17876
21 (strain or strains or variant$ or mutation$).ti,ab,ot,kf,kw. 1916135
22 18 and 21 32095
23 (omikron or Omicron or "B.1.1.529" or "B11529" or xbb$).af. 9846
24 or/20,22-23 48695
25 mortality/ or "cause of death"/ or fatal outcome/ or hospital mortality/ or survival rate/ 388730
26 Death/ 20522
27 (mortalit$ or death$ or fatal$ or survival).ti,ab,ot,kf,kw. 2881687
28 exp Critical Care/ 67547
29 exp Intensive Care Units/ 107853
30 (close attention unit$ or intensive care or respiratory care unit or respiratory care units or special care unit or special care units or high dependency unit or high dependency units).ti,ab,ot,kf,kw. 202827
31 (ICU or ICUs or CCU or CCus or GICU or GICUs or HDU or HDUs or ITU or ITUs).ti,ab,ot,kf,kw. 95415
32 (Hospitaliz$ or Hospitalis$ or (hospital adj2 Admission$) or Critical Care).ti,ab,ot,kf,kw. 436800
33 Hospitalization/ 137866
34 ("length of stay" or "duration of stay" or "extended stay" or "prolonged stay" or "hospital stay").ti,ab,ot,hw. 220112
35 "Length of Stay"/ 103573
36 ("in-patient stay" or "inpatient stay" or "in-patient stays" or "inpatient stays").ti,ab,ot,hw. 3851
37 or/25-36 3560422
38 exp risk/ 1397708
39 odds ratio/ or proportional hazards models/ 185114
40 life tables/ 6619
41 (risk$ or danger$ or association$ or peril or jeopard$ or threat$ or chance or chances or hazard$ or gamble$ or probabilit$ or "at stake" or endanger$ or associat$ or likelihood$ or possibilit$ or correlation$ or odds).ti,ab,ot,kf,kw. 9312430
42 (morbidit$ or comorbid$ or co-morbid$).ti,ab,ot,kf,kw. 716968
43 (incidence or prevalence or predict$ or prognosis).ti,ab,ot,kf,kw. 3993128
44 (HR or RR or aiRR or aOR or "adjusted OR").ti,ab,ot,kf,kw. 471257
45 or/38-44 11628532
46 Epidemiologic studies/ 9457
47 exp case control studies/ 1470108
48 exp cohort studies/ 2556574
49 Case control.tw. 158730
50 Longitudinal.tw. 335439
51 Retrospective.tw. 782068
52 Cross sectional.tw. 540774
53 Cross-sectional studies/ 487981

54 Observational Studies as Topic/ 9294
55 (observational adj3 (study or studies or design or analysis or analyses)).ti,ab,kf. 234202
56 ((follow up or followup) adj7 (study or studies or design or analysis or analyses)).ti,ab,kf. 175238
57 (epidemiologic$ adj3 (study or studies or design or analysis or analyses)).ti,ab,kf. 119921
58 ((longterm or (long adj term)) adj7 (study or studies or design or analysis or analyses or data)).ti,ab,kf. 160631
59 (cohort$ adj3 (study or studies or design or analysis or analyses or data)).ti,ab,kf. 420054
60 (prospective adj7 (study or studies or design or analysis or analyses)).ti,ab,kf. 551875
61 or/46-60 4243864
62 animals/ not (animals/ and humans/) 5148855
63 61 not 62 4139882
64 63 not (case reports or clinical conference or comment or editorial or letter).pt. 3929840
65 limit 64 to yr="2022 -Current" 538850
**66 24 and 37 and 45 and 65 3385**

***The Medline ALL strategy was updated on 1.2.24 (363 records) and 1.3.24 (363 records).***

*COVID facet based on terms from:*

World Health Organization (26 May 2021) WHO COVID-19 Database Search Strategy. Systematic search of the COVID-19 literature performed Monday through Friday for the WHO Database. Search strategy as of 26 May 2021. Searches performed by Tomas Allen, Kavita Kothari, and Martha Knuth. Available from: https://www.who.int/docs/default-source/coronaviruse/who-covid-19-database/who-covid-19_sources_searchstrategy_20210526.pdf?sfvrsn=65209cc2_5

Canadian Agency for Drugs and Technologies in Health (2.9.21) CADTH COVID-19 Search Strings: COVID-19 — EMBASE (Internet). Available from: https://covid.cadth.ca/literature-searching-tools/cadth-covid-19-search-strings/

NICE (18 December 2020) [accessed 17.8.21] COVID-19 rapid guideline: managing the long-term effects of COVID-19 [NG188]. Search history record [PDF]. NICE: London. Available from: https://www.nice.org.uk/guidance/ng188/evidence/search-strategies-pdf-8957634445

*Observational study design filter adapted from:*

Scottish Intercollegiate Guidelines Network (SIGN). Search filters: observational studies. Medline. Edinburgh: SIGN, Last Modified 24/04/17 Available from: https://www.sign.ac.uk/what-we-do/methodology/search-filters/

Observational Studies - Medline. In: CADTH Search Filters Database. Ottawa: CADTH; 2023 [accessed 7.11.23]: https://searchfilters.cadth.ca/list?q=topic%3A%22Observational%20studies%22&p=1&ps=&sort=title_sort%20asc

**PubMed (NLM): 2022-2024/01/03**
**Searched 3.1.24**
**https://pubmed.ncbi.nlm.nih.gov/**

**35 #13 AND #30 AND #34 AND #24 AND #25 1,047**
34 #31 OR #32 OR #33 9,159,936
33 morbidity[Title/Abstract] OR comorbidity[Title/Abstract] OR "co-morbidity"[Title/Abstract] OR morbidities[Title/Abstract] OR comorbidities[Title/Abstract] OR "co-morbidities"[Title/Abstract] OR incidence[Title/Abstract] OR prevalence[Title/Abstract] OR predict[Title/Abstract] OR predicted[Title/Abstract] OR predictive[Title/Abstract] OR prognosis[Title/Abstract] OR prediction[Title/Abstract] OR predictions[Title/Abstract] OR HR[Title/Abstract] OR RR[Title/Abstract] OR aiRR[Title/Abstract] OR aOR[Title/Abstract] OR "adjusted OR"[Title/Abstract] 4,362,222
32 risk[Title/Abstract] OR risks[Title/Abstract] OR risky[Title/Abstract] OR danger[Title/Abstract] OR dangers[Title/Abstract] OR dangerous[Title/Abstract] OR association[Title/Abstract] OR associations[Title/Abstract] OR peril[Title/Abstract] OR jeopardy[Title/Abstract] OR jeopardise[Title/Abstract]

OR jeopardize[Title/Abstract] OR threat[Title/Abstract] OR threats[Title/Abstract] OR chance[Title/Abstract] OR chances[Title/Abstract] OR hazard[Title/Abstract] OR hazards[Title/Abstract] OR hazardous[Title/Abstract] OR gamble[Title/Abstract] OR gambles[Title/Abstract] OR gambling[Title/Abstract] OR probability[Title/Abstract] OR probabilities[Title/Abstract] OR "at stake"[Title/Abstract] OR endanger[Title/Abstract] OR endangers[Title/Abstract] OR endangered[Title/Abstract] OR endangering[Title/Abstract] OR associate[Title/Abstract] OR associates[Title/Abstract] OR associating[Title/Abstract] OR association[Title/Abstract] OR likelihood[Title/Abstract] OR likelihoods[Title/Abstract] OR possibility[Title/Abstract] OR possibilities[Title/Abstract] OR correlation[Title/Abstract] OR correlations[Title/Abstract] OR odds[Title/Abstract] 6,440,552

31 ((("Risk"[Mesh]) OR "Odds Ratio"[Mesh:NoExp]) OR "Proportional Hazards Models"[Mesh:NoExp]) OR "Life Tables"[Mesh:NoExp] 1,481,626

30 #26 OR #27 OR #28 OR #29 3,581,605

29 Hospitalized[Title/Abstract] OR Hospitalised population[Title/Abstract] OR Hospitalization[Title/Abstract] OR Hospitalisation[Title/Abstract] OR Hospitalizations[Title/Abstract] OR Hospitalisations[Title/Abstract] OR "hospital Admission"[Title/Abstract] OR "hospital Admissions"[Title/Abstract] OR "Critical Care"[Title/Abstract] OR "length of stay"[Title/Abstract] OR "duration of stay"[Title/Abstract] OR "extended stay"[Title/Abstract] OR "prolonged stay"[Title/Abstract] OR "hospital stay"[Title/Abstract] OR "in-patient stay"[Title/Abstract] OR "inpatient stay"[Title/Abstract] OR "in-patient stays"[Title/Abstract] OR "inpatient stays"[Title/Abstract] 565,867

28 mortality[Title/Abstract] OR death[Title/Abstract] OR deaths[Title/Abstract] OR fatal[Title/Abstract] OR fatality[Title/Abstract] OR fatalities[Title/Abstract] OR survival[Title/Abstract] OR "close attention unit"[Title/Abstract] OR "close attention units"[Title/Abstract] OR "intensive care"[Title/Abstract] OR "respiratory care unit"[Title/Abstract] OR "respiratory care units"[Title/Abstract] OR "special care unit"[Title/Abstract] OR "special care units"[Title/Abstract] OR "high dependency unit"[Title/Abstract] OR "high dependency units"[Title/Abstract] OR ICU[Title/Abstract] OR ICUs[Title/Abstract] OR CCU[Title/Abstract] OR CCus[Title/Abstract] OR GICU[Title/Abstract] OR GICUs[Title/Abstract] OR HDU[Title/Abstract] OR HDUs[Title/Abstract] OR ITU[Title/Abstract] OR ITUs[Title/Abstract] 3,013,975

27 ((("Length of Stay"[Mesh:NoExp]) OR "Critical Care"[Mesh]) OR "Intensive Care Units"[Mesh]) OR "Hospitalization"[Mesh] 429,629

26 ((((("Mortality"[Mesh:NoExp]) OR "Cause of Death"[Mesh:NoExp]) OR "Fatal Outcome"[Mesh:NoExp]) OR "Hospital Mortality"[Mesh:NoExp]) OR "Survival Rate"[Mesh:NoExp]) OR "Death"[Mesh:NoExp] 408,041

25 pubstatusaheadofprint OR publisher[sb] OR pubmednotmedline[sb] 5,627,356

24 #22 AND #23 493,476

23 ("2022/01/01"[Date - Publication] : "3000"[Date - Publication]) 3,253,097

22 #20 NOT #21 3,739,291

21 LETTER[Publication Type] OR EDITORIAL[Publication Type] OR COMMENT[Publication Type] 2,213,615

20 #16 NOT #19 3,825,199

19 #18 NOT (#18 AND #17) 3,757,858

18 rat[tiab] or rats[tiab] or mouse[tiab] or mice[tiab] or murine[tiab] or rodent[tiab] or rodents[tiab] or hamster[tiab] or hamsters[tiab] or pig[tiab] or pigs[tiab] or porcine[tiab] or rabbit[tiab] or rabbits[tiab] or animal[tiab] or animals[tiab] or dogs[tiab] or dog[tiab] or cats[tiab] or cow[tiab] or bovine[tiab] or sheep[tiab] or ovine[tiab] or monkey[tiab] or monkeys[tiab] 4,715,684

17 Human*[tiab] 3,295,660

16 #14 OR #15 3,914,514

15 ((("Epidemiologic Studies"[Mesh:NoExp]) OR "Case-Control Studies"[Mesh]) OR "Cohort Studies"[Mesh]) OR "Cross-Sectional Studies"[Mesh:NoExp] 3,195,884

14 "Case control"[Title/Abstract] OR "cohort study"[Title/Abstract] OR "cohort studies"[Title/Abstract] OR "cohort analysis"[Title/Abstract] OR "cohort analyses"[Title/Abstract] OR "follow up study"[Title/Abstract] OR "follow up studies"[Title/Abstract] OR "observational study"[Title/Abstract] OR "observational studies"[Title/Abstract] OR Longitudinal[Title/Abstract] OR Retrospective[Title/Abstract] OR "Cross sectional"[Title/Abstract] 2,068,573

13 #9 OR #11 OR #12 47,832

12 omikron[Title/Abstract] OR Omicron[Title/Abstract] OR "B.1.1.529"[Title/Abstract] OR "B11529"[Title/Abstract] OR xbb[Title/Abstract] 9,468

11 #7 AND #10 31,553

10 strain[Title/Abstract] OR strains[Title/Abstract] OR variant[Title/Abstract] OR variants[Title/Abstract] OR mutation[Title/Abstract] OR mutations[Title/Abstract] 1,893,774
9 #7 AND #8 17,832
8 "2022"[Title/Abstract] OR "2023"[Title/Abstract] 158,464
7 #1 OR #4 OR #5 OR #6 412,676
6 "COVID-19 2019"[Title/Abstract] OR ncov[Title/Abstract] OR "sars cov"[Title/Abstract] OR sarscov[Title/Abstract] OR "sars coronavirus"[Title/Abstract] OR coronovirus[Title/Abstract] OR "corono virus"[Title/Abstract] OR "19-nCoV"[Title/Abstract] OR "19nCoV"[Title/Abstract] OR "SARS2"[Title/Abstract] OR "SARS-2"[Title/Abstract] OR SARScoronavirus[Title/Abstract] OR "SARS-coronavirus"[Title/Abstract] OR SARScoronovirus[Title/Abstract] OR "SARS-coronovirus"[Title/Abstract] OR "HCoV-19"[Title/Abstract] OR "HCoV19"[Title/Abstract] OR "HCoV-2019"[Title/Abstract] OR "HCoV2019"[Title/Abstract] OR "2019 novel"[Title/Abstract] OR "Severe Acute Respiratory Syndrome Coronavirus 2"[Title/Abstract] OR "Severe Acute Respiratory Syndrome Corona Virus 2"[Title/Abstract] 141,123
5 "Coronavirus"[Title/Abstract] OR "covid 19"[Title/Abstract] OR "2019-ncov"[Title/Abstract] OR "2019-ncov"[Title/Abstract] OR "2019ncov"[Title/Abstract] OR "corona-virus"[Title/Abstract] OR "cov19"[Title/Abstract] OR "cov-19"[Title/Abstract] OR "19nCoV"[Title/Abstract] OR "COVID19"[Title/Abstract] OR "COVID2019"[Title/Abstract] 382,642
4 #2 AND #3 83,020
3 (("Epidemics"[Mesh:NoExp]) OR "Pandemics"[Mesh:NoExp]) OR "Disease Outbreaks"[Mesh:NoExp] 226,548
2 ((("Coronaviridae"[Mesh:NoExp]) OR "Coronavirus"[Mesh]) OR "Betacoronavirus"[Mesh:NoExp]) OR "Coronavirus Infections"[Mesh:NoExp] 185,366
1 ("COVID-19"[Mesh]) OR "Severe acute respiratory syndrome-related coronavirus"[Mesh] 258,558

***The PubMed strategy was updated on 1.2.24 (92 records) and 1.3.24 (68 records).***

*COVID facet based on terms from:*

World Health Organization (26 May 2021) WHO COVID-19 Database Search Strategy. Systematic search of the COVID-19 literature performed Monday through Friday for the WHO Database. Search strategy as of 26 May 2021. Searches performed by Tomas Allen, Kavita Kothari, and Martha Knuth. Available from: https://www.who.int/docs/default-source/coronaviruse/who-covid-19-database/who-covid-19_sources_searchstrategy_20210526.pdf?sfvrsn=65209cc2_5

Canadian Agency for Drugs and Technologies in Health (2.9.21) CADTH COVID-19 Search Strings: COVID-19 — EMBASE (Internet). Available from: https://covid.cadth.ca/literature-searching-tools/cadth-covid-19-search-strings/

NICE (18 December 2020) [accessed 17.8.21] COVID-19 rapid guideline: managing the long-term effects of COVID-19 [NG188]. Search history record [PDF]. NICE: London. Available from: https://www.nice.org.uk/guidance/ng188/evidence/search-strategies-pdf-8957634445

*Observational study design filter adapted from:*
Scottish Intercollegiate Guidelines Network (SIGN). Search filters: observational studies. Medline. Edinburgh: SIGN, Last Modified 24/04/17 Available from: https://www.sign.ac.uk/what-we-do/methodology/search-filters/

*PubMed limit:*
Duffy S, de Kock S, Misso K, Noake C, Ross J, Stirk L. Supplementary searches of PubMed to improve currency of MEDLINE and MEDLINE In-Process searches via Ovid. J Med Libr Assoc. 2016 Oct;104(4):309-312. doi: 10.3163/1536-5050.104.4.011. https://www.ncbi.nlm.nih.gov/pmc/articles/PMC5079494/

**Europe PMC, including medRxiv and bioRxiv preprints (Internet): up to 2024/01/04**
**Searched 4.1.24**
**https://europepmc.org/**

| **Search terms** | **Results** |
|---|---|
| (TITLE:"COVID-19" OR TITLE:"coronavirus" OR TITLE:"COVID" OR TITLE:"NCOV" OR TITLE:"SARS-CoV-2") AND (TITLE:"risk" OR TITLE:"risks" OR TITLE:"hazard" OR TITLE:"hazards" OR TITLE:"probability" OR TITLE:" probabilities" OR TITLE:"likelihood" OR TITLE:"morbidity" OR TITLE:"comorbidity" OR TITLE:"predict" OR TITLE:"predictive" OR TITLE:"HR" OR TITLE:"RR" OR TITLE:"aiRR" OR TITLE:"aOR" OR TITLE:"adjusted OR") AND (TITLE:"2022" OR TITLE:"2023" OR TITLE:"strain" OR TITLE:"strains" OR TITLE:"variant" OR TITLE:"variants" OR TITLE:"mutation" OR TITLE:"mutations" OR TITLE:"omicron" OR TITLE:"omicron") | **89** |

***The Europe PMC strategy was updated on 1.2.24 (0 records) and 1.3.24 (3 records).***

**Latin American and Caribbean Health Sciences Literature (LILACS) (Internet): 2022-2024/01/07**
**Searched 4.1.24**
https://search.bvsalud.org/portal/?lang=en

Searched Title/Abstract/Subject
Limited to Observational studies
Limited to 2022-2024/01/04
Limited to LILACS only

((COVID OR NCOV OR coronavirus OR COVID19 OR "SARs-COV-2" OR Omicron OR omicron)) AND ((mortality OR death OR deaths OR fatal OR fatality OR fatalities OR survival OR "close attention unit" OR "close attention units" OR "intensive care" OR "respiratory care unit" OR "respiratory care units" OR "special care unit" OR "special care units" OR "high dependency unit" OR "high dependency units" OR ICU OR ICUs OR CCU OR CCus OR GICU OR GICUs OR HDU OR HDUs OR ITU OR ITUs OR Hospitalized OR Hospitalised population OR Hospitalization OR Hospitalisation OR Hospitalizations OR Hospitalisations OR "hospital Admission" OR "hospital Admissions" OR "Critical Care" OR "length of stay" OR "duration of stay" OR "extended stay" OR "prolonged stay" OR "hospital stay" OR "in-patient stay" OR "inpatient stay" OR "in-patient stays" OR "inpatient stays") ) AND ((risk OR risks OR risky OR danger OR dangers OR dangerous OR association OR associations OR peril OR jeopardy OR jeopardise OR jeopardize OR threat OR threats OR chance OR chances OR hazard OR hazards OR hazardous OR gamble OR gambles OR gambling OR probability OR probabilities OR "at stake" OR endanger OR endangers OR endangered OR endangering OR associate OR associates OR associating OR association OR likelihood OR likelihoods OR possibility OR possibilities OR correlation OR correlations OR odds OR morbidity OR comorbidity OR "co-morbidity" OR morbidities OR comorbidities OR "co-morbidities" OR incidence OR prevalence OR predict OR predicted OR predictive OR prognosis OR prediction OR predictions OR HR OR RR OR aiRR OR aOR OR "adjusted OR")) AND ((2022 OR 2023 OR 2024 OR strain OR strains OR variant OR variants OR mutations OR mutation OR omikron OR Omicron OR "B.1.1.529" OR "B11529" OR xbb) )

**N=38**

***The LILACS strategy was updated on 1.2.24 (3 records) and 1.3.24 (0 records).***

**Cochrane COVID-19 Study Register (www): 2022-2024/01/04**
**Searched 4.1.24**
https://covid-19.cochrane.org/

Limited to Observational cohort studies.
Limited 2022-2024/01/04

(mortality OR death OR deaths OR fatal OR fatality OR fatalities OR survival OR "close attention unit" OR "close attention units" OR "intensive care" OR "respiratory care unit" OR "respiratory care units" OR "special care unit" OR "special care units" OR "high dependency unit" OR "high dependency units" OR ICU OR ICUs OR CCU OR CCus OR GICU OR GICUs OR HDU OR HDUs OR ITU OR ITUs OR Hospitalized OR Hospitalised population OR Hospitalization OR Hospitalisation OR Hospitalizations OR Hospitalisations OR "hospital Admission" OR "hospital Admissions" OR "Critical Care" OR "length of stay" OR "duration of stay" OR "extended stay" OR "prolonged stay" OR "hospital stay" OR "in-patient stay" OR "inpatient stay" OR "in-patient stays" OR "inpatient stays") AND (risk OR risks OR risky OR danger OR dangers OR dangerous OR association OR associations OR peril OR jeopardy OR jeopardise OR jeopardize OR threat OR threats OR chance OR chances OR hazard OR hazards OR hazardous OR gamble OR gambles OR gambling OR probability OR probabilities OR "at stake" OR endanger OR endangers OR endangered OR endangering OR associate OR associates OR associating OR association OR likelihood OR likelihoods OR possibility OR possibilities OR correlation OR correlations OR odds OR morbidity OR comorbidity OR "co-morbidity" OR morbidities OR comorbidities OR "co-morbidities" OR incidence OR prevalence OR predict OR predicted OR predictive OR prognosis OR prediction OR predictions OR HR OR RR OR aiRR OR aOR OR "adjusted OR") AND (2022 OR 2023 OR 2024 OR strain OR strains OR variant OR variants OR mutations OR mutation OR omikron OR Omicron OR "B.1.1.529" OR "B11529" OR xbb) AND (cohort OR cohorts OR observational OR "follow up" OR followup OR epidemiologic OR epidemiological OR longterm OR "long term" OR longitudinal OR prospective)

**N=3814**

***The Cochrane COVID-19 Study Register strategy was updated on 1.2.24 (0 records) and 1.3.24 (59 records).***

**WHO COVID-19 (Internet): 2022-2024/01/03**
**Searched 3.1.24**
**https://search.bvsalud.org/global-literature-on-novel-coronavirus-2019-ncov/?lang=en**

Advanced search (title only)

(ti:(mortality OR death OR deaths OR fatal OR fatality OR fatalities OR survival OR "close attention unit" OR "close attention units" OR "intensive care" OR "respiratory care unit" OR "respiratory care units" OR "special care unit" OR "special care units" OR "high dependency unit" OR "high dependency units" OR ICU OR ICUs OR CCU OR CCus OR GICU OR GICUs OR HDU OR HDUs OR ITU OR ITUs OR Hospitalized OR Hospitalised population OR Hospitalization OR Hospitalisation OR Hospitalizations OR Hospitalisations OR "hospital Admission" OR "hospital Admissions" OR "Critical Care" OR "length of stay" OR "duration of stay" OR "extended stay" OR "prolonged stay" OR "hospital stay" OR "in-patient stay" OR "inpatient stay" OR "in-patient stays" OR "inpatient stays")) AND (ti:(risk OR risks OR risky OR danger OR dangers OR dangerous OR association OR associations OR peril OR jeopardy OR jeopardise OR jeopardize OR threat OR threats OR chance OR chances OR hazard OR hazards OR hazardous OR gamble OR gambles OR gambling OR probability OR probabilities OR "at stake" OR endanger OR endangers OR endangered OR endangering OR associate OR associates OR associating OR association OR likelihood OR likelihoods OR possibility OR possibilities OR correlation OR correlations OR odds OR morbidity OR comorbidity OR "co-morbidity" OR morbidities OR comorbidities OR "co-morbidities" OR incidence OR prevalence OR predict OR predicted OR predictive OR prognosis OR prediction OR predictions OR HR OR RR OR aiRR OR aOR OR "adjusted OR"))

Limited to Observational and cohort studies.
Limited 2022-2023/06.

**N=2268**

***** WHO COVID-19 database ceased on June 2023, therefore update searches were not necessary.

**Search 2: Protective effect of vaccination on burden/risk of COVID-19 infection search strategies**

**Embase (Ovid): 2022-2023/12/29**
**Searched 2.1.24**

1 exp coronavirus disease 2019/ 374674
2 sars-related coronavirus/ or exp Severe acute respiratory syndrome coronavirus 2/ 106902
3 (coronavirinae/ or betacoronavirus/ or coronavirus infection/) and (epidemic/ or pandemic/) 10615
4 (Coronavirus$ or "covid 19" or 2019-ncov).ti,ab,kw,kf,ot. 430070
5 (2019-ncov or 2019ncov or corona-virus$ or cov19 or cov-19 or 19nCoV or COVID19 or COVID2019 or "COVID-19 2019").ti,ab,kw,kf,ot. 15833
6 (ncov$ or "sars cov$" or sarscov$ or "sars coronavirus$" or coronovirus$ or corono$ virus$ or "19-nCoV$" or 19nCoV$).ti,ab,kw,kf,ot. 160670
7 (SARS2$ or "SARS-2$" or SARScoronavirus$ or SARS-coronavirus$ or SARScoronovirus$ or SARS-coronovirus$).ti,ab,kw,kf,ot. 2922
8 ("HCoV-19$" or HCoV19$ or "HCoV-2019$" or HCoV2019$).ti,ab,kw,kf,ot. 70
9 ("2019 novel$" or Ncov$).ti,ab,kw,kf,ot. 5843
10 ("Severe Acute Respiratory Syndrome Coronavirus 2" or "Severe Acute Respiratory Syndrome Corona Virus 2").ti,ab,kw,kf,ot. 36797
11 or/1-10 496535
12 ("2022" or "2023").ti,ab,ot. 235450
13 11 and 12 26927
14 (strain or strains or variant$ or mutation$).ti,ab,ot,kf,kw. 2383716
15 11 and 14 36992
16 exp "SARS-CoV-2 (lineage B.1.1)"/ 8191
17 exp "SARS-CoV-2 Omicron"/ 7250
18 (omikron or Omicron or "B.1.1.529" or "B11529" or xbb$).af. 13566
19 or/16-18 14551
20 or/13,15,19 63058
21 Clinical study/ 165138
22 Case control study/ 211296
23 Family study/ 25764
24 Longitudinal study/ 203445
25 Retrospective study/ 1544157
26 Prospective study/ 899644
27 Randomized controlled trials/ 267337
28 26 not 27 888668
29 Cohort analysis/ 1096585
30 (Cohort adj (study or studies)).mp. 493516
31 (Case control adj (study or studies)).tw. 171865
32 (follow up adj (study or studies)).tw. 74874
33 (observational adj (study or studies)).tw. 264634
34 (epidemiologic$ adj (study or studies)).tw. 123898
35 (cross sectional adj (study or studies)).tw. 356386
36 or/21-25,28-35 4072059
37 animal/ or animal experiment/ 4720382
38 (rat or rats or mouse or mice or murine or rodent or rodents or hamster or hamsters or pig or pigs or porcine or rabbit or rabbits or animal or animals or dogs or dog or cats or cow or bovine or sheep or ovine or monkey or monkeys).ti,ab,ot,hw. 7724982
39 or/37-38 7724982
40 human experiment/ or exp humans/ 25973725
41 39 not (39 and 40) 5781960
42 36 not 41 4009600
43 limit 42 to yr="2022 -Current" 754329
44 43 not (letter or editorial or conference or "conference abstract" or "conference paper" or "conference review").pt. 555478
45 mortality/ or death/ or "cause of death"/ or survival rate/ 1478645

46 (mortalit$ or death$ or fatal$ or survival).ti,ab,ot,kf,kw. 4124730
47 exp intensive care/ or exp intensive care unit/ or high dependency unit/ 1066416
48 (close attention unit$ or intensive care or respiratory care unit or respiratory care units or special care unit or special care units or high dependency unit or high dependency units).ti,ab,ot,kf,kw. 298895
49 (ICU or ICUs or CCU or CCus or GICU or GICUs or HDU or HDUs or ITU or ITUs).ti,ab,ot,kf,kw. 187026
50 (Hospitaliz$ or Hospitalis$ or (hospital adj2 Admission$) or Critical Care).ti,ab,ot,kf,kw. 716741
51 hospitalization/ 546684
52 ("length of stay" or "duration of stay" or "extended stay" or "prolonged stay" or "hospital stay").ti,ab,ot,hw. 412206
53 ("in-patient stay" or "inpatient stay" or "in-patient stays" or "inpatient stays").ti,ab,ot,hw. 7835
54 exp risk/ or exp risk factor/ or exp risk assessment/ 3170934
55 hazard ratio/ or odds ratio/ 99395
56 mortality risk/ 46876
57 (risk$ or danger$ or association$ or peril or jeopard$ or threat$ or chance or chances or hazard$ or gamble$ or probabilit$ or "at stake" or endanger$ or associat$ or likelihood$ or possibilit$ or correlation$ or odds or "vaccine effectiveness" or "immune evasion" or "immunity evasion").ti,ab,ot,kf,kw. 12518691
58 (morbidit$ or comorbid$ or co-morbid$).ti,ab,ot,kf,kw. 1138678
59 (incidence or prevalence or predict$ or prognosis).ti,ab,ot,kf,kw. 5507851
60 (HR or RR or aiRR or aOR or "adjusted OR").ti,ab,ot,kf,kw. 753007
61 disease severity assessment/ or covid-19 severity score/ or global severity index/ or quick covid-19 severity index/ or "severity of illness index"/ 32588
62 morbidity/ 416716
63 disease severity/ or critical illness/ or emergency status/ or urgency status/ 772357
64 ((severe or severity) adj3 (disease$ or infection$ or illness$ or complication$ or condition$ or illhealth$ or ill-health$ or sickness or virus or infirmity or affliction$)).ti,ab,ot,kf,kw. 423477
65 morbidit$.ti,ab,ot,kf,kw. 754095
66 or/45-65 18057098
67 Tozinameran/ 11540
68 ((pfizer$ or biontech) adj5 vaccin$).ti,ab,ot,kf,kw,hw,du,dy,tn,rn. 4443
69 (tozinameran or Comirnaty or "Pfizer-BioNTech" or "pf07302048" or "pf-07302048" or " BNT-162b2" or "BNT162b2" or Pidacmeran or "BNT162C2" or "BNT-162C2" or Abdavomeran or "BNT-162B1" or "BNT162B1" or "BNT162A1" or "BNT-162A1").ti,ab,ot,kf,kw,hw,du,dy,tn,rn. 15132
70 "Pfizer-BioNTech".mf. 947
71 "2417899-77-3".af. 12357
72 elasomeran/ 6806
73 (Elasomeran or moderna or Spikevax or davesomeran or imelasomeran or andusomeran or "mRNA 1273" or "mRNA1273" or "mRNA-1273.211" or "mRNA 1273.211" or Spikevax or "M-1273" or "M1273" or "CX-024414" or "CX024414" or "TAK-919" or "TAK919").ti,ab,ot,kf,kw,hw,du,dy,tn,rn. 9771
74 ("2430046-03-8" or "2457298-05-2").af. 6468
75 RNA vaccine/ 5844
76 (("messenger RNA" or mrna or "RNA based") adj4 (vaccin$ or jab or jabs or shot or shots or immunis$ or immuniz$)).ti,ab,ot,kf,kw. 12887
77 or/67-76 25983
**78 and/20,44,66,77 1628**

***The Embase strategy was updated on 1.2.24 (110 records) and 1.3.24 (86 records).***

*COVID facet based on terms from:*

World Health Organization (26 May 2021) WHO COVID-19 Database Search Strategy. Systematic search of the COVID-19 literature performed Monday through Friday for the WHO Database. Search strategy as of 26 May 2021. Searches performed by Tomas Allen, Kavita Kothari, and Martha Knuth. Available from: https://www.who.int/docs/default-source/coronaviruse/who-covid-19-database/who-covid-19_sources_searchstrategy_20210526.pdf?sfvrsn=65209cc2_5

Canadian Agency for Drugs and Technologies in Health (2.9.21) CADTH COVID-19 Search Strings: COVID-19 — EMBASE (Internet). Available from: https://covid.cadth.ca/literature-searching-tools/cadth-covid-19-search-strings/

NICE (18 December 2020) [accessed 17.8.21] COVID-19 rapid guideline: managing the long-term effects of COVID-19 [NG188]. Search history record [PDF]. NICE: London. Available from: https://www.nice.org.uk/guidance/ng188/evidence/search-strategies-pdf-8957634445

*Observational study design filter adapted from:*

Scottish Intercollegiate Guidelines Network (SIGN). Search filters: observational studies. Embase. Edinburgh: SIGN, Last Modified 24/04/17 Available from: https://www.sign.ac.uk/what-we-do/methodology/search-filters/

Observational Studies - Embase. In: CADTH Search Filters Database. Ottawa: CADTH; 2023 [accessed 7.11.23]: https://searchfilters.cadth.ca/link/37

**Medline ALL (Ovid): 2022-2024/01/02**
**Searched 2.1.24**

| | | |
|---|---|---|
| 1 | exp COVID-19/ | 250794 |
| 2 | exp Severe acute respiratory syndrome-related coronavirus/ | 166686 |
| 3 | coronaviridae/ or exp coronavirus/ | 178833 |
| 4 | Betacoronavirus/ | 33277 |
| 5 | Coronavirus Infections/ | 46094 |
| 6 | or/3-5 | 185838 |
| 7 | epidemics/ or pandemics/ | 139078 |
| 8 | Disease Outbreaks/ | 92374 |
| 9 | or/7-8 | 226884 |
| 10 | 6 and 9 | 83151 |
| 11 | (Coronavirus$ or "covid 19" or 2019-ncov).ti,ab,kw,kf,ot. | 385791 |
| 12 | (2019-ncov or 2019ncov or corona-virus$ or cov19 or cov-19 or 19nCoV or COVID19 or COVID2019 or "COVID-19 2019").ti,ab,kw,kf,ot. | 10202 |
| 13 | (ncov$ or "sars cov$" or sarscov$ or "sars coronavirus$" or coronovirus$ or corono$ virus$ or "19-nCoV$" or 19nCoV$).ti,ab,kw,kf,ot. | 140222 |
| 14 | (SARS2$ or "SARS-2$" or SARScoronavirus$ or SARS-coronavirus$ or SARScoronovirus$ or SARS-coronovirus$).ti,ab,kw,kf,ot. | 2635 |
| 15 | ("HCoV-19$" or HCoV19$ or "HCoV-2019$" or HCoV2019$).ti,ab,kw,kf,ot. | 66 |
| 16 | ("2019 novel$" or Ncov$).ti,ab,kw,kf,ot. | 5236 |
| 17 | ("Severe Acute Respiratory Syndrome Coronavirus 2" or "Severe Acute Respiratory Syndrome Corona Virus 2").ti,ab,kw,kf,ot. | 36907 |
| 18 | or/1-2,10-17 | 416995 |
| 19 | ("2022" or "2023").ti,ab,ot. | 153808 |
| 20 | 18 and 19 | 17876 |
| 21 | (strain or strains or variant$ or mutation$).ti,ab,ot,kf,kw. | 1916135 |
| 22 | 18 and 21 | 32095 |
| 23 | (omikron or Omicron or "B.1.1.529" or "B11529" or xbb$).af. | 9846 |
| 24 | or/20,22-23 | 48695 |
| 25 | Epidemiologic studies/ | 9457 |
| 26 | exp case control studies/ | 1470108 |
| 27 | exp cohort studies/ | 2556574 |
| 28 | Case control.tw. | 158730 |
| 29 | Longitudinal.tw. | 335439 |
| 30 | Retrospective.tw. | 782068 |
| 31 | Cross sectional.tw. | 540774 |
| 32 | Cross-sectional studies/ | 487981 |
| 33 | Observational Studies as Topic/ | 9294 |
| 34 | (observational adj3 (study or studies or design or analysis or analyses)).ti,ab,kf. | 234202 |
| 35 | ((follow up or followup) adj7 (study or studies or design or analysis or analyses)).ti,ab,kf. | 175238 |

36 (epidemiologic$ adj3 (study or studies or design or analysis or analyses)).ti,ab,kf. 119921
37 ((longterm or (long adj term)) adj7 (study or studies or design or analysis or analyses or data)).ti,ab,kf. 160631
38 (cohort$ adj3 (study or studies or design or analysis or analyses or data)).ti,ab,kf. 420054
39 (prospective adj7 (study or studies or design or analysis or analyses)).ti,ab,kf. 551875
40 or/25-39 4243864
41 animals/ not (animals/ and humans/) 5148855
42 40 not 41 4139882
43 42 not (case reports or clinical conference or comment or editorial or letter).pt. 3929840
44 limit 43 to yr="2022 -Current" 538850
45 mortality/ or "cause of death"/ or fatal outcome/ or hospital mortality/ or survival rate/ or Death/ 408166
46 (mortalit$ or death$ or fatal$ or survival).ti,ab,ot,kf,kw. 2881687
47 exp Critical Care/ or exp Intensive Care Units/ 159166
48 (close attention unit$ or intensive care or respiratory care unit or respiratory care units or special care unit or special care units or high dependency unit or high dependency units).ti,ab,ot,kf,kw. 202827
49 (ICU or ICUs or CCU or CCus or GICU or GICUs or HDU or HDUs or ITU or ITUs).ti,ab,ot,kf,kw. 95415
50 (Hospitaliz$ or Hospitalis$ or (hospital adj2 Admission$) or Critical Care).ti,ab,ot,kf,kw. 436800
51 Hospitalization/ or "Length of Stay"/ 228750
52 ("length of stay" or "duration of stay" or "extended stay" or "prolonged stay" or "hospital stay").ti,ab,ot,hw. 220112
53 ("in-patient stay" or "inpatient stay" or "in-patient stays" or "inpatient stays").ti,ab,ot,hw. 3851
54 exp risk/ or odds ratio/ or proportional hazards models/ or life tables/ 1482224
55 (risk$ or danger$ or association$ or peril or jeopard$ or threat$ or chance or chances or hazard$ or gamble$ or probabilit$ or "at stake" or endanger$ or associat$ or likelihood$ or possibilit$ or correlation$ or odds or "vaccine effectiveness" or "immune evasion" or "immunity evasion").ti,ab,ot,kf,kw. 9321739
56 (morbidit$ or comorbid$ or co-morbid$).ti,ab,ot,kf,kw. 716968
57 (incidence or prevalence or predict$ or prognosis).ti,ab,ot,kf,kw. 3993128
58 (HR or RR or aiRR or aOR or "adjusted OR").ti,ab,ot,kf,kw. 471257
59 "severity of illness index"/ or sickness impact profile/ 277780
60 patient acuity/ or early warning score/ 3345
61 Morbidity/ 34586
62 Critical Illness/ 39796
63 Emergencies/ 43459
64 ((severe or severity) adj3 (disease$ or infection$ or illness$ or complication$ or condition$ or illhealth$ or ill-health$ or sickness or virus or infirmity or affliction$)).ti,ab,ot,kf,kw. 285081
65 or/45-64 13120377
66 exp mRNA Vaccines/ 5326
67 ((pfizer$ or biontech) adj5 vaccin$).ti,ab,ot,kf,kw,hw,nm,rn. 3046
68 (tozinameran or Comirnaty or "Pfizer-BioNTech" or "pf07302048" or "pf-07302048" or " BNT-162b2" or "BNT162b2" or Pidacmeran or "BNT162C2" or "BNT-162C2" or Abdavomeran or "BNT-162B1" or "BNT162B1" or "BNT162A1" or "BNT-162A1").ti,ab,ot,kf,kw,hw,nm,rn. 6421
69 "Pfizer-BioNTech".af. 2384
70 "2417899-77-3".af. 0
71 (Elasomeran or moderna or Spikevax or davesomeran or imelasomeran or andusomeran or "mRNA 1273" or "mRNA1273" or "mRNA-1273.211" or "mRNA 1273.211" or Spikevax or "M-1273" or "M1273" or "CX-024414" or "CX024414" or "TAK-919" or "TAK919").ti,ab,ot,kf,kw,hw,nm,rn. 4118
72 ("2430046-03-8" or "2457298-05-2").af. 0
73 (("messenger RNA" or mrna or "RNA based") adj4 (vaccin$ or jab or jabs or shot or shots or immunis$ or immuniz$)).ti,ab,ot,kf,kw. 10148
74 or/66-73 15059
**75 24 and 44 and 74 and 65 825**

***The Medline ALL strategy was updated on 1.2.24 (51 records) and 1.3.24 (63 records).***

*COVID facet based on terms from:*

World Health Organization (26 May 2021) WHO COVID-19 Database Search Strategy. Systematic search of the COVID-19 literature performed Monday through Friday for the WHO Database. Search strategy as of 26 May 2021. Searches performed by Tomas Allen, Kavita Kothari, and Martha Knuth. Available from: https://www.who.int/docs/default-source/coronaviruse/who-covid-19-database/who-covid-19_sources_searchstrategy_20210526.pdf?sfvrsn=65209cc2_5

Canadian Agency for Drugs and Technologies in Health (2.9.21) CADTH COVID-19 Search Strings: COVID-19 — EMBASE (Internet). Available from: https://covid.cadth.ca/literature-searching-tools/cadth-covid-19-search-strings/

NICE (18 December 2020) [accessed 17.8.21] COVID-19 rapid guideline: managing the long-term effects of COVID-19 [NG188]. Search history record [PDF]. NICE: London. Available from: https://www.nice.org.uk/guidance/ng188/evidence/search-strategies-pdf-8957634445

*Observational study design filter adapted from:*

Observational study design filter adapted from:
Scottish Intercollegiate Guidelines Network (SIGN). Search filters: observational studies. Medline. Edinburgh: SIGN, Last Modified 24/04/17 Available from: https://www.sign.ac.uk/what-we-do/methodology/search-filters/

Observational Studies - Medline. In: CADTH Search Filters Database. Ottawa: CADTH; 2023 [accessed 7.11.23]: https://searchfilters.cadth.ca/list?q=topic%3A%22Observational%20studies%22&p=1&ps=&sort=title_sort%20asc

**PubMed (NLM): 2022-2024/01/03**
**Searched 3.1.24**
**https://pubmed.ncbi.nlm.nih.gov/**

**32 #13 AND #24 AND #31 AND #25 281**
31 #26 OR #27 OR #28 OR #29 OR #30 19,625
30 (("messenger RNA"[Title/Abstract] OR mrna[Title/Abstract] OR "RNA based"[Title/Abstract]) AND (vaccine[Title/Abstract] OR vaccines[Title/Abstract] OR vaccination[Title/Abstract] OR jab[Title/Abstract] OR jabs[Title/Abstract] OR shot[Title/Abstract] OR shots[Title/Abstract] OR immunise[Title/Abstract] OR immunised[Title/Abstract] OR immunisation[Title/Abstract] OR immunize[Title/Abstract] OR immunized[Title/Abstract] OR immunization[Title/Abstract])) 15,979
29 Elasomeran[Title/Abstract] OR moderna[Title/Abstract] OR Spikevax[Title/Abstract] OR davesomeran[Title/Abstract] OR imelasomeran[Title/Abstract] OR andusomeran[Title/Abstract] OR "mRNA 1273"[Title/Abstract] OR "mRNA1273"[Title/Abstract] OR "mRNA-1273.211"[Title/Abstract] OR "mRNA 1273.211"[Title/Abstract] OR Spikevax[Title/Abstract] OR "M-1273"[Title/Abstract] OR "M1273"[Title/Abstract] OR "CX-024414"[Title/Abstract] OR "CX024414"[Title/Abstract] OR "TAK-919"[Title/Abstract] OR "TAK919"[Title/Abstract] OR "2430046-03-8"[Title/Abstract] OR "2457298-05-2"[Title/Abstract]3,072
28 tozinameran[Title/Abstract] OR Comirnaty[Title/Abstract] OR "Pfizer-BioNTech"[Title/Abstract] OR "pf07302048"[Title/Abstract] OR "pf-07302048"[Title/Abstract] OR " BNT-162b2"[Title/Abstract] OR "BNT162b2"[Title/Abstract] OR Pidacmeran[Title/Abstract] OR "BNT162C2"[Title/Abstract] OR "BNT-162C2"[Title/Abstract] OR Abdavomeran[Title/Abstract] OR "BNT-162B1"[Title/Abstract] OR "BNT162B1"[Title/Abstract] OR "BNT162A1"[Title/Abstract] OR "BNT-162A1"[Title/Abstract] OR "Pfizer-BioNTech"[Title/Abstract] OR "2417899-77-3"[Title/Abstract] 6,392
27 ((pfizer[Title/Abstract] OR biontech[Title/Abstract]) AND (vaccine[Title/Abstract] OR vaccines[Title/Abstract])) 3,668
26 "mRNA Vaccines"[Mesh] 5,307
25 pubstatusaheadofprint OR publisher[sb] OR pubmednotmedline[sb] 5,627,356
24 #22 AND #23 493,476
23 ("2022/01/01"[Date - Publication] : "3000"[Date - Publication]) 3,253,097
22 #20 NOT #21 3,739,291
21 LETTER[Publication Type] OR EDITORIAL[Publication Type] OR COMMENT[Publication Type] 2,213,615

20 #16 NOT #19 3,825,199
19 #18 NOT (#18 AND #17) 3,757,858
18 rat[tiab] or rats[tiab] or mouse[tiab] or mice[tiab] or murine[tiab] or rodent[tiab] or rodents[tiab] or hamster[tiab] or hamsters[tiab] or pig[tiab] or pigs[tiab] or porcine[tiab] or rabbit[tiab] or rabbits[tiab] or animal[tiab] or animals[tiab] or dogs[tiab] or dog[tiab] or cats[tiab] or cow[tiab] or bovine[tiab] or sheep[tiab] or ovine[tiab] or monkey[tiab] or monkeys[tiab] 4,715,684
17 Human*[tiab] 3,295,660
16 #14 OR #15 3,914,514
15 ((("Epidemiologic Studies"[Mesh:NoExp]) OR "Case-Control Studies"[Mesh]) OR "Cohort Studies"[Mesh]) OR "Cross-Sectional Studies"[Mesh:NoExp] 3,195,884
14 "Case control"[Title/Abstract] OR "cohort study"[Title/Abstract] OR "cohort studies"[Title/Abstract] OR "cohort analysis"[Title/Abstract] OR "cohort analyses"[Title/Abstract] OR "follow up study"[Title/Abstract] OR "follow up studies"[Title/Abstract] OR "observational study"[Title/Abstract] OR "observational studies"[Title/Abstract] OR Longitudinal[Title/Abstract] OR Retrospective[Title/Abstract] OR "Cross sectional"[Title/Abstract] 2,068,573
13 #9 OR #11 OR #12 47,832
12 omikron[Title/Abstract] OR Omicron[Title/Abstract] OR "B.1.1.529"[Title/Abstract] OR "B11529"[Title/Abstract] OR xbb[Title/Abstract] 9,468
11 #7 AND #10 31,553
10 strain[Title/Abstract] OR strains[Title/Abstract] OR variant[Title/Abstract] OR variants[Title/Abstract] OR mutation[Title/Abstract] OR mutations[Title/Abstract] 1,893,774
9 #7 AND #8 17,832
8 "2022"[Title/Abstract] OR "2023"[Title/Abstract] 158,464
7 #1 OR #4 OR #5 OR #6 412,676
6 "COVID-19 2019"[Title/Abstract] OR ncov[Title/Abstract] OR "sars cov"[Title/Abstract] OR sarscov[Title/Abstract] OR "sars coronavirus"[Title/Abstract] OR coronovirus[Title/Abstract] OR "corono virus"[Title/Abstract] OR "19-nCoV"[Title/Abstract] OR "19nCoV"[Title/Abstract] OR "SARS2"[Title/Abstract] OR "SARS-2"[Title/Abstract] OR SARScoronavirus[Title/Abstract] OR "SARS-coronavirus"[Title/Abstract] OR SARScoronovirus[Title/Abstract] OR "SARS-coronovirus"[Title/Abstract] OR "HCoV-19"[Title/Abstract] OR "HCoV19"[Title/Abstract] OR "HCoV-2019"[Title/Abstract] OR "HCoV2019"[Title/Abstract] OR "2019 novel"[Title/Abstract] OR "Severe Acute Respiratory Syndrome Coronavirus 2"[Title/Abstract] OR "Severe Acute Respiratory Syndrome Corona Virus 2"[Title/Abstract] 141,123
5 "Coronavirus"[Title/Abstract] OR "covid 19"[Title/Abstract] OR "2019-ncov"[Title/Abstract] OR "2019-ncov"[Title/Abstract] OR "2019ncov"[Title/Abstract] OR "corona-virus"[Title/Abstract] OR "cov19"[Title/Abstract] OR "cov-19"[Title/Abstract] OR "19nCoV"[Title/Abstract] OR "COVID19"[Title/Abstract] OR "COVID2019"[Title/Abstract] 382,642
4 #2 AND #3 83,020
3 (("Epidemics"[Mesh:NoExp]) OR "Pandemics"[Mesh:NoExp]) OR "Disease Outbreaks"[Mesh:NoExp] 226,548
2 ((("Coronaviridae"[Mesh:NoExp]) OR "Coronavirus"[Mesh]) OR "Betacoronavirus"[Mesh:NoExp]) OR "Coronavirus Infections"[Mesh:NoExp] 185,366
1 ("COVID-19"[Mesh]) OR "Severe acute respiratory syndrome-related coronavirus"[Mesh] 258,558

***The PubMed strategy was updated on 1.2.24 (20 records) and 1.3.24 (17 records).***

*COVID facet based on terms from:*

World Health Organization (26 May 2021) WHO COVID-19 Database Search Strategy. Systematic search of the COVID-19 literature performed Monday through Friday for the WHO Database. Search strategy as of 26 May 2021. Searches performed by Tomas Allen, Kavita Kothari, and Martha Knuth. Available from: https://www.who.int/docs/default-source/coronaviruse/who-covid-19-database/who-covid-19_sources_searchstrategy_20210526.pdf?sfvrsn=65209cc2_5

Canadian Agency for Drugs and Technologies in Health (2.9.21) CADTH COVID-19 Search Strings: COVID-19 — EMBASE (Internet). Available from: https://covid.cadth.ca/literature-searching-tools/cadth-covid-19-search-strings/

NICE (18 December 2020) [accessed 17.8.21] COVID-19 rapid guideline: managing the long-term effects of COVID-19 [NG188]. Search history record [PDF]. NICE: London. Available from: https://www.nice.org.uk/guidance/ng188/evidence/search-strategies-pdf-8957634445

*Observational study design filter adapted from:*
Scottish Intercollegiate Guidelines Network (SIGN). Search filters: observational studies. Medline. Edinburgh: SIGN, Last Modified 24/04/17 Available from: https://www.sign.ac.uk/what-we-do/methodology/search-filters/

*PubMed limit:*
Duffy S, de Kock S, Misso K, Noake C, Ross J, Stirk L. Supplementary searches of PubMed to improve currency of MEDLINE and MEDLINE In-Process searches via Ovid. J Med Libr Assoc. 2016 Oct;104(4):309-312. doi: 10.3163/1536-5050.104.4.011. https://www.ncbi.nlm.nih.gov/pmc/articles/PMC5079494/

**Europe PMC, including medRxiv and bioRxiv preprints (Internet): up to 2024/01/04**
**Searched 4.1.24**
**https://europepmc.org/**

| **Search terms** | **Results** |
|---|---|
| (TITLE:"COVID-19" OR TITLE:"coronavirus" OR TITLE:"COVID" OR TITLE:"NCOV" OR TITLE:"SARS-CoV-2") AND (TITLE:"2022" OR TITLE:"2023" OR TITLE:"strain" OR TITLE:"strains" OR TITLE:"variant" OR TITLE:"variants" OR TITLE:"mutation" OR TITLE:"mutations" OR TITLE:"omicron" OR TITLE:"omicron") AND (TITLE:"pfizer" OR TITLE:"biontech" OR TITLE:" tozinameran" OR TITLE:" Comirnaty" OR TITLE:" Pfizer-BioNTech" OR TITLE:" BNT-162b2" OR TITLE:" BNT162b2" OR TITLE:"BNT162C2" OR TITLE:"BNT-162C2" OR TITLE:"Abdavomeran" OR TITLE:"Elasomeran" OR TITLE:"moderna" OR TITLE:"spikevax" OR TITLE:"TAK-919" OR TITLE:"TAK919" OR TITLE:"RNA vaccine" OR TITLE:"RNA vaccines" OR TITLE:"mrna vaccine" OR TITLE:"mrna vaccines") AND (TITLE:"cohort" OR TITLE:"observational" OR TITLE:"longitudinal" OR TITLE:"followup" OR TITLE:"follow up" OR TITLE:"epidemiologic" OR TITLE:"epidemiological" OR TITLE:"longterm" OR TITLE:"long term" OR TITLE:"prospective") | **10** |

***The Europe PMC strategy was updated on 1.2.24 (1 record) and 1.3.24 (0 records).***

**Latin American and Caribbean Health Sciences Literature (LILACS) (Internet): 2022-2024/01/04**
**Searched 4.1.24**
https://search.bvsalud.org/portal/?lang=en

Searched Title/Abstract/Subject
Limited to Observational studies
Limited to 2022-2024/01/04
Limited to LILACS only

((COVID OR NCOV OR coronavirus OR COVID19 OR "SARs-COV-2" OR Omicron OR omicron)) AND ((mortality OR death OR deaths OR fatal OR fatality OR fatalities OR survival OR "close attention unit" OR "close attention units" OR "intensive care" OR "respiratory care unit" OR "respiratory care units" OR "special care unit" OR "special care units" OR "high dependency unit" OR "high dependency units" OR ICU OR ICUs OR CCU OR CCus OR GICU OR GICUs OR HDU OR HDUs OR ITU OR ITUs OR Hospitalized OR Hospitalised population OR Hospitalization OR Hospitalisation OR Hospitalizations OR Hospitalisations OR "hospital Admission" OR "hospital Admissions" OR "Critical Care" OR "length of stay" OR "duration of stay" OR "extended stay" OR "prolonged stay" OR "hospital stay" OR "in-patient stay" OR "inpatient stay" OR "in-patient stays" OR "inpatient stays" OR risk OR risks OR risky OR danger OR dangers OR dangerous OR association OR associations OR peril OR jeopardy OR jeopardise OR jeopardize OR threat OR threats OR chance OR chances OR hazard OR hazards OR hazardous OR gamble OR gambles OR gambling OR probability OR probabilities OR "at stake" OR endanger OR endangers OR endangered OR endangering OR associate OR associates OR associating OR association OR likelihood OR likelihoods OR possibility OR possibilities OR correlation OR correlations OR odds OR morbidity OR comorbidity OR "co-morbidity" OR morbidities OR comorbidities OR "co-morbidities" OR incidence OR prevalence OR predict OR predicted OR predictive OR prognosis OR prediction OR predictions OR HR OR RR OR aiRR OR aOR OR "adjusted OR" OR "disease severity" OR "severity score" OR "severity index" OR "severity of illness" OR "critical illness" OR "emergency status" OR "urgency status" OR "severe complication" OR "severe complications") ) AND ((pfizer OR biontech OR tozinameran OR Comirnaty OR "Pfizer-BioNTech" OR "pf07302048" OR "pf-07302048" OR " BNT-162b2" OR "BNT162b2" OR Pidacmeran OR "BNT162C2" OR "BNT-162C2" OR Abdavomeran OR "BNT-162B1" OR "BNT162B1" OR "BNT162A1" OR "BNT-162A1" OR "Pfizer-BioNTech" OR "2417899-77-3" OR Elasomeran OR moderna OR Spikevax OR davesomeran OR imelasomeran OR andusomeran OR "mRNA 1273" OR "mRNA1273" OR "mRNA-1273.211" OR "mRNA 1273.211" OR Spikevax OR "M-1273" OR "M1273" OR "CX-024414" OR "CX024414" OR "TAK-919" OR "TAK919" OR "2430046-03-8" OR "2457298-05-2" OR "RNA vaccine" OR "RNA vaccines" OR "RNA vaccination" OR "RNA shot" OR "RNA shots" OR "mRNA vaccine" OR "mRNA vaccines" OR "mRNA vaccination" OR "mRNA shot" OR "mRNA shots") ) AND ((2022 OR 2023 OR 2024 OR strain OR strains OR variant OR variants OR mutations OR mutation OR omikron OR Omicron OR "B.1.1.529" OR "B11529" OR xbb) )

**N=3**

***The LILACS strategy was updated on 1.2.24 (0 records) and 1.3.24 (0 records).***

**Cochrane COVID-19 Study Register (www): 2022-2024/01/04**
**Searched 4.1.24**
https://covid-19.cochrane.org/

Limited to Observational cohort studies.
Limited 2022-2024/01/04

(mortality OR death OR deaths OR fatal OR fatality OR fatalities OR survival OR "close attention unit" OR "close attention units" OR "intensive care" OR "respiratory care unit" OR "respiratory care units" OR "special care unit" OR "special care units" OR "high dependency unit" OR "high dependency units" OR ICU OR ICUs OR CCU OR CCus OR GICU OR GICUs OR HDU OR HDUs OR ITU OR ITUs OR Hospitalized OR Hospitalised population OR Hospitalization OR Hospitalisation OR Hospitalizations OR Hospitalisations OR "hospital Admission" OR "hospital Admissions" OR "Critical Care" OR "length of stay" OR "duration of stay" OR "extended stay" OR "prolonged stay" OR "hospital stay" OR "in-patient stay" OR "inpatient stay" OR "in-patient stays" OR "inpatient stays" OR risk OR risks OR risky OR danger OR dangers OR dangerous OR association OR associations OR peril OR jeopardy OR jeopardise OR jeopardize OR threat OR threats OR chance OR chances OR hazard OR hazards OR hazardous OR gamble OR gambles OR gambling OR probability OR probabilities OR "at stake" OR endanger OR endangers OR endangered OR endangering OR associate OR associates OR associating OR association OR likelihood OR likelihoods OR possibility OR possibilities OR correlation OR correlations OR odds OR morbidity OR comorbidity OR "co-morbidity" OR morbidities OR comorbidities OR "co-morbidities" OR incidence OR prevalence OR predict OR predicted OR predictive OR prognosis OR prediction OR predictions OR HR OR RR OR aiRR OR aOR OR "adjusted OR" OR "disease severity" OR "severity score" OR "severity index" OR "severity of illness" OR "critical illness" OR "emergency status" OR "urgency status" OR "severe complication" OR "severe complications") AND (pfizer OR biontech OR tozinameran OR Comirnaty OR "Pfizer-BioNTech" OR "pf07302048" OR "pf-07302048" OR " BNT-162b2" OR "BNT162b2" OR Pidacmeran OR "BNT162C2" OR "BNT-162C2" OR Abdavomeran OR "BNT-162B1" OR "BNT162B1" OR "BNT162A1" OR "BNT-162A1" OR "Pfizer-BioNTech" OR "2417899-77-3" OR Elasomeran OR moderna OR Spikevax OR davesomeran OR imelasomeran OR andusomeran OR "mRNA 1273" OR "mRNA1273" OR "mRNA-1273.211" OR "mRNA 1273.211" OR Spikevax OR "M-1273" OR "M1273" OR "CX-024414" OR "CX024414" OR "TAK-919" OR "TAK919" OR "2430046-03-8" OR "2457298-05-2" OR "RNA vaccine" OR "RNA vaccines" OR "RNA vaccination" OR "RNA shot" OR "RNA shots" OR "mRNA vaccine" OR "mRNA vaccines" OR "mRNA vaccination" OR "mRNA shot" OR "mRNA shots") AND (2022 OR 2023 OR 2024 OR strain OR strains OR variant OR variants OR mutations OR mutation OR omikron OR Omicron OR "B.1.1.529" OR "B11529" OR xbb) AND (cohort OR cohorts OR observational OR "follow up" OR followup OR epidemiologic OR epidemiological OR longterm OR "long term" OR longitudinal OR prospective)

**N=(1551 studies) 1792 references**

***The Cochrane COVID-19 Study Register strategy was updated on 1.2.24 (1 record) and 1.3.24 (0 records).***

**WHO COVID-19 (Internet): 2022-2024/01/03**
**Searched 3.1.24**
**https://search.bvsalud.org/global-literature-on-novel-coronavirus-2019-ncov/?lang=en**

Limited to Observational and cohort studies.
Limited 2022-2023.

(ti:(mortality OR death OR deaths OR fatal OR fatality OR fatalities OR survival OR "close attention unit" OR "close attention units" OR "intensive care" OR "respiratory care unit" OR "respiratory care units" OR "special care unit" OR "special care units" OR "high dependency unit" OR "high dependency units" OR ICU OR ICUs OR CCU OR CCus OR GICU OR GICUs OR HDU OR HDUs OR ITU OR ITUs OR Hospitalized OR Hospitalised population OR Hospitalization OR Hospitalisation OR Hospitalizations OR Hospitalisations OR "hospital Admission" OR "hospital Admissions" OR "Critical Care" OR "length of stay" OR "duration of stay" OR "extended stay" OR "prolonged stay" OR "hospital stay" OR "in-patient stay" OR "inpatient stay" OR "in-patient stays" OR "inpatient stays" OR risk OR risks OR risky OR danger OR dangers OR dangerous OR association OR associations OR peril OR jeopardy OR jeopardise OR jeopardize OR threat OR threats OR chance OR chances OR hazard OR hazards OR hazardous OR gamble OR gambles OR gambling OR probability OR probabilities OR "at stake" OR endanger OR endangers OR endangered OR endangering OR associate OR associates OR associating OR association OR likelihood OR likelihoods OR possibility OR possibilities OR correlation OR correlations OR odds OR morbidity OR comorbidity OR "co-morbidity" OR morbidities OR comorbidities OR "co-morbidities" OR incidence OR prevalence OR predict OR predicted OR predictive OR prognosis OR prediction OR predictions OR HR OR RR OR aiRR OR aOR OR "adjusted OR" OR "disease severity" OR "severity score" OR "severity index" OR "severity of illness" OR "critical illness" OR "emergency status" OR "urgency status" OR "severe complication" OR "severe complications")) AND (pfizer$ OR biontech OR tozinameran OR Comirnaty OR "Pfizer-BioNTech" OR "pf07302048" OR "pf-07302048" OR " BNT-162b2" OR "BNT162b2" OR Pidacmeran OR "BNT162C2" OR "BNT-162C2" OR Abdavomeran OR "BNT-162B1" OR "BNT162B1" OR "BNT162A1" OR "BNT-162A1" OR "Pfizer-BioNTech" OR "2417899-77-3" OR Elasomeran OR moderna OR Spikevax OR davesomeran OR imelasomeran OR andusomeran OR "mRNA 1273" OR "mRNA1273" OR "mRNA-1273.211" OR "mRNA 1273.211" OR Spikevax OR "M-1273" OR "M1273" OR "CX-024414" OR "CX024414" OR "TAK-919" OR "TAK919" OR "2430046-03-8" OR "2457298-05-2" OR "RNA vaccine" OR "RNA vaccines" OR "RNA vaccination" OR "RNA shot" OR "RNA shots" OR "mRNA vaccine" OR "mRNA vaccines" OR "mRNA vaccination" OR "mRNA shot" OR "mRNA shots")

**N=477**

***** WHO COVID-19 database ceased on June 2023, therefore update searches were not necessary.

##### 1.2. Study Selection

Identified studies were first assessed at title and abstract level using the Rayyan tool[1] by 2 independent reviewers to determine whether they met the inclusion criteria. Disagreements were resolved by discussion until a consensus was reached. Full text screening was performed on all studies that met the inclusion criteria at title and abstract level.

##### 1.3. Data Extraction

One reviewer extracted data on study and patient characteristics, and outcomes of interest into a predefined table. A second reviewer verified the accuracy of extraction. For studies that also coincided with other SARS-CoV-2 variants, only data relating to the Omicron variant were extracted, as defined by the study.

##### 1.4. Quality Assessment

Studies were assessed as having high, moderate, or low risk of bias using the Newcastle-Ottawa scale for cohort and case-control studies[2] and the Joanna Briggs Institute checklist for cross-sectional studies.[3] A single reviewer conducted the assessments, and a second reviewer verified the results. Discrepancies were resolved by discussion.

##### 1.5. Data Analysis

###### *1.5.1 Statistical Analysis*

Odds ratios (ORs), hazard ratios (HRs), and rate ratios are considered equal estimates and were therefore combined.[4] Weights were calculated using the inverse variance method (weight= 1/variance). Random--effects DerSimonian and Laird models[5] were fitted to calculate pooled relative risk (RR) and 95% confidence interval for all outcomes. Population analyses that included fewer than 5 subgroups were not conducted due to the challenges associated with combining few studies. ‘Subgroup’ refers to the types of immunocompromising/immunosuppressive conditions of interest. It often contains any additional information that differentiates the condition from another one included in the same meta-analysis by the same study (e.g. ‘men who take immunosuppressants’ and ‘women who take immunosuppressants’). ‘Subgroup’ is often used interchangeably with ‘study’. However, considering that some studies included multiple subgroups that are included in the same meta-analysis, for clarity they are referred to in this report as ‘subgroups’.

Heterogeneity was measured using Cochran’s Q statistic, with statistical significance set as $P < .05$, and quantified by the $I^2$ test. The $I^2$ statistic as defined by the Cochrane Handbook for Systematic Reviews of Interventions[6] was used for thresholds for interpretation: 0% to 40% might not be important; 40% to 60% may represent moderate heterogeneity; 60% to 90% may represent substantial heterogeneity; 90% to 100% may represent considerable heterogeneity. Publication bias was assessed using funnel plots and the Egger’s test.[7]

###### *1.5.2 Additional Subgroup Analysis*

Studies that exclusively included children were excluded from all analyses. For analyses for which those studies were eligible, an additional sensitivity analysis called ‘Including children’ was conducted. Additional subgroup analyses included the ‘> 50 years’ population included only participants older than 50 years, the ‘COVID-19-related outcomes only’ population included only outcomes that were explicitly caused by COVID-19. Patients with renal disease were further analyzed for the ‘Death’ outcome in 2 population subgroups: ‘chronic kidney disease (CKD) stage 3’ (included CKD stage 3 subgroups) and ‘advanced renal disease’ (included CKD stage 5, renal failure, end-stage kidney disease, and renal replacement therapy subgroups).

#### 2. Supplemental eResults

##### 2.1. Studies With Insufficient Numbers for Meta-Analysis

Due to insufficient number of subgroups, meta-analyses were not conducted for the following outcomes and immunocompromising/immunosuppressing (IC/IS) conditions: intensive care unit (ICU) admission for people with autoimmune diseases (n = 2) and transplant (n = 2); death (n = 1), hospitalization (n = 3), ICU (n = 0), or combined outcomes (n = 2) for people with HIV; and hospitalization for people with liver disease (n = 2) (**Table 3**). Studies and subgroups identified for each of these conditions are narratively described in **Section 2.6** of the Supplement.

##### 2.2. Risk of Death – Other Subgroup Analyses

Subgroup analyses differed from the main analysis and showed that people with either autoimmune or liver diseases did not have a significantly higher risk of death when they were hospitalized before the start of the study (**Supplemental eTable 5**).

##### 2.3. Risk of ICU – Sensitivity and Subgroup Analyses

Statistical significance was lost in the group of people with cancer in the least adjusted analysis ($P$ = .0647) (**Supplemental eTable 8**). The ‘Leave-1-out’ sensitivity analyses assessed the effect of removing individual studies on pooled estimates. Removing the Starkey 2023 study[8] from the cancer group analysis and the Russell 2023[9] or Mayer 2023[10] studies from the renal disease group resulted in a loss of significance for each analysis; these studies were not outliers but had more weight than other studies.

Subgroup analyses showed that people with cancer who were hospitalized before the study did not have a significant risk of ICU admission (**Supplemental eTable 9**). Additionally, people with renal disease did not have a significant risk of ICU admission in any of the analyzed subgroups, which is likely due to the low number of studies included for each subgroup.

##### 2.4. Risk of Any Combination of Outcomes (Death, Hospitalization, or ICU Admission)

For all assessed IC/IS conditions (autoimmune diseases, cancer, IC/IS, liver disease, renal diseases, or transplant), patients with the condition had a significantly increased risk of the ‘Combined’ outcome ($P < .05$; **Figure 2D**) in comparison with patients without the condition. Statistical heterogeneity was substantial for studies of patients with autoimmune diseases, and considerable for all other groups (**Figure 2D**). Publication bias was detected in studies of patients with cancer or liver disease, and could not be assessed for studies of patients with autoimmune diseases or transplant recipients (**Supplemental eTable 10**).

All sensitivity analyses results had the same degree of significance as the main analysis for the risk of hospitalization outcome (**Supplemental eTable 10**), indicating the robustness of the main analysis results. Subgroup analyses demonstrated that patients with autoimmune diseases or cancer did not have a significant risk of combined outcomes if they were hospitalized prior to the study (**Supplemental eTable 11**). Additionally, patients with liver disease did not have a higher risk of combined outcomes for COVID-19-related only outcomes or if they were not hospitalized before the study.

#### 2.5. Supplemental eTables

##### *2.5.1. Supplemental eTable 1. Inclusion and Exclusion Criteria for Studies*

##### *Included in This Analysis*

| Characteristic | Inclusion criteria | Exclusion criteria |
|---|---|---|
| Population | ▪ Immunocompromised/immunosuppressed:<br>▫ Immunocompromised/immunosuppressed groups<br>▫ Solid organ transplant (current or historic)<br>▫ Stem cells transplant (current or historic)<br>▫ Bone marrow transplant (current or historic)<br>▫ Any malignancy (current or historic)<br>▫ Autoimmune conditions<br>▫ Any liver diseases<br>▫ Chronic kidney disease<br>▫ Advanced or untreated HIV<br>▫ End-stage kidney disease | ▪ All patients with conditions of interest to prevent synergizing effects<br>▪ High risk patient groups, i.e. patients who are eligible to take antivirals to prevent severe COVID-19<br>▪ Highly vulnerable populations, such as the elderly in nursing homes |
| Comparators | ▪ Non-immunocompromised/non-immunosuppressed people<br>▪ General population | ▪ Non-comparative studies |
| Outcomes | ▪ Risk of:<br>▫ Hospitalization<br>▫ ICU admission<br>▫ Death<br>▫ Combined: other potential COVID-19 severity outcomes, where severity is measured by any of the outcomes outlined above<br>Outcomes that were not explicit 'Hospitalization' or 'ICU', such as mechanical ventilation, were also included in this review as part of 'ICU' or 'Hospitalization' outcomes, depending on the definitions | ▪ Studies that do not report at least 1 relevant outcome of interest |
| Outcome measures | ▪ Risk ratio<br>▪ Rate ratio<br>▪ Hazard ratio<br>▪ Odds ratio<br>▪ Incidence rate ratios | - |

| | | |
|---|---|---|
| Study design | ▪ Observational (cohort, case-control, cross-sectional) | ▪ Interventional studies, such as randomized/non-randomized controlled trials |
| Language | ▪ Studies with full text published in English will be included. All potentially relevant publications without English language full text will be listed | ▪ Studies without a full text published in English |
| Timeframe | ▪ Majority of cases contracted the Omicron variant, as defined by study authors. When the study period was not described as predominantly Omicron, the cut-off date of December 2021, when the WHO declared Omicron as a new variant of concern,[11] was chosen | ▪ Studies not reporting data on the Omicron variant |
| Publication date | ▪ Studies published after 1 Jan 2022 | ▪ Studies published prior to 1 Jan 2022 |
| Publication type | ▪ Full-text articles | ▪ Conference abstracts<br>▪ Letters<br>▪ Case reports<br>▪ Editorials |
| Countries | ▪ Any | - |

ICU, intensive care unit; WHO, World Health Organization.

##### *2.5.2 Supplemental eTable 2. Omicron Period in the Included Studies*

| Year | 2021 | | 2022 | | | | | | | | | | | | 2023 | | | | | | | | | | | |
|---|---|---|---|---|---|---|---|---|---|---|---|---|---|---|---|---|---|---|---|---|---|---|---|---|---|---|
| **Study/month** | **11** | **12** | **1** | **2** | **3** | **4** | **5** | **6** | **7** | **8** | **9** | **10** | **11** | **12** | **1** | **2** | **3** | **4** | **5** | **6** | **7** | **8** | **9** | **10** | **11** | **12** |
| Agrawal 2022 | | | | | | | | | | | | | | | | | | | | | | | | | | |
| AlBahrani 2022 | | | | | | | | | | | | | | | | | | | | | | | | | | |
| Arbel 2023 | | | | | | | | | | | | | | | | | | | | | | | | | | |
| Arbel 2022 | | | | | | | | | | | | | | | | | | | | | | | | | | |
| Bahremand 2023 | | | | | | | | | | | | | | | | | | | | | | | | | | |
| Bao 2022 | | | | | | | | | | | | | | | | | | | | | | | | | | |
| Bedston 2024 | | | | | | | | | | | | | | | | | | | | | | | | | | |
| Benites-Godínez 2023 | | | | | | | | | | | | | | | | | | | | | | | | | | |
| Beppu 2024 | | | | | | | | | | | | | | | | | | | | | | | | | | |
| Beraud 2023 | | | | | | | | | | | | | | | | | | | | | | | | | | |
| Bournia 2023 | | | | | | | | | | | | | | | | | | | | | | | | | | |
| Briciu 2023 | | | | | | | | | | | | | | | | | | | | | | | | | | |
| Brosh-Nissimov 2023 | | | | | | | | | | | | | | | | | | | | | | | | | | |
| Chen 2023 | | | | | | | | | | | | | | | | | | | | | | | | | | |
| Chen 2024 | | | | | | | | | | | | | | | | | | | | | | | | | | |
| Choi 2023 | | | | | | | | | | | | | | | | | | | | | | | | | | |
| de Prost 2022 | | | | | | | | | | | | | | | | | | | | | | | | | | |
| de Prost 2023 | | | | | | | | | | | | | | | | | | | | | | | | | | |
| Drummond 2023 | | | | | | | | | | | | | | | | | | | | | | | | | | |
| Elamin 2024 | | | | | | | | | | | | | | | | | | | | | | | | | | |
| Ellis 2023 | | | | | | | | | | | | | | | | | | | | | | | | | | |

| Year | 2021 | | 2022 | | | | | | | | | | | | 2023 | | | | | | | | | | | |
|---|---|---|---|---|---|---|---|---|---|---|---|---|---|---|---|---|---|---|---|---|---|---|---|---|---|---|
| **Study/month** | **11** | **12** | **1** | **2** | **3** | **4** | **5** | **6** | **7** | **8** | **9** | **10** | **11** | **12** | **1** | **2** | **3** | **4** | **5** | **6** | **7** | **8** | **9** | **10** | **11** | **12** |
| Evans 2023 | | | | | | | | | | | | | | | | | | | | | | | | | | |
| Favia 2023 | | | | | | | | | | | | | | | | | | | | | | | | | | |
| Flacco 2023 | | | | | | | | | | | | | | | | | | | | | | | | | | |
| Gazit 2022 | | | | | | | | | | | | | | | | | | | | | | | | | | |
| Griggs 2024 | | | | | | | | | | | | | | | | | | | | | | | | | | |
| Guo 2023 | | | | | | | | | | | | | | | | | | | | | | | | | | |
| Helmy 2023[a] | | | | | | | | | | | | | | | | | | | | | | | | | | |
| Hippisley-Cox 2023 | | | | | | | | | | | | | | | | | | | | | | | | | | |
| Jamaati 2023 | | | | | | | | | | | | | | | | | | | | | | | | | | |
| Kang 2023 | | | | | | | | | | | | | | | | | | | | | | | | | | |
| Karageorgou 2023 | | | | | | | | | | | | | | | | | | | | | | | | | | |
| Kim 2023 | | | | | | | | | | | | | | | | | | | | | | | | | | |
| Klein 2023 | | | | | | | | | | | | | | | | | | | | | | | | | | |
| Konermann 2023 | | | | | | | | | | | | | | | | | | | | | | | | | | |
| Lee 2024 | | | | | | | | | | | | | | | | | | | | | | | | | | |
| Li 2024 | | | | | | | | | | | | | | | | | | | | | | | | | | |
| Li 2023 | | | | | | | | | | | | | | | | | | | | | | | | | | |
| Liu 2023 | | | | | | | | | | | | | | | | | | | | | | | | | | |
| Lu 2022 | | | | | | | | | | | | | | | | | | | | | | | | | | |
| Mayer 2023 | | | | | | | | | | | | | | | | | | | | | | | | | | |
| Mendoza-Cano 2023 | | | | | | | | | | | | | | | | | | | | | | | | | | |
| Mizrahi Reuveni 2023 | | | | | | | | | | | | | | | | | | | | | | | | | | |
| Morris 2023 | | | | | | | | | | | | | | | | | | | | | | | | | | |
| Nab 2023 | | | | | | | | | | | | | | | | | | | | | | | | | | |

| **Year** | **2021** | | **2022** | | | | | | | | | | | | **2023** | | | | | | | | | | | |
|---|---|---|---|---|---|---|---|---|---|---|---|---|---|---|---|---|---|---|---|---|---|---|---|---|---|---|
| **Study/month** | **11** | **12** | **1** | **2** | **3** | **4** | **5** | **6** | **7** | **8** | **9** | **10** | **11** | **12** | **1** | **2** | **3** | **4** | **5** | **6** | **7** | **8** | **9** | **10** | **11** | **12** |
| Nevejan 2022 | | | | | | | | | | | | | | | | | | | | | | | | | | |
| O'Leary 2023 | | | | | | | | | | | | | | | | | | | | | | | | | | |
| Overvad 2022 | | | | | | | | | | | | | | | | | | | | | | | | | | |
| Parajuli 2023 | | | | | | | | | | | | | | | | | | | | | | | | | | |
| Parra-Bracamonte 2023 | | | | | | | | | | | | | | | | | | | | | | | | | | |
| Patton 2023 | | | | | | | | | | | | | | | | | | | | | | | | | | |
| Puyat 2023 | | | | | | | | | | | | | | | | | | | | | | | | | | |
| Radhakrishnan 2023 | | | | | | | | | | | | | | | | | | | | | | | | | | |
| Rasmussen 2023 | | | | | | | | | | | | | | | | | | | | | | | | | | |
| Risk 2022 | | | | | | | | | | | | | | | | | | | | | | | | | | |
| Russell 2023 | | | | | | | | | | | | | | | | | | | | | | | | | | |
| Shakor 2023 | | | | | | | | | | | | | | | | | | | | | | | | | | |
| Shi 2023 | | | | | | | | | | | | | | | | | | | | | | | | | | |
| Simmons 2023 | | | | | | | | | | | | | | | | | | | | | | | | | | |
| Skarbinski 2022 | | | | | | | | | | | | | | | | | | | | | | | | | | |
| Starkey 2023 | | | | | | | | | | | | | | | | | | | | | | | | | | |
| Svensson 2023 | | | | | | | | | | | | | | | | | | | | | | | | | | |
| Tsujimoto 2023 | | | | | | | | | | | | | | | | | | | | | | | | | | |
| Vo 2022 | | | | | | | | | | | | | | | | | | | | | | | | | | |
| Wang 2023 | | | | | | | | | | | | | | | | | | | | | | | | | | |
| Ward 2024 | | | | | | | | | | | | | | | | | | | | | | | | | | |
| Xin 2024 | | | | | | | | | | | | | | | | | | | | | | | | | | |
| Xing 2023 | | | | | | | | | | | | | | | | | | | | | | | | | | |
| Yang 2023 | | | | | | | | | | | | | | | | | | | | | | | | | | |

| **Year** | **2021** | | **2022** | | | | | | | | | | | | **2023** | | | | | | | | | | | |
|---|---|---|---|---|---|---|---|---|---|---|---|---|---|---|---|---|---|---|---|---|---|---|---|---|---|---|
| **Study/month** | **11** | **12** | **1** | **2** | **3** | **4** | **5** | **6** | **7** | **8** | **9** | **10** | **11** | **12** | **1** | **2** | **3** | **4** | **5** | **6** | **7** | **8** | **9** | **10** | **11** | **12** |
| Zhang 2022 | | | | | | | | | | | | | | | | | | | | | | | | | | |
| Zhao 2023 | | | | | | | | | | | | | | | | | | | | | | | | | | |
| Zhu 2023 | | | | | | | | | | | | | | | | | | | | | | | | | | |

[a]Helmy 2023 did not explicitly mention the Omicron variant and study period; however, it was included based on the following sentence: "This prospective observational study was conducted in a university hospital after the institutional research ethics board approval (N-24-2022)". Green indicates the Omicron study period of the studies included in the analyses; Orange indicates the Omicron study period of the studies excluded from the analyses.

##### *2.5.3. Supplemental eTable 3. Risk of Bias of the Included Studies Using NOS and JBI Tools*

| Study | Low | Medium | High |
|---|---|---|---|
| Agrawal 2022[12] | ● | | |
| AlBahrani 2022[13] | ● | | |
| Arbel 2022[14] | ● | | |
| Arbel 2023[15] | ● | | |
| Bahremand 2023[16] | ● | | |
| Bao 2022[17a] | ● | | |
| Bedston 2024[18] | ● | | |
| Benites-Godínez 2023[19] | ● | | |
| Beppu 2024[20] | ● | | |
| Beraud 2023[21] | ● | | |
| Bournia 2023[22] | ● | | |
| Briciu 2023[23] | ● | | |
| Brosh-Nissimov 2023[24] | ● | | |
| Chen 2023[25a] | ● | | |
| Chen 2024[26] | ● | | |
| Choi 2023[27] | ● | | |
| de Prost 2022[28] | ● | | |
| de Prost 2023[29] | ● | | |
| Drummond 2023[30] | ● | | |
| Elamin 2024[31] | ● | | |
| Ellis 2023[32] | ● | | |
| Evans 2023[33] | ● | | |
| Favia 2023[34] | ● | | |
| Flacco 2023[35] | ● | | |
| Gazit 2022[36] | | ● | |
| Griggs 2024[37] | | ● | |
| Guo 2023[38] | ● | | |
| Helmy 2023[39] | | ● | |
| Hippisley-Cox 2023[40] | | ● | |
| Jamaati 2023[41] | | ● | |
| Kang 2023[42] | ● | | |
| Karageorgou 2023[43] | ● | | |
| Kim 2023[44] | ● | | |

| Study | Low | Medium | High |
|---|---|---|---|
| Klein 2023[45] | ● | | |
| Konermann 2023[46] | ● | | |
| Lee 2024[47] | ● | | |
| Li 2023[48a] | ● | | |
| Li 2024[49] | | ● | |
| Liu 2023[50] | ● | | |
| Lu 2022[51a] | ● | | |
| Mayer 2023[10] | | ● | |
| Mendoza-Cano 2023[52] | ● | | |
| Mizrahi Reuveni 2023[53] | ● | | |
| Morris 2023[54b] | ● | | |
| Nab 2023[55] | ● | | |
| Nevejan 2022[56] | ● | | |
| O'Leary 2023[57] | ● | | |
| Overvad 2022[58] | | ● | |
| Parajuli 2023[59] | ● | | |
| Parra-Bracamonte 2023[60] | ● | | |
| Patton 2023[61] | | ● | |
| Puyat 2023[62] | ● | | |
| Radhakrishnan 2023[63] | | ● | |
| Rasmussen 2023[64] | ● | | |
| Risk 2022[65] | ● | | |
| Russell 2023[9] | | ● | |
| Shakor 2023[66a,b] | | | ● |
| Shi 2023[67] | ● | | |
| Simmons 2023[68] | ● | | |
| Skarbinski 2022[69] | ● | | |
| Starkey 2023[8] | | ● | |
| Svensson 2023[70] | ● | | |
| Tsujimoto 2023[71] | ● | | |
| Vo 2022[72] | ● | | |
| Wang 2023[73] | ● | | |
| Ward 2024[74] | ● | | |
| Xin 2024[75] | ● | | |
| Xing 2023[76b] | ● | | |
| Yang 2023[77] | ● | | |

| Study | Low | Medium | High |
|---|---|---|---|
| Zhang 2022[78] | ● | | |
| Zhao 2023[79] | ● | | |
| Zhu 2023[80] | ● | | |

Abbreviations: JBI, Joanna Briggs Institute; NOS, Newcastle-Ottawa scale.

[a]Risk of bias was assessed using the JBI critical appraisal checklist for analytical cross-sectional studies. Green indicates low risk of bias, amber – medium, and red – high.

[b]Shakor 2023 was assessed using the JBI critical appraisal checklist for analytical cross-sectional studies. For 2 of the questions the study was unclear (3. Was the exposure measured in a valid and reliable way?; 4. Were objective, standard criteria used for measurement of the condition?). For 1 question the answer was no (5. Were confounding factors identified?). For 1 question, the answer was not applicable (6. Were strategies to deal with confounding factors stated [adjustment during analysis]?).

##### *2.5.4. Supplemental eTable 4. Main and Sensitivity Analyses Results of the 'Death' Outcome*

| | Main analysis (RR (95% CI), *P* value, $I^2$) | Statistical evidence of publication bias (yes/no) | Least adjusted (RR [95% C]), *P* value, $I^2$) | Only adjusted (RR [95% C]), *P* value, $I^2$) | Excluding studies for population overlap (RR [95% CI], *P* value, $I^2$) | Leave-1-out (yes/no for significant difference of removing a subgroup) |
|---|---|---|---|---|---|---|
| **Autoimmune diseases** | 1.33 (1.20, 1.48), $P < .05$, $I^2$ = 65% | No | 1.39 (1.29, 1.49), $P < .001$, $I^2$ = 31.92% | 1.34 (1.20, 1.48), $P < .001$, $I^2$ = 68.15% | 1.33 (1.21, 1.48), $P < .001$, $I^2$ = 64.61% | No |
| **Cancer** | 2.40 (1.22, 2.74), $P < .05$, $I^2$ = 97% | No | 2.49 (1.90, 3.26), $P < .001$, $I^2$ = 98.94% | 2.53 (2.21, 2.90), $P < .001$, $I^2$ = 96.94% | 2.40 (2.11, 2.74), $P < .001$, $I^2$ = 96.81% | No |
| **Immunocompromised/ immunosuppressed** | 2.17 (1.77, 2.66), $P < .05$, $I^2$ = 97% | No | 2.25 (1.83, 2.75), $P < .001$, $I^2$ = 97.02% | 2.20 (1.77, 2.72), $P < .001$, $I^2$ = 97.16% | 2.18 (1.77, 2.67), $P < .001$, $I^2$ = 96.9% | No |
| **Liver disease** | 1.79 (1.31, 2.43), $P < .05$, $I^2$ = 96% | No | 1.99 (1.52, 2.59), $P < .001$, $I^2$ = 94.63% | 1.85 (1.35, 2.53), $P < .001$, $I^2$ = 96.47% | 1.79 (1.31, 2.43), $P < .001$, $I^2$ = 95.83% | No |
| **Renal disease** | 2.15 (1.88, 2.46), $P < .05$, $I^2$ = 98% | Yes | 2.21 (1.93, 2.53), $P < .001$, $I^2$ = 98.28% | 2.24 (1.94, 2.58), $P < .001$, $I^2$ = 98.41% | 2.26 (1.96, 2.60), $P < .001$, $I^2$ = 98.29% | No |
| **Transplant** | 6.78 (4.41, 10.43), $P < .05$, $I^2$ = 80% | Yes | 6.78 (4.41, 10.43), $P < .001$, $I^2$ = 79.86% | 8.50 (5.70, 12.67), $P < .001$, $I^2$ = 74.81% | 6.78 (4.41, 10.43), $P < .001$, $I^2$ = 79.86% | No |

Abbreviations: CI, confidence interval; RR, risk ratio.
All estimates rounded to 2 decimal places. Green indicates that the significance of the result is the same as of the 'Main' analysis.

#### *2.5.5. Supplemental eTable 5. Main and Population Subgroup Analyses Results of the 'Death' Outcome for All Population*

##### *Subgroups of Interest*

| | Main analysis (RR [95% CI], *P* value, $I^2$) | General population (RR [95% CI], *P* value, $I^2$) | Hospitalized population (RR [95% CI], *P* value, $I^2$) | COVID-19 related only (RR [95% CI], *P* value, $I^2$) | Subgroup of a population older than 50 years (RR [95% CI], *P* value, $I^2$) | CKD stage 3 (RR [95% CI], *P* value, $I^2$) | Advanced renal disease (RR [95% CI], *P* value, $I^2$) |
|---|---|---|---|---|---|---|---|
| **Autoimmune diseases** | 1.33 (1.20, 1.48), $P < .05$, $I^2 = 65\%$ | 1.33 (1.20, 1.48), $P < .001$, $I^2 = 70.93\%$ | 1.84 (0.72, 4.69), $P = .202$, $I^2 = 0\%$ | 1.38 (1.27, 1.51), $P < .001$, $I^2 = 50.04\%$ | 1.46 (1.24, 1.71), $P < .001$, $I^2 = 0\%$ | NA | NA |
| **Cancer** | 2.40 (1.22, 2.74), $P < .05$, $I^2 = 97\%$ | 2.61 (2.16, 3.16), $P < .001$, $I^2 = 97.48\%$ | 2.07 (1.55, 2.77), $P < .001$, $I^2 = 91.12\%$ | 2.78 (2.20, 3.51), $P < .001$, $I^2 = 97.6\%$ | 2.81 (2.39, 3.31), $P < .001$, $I^2 = 21.09\%$ | NA | NA |
| **Immunocompromised/ immunosuppressed** | 2.17 (1.77, 2.66), $P < .05$, $I^2 = 97\%$ | 2.303 (1.77, 3.00), $P < .001$, $I^2 = 96.89\%$ | 1.82 (1.38, 2.41), $P < .001$, $I^2 = 60.62\%$ | 2.61 (2.09, 3.25), $P < .001$, $I^2 = 93.66\%$ | NA | NA | NA |
| **Liver disease** | 1.79 (1.31, 2.43), $P < .05$, $I^2 = 96\%$ | 2.767 (2.19, 3.50), $P < .001$, $I^2 = 70.63\%$ | 1.20 (0.94, 1.53), $P = .142$, $I^2 = 85.03\%$ | 3.34 (2.95, 3.78), $P < .001$, $I^2 = 0\%$ | 2.677 (2.00, 3.59), $P < .001$, $I^2 = 0\%$ | NA | NA |
| **Renal disease** | 2.15 (1.88, 2.46), $P < .05$, $I^2 = 98\%$ | 2.558 (2.09, 3.14), $P < .001$, $I^2 = 98.02\%$ | 1.35 (1.18, 1.55), $P < .001$, $I^2 = 93.47\%$ | 2.51 (2.00, 3.16), $P < .001$, $I^2 = 98.95\%$ | 1.88 (1.23, 2.87), $P = .00367$, $I^2 = 68.48\%$ | 1.38 (1.17, 1.64), $P < .001$, $I^2 = 94\%$ | 3.57 (2.06, 6.19), $P < .001$, $I^2 = 98\%$ |
| **Transplant** | 6.78 (4.41, 10.43), $P < .05$, $I^2 = 80\%$ | 8.499 (5.70, 12.67), $P < .001$, $I^2 = 74.81\%$ | 2.30 (1.08, 4.88), $P = .03$, $I^2 = 0\%$ | 8.50 (5.70, 12.67), $P < .001$, $I^2 = 74.81\%$ | NA | NA | NA |

Abbreviations: CI, confidence interval; CKD, chronic kidney disease; NA, not applicable; RR, risk ratio.
All estimates rounded to 2 decimal places. Green indicates that the significance of the result is the same as of the 'Main' analysis; Orange indicates that the significance of the result is different from the main analysis.

#### *2.5.6. Supplemental eTable 6. Main and Sensitivity Analyses Results of the 'Hospitalization' Outcome*

| | Main analysis (RR [95% CI], *P* value, $I^2$) | Statistical evidence of publication bias (yes/no) | Least adjusted (RR [95% CI], *P* value, $I^2$) | Only adjusted (RR [95% CI], *P* value, $I^2$) | Excluding studies for population overlap (RR [95% CI], *P* value, $I^2$) | Leave-1-out (yes/no for significant difference of removing a subgroup) |
|---|---|---|---|---|---|---|
| **Autoimmune diseases** | 1.76 (1.42, 2.16), $P < .05$, $I^2$ = 97% | NA | 1.79 (1.44, 2.22), $P < .001$, $I^2$ = 97.54% | 1.76 (1.42, 2.17), $P < .001$, $I^2$ = 97.36% | 1.76 (1.42, 2.17), $P < .001$, $I^2$ = 97.36% | No |
| **Cancer** | 2.18 (1.82, 2.61), $P < .05$, $I^2$ = 98% | No | 2.30 (1.74, 3.03), $P < .001$, $I^2$ = 99.25% | 2.18 (1.82, 2.61), $P < .001$, $I^2$ = 98.02% | 2.18 (1.82, 2.61), $P < .001$, $I^2$ = 98.02% | No |
| **Immunocompromised/ immunosuppressed** | 2.75 (2.11, 3.59), $P < .05$, $I^2$ = 98% | No | 2.77 (2.14, 3.59), $P < .001$, $I^2$ = 97.93% | 2.75 (2.11, 3.59), $P < .001$, $I^2$ = 98.05% | 2.75 (2.11, 3.59), $P < .001$, $I^2$ = 98.05% | No |
| **Renal disease** | 2.13 (1.64, 2.76), $P < .05$, $I^2$ = 99% | No | 2.14 (1.65, 2.76), $P < .001$, $I^2$ = 98.7% | 2.13 (1.64, 2.76), $P < .001$, $I^2$ = 98.72% | 2.13 (1.64, 2.76), $P < .001$, $I^2$ = 98.72% | No |
| **Transplant** | 6.75 (3.41, 13.37), $P < .05$, $I^2$ = 98% | NA | 6.75 (3.4, 13.37), $P < .001$, $I^2$ = 97.96% | 5.34 (2.98, 9.56), $P < .001$, $I^2$ = 93.81% | 6.75 (3.41, 13.37), $P < .001$, $I^2$ = 97.96% | No |

Abbreviations: CI, confidence interval; NA, not applicable; RR, risk ratio.
All estimates rounded to 2 decimal places. Green indicates that the significance of the result is the same as of the 'Main' analysis.

##### *2.5.7. Supplemental eTable 7. Main and Population Subgroup Analyses Results of the 'Hospitalization' Outcome for All Subgroups of Interest*

| | Main analysis (RR [95% CI], $P$ value, $I^2$) | COVID-19 related only (RR [95% CI], $P$ value, $I^2$) | Subgroup of a population older than 50 years (RR [95% CI], $P$ value, $I^2$) | Including children (RR [95% CI], $P$ value, $I^2$) |
|---|---|---|---|---|
| **Autoimmune diseases** | 1.76 (1.42, 2.16), $P < .05$, $I^2 = 97\%$ | 1.81 (1.48, 2.20), $P < .001$, $I^2 = 94.3\%$ | NA | NA |
| **Cancer** | 2.18 (1.82, 2.61), $P < .05$, $I^2 = 98\%$ | 2.52 (1.93, 3.30), $P < .001$, $I^2 = 98.39\%$ | NA | NA |
| **Immunocompromised/ immunosuppressed** | 2.76 (2.11, 3.59), $P < .05$, $I^2 = 98\%$ | 2.88 (2.06, 4.034), $P < .001$, $I^2 = 98.41\%$ | NA | 2.92 (2.24, 3.81), $P < .001$, $I^2 = 97.96\%$ |
| **Renal disease** | 2.13 (1.64, 2.76), $P < .05$, $I^2 = 99\%$ | 2.25 (1.57, 3.21), $P < .001$, $I^2 = 98.96\%$ | 1.79 (1.14, 2.82), $P = .0114$, $I^2 = 87.14\%$ | NA |
| **Transplant** | 6.75 (3.41, 13.37), $P < .05$, $I^2 = 98\%$ | 6.75 (3.41, 13.37), $P < .001$, $I^2 = 97.96\%$ | NA | NA |

Abbreviations: CI, confidence interval; NA, not applicable; RR, risk ratio.
All estimates rounded to 2 decimal places. Green indicates that the significance of the result is the same as of the 'Main' analysis.

##### *2.5.8. Supplemental eTable 8. Main and Sensitivity Analyses Results of the 'ICU Admission' Outcome*

| | Main analysis (RR [95% CI], *P* value, $I^2$) | Statistical evidence of publication bias (yes/no) | Least adjusted (RR [95% CI], *P* value, $I^2$) | Only adjusted (RR [95% CI], *P* value, $I^2$) | Excluding studies for population overlap (RR [95% CI], *P* value, $I^2$) | Leave-1-out (yes/no for significant difference of removing a subgroup) |
|---|---|---|---|---|---|---|
| **Cancer** | 2.09 (1.13, 3.89), $P < .05$, $I^2$ = 98% | NA | 2.00 (0.96, 4.16), $P$ = .0647, $I^2$ = 98.93% | 2.66 (1.34, 5.26), $P$ = .00498, $I^2$ = 98.72% | 2.10 (1.13, 3.89), $P$ = .019, $I^2$ = 98.3% | Yes |
| **Immunocompromised/ immunosuppressed** | 3.38 (2.37, 4.83), $P < .05$, $I^2$ = 79% | NA | 3.43 (2.41, 4.88), $P < .001$, $I^2$ = 78.32% | 4.42 (3.92, 4.99), $P < .001$, $I^2$= 0% | 3.38 (2.37, 4.83), $P < .001$, $I^2$ = 78.72% | No |
| **Liver disease** | 1.27 (1.07, 1.52), $P < .05$, $I^2$ = 68% | NA | 1.41 (1.14, 1.75), $P$ = .0017, $I^2$ = 86.79% | 1.28 (1.07, 1.52), $P$ = .00741, $I^2$ = 68.18% | 1.28 (1.07, 1.52), $P$ = .00741, $I^2$ = 68.18% | No |
| **Renal disease** | 1.96 (1.19, 3.22), $P < .05$, $I^2$ = | No | 2.25 (1.33, 3.79), $P$ = .00236, $I^2$ = 96.8% | 2.89 (1.56, 5.34), $P < .001$, $I^2$ = 97.74% | 1.96 (1.19, 3.22), $P$ = .00802, $I^2$ = 96.05% | Yes |

Abbreviations: CI, confidence interval; NA, not applicable; RR, risk ratio.
All estimates rounded to 2 decimal places. Green indicates that the significance of the result is the same as of the 'Main' analysis; Orange indicates that the significance of the result is different from the main analysis.

##### *2.5.9. Supplemental eTable 9. Main and Population Subgroup Analyses Results of the 'ICU' Outcome for All Population*

##### *Subgroups of Interest*

| | **Main analysis (RR [95% CI], *P* value, $I^2$)** | **General population (RR [95% CI], *P* value, $I^2$)** | **Hospitalized population (RR [95% CI], *P* value, $I^2$)** | **COVID-19 related only (RR [95% CI], *P* value, $I^2$)** | **Subgroup of a population older than 50 years (RR [95% CI], *P* value, $I^2$)** |
|---|---|---|---|---|---|
| **Cancer** | 2.09 (1.13, 3.89), *P* < .05, $I^2$ = 98% | 3.37 (1.10, 10.37), *P* = .0337, $I^2$ = 98.36% | 1.59 (0.83, 3.06), *P* = .163, $I^2$ = 85.56% | 3.85 (1.48, 10.05), *P* = .00581, $I^2$ = 97.6% | NA |
| **Immunocompromised/ immunosuppressed** | 3.38 (2.37, 4.83), *P* < .05, $I^2$ = 79% | 4.49 (3.97, 5.08), *P* < .001, $I^2$ = 0% | 2.39 (1.05, 5.46), *P* = .0379, $I^2$ = 71.07% | 4.42 (3.91, 4.98), *P* < .001, $I^2$ = 0% | NA |
| **Liver disease** | 1.27 (1.07, 1.52), *P* < .05, $I^2$ = 68% | NA | 1.28 (1.07, 1.52), *P* = .00741, $I^2$ = 68.18% | NA | NA |
| **Renal disease** | 1.96 (1.19, 3.22), *P* < .05, $I^2$ = 96% | 3.00 (0.45, 20.11), *P* = .259, $I^2$ = 98.61% | 1.53 (0.89, 2.62), *P* = .122, $I^2$ = 81.33% | 3.20 (0.92, 11.07), *P* = .067, $I^2$ = 91.86% | NA |

Abbreviations: CI, confidence interval; NA, not applicable; RR, risk ratio.
All estimates rounded to 2 decimal places. Green indicates that the significance of the result is the same as of the 'Main' analysis; Orange indicates that the significance of the result is different from the main analysis.

##### *2.5.10. Supplemental eTable 10. Main and Sensitivity Analyses Results of the 'Combined' Outcome*

| | Main analysis (RR [95% CI], *P* value, $I^2$) | Statistical evidence of publication bias (yes/no) | Least adjusted (RR [95% CI], *P* value, $I^2$) | Only adjusted (RR [95% CI], *P* value, $I^2$) | Excluding studies for population overlap (RR [95% CI], *P* value, $I^2$) | Leave-1-out (yes/no for significant difference of removing a subgroup) |
|---|---|---|---|---|---|---|
| **Autoimmune diseases** | 2.21 (1.61, 3.04), $P < .05$, $I^2 = 89\%$ | NA | 2.21 (1.61, 3.04), $P < .001$, $I^2 = 88.61\%$ | 2.22 (1.60, 3.08), $P < .001$, $I^2 = 93.95\%$ | 2.21 (1.61, 3.04), $P < .001$, $I^2 = 88.61\%$ | No |
| **Cancer** | 1.78 (1.45, 2.19), $P < .05$, $I^2 = 96\%$ | Yes | 1.72 (1.40, 2.11), $P < .001$, $I^2 = 95.85\%$ | 1.89 (1.52, 2.34), $P < .001$, $I^2 = 96.35\%$ | 1.83 (1.48, 2.24), $P < .001$, $I^2 = 96.02\%$ | No |
| **Immunocompromised/ immunosuppressed** | 2.04 (1.42, 2.94), $P < .05$, $I^2 = 99\%$ | No | 2.05 (1.42, 2.96), $P < .001$, $I^2 = 99.25\%$ | 1.94 (1.33, 2.83), $P < .001$, $I^2 = 99.29\%$ | 2.05 (1.42, 2.94), $P < .001$, $I^2 = 99.25\%$ | No |
| **Liver disease** | 1.50 (1.12, 2.00), $P < .05$, $I^2 = 97\%$ | No | 1.48 (1.10, 1.97), $P = .00864$, $I^2 = 96.64\%$ | 1.56 (1.16, 2.11), $P = .00364$, $I^2 = 97.29\%$ | 1.50 (1.12, 2.00), $P = .00678$, $I^2 = 96.65\%$ | No |
| **Renal disease** | 1.86 (1.55, 2.23), $P < .005$, $I^2 = 95\%$ | Yes | 1.81 (1.51, 2.16), $P < .001$, $I^2 = 94.67\%$ | 1.93 (1.58, 2.36), $P < .001$, $I^2 = 96.13\%$ | 1.81 (1.50, 2.17), $P < .001$, $I^2 = 95\%$ | No |
| **Transplant** | 8.65 (4.01, 18.65), $P < .05$, $I^2 = 97\%$ | NA | 8.65 (4.01, 18.65), $P < .001$, $I^2 = 97\%$ | 7.90 (3.21, 19.41), $P < .001$, $I^2 = 97.96\%$ | 8.65 (4.01, 18.65), $P < .001$, $I^2 = 97.32\%$ | No |

Abbreviations: CI, confidence interval; NA, not applicable; RR, risk ratio.
All estimates rounded to 2 decimal places. Green indicates that the significance of the result is the same as of the 'Main' analysis; Orange indicates that the significance of the result is different from the main analysis.

##### *2.5.11. Supplemental eTable 11. Main and Population Subgroup Analyses Results of the 'Combined' Outcome for All Population Subgroups of Interest*

| | Main analysis (RR [95% CI], *P* value, $I^2$) | General population (RR [95% CI], *P* value, $I^2$) | Hospitalized population (RR [95% CI], *P* value, $I^2$) | COVID-19 related only (RR [95% CI], *P* value, $I^2$) | Subgroup of a population older than 50 years (RR [95% CI], *P* value, $I^2$) | Including children (RR [95% CI], *P* value, $I^2$) |
|---|---|---|---|---|---|---|
| **Autoimmune diseases** | 2.21 (1.61, 3.04), $P < .05$, $I^2 = 89\%$ | 2.22 (1.60, 3.08), $P < .001$, $I^2 = 93.95\%$ | 1.91 (0.29, 12.39), $P = .5$, $I^2 = 50.49\%$ | 2.21 (1.61, 3.04), $P < .001$, $I^2 = 88.61\%$ | NA | NA |
| **Cancer** | 1.78 (1.45, 2.19), $P < .05$, $I^2 = 96\%$ | 1.96 (1.58, 2.43), $P < .001$, $I^2 = 96.71\%$ | 1.09 (0.51, 2.32), $P = .822$, $I^2 = 75.58\%$ | 1.96 (1.59, 2.43), $P < .001$, $I^2 = 96.4\%$ | NA | 1.75 (1.43, 2.14), $P < .001$, $I^2 = 95.62\%$ |
| **Immunocompromised/ immunosuppressed** | 2.04 (1.42, 2.94), $P < .05$, $I^2 = 99\%$ | 2.49 (1.65, 3.73), $P < .001$, $I^2 = 98.85\%$ | 1.26 (1.18, 1.35), $P < .001$, $I^2 = 38.73\%$ | 2.34 (1.55, 3.53), $P < .001$, $I^2 = 98.85\%$ | NA | NA |
| **Liver disease** | 1.50 (1.12, 2.00), $P < .05$, $I^2 = 97\%$ | 1.73 (0.91, 3.29), $P = 0.0921$, $I^2 = 98.26\%$ | 1.36 (1.25, 1.48), $P < .001$, $I^2 = 34.98\%$ | 1.59 (0.91, 2.79), $P = .105$, $I^2 = 96.63\%$ | NA | NA |
| **Renal disease** | 1.86 (1.55, 2.23), $P < .005$, $I^2 = 95\%$ | 2.39 (1.92, 2.96), $P < .001$, $I^2 = 91.59\%$ | 1.20 (1.05, 1.37), $P = .00688$, $I^2 = 75.43\%$ | 2.41 (1.96, 2.96), $P < .001$, $I^2 = 89.63\%$ | 2.46 (1.46, 4.17), $P < .001$, $I^2 = 42.3\%$ | NA |
| **Transplant** | 8.65 (4.01, 18.65), $P < .05$, $I^2 = 97\%$ | 8.65 (4.01, 18.65), $P < .001$, $I^2 = 97\%$ | NA | 8.65 (4.01, 18.65), $P < .001$, $I^2 = 97\%$ | NA | 7.74 (3.75, 15.99), $P < .001$, $I^2 = 96.81\%$ |

Abbreviations: CI, confidence interval; NA, not applicable; RR, risk ratio.
All estimates rounded to 2 decimal places. Green indicates that the significance of the result is the same as of the 'Main' analysis; Orange indicates that the significance of the result is different from the 'Main' analysis.

#### 2.6. Meta-analyses for Each IC/IS Condition: Forest Plots of Pooled RR or Narrative Descriptions for Each IC/IS Condition and Outcome

##### *2.6.1. Autoimmune Disease*

*2.6.1.1. Supplemental eFigure 1. Autoimmune Disease, Death Outcome (No. of Subgroups: 11)*

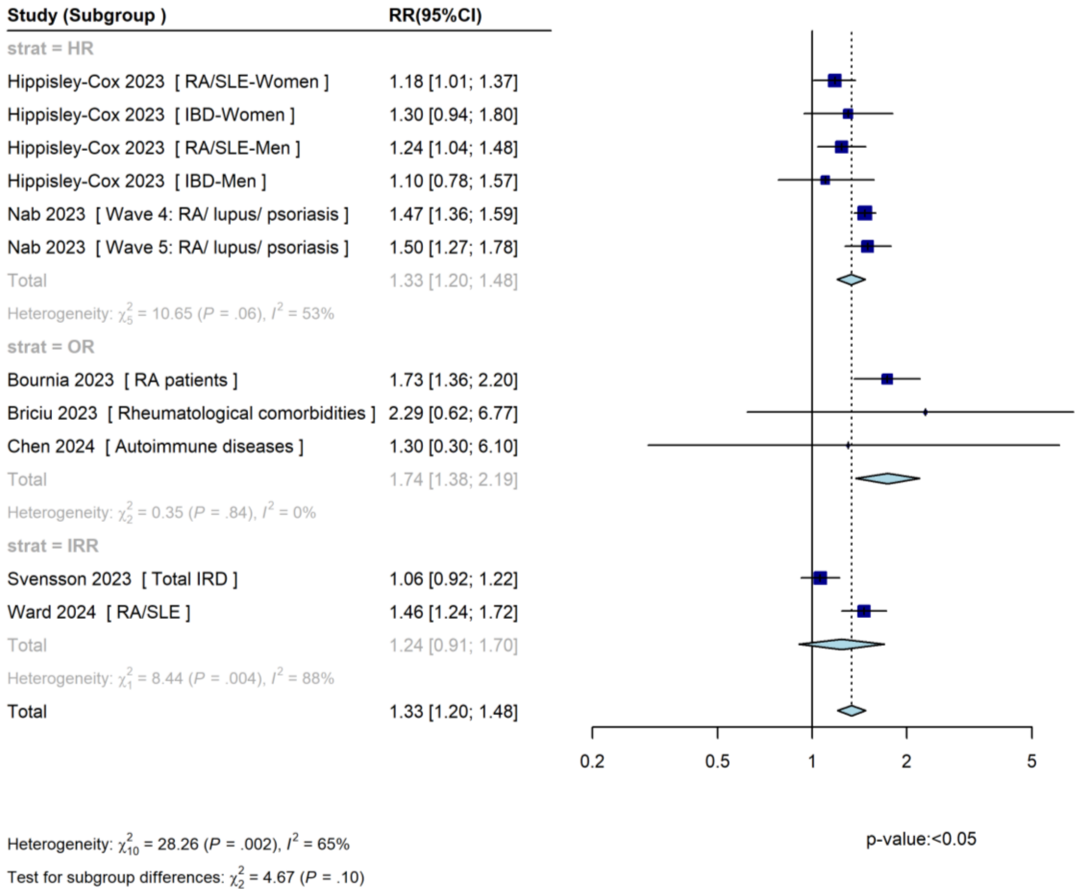

CI, confidence interval; HR, hazard ratio; IBD, inflammatory bowel disease; IRD, inflammatory rheumatic diseases; IRR, incidence rate ratio; OR, odds ratio; RA, rheumatoid arthritis; RR, risk ratio; SLE, systemic lupus erythematosus.

*2.6.1.2. Supplemental eFigure 2. Autoimmune Disease, Hospitalization Outcome (No. of Subgroups: 8)*

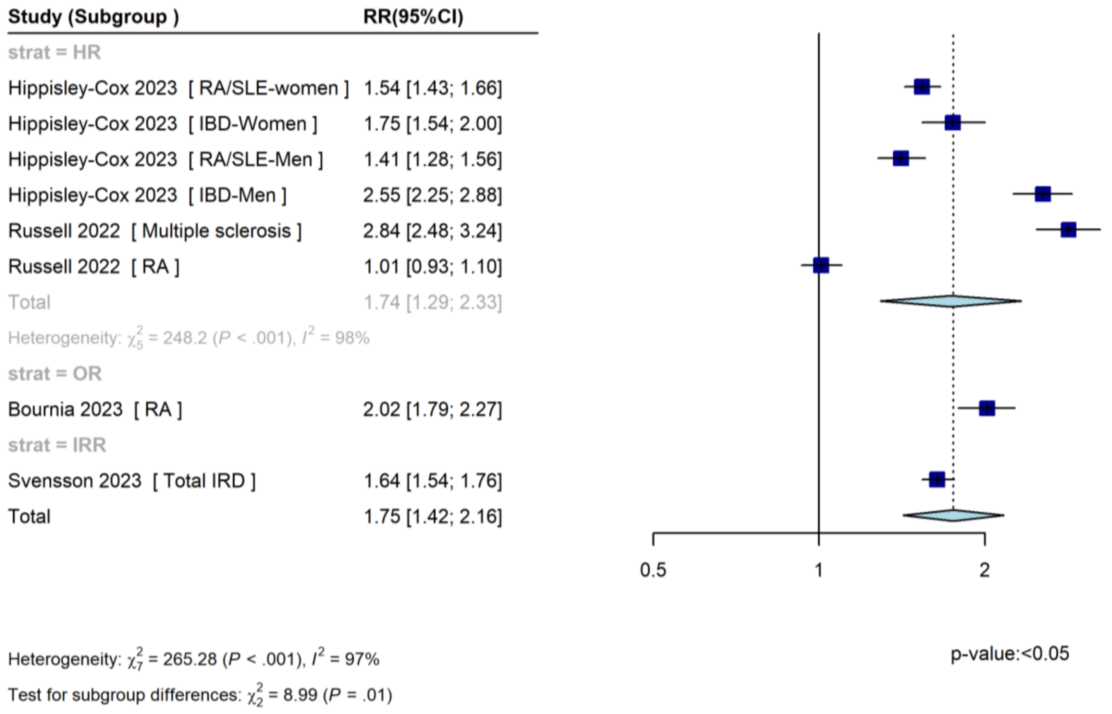

CI, confidence interval; HR, hazard ratio; IBD, inflammatory bowel disease; IRD, inflammatory rheumatic diseases; IRR, incidence rate ratio; OR, odds ratio; RA, rheumatoid arthritis; RR, risk ratio; SLE, systemic lupus erythematosus.

*2.6.1.3. Autoimmune Disease, ICU Admission Outcome (No. of Studies: 2)*

The studies reported the following results for the 'ICU' outcome in the 'Autoimmune disease' populations:

- Briciu 2023[23] (rheumatological comorbidities): OR (95% CI): 1.60 (0.51, 4.17)
- Russell 2023[9] (rheumatoid arthritis): HR (95% CI): 1.15 (0.91, 1.47)

While both studies indicated the same direction of effect – increases in the risk of the 'ICU' outcome, based on the CIs, the results were not statistically significant.

*2.6.1.4. Supplemental eFigure 3. Autoimmune Disease, Combined Outcome (No. of Subgroups: 5)*

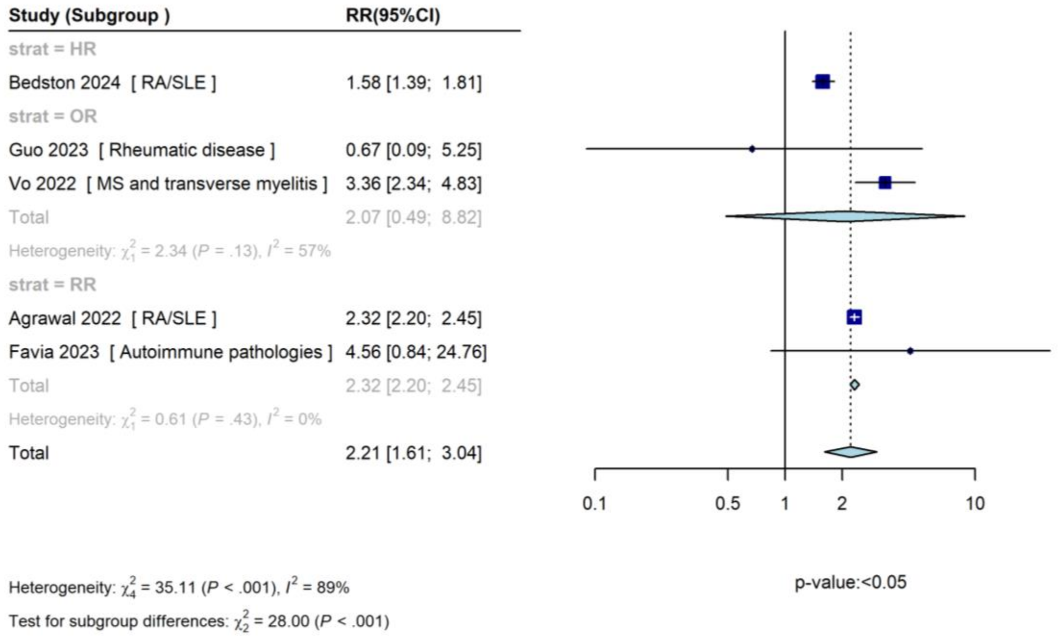

CI, confidence interval; HR, hazard ratio; MS, multiple sclerosis; OR, odds ratio; RA, rheumatoid arthritis; RR, risk ratio; SLE, systemic lupus erythematosus.

##### *2.6.2. Cancer*

*2.6.2.1. Supplemental eFigure 4. Cancer, Death Outcome (No. of Subgroups: 39)*

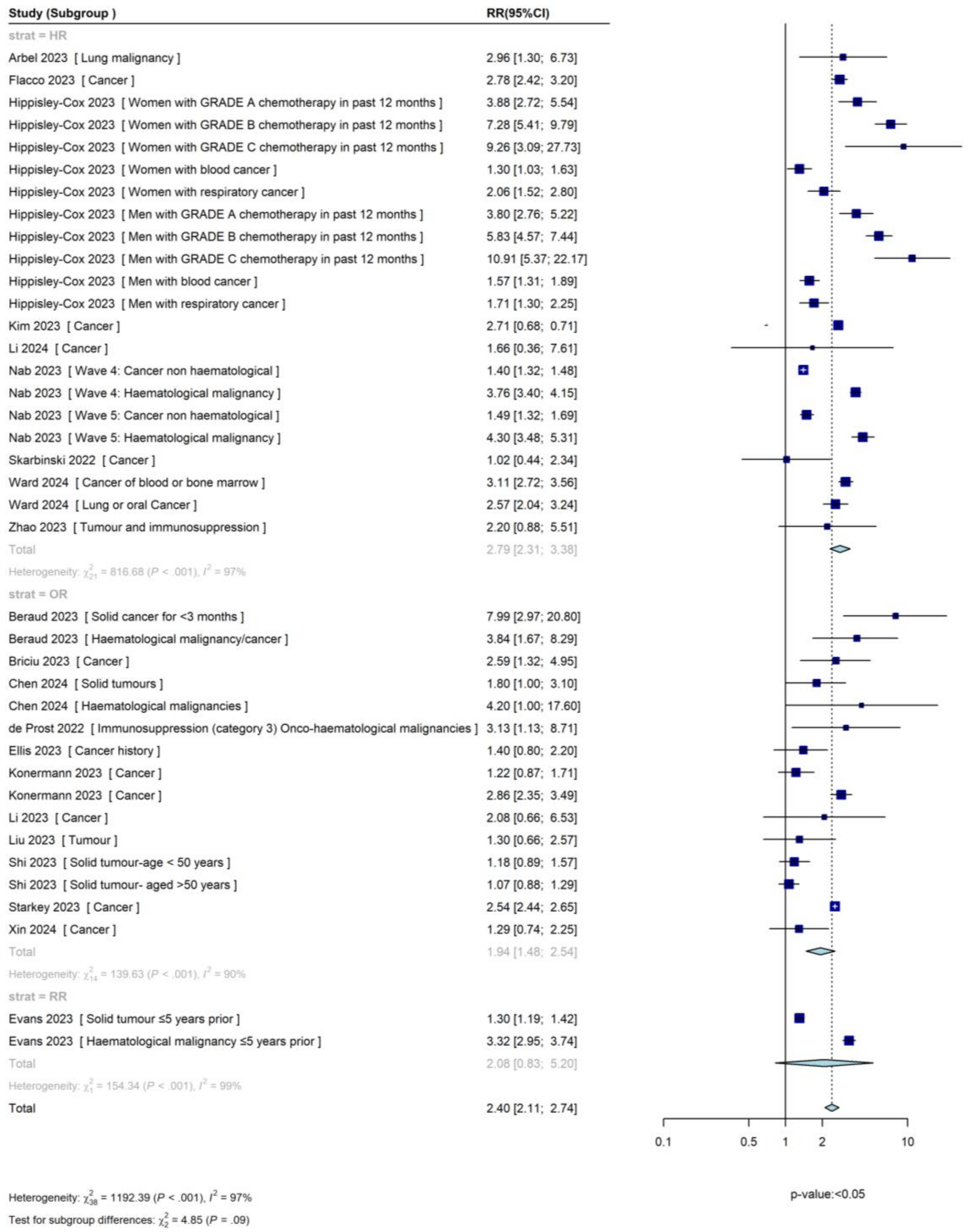

CI, confidence interval; HR, hazard ratio; IRR, incidence rate ratio; OR, odds ratio; RR, risk ratio.

*2.6.2.2. Supplemental eFigure 5. Cancer, Hospitalization Outcome (No. of Subgroups: 18)*

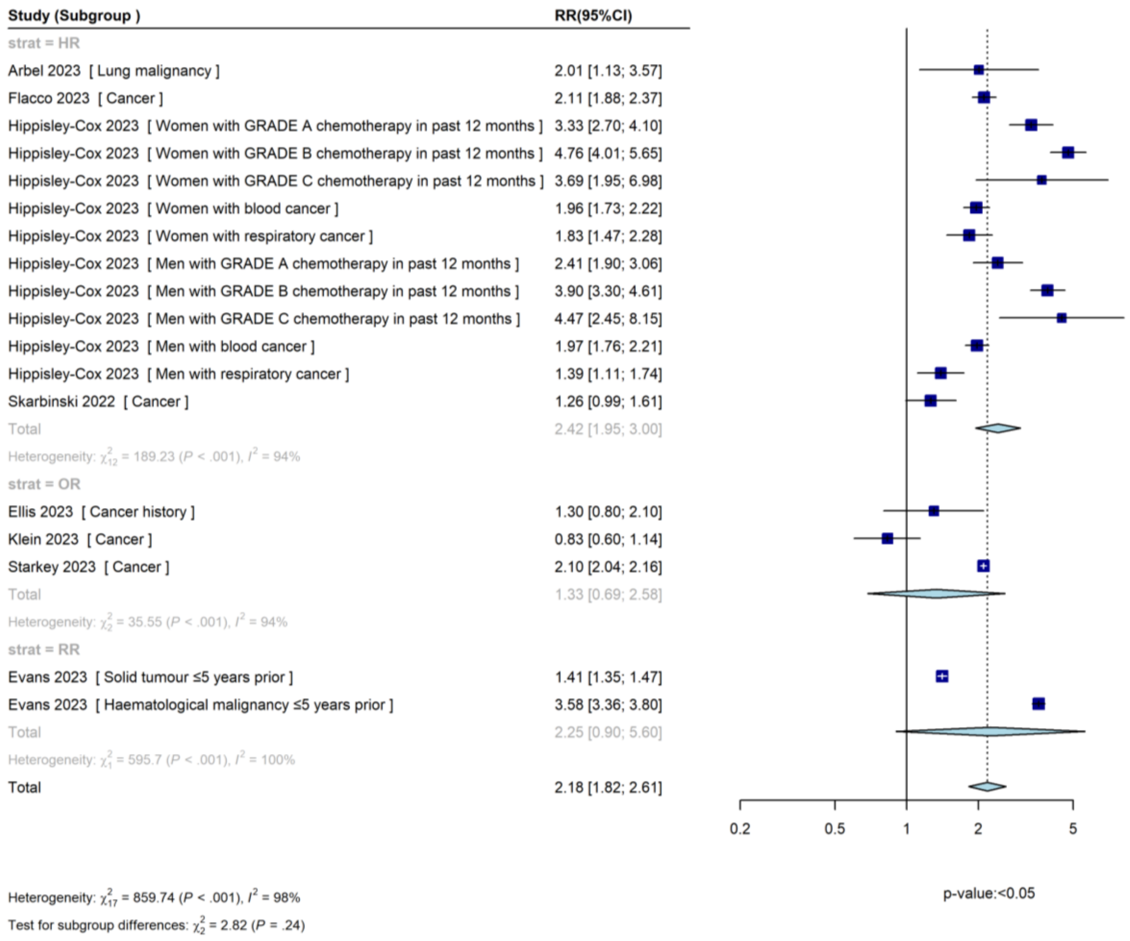

CI, confidence interval; HR, hazard ratio; IRR, incidence rate ratio; OR, odds ratio; RR, risk ratio.

*2.6.2.3. Supplemental eFigure 6. Cancer, ICU Admission Outcome (No. of Subgroups: 9)*

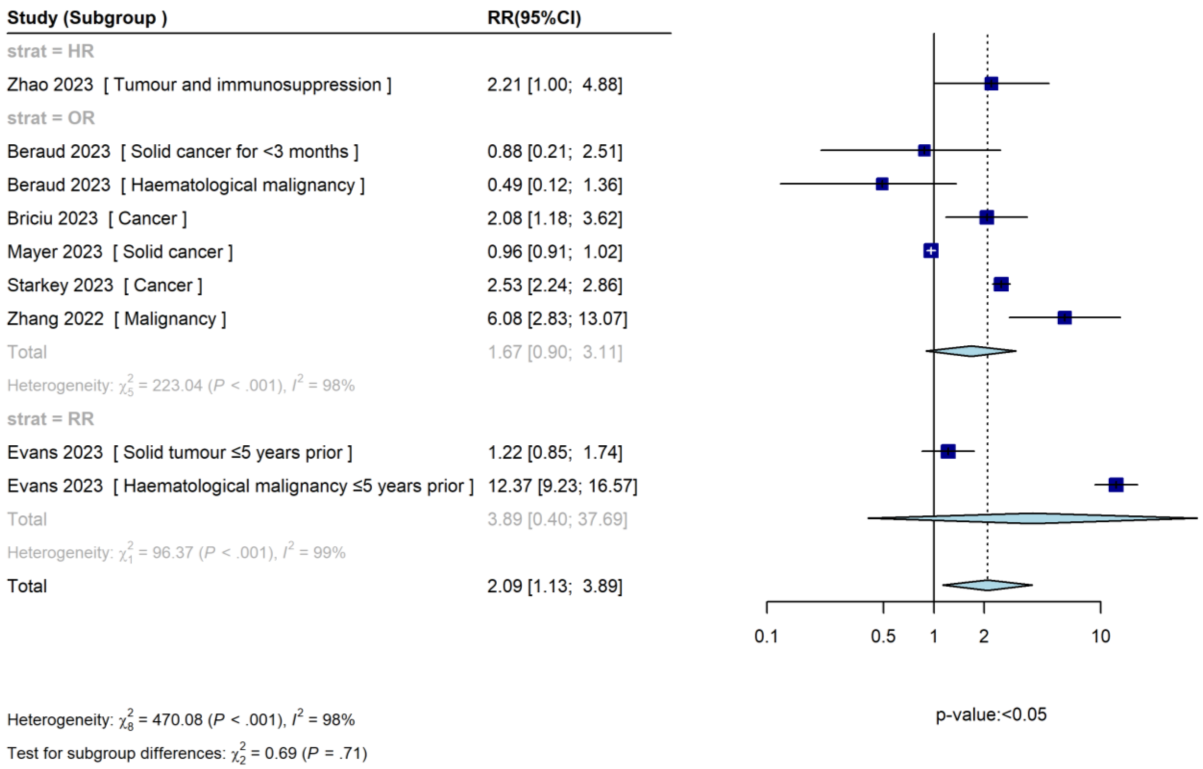

CI, confidence interval; HR, hazard ratio; ICU, intensive care unit; IRR, incidence rate ratio; OR, odds ratio; RR, risk ratio.

*2.6.2.4. Supplemental eFigure 7. Cancer, Combined Outcome (No. of Subgroups: 18)*

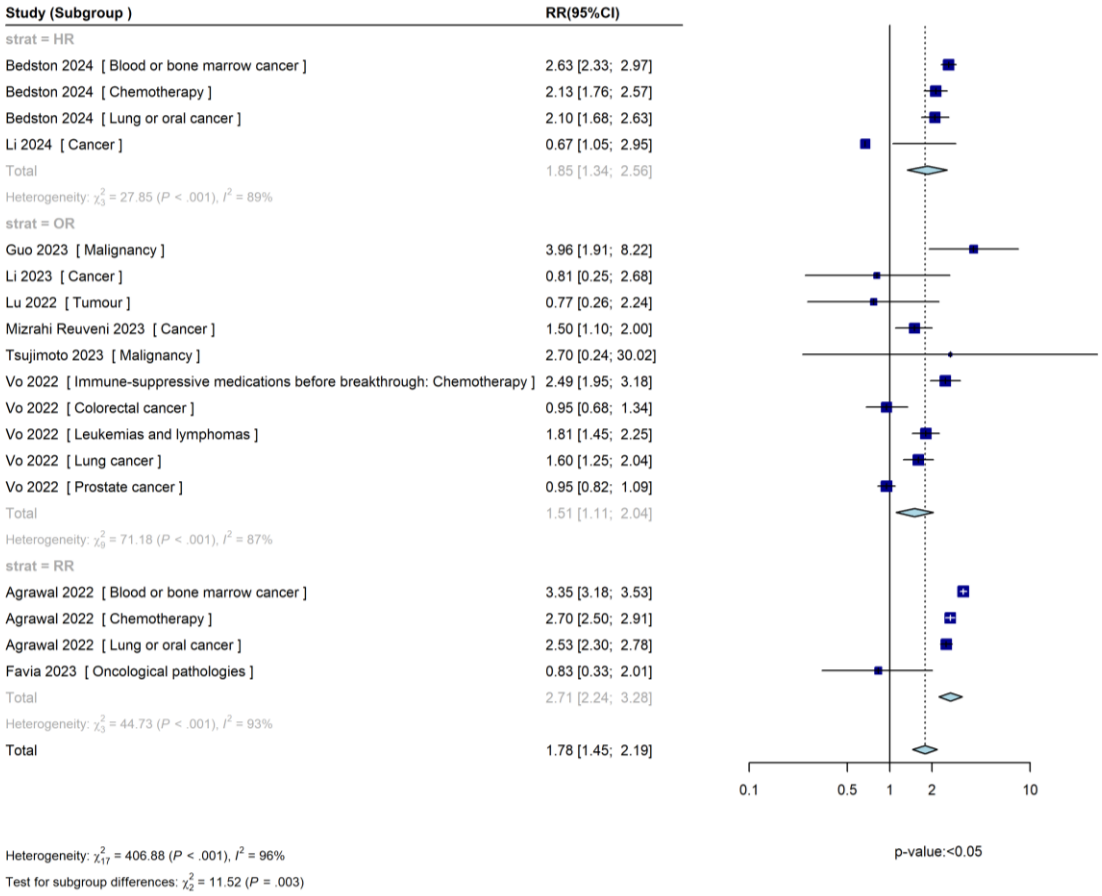

CI, confidence interval; HR, hazard ratio; IRR, incidence rate ratio; OR, odds ratio; RR, risk ratio.

###### ***2.6.3. HIV***

###### *2.6.3.1. HIV, Death Outcome (No. of Studies: 1; No. of Subgroups: 1)*

The study reported the following results for the ‘Death’ outcome in the ‘HIV’ populations:

- Evans 2023[33] (advanced or untreated HIV): incidence rate ratio (IRR) (95% CI): 3.12 (1.13, 11.39)

Based on the CI, Evans 2023 showed a statistically significant increase in the risk of the ‘Death’ outcome in the ‘HIV’ populations.

###### *2.6.3.2. HIV, Hospitalization Outcome (No. of Studies: 3; No. of Subgroups: 3)*

The studies reported the following results for the ‘Hospitalization’ outcome in the ‘HIV’ populations:

- Evans 2023[33] (advanced or untreated HIV): IRR (95% CI): 2.26 (1.11, 4.58)
- Puyat 2023[62] (all people living with HIV): HR (95% CI): 1.01 (0.78, 1.29)
- Rasmussen 2023[64] (people with HIV at first half of year 2022): IRR (95% CI): 2.0 (1.4, 2.8)

All studies indicated the same direction of effect – increases in the risk of the ‘Hospitalization’ outcome, based on the CIs, but the result of Puyat 2023 was not statistically significant.

###### *2.6.3.3. HIV, ICU Admission Outcome (No. of Studies: 0; No. of Subgroups: 0)*

The analysis of the ‘ICU’ outcome in the ‘HIV’ populations is not feasible as no studies were included in this analysis.

###### *2.6.3.4. HIV, Combined Outcome (No. of Studies: 2; No. of Subgroups: 2)*

The studies reported the following results for the ‘Combined’ outcome in the ‘HIV’ populations:

- Agrawal 2022[12] (HIV): RR (95% CI): 0.61 (0.35, 1.08)
- Vo 2022[72] (HIV/AIDS): OR (95% CI): 1.23 (0.84, 1.80)

The analysis shows conflicting results in terms of the direction of effects of both studies. However, both results are not statistically significant.

### *2.6.4. IC/IS*

*2.6.4.1. Supplemental eFigure 8. IC/IS, Death Outcome (No. of Subgroups: 24)*

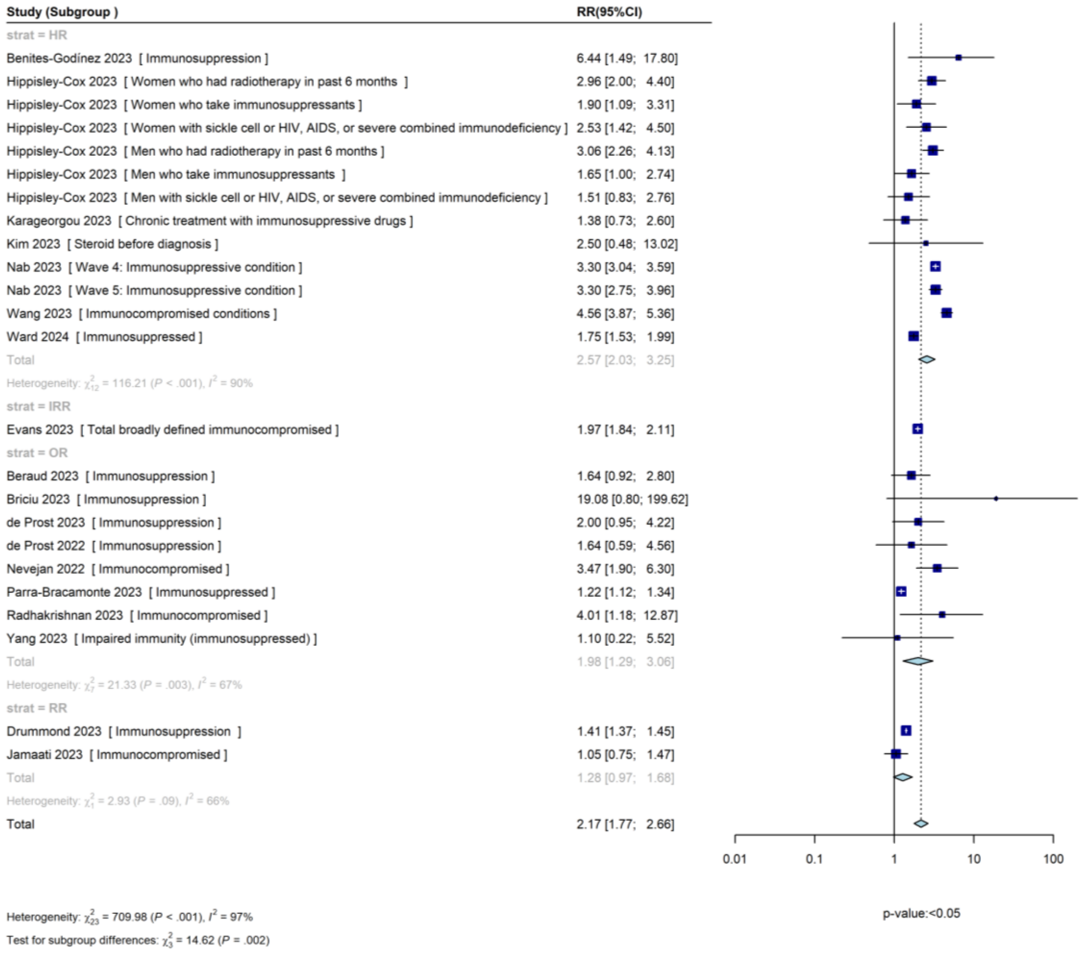

AIDS, acquired immune deficiency syndrome; CI, confidence interval; HR, hazard ratio; IC/IS, immunocompromising/immunosuppressing; IRR, incidence rate ratio; OR, odds ratio; RR, risk ratio.

*2.6.4.2. Supplemental eFigure 9. IC/IS, Hospitalization Outcome (No. of Subgroups: 16)*

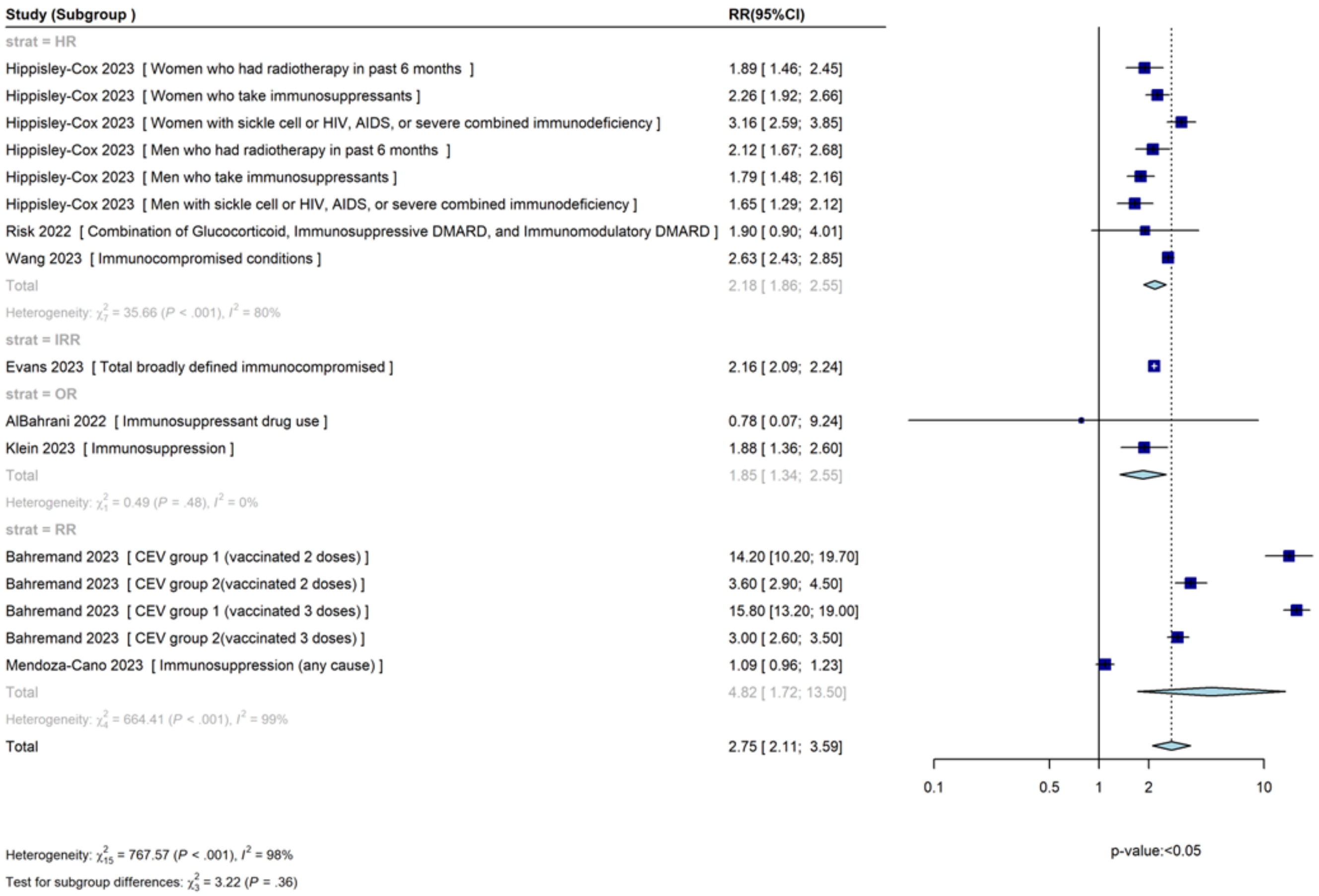

AIDS, acquired immune deficiency syndrome; CEV, clinically extremely vulnerable; CI, confidence interval; DMARD, disease-modifying anti-rheumatic drugs; HR, hazard ratio; IC/IS, immunocompromising/immunosuppressive; IRR, incidence rate ratio; OR, odds ratio; RR, risk ratio.

*2.6.4.3. Supplemental eFigure 10. IC/IS, ICU Admission Outcome (No. of Subgroups: 5)*

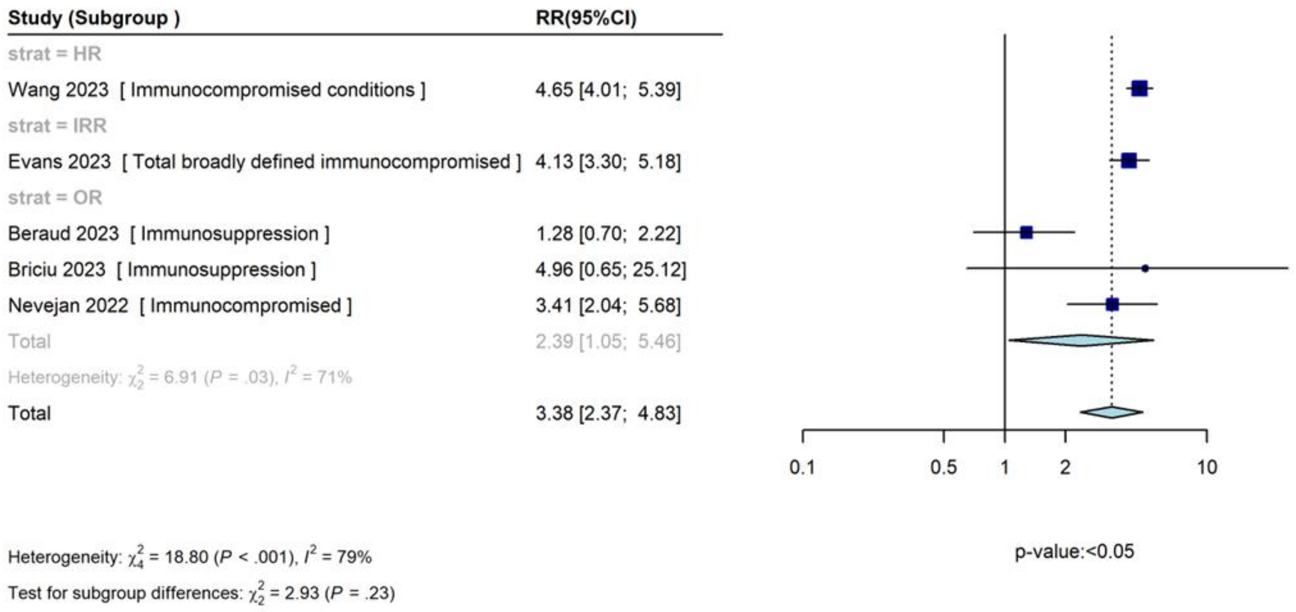

CI, confidence interval; HR, hazard ratio; IC/IS, immunocompromising/immunosuppressing; ICU, intensive care unit; IRR, incidence rate ratio; OR, odds ratio; RR, risk ratio.

*2.6.4.4. Supplemental eFigure 11. IC/IS, Combined Outcome (No. of subgroups: 17)*

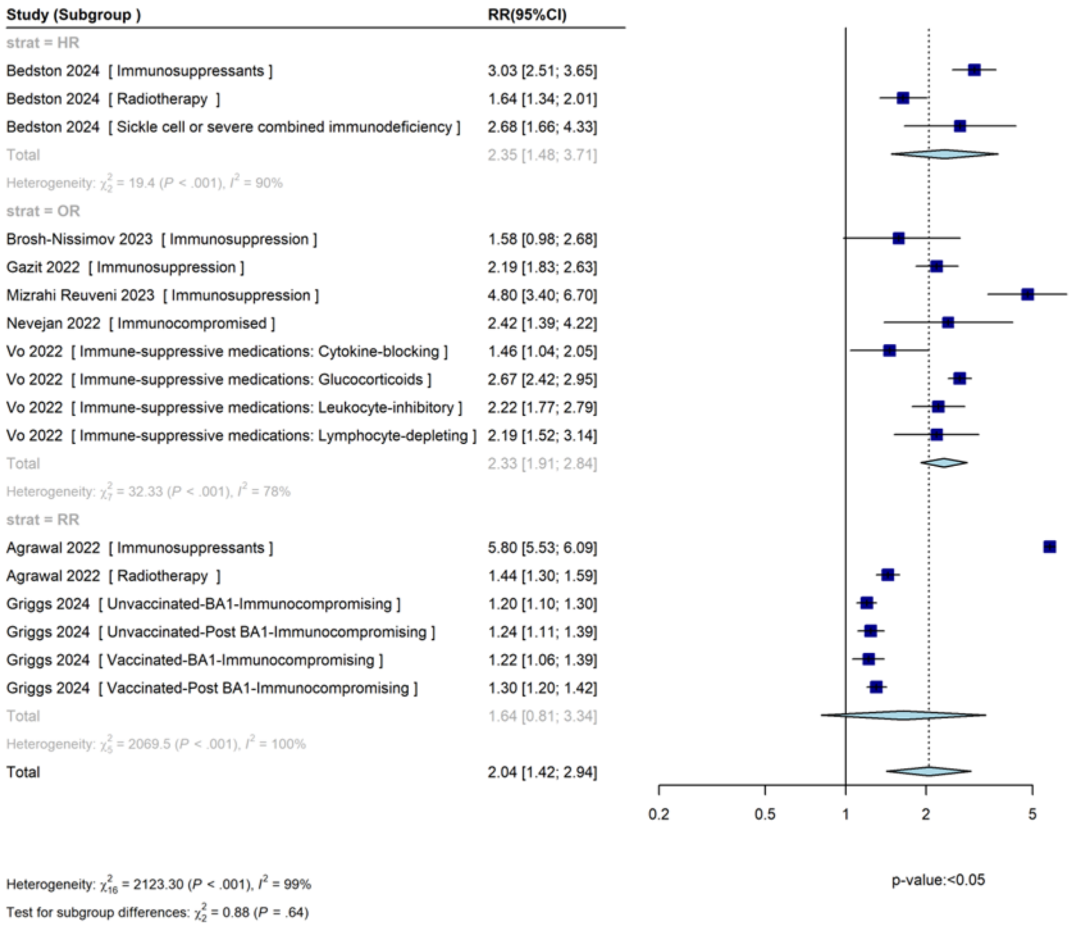

CI, confidence interval; HR, hazard ratio; IC/IS, immunocompromising/immunosuppressing; IRR, incidence rate ratio; OR, odds ratio; RR, risk ratio.

##### *2.6.5. Liver Disease*

*2.6.5.1. Supplemental eFigure 12. Liver Disease, Death Outcome (No. of Subgroups: 14)*

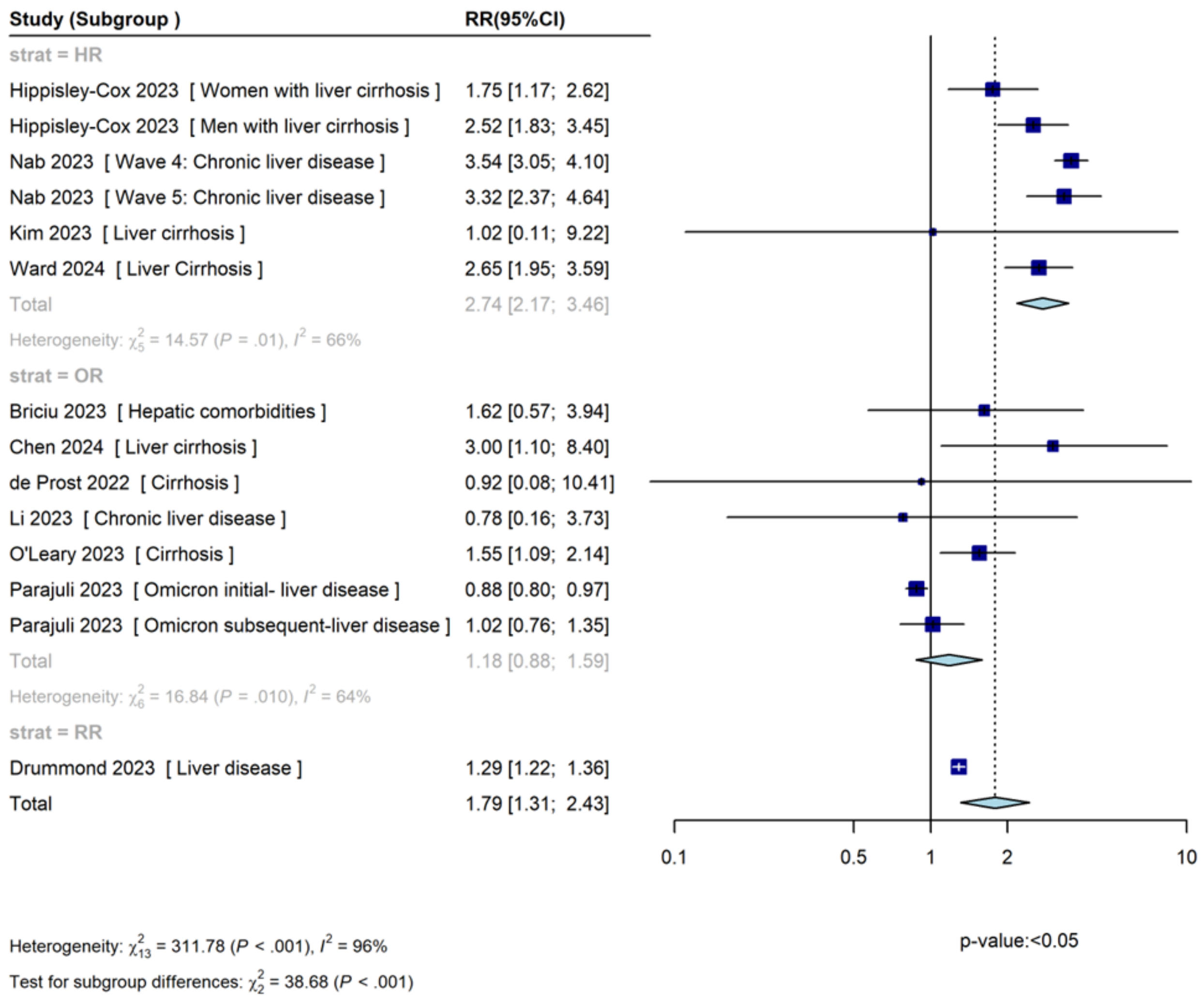

CI, confidence interval; HR, hazard ratio; IRR, incidence rate ratio; OR, odds ratio; RR, risk ratio.

*2.6.5.2. Liver Disease, Hospitalization Outcome (No. of Studies: 1; No. of Subgroups: 2)*

The studies reported the following results for the 'Hospitalization' outcome in the 'Liver disease' populations:

- Hippisley-Cox 2023[40] (women with liver cirrhosis): HR (95% CI): 2.40 (1.97, 2.92)
- Hippisley-Cox 2023[40] (men with liver cirrhosis): HR (95% CI): 2.18 (1.80, 2.63)

Both liver cirrhosis subgroups in the Hippisley-Cox 2023 study found statistically significant increases in the risk of 'Hospitalization'.

*2.6.5.3. Supplemental eFigure 13. Liver Disease, ICU Admission Outcome (No. of Subgroups: 5)*

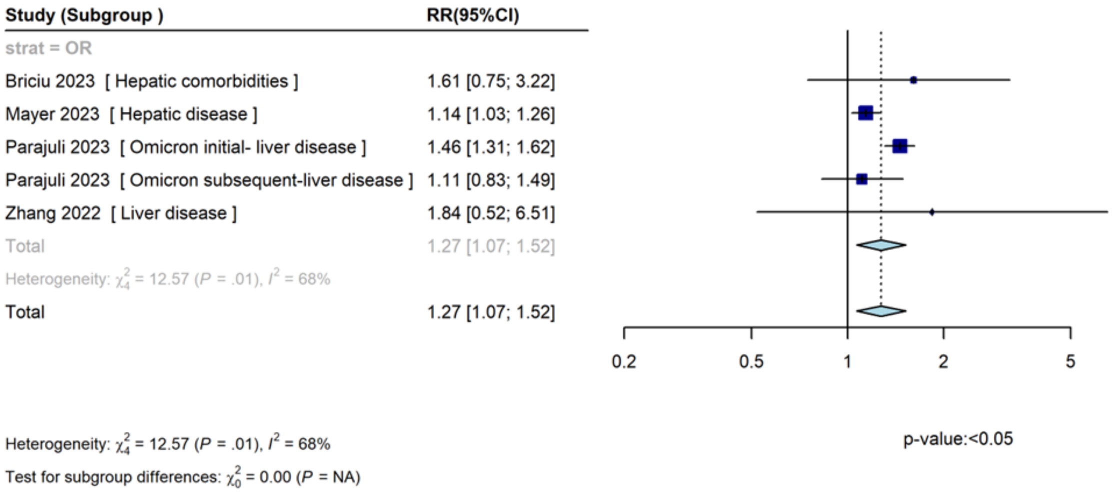

CI, confidence interval; HR, hazard ratio; ICU, intensive care unit; IRR, incidence rate ratio; NA, not applicable; OR, odds ratio; RR, risk ratio.

*2.6.5.4. Supplemental eFigure 14. Liver Disease, Combined Outcome (No. of Subgroups: 11)*

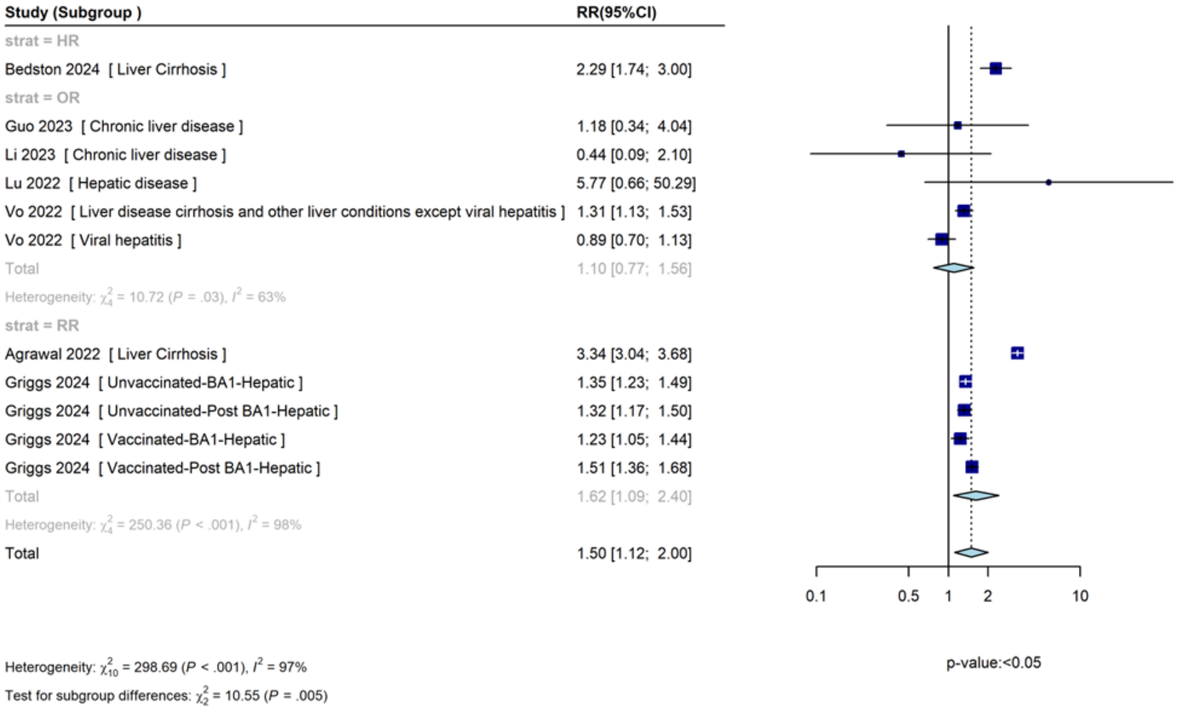

CI, confidence interval; HR, hazard ratio; IRR, incidence rate ratio; OR, odds ratio; RR, risk ratio.

1169 ***2.6.6. Renal Disease***

1170 *2.6.6.1. Supplemental eFigure 15. Renal Disease, Death Outcome (No. of Subgroups: 44)*

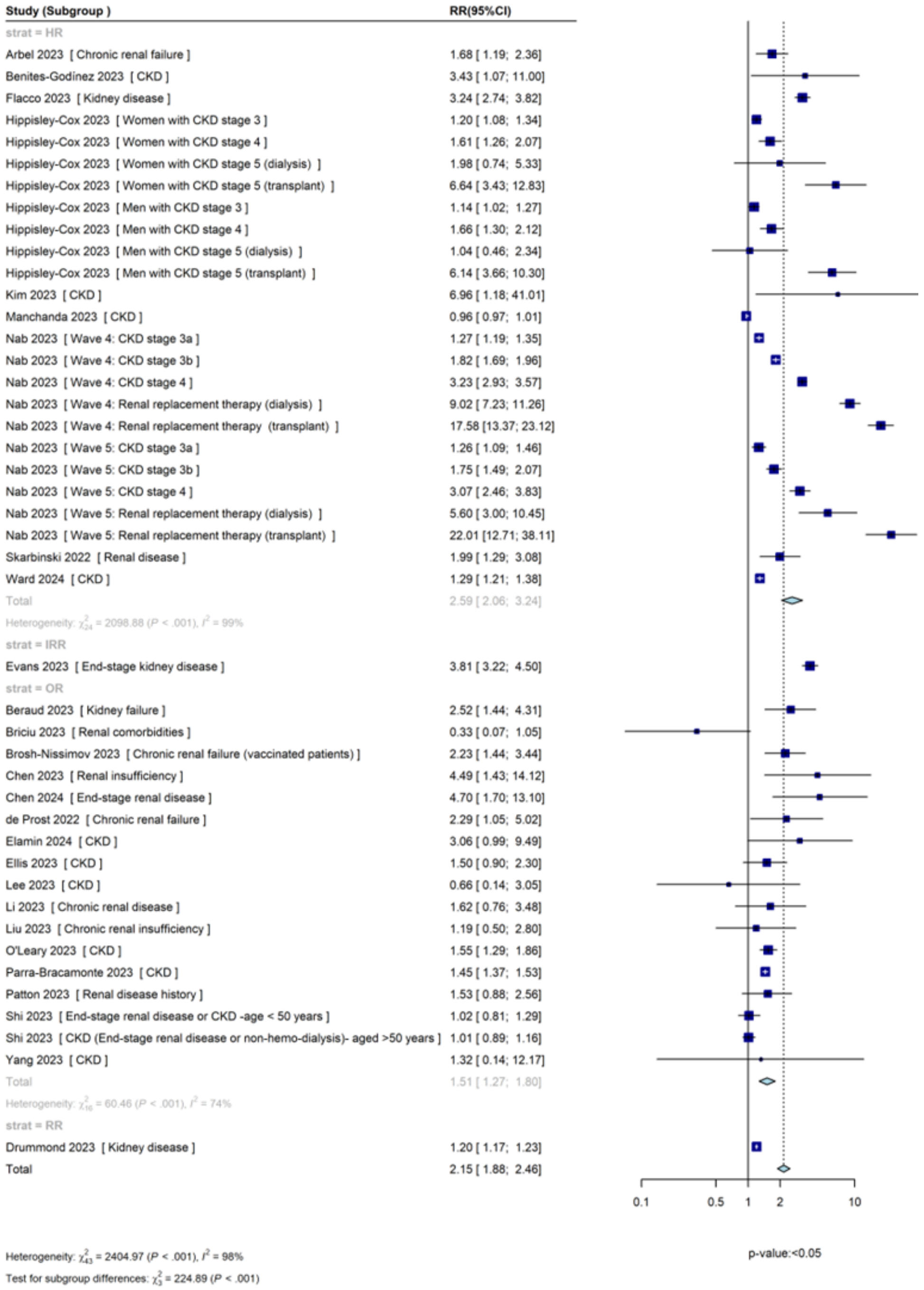

1171

1172 CI, confidence interval; CKD, chronic kidney disease; HR, hazard ratio; IRR, incidence rate ratio; OR, odds ratio; RR, risk ratio.

*2.6.6.2. Supplemental eFigure 16. Renal Disease, Hospitalization Outcome (No. of Subgroups: 17)*

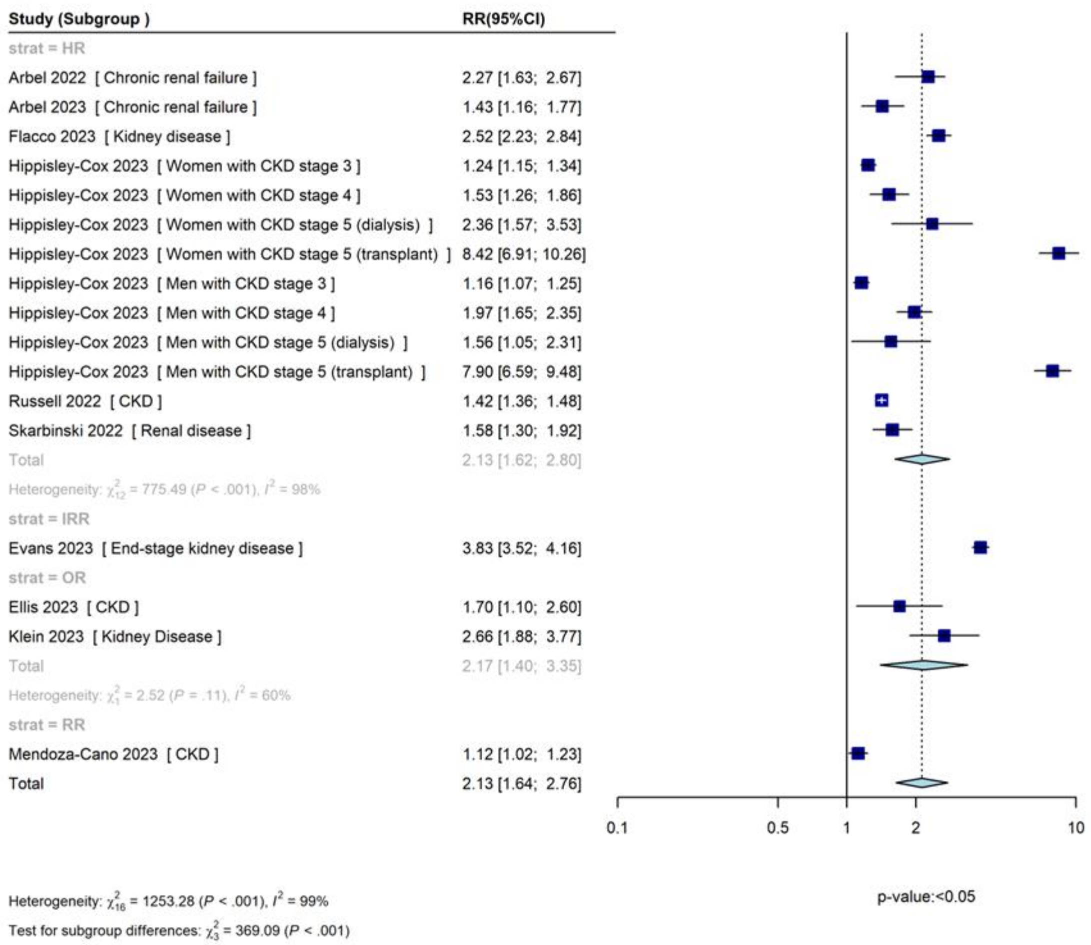

CI, confidence interval; CKD, chronic kidney disease; HR, hazard ratio; IRR, incidence rate ratio; OR, odds ratio; RR, risk ratio.

*2.6.6.3. Supplemental eFigure 17. Renal Disease, ICU Admission Outcome (No. of Subgroups: 10)*

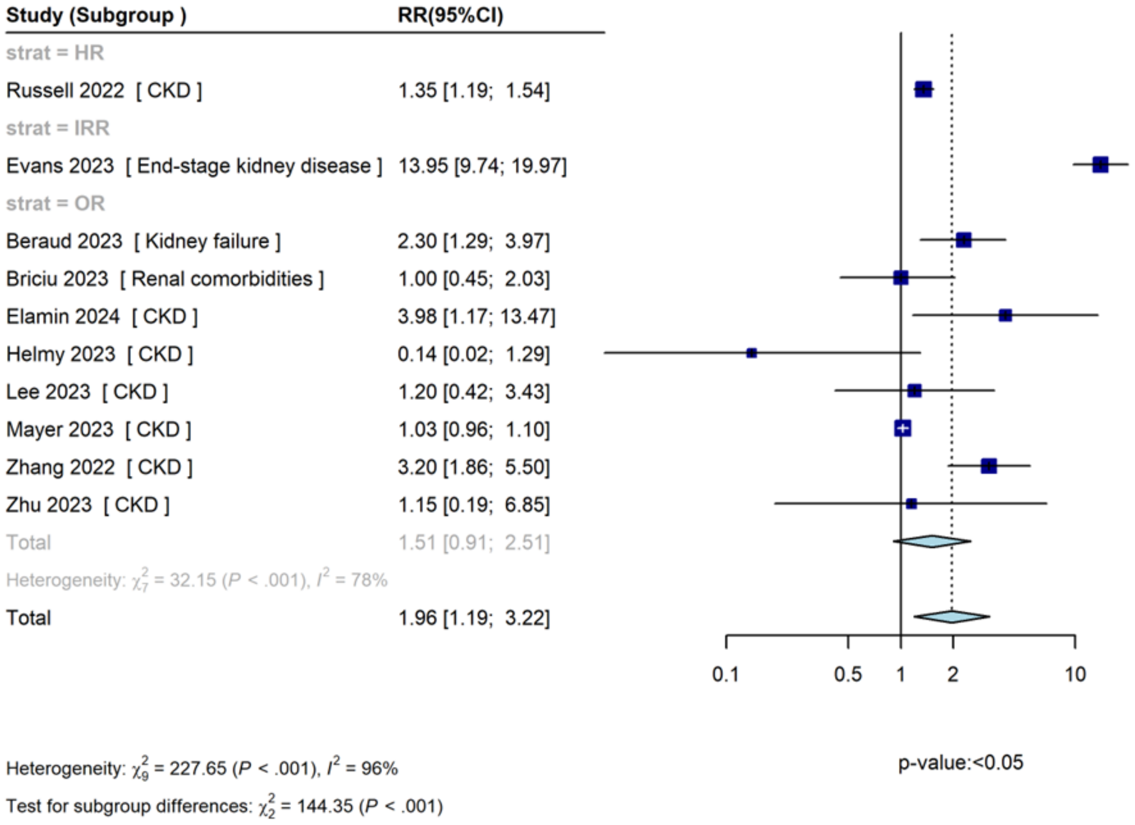

CI, confidence interval; CKD, chronic kidney disease; HR, hazard ratio; ICU, intensive care unit; IRR, incidence rate ratio; OR, odds ratio; RR, risk ratio.

*2.6.6.4. Supplemental eFigure 18. Renal Disease, Combined Outcome (No. of Subgroups: 21)*

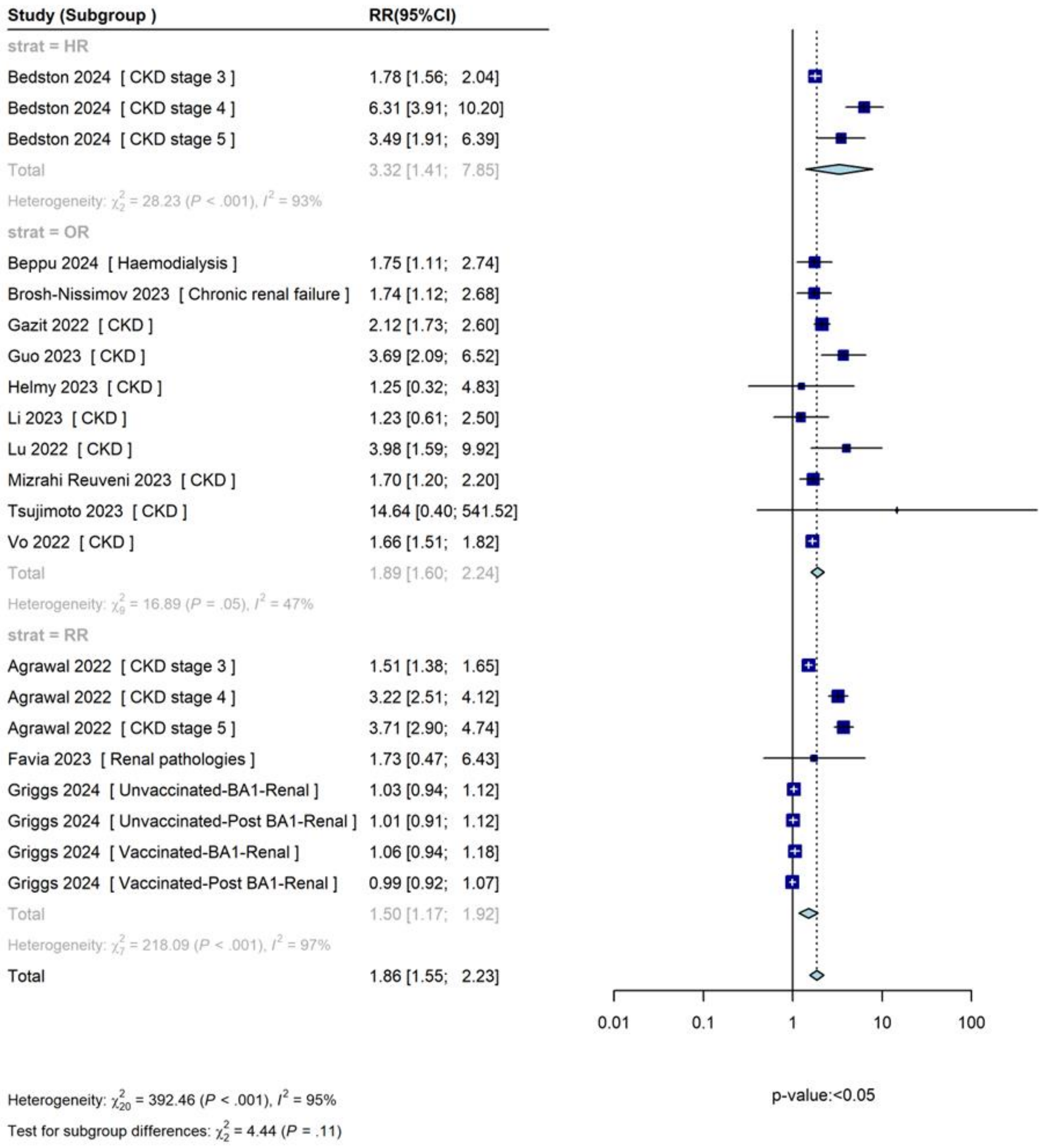

CI, confidence interval; CKD, chronic kidney disease; HR, hazard ratio; IRR, incidence rate ratio; OR, odds ratio; RR, risk ratio.

##### *2.6.7. Transplant*

*2.6.7.1. Supplemental eFigure 19. Transplant, Death Outcome (No. of Subgroups: 13)*

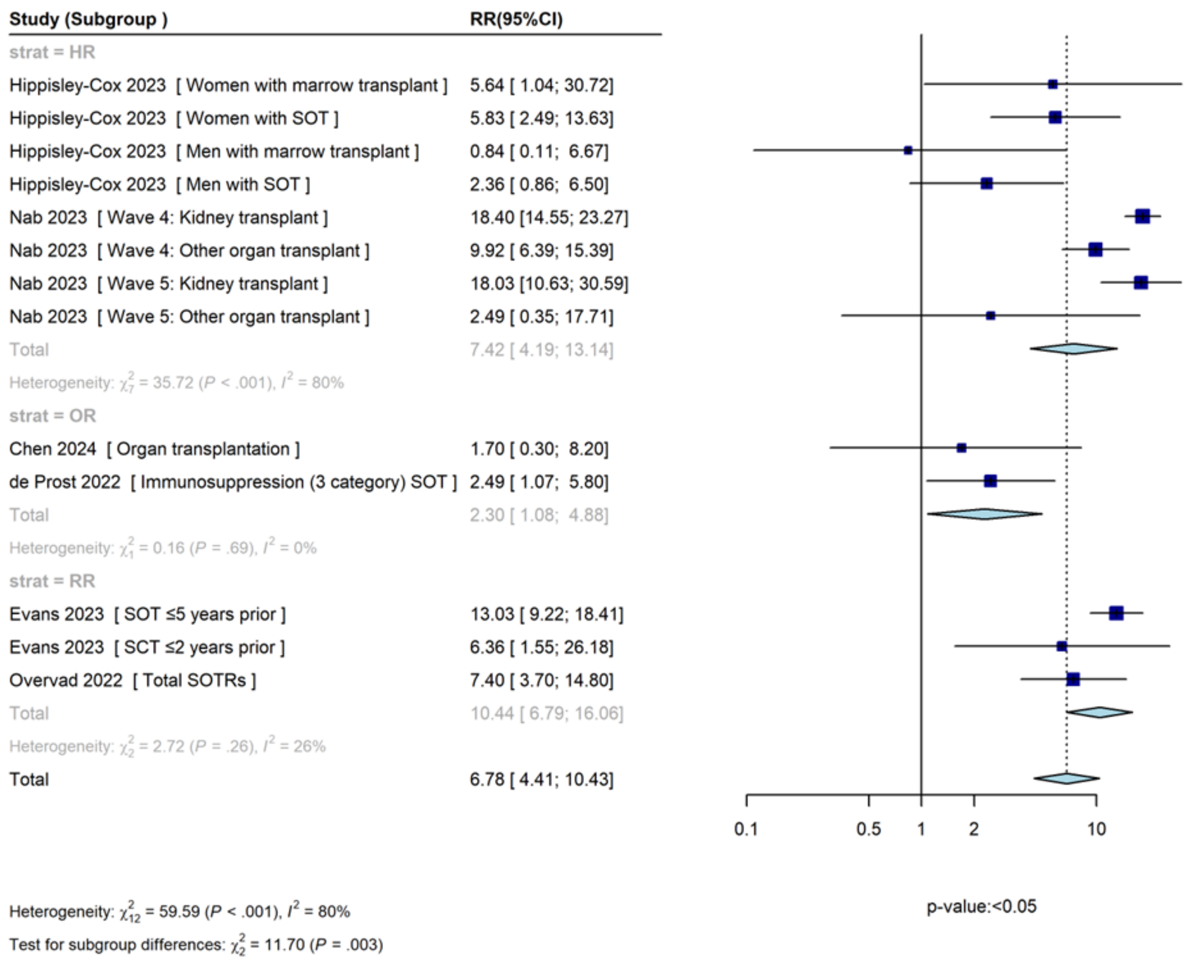

CI, confidence interval; HR, hazard ratio; IRR, incidence rate ratio; OR, odds ratio; RR, risk ratio; SCT, stem cell transplant; SOT, solid organ transplant; SOTR, solid organ transplant recipients.

*2.6.7.2. Supplemental eFigure 20. Transplant, Hospitalization Outcome (No. of Subgroups: 8)*

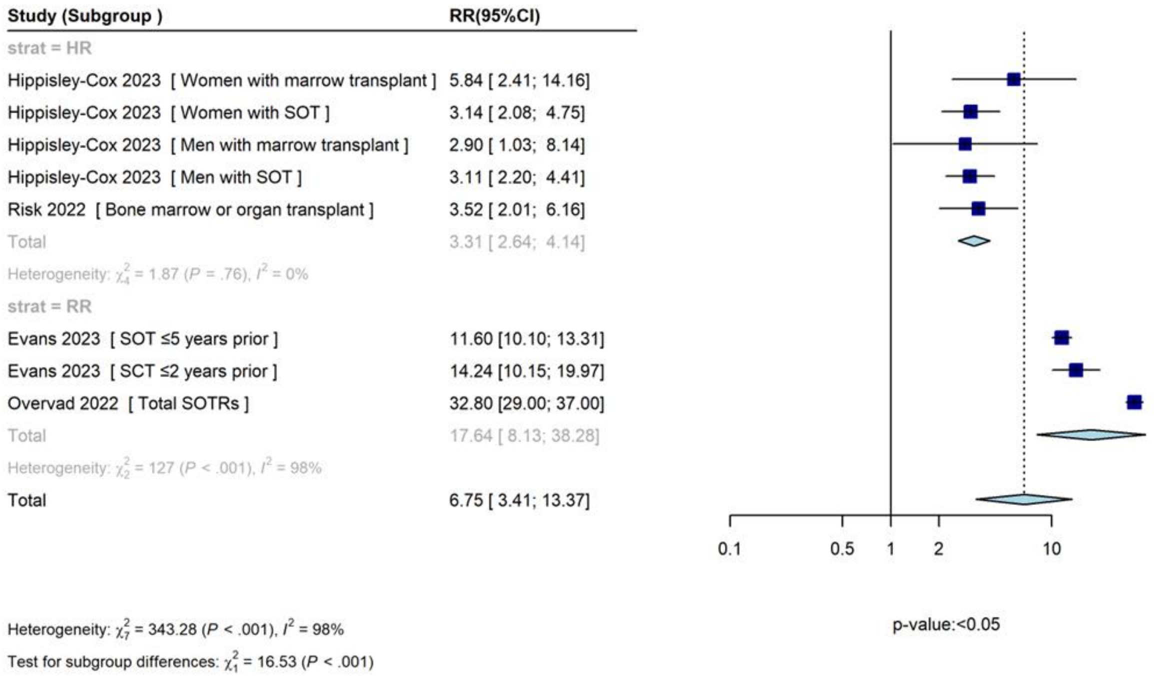

CI, confidence interval; HR, hazard ratio; IRR, incidence rate ratio; OR, odds ratio; RR, risk ratio; SCT, stem cell transplant; SOT, solid organ transplant; SOTR, solid organ transplant recipients.

*2.6.7.3. Transplant, ICU Admission Outcome (No. of Studies: 1; No. of Subgroups: 2)*

The studies reported the following results for the 'ICU' outcome in the 'Transplant' populations:

- Evans 2023[33] (solid organ transplant ≤5 years prior): IRR (95% CI): 24.74 (14.28, 42.85)
- Evans 2023[33] (stem cell transplant ≤2 years prior): IRR (95% CI): 24.92 (6.03, 102.98)

Both 'Transplant' subgroups in the Evans 2023 study found statistically significant increases in the risk of the 'ICU' outcome. However, both subgroups, the 'Stem cell transplant' subgroup in particular, had very wide CIs, indicating a low number of 'Transplant' participants who were admitted to the ICU.

*2.6.7.4. Supplemental eFigure 21. Transplant, Combined Outcome (No. of Subgroups: 5)*

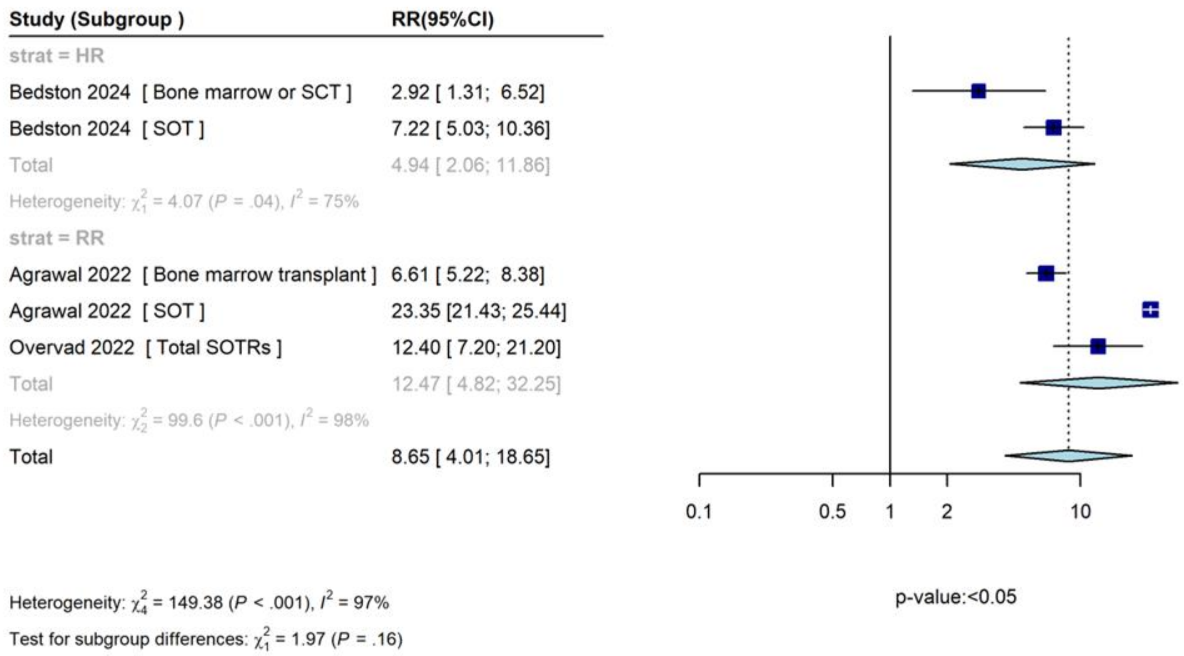

CI, confidence interval; HR, hazard ratio; IRR, incidence rate ratio; OR, odds ratio; RR, risk ratio; SCT, stem cell transplant; SOT, solid organ transplant; SOTR, solid organ transplant recipients.

#### References

**1.** Ouzzani M, Hammady H, Fedorowicz Z, Elmagarmid A. Rayyan-a web and mobile app for systematic reviews. *Syst Rev*. 2016;5(1):210.

**2.** Wells G SB, O'Connell D, Peterson J, Welch V, Losos M. The Newcastle-Ottawa Scale (NOS) for assessing the quality of nonrandomised studies in meta-analyses. https://www.ohri.ca/programs/clinical_epidemiology/oxford.asp.

**3.** JBI. Critical appraisal tools. https://jbi.global/critical-appraisal-tools.

**4.** Greenland S. Quantitative methods in the review of epidemiologic literature. *Epidemiol Rev*. 1987;9:1-30.

**5.** DerSimonian R, Laird N. Meta-analysis in clinical trials. *Control Clin Trials*. 1986;7(3):177-188.

**6.** Higgins JPT TJ, Chandler J, Li T, Page MJ, Welch V, eds. *Cochrane Handbook for Systematic Reviews of Interventions [Internet]. Version 6.4 Cochrane, updated August 2023. [cited 2024 May 28]. Available from: https://training.cochrane.org/handbook/current.*

**7.** Stuck AE, Rubenstein LZ, Wieland D. Bias in meta-analysis detected by a simple, graphical test. Asymmetry detected in funnel plot was probably due to true heterogeneity. *BMJ*. 1998;316(7129):469; author reply 470-461.

**8.** Starkey T, Ionescu MC, Tilby M, et al. A population-scale temporal case–control evaluation of COVID-19 disease phenotype and related outcome rates in patients with cancer in England (UKCCP). *Sci Rep*. 2023;13(1):11327.

**9.** Russell SL, Klaver BRA, Harrigan SP, et al. Clinical severity of Omicron subvariants BA.1, BA.2, and BA.5 in a population-based cohort study in British Columbia, Canada. *J Med Virol*. 2023;95(1):e28423.

**10.** Mayer C, Woo MS, Brehm TT, et al. History of cerebrovascular disease but not dementia increases the risk for secondary vascular events during SARS-CoV-2 infection with presumed Omicron variant: a retrospective observational study. *Eur J Neurol*. 2023;30(8):2297-2304.

**11.** World Health Organization. Classification of Omicron (B.1.1.529): SARS-CoV-2 variant of concern. Updated November 26, 2021.https://www.who.int/news/item/26-11-2021-classification-of-omicron-(b.1.1.529)-sars-cov-2-variant-of-concern. Accessed July 3, 2024.

**12.** Agrawal U, Bedston S, McCowan C, et al. Severe COVID-19 outcomes after full vaccination of primary schedule and initial boosters: pooled analysis of national prospective cohort studies of 30 million individuals in England, Northern Ireland, Scotland, and Wales. *Lancet*. 2022;400(10360):1305-1320.

**13.** AlBahrani S, AlBarrak A, Al-Musawi T, et al. COVID-19 vaccine had a significant positive impact on patients with SARS-COV-2 during the third (Omicron) wave in Saudi Arabia. *J Infect Public Health*. 2022;15(11):1169-1174.

**14.** Arbel R, Sergienko R, Friger M, et al. Effectiveness of a second BNT162b2 booster vaccine against hospitalization and death from COVID-19 in adults aged over 60 years. *Nat Med*. 2022;28(7):1486-1490.

**15.** Arbel R, Peretz A, Sergienko R, et al. Effectiveness of a bivalent mRNA vaccine booster dose to prevent severe COVID-19 outcomes: a retrospective cohort study. *Lancet Infect Dis*. 2023;23(8):914-921.

**16.** Bahremand T, Yao JA, Mill C, et al. COVID-19 hospitalisations in immunocompromised individuals in the Omicron era: a population-based observational study using surveillance data in British Columbia, Canada. *Lancet Reg Health Am*. 2023;20:100461.

**17.** Bao S, Lu G, Kang Y, et al. A diagnostic model for serious COVID-19 infection among older adults in Shanghai during the Omicron wave. *Front Med (Lausanne)*. 2022;9:1018516.

**18.** Bedston S, Almaghrabi F, Patterson L, et al. Risk of severe COVID-19 outcomes after autumn 2022 COVID-19 booster vaccinations: a pooled analysis of national prospective cohort studies involving 7.4 million adults in England, Northern Ireland, Scotland and Wales. *Lancet Reg Health Eur*. 2024;37:100816.

**19.** Benites-Godínez V, Mendoza-Cano O, Trujillo X, et al. Survival analysis and contributing factors among PCR-confirmed adult inpatients during the endemic phase of COVID-19. *Diseases*. 2023;11(3):119.

**20.** Beppu H, Fukuda T, Otsubo N, et al. Comparative outcomes of hemodialysis patients facing pre-Omicron and Omicron COVID-19 epidemics. *Ther Apher Dial*. 2024;28(1):51-60.

**21.** Beraud G, Bouetard L, Civljak R, et al. Impact of vaccination on the presence and severity of symptoms in hospitalized patients with an infection of the Omicron variant (B.1.1.529) of the SARS-CoV-2 (subvariant BA.1). *Clin Microbiol Infect*. 2023;29(5):642-650.

**22.** Bournia V-K, Fragoulis GE, Mitrou P, et al. Outcomes of COVID-19 Omicron variant in patients with rheumatoid arthritis: a nationwide Greek cohort study. *Rheumatology*. 2023;63(4):1130-1138.

**23.** Briciu V, Topan A, Calin M, et al. Comparison of COVID-19 severity in vaccinated and unvaccinated patients during the Delta and Omicron wave of the pandemic in a Romanian tertiary infectious diseases hospital. *Healthcare*. 2023;11(3):373.

**24.** Brosh-Nissimov T, Hussein K, Wiener-Well Y, et al. Hospitalized patients with severe coronavirus disease 2019 during the Omicron wave in Israel: benefits of a fourth vaccine dose. *Clin Infect Dis*. 2022;76(3):e234-e239.

**25.** Chen Z, Tian F, Zeng Y. Polypharmacy, potentially inappropriate medications, and drug-drug interactions in older COVID-19 inpatients. *BMC Geriatrics*. 2023;23(1):774.

**26.** Chen CL, Teng CK, Chen WC, et al. Clinical characteristics and treatment outcomes among the hospitalized elderly patients with COVID-19 during the late pandemic phase in central Taiwan. *J Microbiol Immunol Infect*. 2024;57(2):257-268.

**27.** Choi S-H, Choi JH, Lee JK, et al. Clinical characteristics and outcomes of children with SARS-CoV-2 infection during the Delta and Omicron variant-dominant periods in Korea. *J Korean Med Sci*. 2023;38(9):e65.

**28.** de Prost N, Audureau E, Heming N, et al. Clinical phenotypes and outcomes associated with SARS-CoV-2 variant Omicron in critically ill French patients with COVID-19. *Nat Commun*. 2022;13(1):6025.

**29.** de Prost N, Audureau E, Préau S, et al. Clinical phenotypes and outcomes associated with SARS-CoV-2 Omicron variants BA.2, BA.5 and BQ.1.1 in critically ill patients with COVID-19: a prospective, multicenter cohort study. *Intensive Care Med Exp*. 2023;11(1):48.

**30.** Drummond PD, de Salles DB, de Souza NSH, et al. Profile and outcomes of hospitalized COVID-19 patients during the prevalence of the Omicron variant according to the Brazilian regions: a retrospective cohort study from 2022. *Vaccines*. 2023;11(10):1568.

**31.** Elamin MY, Maslamani YA, Alsheikh FA, et al. Impact of vaccination on morbidity and mortality in adults hospitalized with COVID-19 during the Omicron wave in the Jazan Region, Saudi Arabia. *Saudi Med J*. 2024;45(2):179-187.

**32.** Ellis RJ, Moffatt CR, Aaron LT, et al. Factors associated with hospitalisations and deaths of residential aged care residents with COVID-19 during the Omicron (BA.1) wave in Queensland. *Med J Aust*. 2023;218(4):174-179.

**33.** Evans RA, Dube S, Lu Y, et al. Impact of COVID-19 on immunocompromised populations during the Omicron era: insights from the observational population-based INFORM study. *Lancet Reg Health Eur*. 2023;35:100747.

**34.** Favia G, Barile G, Tempesta A, et al. Relationship between oral lesions and severe SARS-CoV-2 infection in intensive care unit patients. *Oral Dis*. 2024;30(3):1296-1303.

**35.** Flacco ME, Acuti Martellucci C, Soldato G, et al. Predictors of SARS-CoV-2 infection and severe and lethal COVID-19 after three years of follow-up: a population-wide study. *Viruses*. 2023;15(9):1794.

**36.** Gazit S, Saciuk Y, Perez G, et al. Short term, relative effectiveness of four doses versus three doses of BNT162b2 vaccine in people aged 60 years and older in Israel: retrospective, test negative, case-control study. *BMJ*. 2022;377:e071113.

**37.** Griggs EP, Mitchell PK, Lazariu V, et al. Clinical epidemiology and risk factors for critical outcomes among vaccinated and unvaccinated adults hospitalized with COVID-19-VISION Network, 10 states, June 2021-March 2023. *Clin Infect Dis*. 2024;78(2):338-348.

**38.** Guo Y, Guo Y, Ying H, et al. In-hospital adverse outcomes and risk factors among chronic kidney disease patients infected with the omicron variant of SARS-CoV-2: a single-center retrospective study. *BMC Infect Dis*. 2023;23(1):698.

**39.** Helmy MA, Milad LM, Hasanin AM, et al. Parasternal intercostal thickening at hospital admission: a promising indicator for mechanical ventilation risk in subjects with severe COVID-19. *J Clin Monit Comput*. 2023;37(5):1287-1293.

**40.** Hippisley-Cox J, Khunti K, Sheikh A, Nguyen-Van-Tam JS, Coupland CAC. Risk prediction of covid-19 related death or hospital admission in adults testing positive for SARS-CoV-2 infection during the omicron wave in England (QCOVID4): cohort study. *BMJ*. 2023;381:e072976.

**41.** Jamaati H, Karimi S, Ghorbani F, et al. Effectiveness of different vaccine platforms in reducing mortality and length of ICU stay in severe and critical cases of COVID-19 in the Omicron variant era: a national cohort study in Iran. *J Med Virol*. 2023;95(3):e28607.

**42.** Kang J-M, Kang M, Kim Y-E, et al. Severe coronavirus disease 2019 in pediatric solid organ transplant recipients: big data convergence study in Korea (K-COV-N cohort). *Int J Infect Dis*. 2023;134:220-227.

**43.** Karageorgou V, Papaioannou AI, Kallieri M, et al. Patients hospitalized for COVID-19 in the periods of Delta and Omicron variant dominance in Greece: determinants of severity and mortality. *J Clin Med*. 2023;12(18):5904.

**44.** Kim SH, Kim T, Choi H, Shin TR, Sim YS. Clinical outcome and prognosis of a nosocomial outbreak of COVID-19. *J Clin Med*. 2023;12(6):2279.

**45.** Klein EY, Fall A, Norton JM, et al. Severity outcomes associated with SARS-CoV-2 XBB variants, an observational analysis. *J Clin Virol*. 2023;165:105500.

**46.** Konermann FM, Gessler N, Wohlmuth P, et al. High in-hospital mortality in SARS-CoV-2-infected patients with active cancer disease during Omicron phase of the pandemic: insights from the CORONA Germany study. *Oncol Res Treat*. 2023;46(5):201-210.

**47.** Lee CM, Kim M, Park SW, et al. Clinical outcomes and immunological features of COVID-19 patients receiving B-cell depletion therapy during the Omicron era. *Infect Dis (Lond)*. 2024;56(2):116-127.

**48.** Li H, Jia X, Wang Y, et al. Differences in the severity and mortality risk factors for patients hospitalized for COVID-19 pneumonia between the early wave and the very late stage of the pandemic. *Front Med (Lausanne)*. 2023;10:1238713.

**49.** Li D-J, Zhou C-C, Huang F, Shen F-M, Li Y-C. Clinical features of omicron SARS-CoV-2 variants infection associated with co-infection and ICU-acquired infection in ICU patients. *Front Public Health*. 2024;11:1320340.

**50.** Liu Y, Qi Z, Bai M, et al. Combination of chest computed tomography value and clinical laboratory data for the prognostic risk evaluation of patients with COVID-19. *Int J Gen Med*. 2023;16:3829-3842.

**51.** Lu G, Zhang Y, Zhang H, et al. Geriatric risk and protective factors for serious COVID-19 outcomes among older adults in Shanghai Omicron wave. *Emerg Microbes Infect*. 2022;11(1):2045-2054.

**52.** Mendoza-Cano O, Trujillo X, Ríos-Silva M, et al. Association between vaccination status for COVID-19 and the risk of severe symptoms during the endemic phase of the disease. *Vaccines*. 2023;11(10):1512.

**53.** Mizrahi Reuveni M, Kertes J, Shapiro Ben David S, et al. Risk stratification model for severe COVID-19 disease: a retrospective cohort study. *Biomedicines*. 2023;11(3):767.

**54.** Morris CP, Eldesouki RE, Sachithanandham J, et al. Omicron subvariants: clinical, laboratory, and cell culture characterization. *Clin Infect Dis*. 2023;76(7):1276-1284.

**55.** Nab L, Parker EPK, Andrews CD, et al. Changes in COVID-19-related mortality across key demographic and clinical subgroups in England from 2020 to 2022: a retrospective cohort study using the OpenSAFELY platform. *Lancet Public Health*. 2023;8(5):e364-e377.

**56.** Nevejan L, Ombelet S, Laenen L, et al. Severity of COVID-19 among hospitalized patients: Omicron remains a severe threat for immunocompromised hosts. *Viruses*. 2022;14(12):2736.

**57.** O'Leary AL, Wattengel BA, Carter MT, Drye AF, Mergenhagen KA. Risk factors associated with mortality in hospitalized patients with laboratory confirmed SARS-CoV-2 infection during the period of omicron (B.1.1.529) variant predominance. *Am J Infect Control*. 2023;51(6):603-606.

**58.** Overvad M, Koch A, Jespersen B, et al. Outcomes following SARS-CoV-2 infection in individuals with and without solid organ transplantation-A Danish nationwide cohort study. *Am J Transplant*. 2022;22(11):2627-2636.

**59.** Parajuli P, Sabo R, Alsaadawi R, et al. Fibrosis-4 (FIB-4) index as a predictor for mechanical ventilation and 30-day mortality across COVID-19 variants. *J Clin Transl Sci*. 2023;7(1):e213.

**60.** Parra-Bracamonte GM, Lopez-Villalobos N, Velazquez MA, et al. Comparative analysis of risk factors for COVID-19 mortality before, during and after the vaccination programme in Mexico. *Public Health*. 2023;215:94-99.

**61.** Patton MJ, Orihuela CJ, Harrod KS, et al. COVID-19 bacteremic co-infection is a major risk factor for mortality, ICU admission, and mechanical ventilation. *Crit Care*. 2023;27(1):34.

**62.** Puyat JH, Fowokan A, Wilton J, et al. Risk of COVID-19 hospitalization in people living with HIV and HIV-negative individuals and the role of COVID-19 vaccination: a retrospective cohort study. *Int J Infect Dis*. 2023;135:49-56.

**63.** Radhakrishnan N, Liu M, Idowu B, et al. Comparison of the clinical characteristics of SARS-CoV-2 Delta (B.1.617.2) and Omicron (B.1.1.529) infected patients from a single hospitalist service. *BMC Infect Dis*. 2023;23(1):747.

**64.** Rasmussen LD, Cowan S, Gerstoft J, et al. Outcomes following severe acute respiratory syndrome coronavirus 2 infection among individuals with and without HIV in Denmark. *AIDS*. 2023;37(2):311-321.

**65.** Risk M, Hayek SS, Schiopu E, et al. COVID-19 vaccine effectiveness against omicron (B.1.1.529) variant infection and hospitalisation in patients taking immunosuppressive medications: a retrospective cohort study. *Lancet Rheumatol*. 2022;4(11):e775-e784.

**66.** Shakor ASaA, Samsudin EZ, Chen XW, Ghazali MH. Factors associated with COVID-19 brought-in deaths: a data-linkage comparative cross-sectional study. *J Infect Public Health*. 2023;16(12):2068-2078.

**67.** Shi HJ, Yang J, Eom JS, et al. Clinical characteristics and risk factors for mortality in critical COVID-19 patients aged 50 years or younger during Omicron wave in Korea: comparison with patients older than 50 years of age. *J Korean Med Sci*. 2023;38(28):e217.

**68.** Simmons AE, Amoako A, Grima AA, et al. Vaccine effectiveness against hospitalization among adolescent and pediatric SARS-CoV-2 cases between May 2021 and January 2022 in Ontario, Canada: a retrospective cohort study. *PLoS One*. 2023;18(3):e0283715.

**69.** Skarbinski J, Wood MS, Chervo TC, et al. Risk of severe clinical outcomes among persons with SARS-CoV-2 infection with differing levels of vaccination during widespread Omicron (B.1.1.529) and Delta (B.1.617.2) variant circulation in Northern California: a retrospective cohort study. *Lancet Reg Health Am*. 2022;12:100297.

**70.** Svensson ALL, Emborg H-D, Bartels LE, et al. Outcomes following SARS-CoV-2 infection in individuals with and without inflammatory rheumatic diseases: a Danish nationwide cohort study. *Ann Rheum Dis*. 2023;82(10):1359-1367.

**71.** Tsujimoto Y, Kobayashi M, Oku T, et al. Outcomes in novel hospital-at-home model for patients with COVID-19: a multicentre retrospective cohort study. *Fam Pract*. 2023;40(5-6):662-670.

**72.** Vo AD, La J, Wu JT, et al. Factors associated with severe COVID-19 among vaccinated adults treated in US veterans affairs hospitals. *JAMA Netw Open*. 2022;5(10):e2240037.

**73.** Wang X, Zein J, Ji X, Lin DY. Impact of vaccination, prior infection, and therapy on Omicron infection and mortality. *J Infect Dis*. 2023;227(8):970-976.

**74.** Ward IL, Robertson C, Agrawal U, et al. Risk of COVID-19 death in adults who received booster COVID-19 vaccinations in England. *Nat Commun*. 2024;15(1):398.

**75.** Xin S, Chen W, Yu Q, Gao L, Lu G. Effect of the number of coronavirus disease 2019 (COVID-19) vaccination shots on the occurrence of pneumonia, severe pneumonia, and death in SARS-CoV-2-infected patients. *Front Public Health*. 2024;11:1330106.

**76.** Xing Y, Sun Y, Tang M, et al. Variables associated with 30-day mortality in very elderly COVID-19 patients. *Clin Interv Aging*. 2023;18:1155-1162.

**77.** Yang H, Wang Z, Zhang Y, et al. Clinical characteristics and factors for serious outcomes among outpatients infected with the Omicron subvariant BF.7. *J Med Virol*. 2023;95(8):e28977.

**78.** Zhang Y, Han J, Sun F, et al. A practical scoring model to predict the occurrence of critical illness in hospitalized patients with SARS-CoV-2 omicron infection. *Front Microbiol*. 2022;13:1031231.

**79.** Zhao Q, Zheng B, Han B, et al. Is azvudine comparable to nirmatrelvir-ritonavir in real-world efficacy and safety for hospitalized patients with COVID-19? a retrospective cohort study. *Infect Dis Ther*. 2023;12(8):2087-2102.

**80.** Zhu Z, Cai J, Tang Q, et al. Circulating eosinophils associated with responsiveness to COVID-19 vaccine and the disease severity in patients with SARS-CoV-2 omicron variant infection. *BMC Pulm Med*. 2023;23(1):177.
